# Social determinants of injection drug use-associated bacterial infections and treatment outcomes: systematic review and meta-analysis

**DOI:** 10.1101/2024.09.20.24313898

**Authors:** Thomas D. Brothers, Dan Lewer, Matthew Bonn, Inhwa Kim, Emilie Comeau, Mary Figgatt, William Eger, Duncan Webster, Andrew Hayward, Magdalena Harris

## Abstract

**Background:** Individual injecting practices (e.g., intramuscular injecting, lack of skin cleaning) are known risk factors for injection drug use-associated bacterial and fungal infections; however, social contexts shape individual behaviours and health outcomes. We sought to synthesize studies assessing potential social determinants of injecting-related infections and treatment outcomes.

**Methods:** We searched five databases for studies published between 1 January 2000 and 18 February 18 2021 (PROSPERO CRD42021231411). We included studies of association (aetiology), assessing social determinants, substance use, and health services exposures influencing development of injecting-related infections and treatment outcomes. We pooled effect estimates via random effects meta-analyses.

**Results:** We screened 4,841 abstracts and included 107 studies. Several factors were associated with incident or prevalent injecting-related infections: woman/female gender/sex (adjusted odds ratio [aOR] 1.57, 95% confidence interval [CI] 1.36-1.83; n=20 studies), homelessness (aOR 1.29, 95%CI 1.16-1.45; n=13 studies), cocaine use (aOR 1.31, 95%CI 1.02–1.69; n=10 studies), amphetamine use (aOR 1.74, 95%CI 1.39-2.23; n=2 studies), public injecting (aOR 1.40, 95%CI 1.05–1.88; n=2 studies), requiring injecting assistance (aOR 1.78, 95%CI 1.40–2.27; n=8 studies), and use of opioid agonist treatment (aOR 0.92, 95%CI 0.89–0.95; n=9 studies). Studies assessing outcomes during treatment (e.g., premature hospital discharge) or afterward (e.g., rehospitalization; all-cause mortality) typically had smaller sample sizes and imprecise effect estimates.

**Conclusions:** Injecting-related infections and treatment outcomes may be shaped by multiple social contextual factors. Approaches to prevention and treatment should look beyond individual injecting practices towards addressing the social and material conditions within which people live, acquire and consume drugs, and access health care.

## Introduction

Injecting-related bacterial and fungal infections cause significant morbidity and mortality among people who inject drugs, with 6-32% of people reporting an infection in the past month and up to 64% reporting a skin and soft-tissue infection (SSTI) in the past year.^1^ Incidence of these infections is increasing in North America, Europe, and Australia.^2–5^ Specific drug preparation and injecting practices (including subcutaneous or intramuscular injecting, reusing blunted or contaminated needles and syringes, and not sterilizing skin before injecting) are known risk factors.^1,6^ Individual-level educational interventions have been developed to promote safer drug injecting techniques, but these have shown inconsistent efficacy.^7–9^ Individual-level interventions may have limited impact because drug injecting practices are shaped by social contexts and material resources.^10^ For example, people without secure housing are more likely to inject in public or unhygienic spaces (e.g., abandoned buildings).^10,11^ People with insufficient access to needle and syringe programs may need to reuse contaminated equipment.^10,12,13^

Treatment of injection drug use-associated bacterial and fungal infections is also often suboptimal.^10,14–16^ People who inject drugs describe negative experiences of untreated pain and withdrawal in health care settings, which discourages access to care.^10,17–19^ Clinicians caring for people with injecting-related infections describe not knowing how best to help.^16,20^ Poor care experiences and outcomes are also affected by social determinants and system-level factors.^10,14,21,22^ Most acute care hospitals do not integrate substance use and addiction care, and many hospitals have abstinence-based policies that lead patients to surreptitiously use drugs (e.g., in locked bathrooms) or leave the hospital prematurely.^16,22–27^ As a result, risk of fatal opioid overdose is two to four times higher in the days after hospital discharge compared to other times,^27,28^ and rehospitalizations with recurrent injecting-related infections are common.^29,30^

Prevention and treatment strategies for injecting-related infections may be greatly improved if clinicians and health systems look more broadly to the social and structural factors that shape individual injecting practices and treatment outcomes. Such approaches have informed prevention and treatment strategies for HIV^31–34^ and hepatitis C virus^32,35^, and prevention strategies for opioid overdose^33,36^, but less is known about how social determinants influence injecting-related bacterial and fungal infections.^10,37^

To better understand and quantify the influence of social determinants on injecting-related infections and treatment outcomes (and to identify opportunities for novel interventions to improve prevention and treatment), we sought to systematically review and meta-analyse the quantitative literature on this topic. This review seeks to answer the question, “Among people who inject drugs, what social and structural factors influence the development of, treatment of, and outcomes of injecting-related bacterial and fungal infections?”

## Methods

We registered (PROSPERO CRD42021231411) and published a protocol^37^ and before conducting the search. We modified the protocol after full-text review. The protocol specified a “mixed studies” review of quantitative, qualitative, and mixed-methods sources.^38^ As we identified more studies than anticipated, we separately reviewed and synthesized quantitative and qualitative studies. Here, we report the quantitative systematic review and meta-analyses. Qualitative results are reported elsewhere.^10^ Our approach is informed by the Conducting Systematic Reviews and Meta-Analyses of Observational Studies of Etiology (COSMOS-E)^39^ guidance. We followed Preferred Reporting Items for Systematic reviews and Meta-Analyses (PRISMA) reporting guidelines.^40^

### Eligibility criteria

Eligibility criteria are detailed in the protocol.^37^ Briefly, we included quantitative studies of association (aetiology),^39,41^ published in peer-reviewed journals. We followed the Population, Exposures, Outcomes approach.^41^ “Participants” were people who inject psychoactive substances. “Exposures” were social or environmental factors that could affect risk for developing infections or influence their treatment. This includes social, economic, and political factors such as housing, income, drug policies, and other socially-patterned exposures such as availability of (or use of) different substances, harm reduction services, drug treatment, and health care. Informed by a socio-ecological model (the “intersectional risk environment”^33,34^), we also considered socially-constructed identities and locations within social power hierarchies, including by gender/sex and race/ethnicity.^33^ While some sociodemographic characteristics (like gender, sex, or age) may confer effects through both biological and social-structural pathways, these are often interlinked and we expected that quantitative studies would not attempt to isolate purported biological-only effects.^33,36^ Potential differences by gender/sex may reflect structural sexism, and potential differences by race/ethnicity may reflect structural racism.^33,36,42^ “Outcomes” were injecting-related bacterial or fungal infections (and any reported outcomes during and after treatment of these infections), including SSTI (abscess, cellulitis, necrotizing fasciitis), sepsis or bacteraemia, vascular infections (endocarditis, septic phlebitis), bone and joint infections (osteomyelitis, septic arthritis, discitis), and central nervous system infections (epidural abscess, brain abscess, meningitis, encephalitis). We included studies published between 1 January 2000 and 18 February 2021, in English or French.

### Information sources, search strategy, and data management

We searched PubMed, EMBASE, Scopus, CINAHL, and PsycINFO databases. We supplemented this with forward and backward citation chaining and included articles from the review team’s personal files. We developed the final search strategy in consultation with a health sciences librarian (Appendix 1).

Search results were uploaded into Covidence and automatically de-duplicated. Two reviewers (TDB and either MB, EC, IK, or DL) screened abstracts against the inclusion criteria, resolving discrepancies through consensus. TDB assessed full-text reports for inclusion and recorded reasons for exclusion.

### Data collection

We developed and pilot-tested a data extraction form for studies with quantitative data. Data was extracted by TDB and checked independently by WE or MF. We extracted data on:

- First author and publication year
- Social and structural exposures included in the review
- Main exposure or estimand of the study (and whether all exposures assessed in our review reflect the study estimand)
- Infection types (e.g., SSTI, endocarditis, osteomyelitis, etc.)
- Infection-related outcomes
- Country (city) where study took place
- Sample size
- Sampling method (and parent study name, if applicable)
- Data collection period
- Inclusion criteria
- Proportion of sample that are women/female
- Age of sample
- Drugs used by ≥50% of the sample

As suggested in COSMOS-E guidance^39^, we manually extracted both unadjusted and fully covariate-adjusted effect estimates (with 95% confidence intervals), wherever possible. Following Kaufman^43^, we conceptualized unadjusted analyses as associative (e.g., “are people experiencing homelessness more likely to have injecting-related infections?”) and covariate-adjusted effect estimates as attempting to identify causal relationships (aetiology; e.g., “does homelessness contribute to injecting-related infections?”).

Where summary effect estimates (e.g., odds ratios) were not reported, we extracted frequencies to calculate unadjusted odds ratios and standard errors. When studies reported only stratified analyses (e.g., separate effect estimates among women and men) we kept both effect estimates for meta-analyses. When studies reported highly related effect estimates (e.g., for measures of both “lifetime” and “past six months” homelessness) from the same sample, we included only one in meta-analysis to avoid double-counting and documented reasons for inclusion or exclusion.^44^ Specifically, we selected the exposure-outcome pair with the most congruent timelines (e.g., “past six months” for exposure and outcome). As most studies reported effect estimates in odds ratios, we treated all relative effect estimates (including hazard ratios and rate ratios) as if they were odds ratios for meta-analysis.^39^ When studies did not report statistics for null or nonsignificant findings, (instead reporting, e.g., “no associations found”), we recorded where this was reported but we could not include these in meta-analyses.

### Critical appraisal

We applied the Mixed Methods Appraisal Tool^45^ (MMAT; which is designed for use with quantitative, qualitative, and mixed-methods studies), following the “user guide”.^46^ We included all studies which met both screening questions: “Are there clear research questions?” and “Do the collected data allow to address the research questions?”. For quantitative studies of association (aetiology), five criteria questions focus on whether the sample is representative of the target population, potential measurement error, if confounders are accounted for, and whether the exposure occurred as intended. The MMAT is scored only once per study, but many studies contain multiple exposure-outcome analyses that might have differing risks of bias. We scored a question as “No” if any exposure-outcome analysis in the study did not meet the criteria (e.g., if the timeline of exposure and outcome ascertainment did not align for one of many exposures assessed). Also, the MMAT only asks whether confounders were considered, not how they were selected or if they are appropriate. We scored “Yes” for studies that included covariate-adjusted analyses, no matter how covariates were selected. We assigned each study a score out of 5, by summing the questions answered “Yes”. We report MMAT scores for each study but did not otherwise use critical appraisal in syntheses.

### Meta-analyses

We conducted separate inverse variance meta-analyses for each exposure-outcome pair in *R* (version 4.2.2), using the *meta* package.^44^ We performed random-effects meta-analyses because we assumed there would be between-study heterogeneity (including different exposure and outcome definitions, study settings, and sampling frames).^39^ We applied the DerSimonian-Laird estimator for τ^2^, ^47^ with the Knapp-Harding adjustment (which assumes a t-distribution of the standard error of the pooled effect size and reduces the chance of false positives).^48^ We measured the percentage of total statistical variability attributable to between-study heterogeneity using *I^2^* statistics.^49^ We identified individual effect estimates as outliers if its confidence interval did not overlap with the confidence interval of the pooled effects (i.e., the effect size of the outlier is so extreme that it differs significantly from the meta-analysis summary effect).^50^

## Results

### Summary of included studies

The mixed-studies search identified 8,167 references; after de-duplication, we screened 4,841 abstracts and 631 full-text reports. We reviewed 16 additional full-text reports identified outside the search (Appendix 2). We excluded four quantitative studies because they failed the MMAT screening questions (Appendix 3).

We finally included 107 studies (Figure 1). There were 60 studies assessing incident or prevalent infections; 26 studies assessing outcomes during treatment (e.g., in-hospital mortality, premature hospital discharge); 29 studies assessing outcomes after treatment (e.g., infection-related rehospitalization, all-cause mortality); and five studies assessing colonisation with pathogenic bacteria.

**Figure 1.**
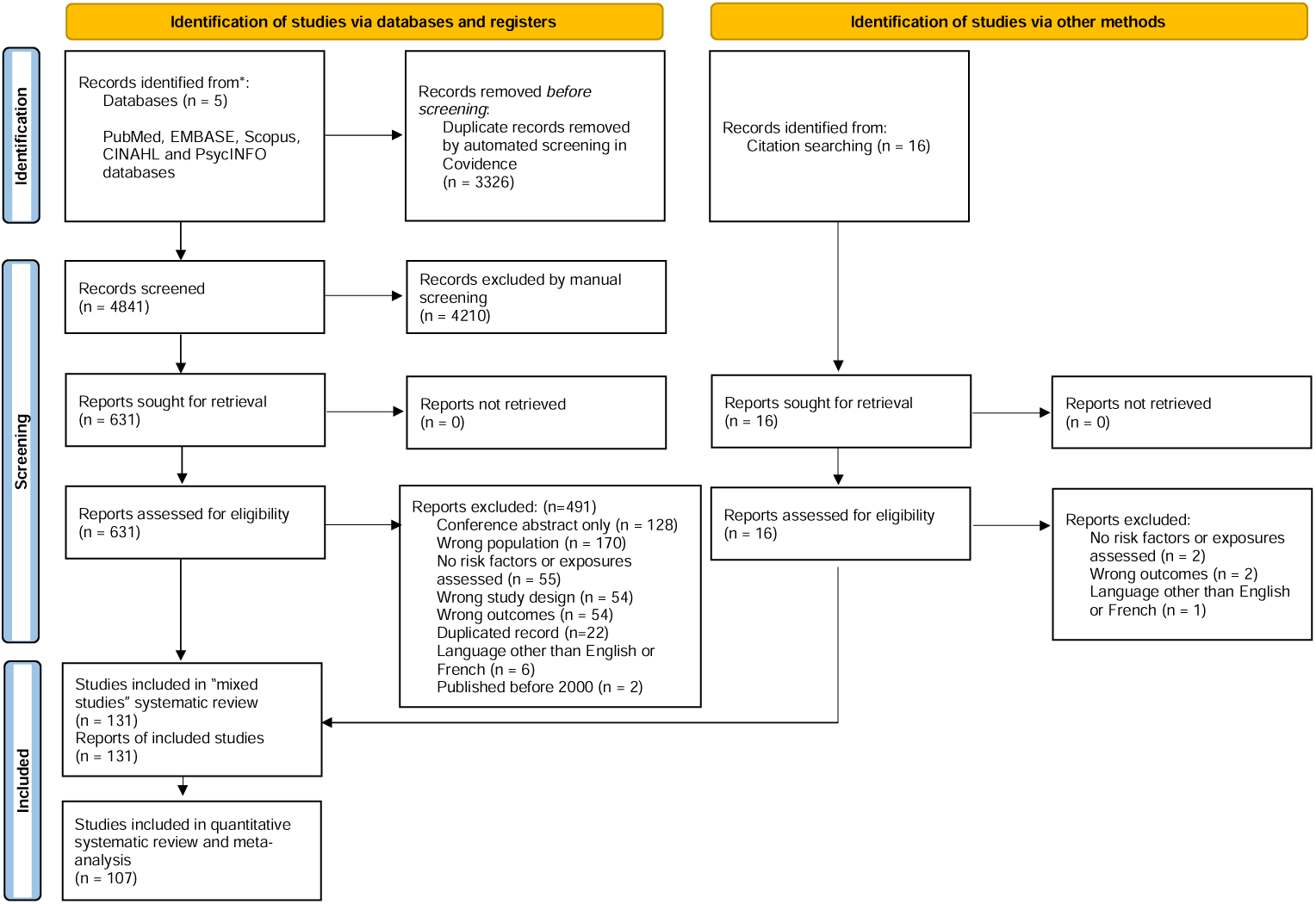
PRISMA 2020 flow diagram of included studies in quantitative systematic review of injecting-related infections.

See Table 1 for summary of included studies and Appendices 4-7 for detailed characteristics of each study. Two studies incorporated fungal infections (fungaemia and endogenous endophthalmitis); the remaining 105 studies focused only on bacterial infections. Among the 60 studies where the outcome was incident or prevalent injecting-related infections, 83% (n=50) included SSTI and samples were median 28.6% women/female. Among studies where the outcomes occurred during or after infection treatment, most focused on endocarditis (73% and 86%, respectively) and woman/female participants were more common (48.2% and 43.2%, respectively). Overall, most studies were from North America, followed by Europe (especially the United Kingdom). Few studies came from Asia (n=2) or Africa (n=1), and none from South America.

**Table 1.**
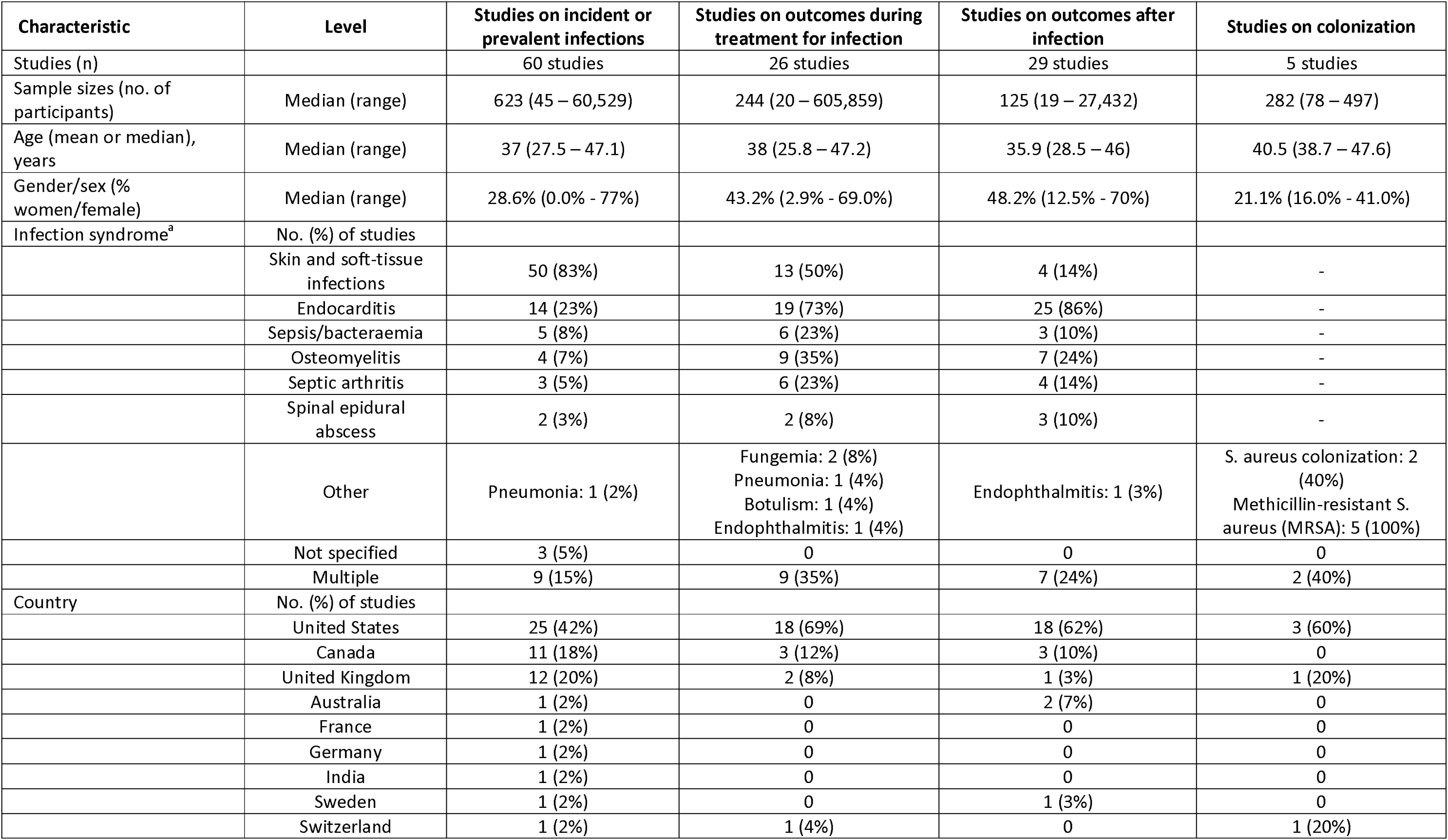
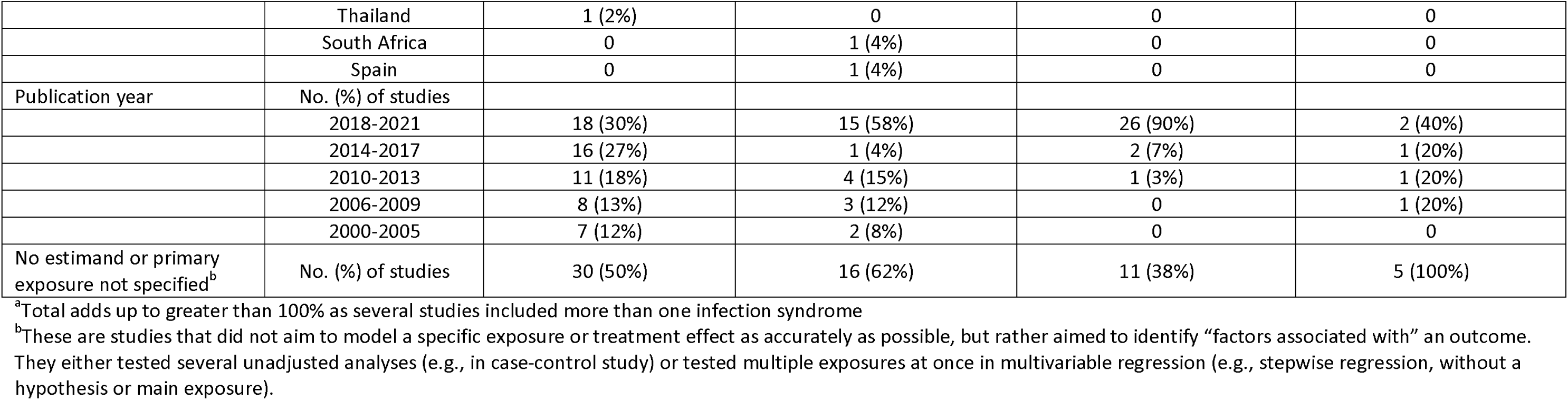
Summary of included studies in quantitative systematic review on injecting-related bacterial and fungal infections.

| Characteristic | Level | Studies on incident or prevalent infections | Studies on outcomes during treatment for infection | Studies on outcomes after infection | Studies on colonization |
| --- | --- | --- | --- | --- | --- |
| Studies (n) |  | 60 studies | 26 studies | 29 studies | 5 studies |
| Sample sizes (no. of participants) | Median (range) | 623 (45 – 60,529) | 244 (20 – 605,859) | 125 (19 – 27,432) | 282 (78 – 497) |
| Age (mean or median), years | Median (range) | 37 (27.5 – 47.1) | 38 (25.8 – 47.2) | 35.9 (28.5 – 46) | 40.5 (38.7 – 47.6) |
| Gender/sex (% women/female) | Median (range) | 28.6% (0.0% - 77%) | 43.2% (2.9% - 69.0%) | 48.2% (12.5% - 70%) | 21.1% (16.0% - 41.0%) |
| Infection syndrome<sup>a</sup> | No. (%) of studies |  |  |  |  |
|  | Skin and soft-tissue infections | 50 (83%) | 13 (50%) | 4 (14%) | - |
|  | Endocarditis | 14 (23%) | 19 (73%) | 25 (86%) | - |
|  | Sepsis/bacteraemia | 5 (8%) | 6 (23%) | 3 (10%) | - |
|  | Osteomyelitis | 4 (7%) | 9 (35%) | 7 (24%) | - |
|  | Septic arthritis | 3 (5%) | 6 (23%) | 4 (14%) | - |
|  | Spinal epidural abscess | 2 (3%) | 2 (8%) | 3 (10%) | - |
|  | Other | Pneumonia: 1 (2%) | Fungemia: 2 (8%)<br>Pneumonia: 1 (4%)<br>Botulism: 1 (4%)<br>Endophthalmitis: 1 (4%) | Endophthalmitis: 1 (3%) | S. aureus colonization: 2 (40%)<br>Methicillin-resistant S. aureus (MRSA): 5 (100%) |
|  | Not specified | 3 (5%) | 0 | 0 | 0 |
|  | Multiple | 9 (15%) | 9 (35%) | 7 (24%) | 2 (40%) |
| Country | No. (%) of studies |  |  |  |  |
|  | United States | 25 (42%) | 18 (69%) | 18 (62%) | 3 (60%) |
|  | Canada | 11 (18%) | 3 (12%) | 3 (10%) | 0 |
|  | United Kingdom | 12 (20%) | 2 (8%) | 1 (3%) | 1 (20%) |
|  | Australia | 1 (2%) | 0 | 2 (7%) | 0 |
|  | France | 1 (2%) | 0 | 0 | 0 |
|  | Germany | 1 (2%) | 0 | 0 | 0 |
|  | India | 1 (2%) | 0 | 0 | 0 |
|  | Sweden | 1 (2%) | 0 | 1 (3%) | 0 |
|  | Switzerland | 1 (2%) | 1 (4%) | 0 | 1 (20%) |
|  | Thailand | 1 (2%) | 0 | 0 | 0 |
|  | South Africa | 0 | 1 (4%) | 0 | 0 |
|  | Spain | 0 | 1 (4%) | 0 | 0 |
| Publication year | No. (%) of studies |  |  |  |  |
|  | 2018-2021 | 18 (30%) | 15 (58%) | 26 (90%) | 2 (40%) |
|  | 2014-2017 | 16 (27%) | 1 (4%) | 2 (7%) | 1 (20%) |
|  | 2010-2013 | 11 (18%) | 4 (15%) | 1 (3%) | 1 (20%) |
|  | 2006-2009 | 8 (13%) | 3 (12%) | 0 | 1 (20%) |
|  | 2000-2005 | 7 (12%) | 2 (8%) | 0 | 0 |
| No estimand or primary exposure not specified<sup>b</sup> | No. (%) of studies | 30 (50%) | 16 (62%) | 11 (38%) | 5 (100%) |

<sup>a</sup>Total adds up to greater than 100% as several studies included more than one infection syndrome
<sup>b</sup>These are studies that did not aim to model a specific exposure or treatment effect as accurately as possible, but rather aimed to identify “factors associated with” an outcome. They either tested several unadjusted analyses (e.g., in case-control study) or tested multiple exposures at once in multivariable regression (e.g., stepwise regression, without a hypothesis or main exposure).

See Appendices 8-11 for full MMAT critical appraisal results. See Appendices 12-15 for all extracted effect estimates and Appendix 16 for details of selecting and transforming effect estimates for inclusion in meta-analyses.

### Incident or prevalent injecting-related infections

Sixty studies assessed factors associated with incident or prevalent injecting-related infections, including sociodemographic factors (gender/sex, age, race/ethnicity, education, income/employment, relationship status, migration), social and housing support characteristics (incarceration, sex work, unstable housing/homelessness, food insecurity, health insurance), substance use-related factors (overdose history, heroin use, heroin formulation [i.e., tar vs. powder], prescription-type opioids, cocaine [including crack and powder], amphetamines [including methamphetamines], prescription-type stimulants, novel psychoactive stimulants, speedball/goofball use, alcohol, smoking), drug policy and injecting contexts (drug policy changes, drug purchasing network, public injecting, use of shooting galleries, police contacts/arrests, requiring or receiving injecting assistance, injecting with others), and harm reduction and drug treatment (needle and syringe programs, opioid agonist treatment, supervised consumption sites).

See Figure 2 for a summary of exposures and associated meta-analytic effect estimates. See Figure 3 for forest plots of unadjusted and adjusted effect estimates for three selected exposures: incarceration history, unstable housing or homelessness, and needle and syringe program use. See Appendix 17 for full results, including forest plots for each exposure.

**Figure 2.**
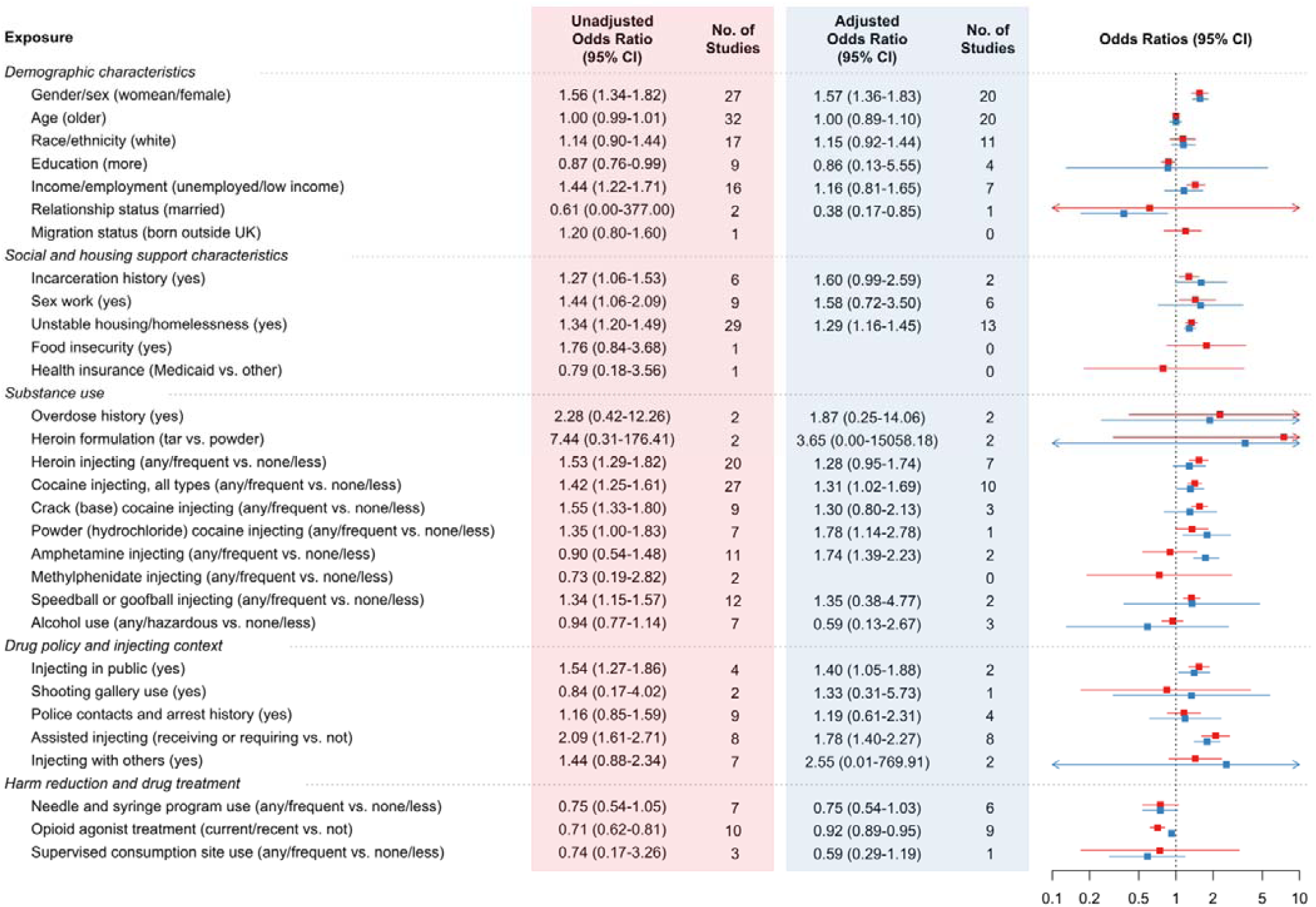
Summary of exposures and meta-analytic effect estimates among studies where outcome is incident or prevalent injecting-related bacterial infection. CI: confidence interval. See supplementary appendices for data on the remaining exposures (injecting prescription-type opioids; injecting other prescription-type stimulants, injecting novel psychoactive stimulants, smoking, drug policy changes, for drug-purchasing network) that could not be presented as single summary effect estimates.

**Figure 3.**
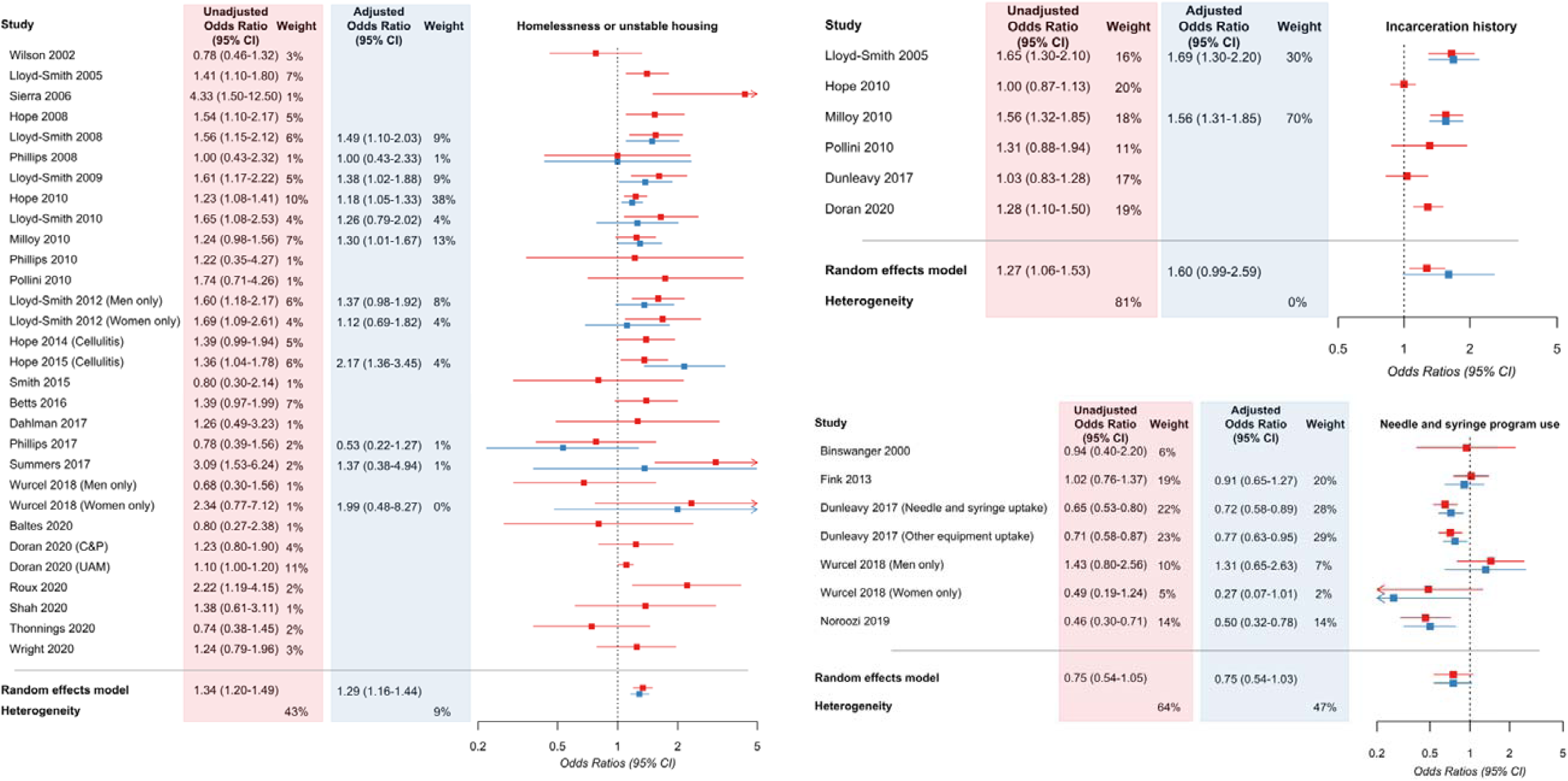
Forest plots for meta-analyses of selected exposures among studies where outcome is incident or prevalent injecting-related bacterial or fungal infections. In the panel on the left, the exposure is unstable housing or homelessness; in the panel on the top right, it is incarceration history; in the panel on the bottom right, the exposure is needle and syringe program use. CI: confidence interval. C&P: Care & Prevent cohort, UAM: Unlinked Anonymous Monitoring survey cohort (both reported in Doran 2020).

Briefly, we identified evidence to support associations between several factors with incident or prevalent injecting-related infections, in meta-analyses of covariate-adjusted effect estimates: woman/female gender/sex (adjusted odds ratio [aOR] 1.57, 95% confidence interval [CI] 1.36-1.83; I^2^ 47%; n=20 studies), unstable housing and homelessness (aOR 1.29, 95%CI 1.16-1.45; I^2^ 9%; n=13 studies; Figure 3), cocaine use (aOR 1.31, 95%CI 1.02–1.69; I^2^ 75%; n=10 studies), amphetamine use (aOR 1.74, 95%CI 1.39-2.23; I^2^ 0%; n=2 studies), public injecting (aOR 1.40, 95%CI 1.05–1.88; I^2^ 0%; n=2 studies), requiring/receiving injecting assistance (aOR 1.78, 95%CI 1.40–2.27; I^2^ 48%; n=8 studies), and use of opioid agonist treatment (aOR 0.92, 95%CI 0.89–0.95; I^2^ 50%; n=9 studies). For several other exposures, we identified evidence to support an association only in meta-analyses of unadjusted (but not covariate-adjusted) effect estimates: lower income/unemployment (unadjusted odds ratio [uOR] 1.44, 95%CI 1.22-1.71; I^2^ 79%; n=16 studies), incarceration history (uOR 1.27, 95%CI 1.06-1.53; I^2^ 81%; n=6 studies; Figure 3), sex work (uOR 1.49, 95%CI 1.06-2.09; I^2^ 89%; n=8 studies), heroin use (uOR 1.35, 95%CI 1.13-1.61; I^2^ 75%; n=20 studies), speedball (heroin and cocaine together) or goofball (heroin and methamphetamines together) use (uOR 1.34, 95%CI 1.15-1.57; I^2^ 51%; n=12 studies). For all other exposures (including needle and syringe program use, aOR 0.75, 95%CI 0.54-1.03; Figure 3), meta-analyses of unadjusted or covariate-adjusted effect estimates included null effect within their 95% confidence intervals.

### Outcomes occurring during treatment for injecting-related infections

We identified 26 studies assessing several outcomes during treatment/care of injecting-related bacterial infections. See Table 2 for the eight outcomes and list of exposures assessed in relation to each.

**Table 2.** Summary of outcomes and associated exposures assessed among studies where outcome occurs during treatment for injecting-related infections.

| Outcomes | Exposures assessed | Number of studies |
| --- | --- | --- |
| Health care-seeking for injecting-related infections | gender/sex; age; income/employment; sex work; unstable housing; incarceration; overdose history; migration status; heroin use; cocaine use; amphetamine use; opioid agonist treatment; supervised consumption site use | 4 |
| Self-treatment of abscess | gender/sex; age; race/ethnicity; unstable housing; heroin use; cocaine use; needle and syringe programs; several measures of access to health care (e.g., having a primary care provider or having health insurance) | 2 |
| Hospital admissions among people with an injecting-related SSTI | gender/sex; age; race/ethnicity; education; income/employment; sex work; migration status; unstable housing/homelessness; incarceration history; overdose history; heroin; cocaine; amphetamines; alcohol use; needle and syringe program use; access to health care (e.g., insurance, having a primary care provider); self-treatment of infections; hospital admission history | 2 |
| Premature hospital discharges, among people hospitalized with injecting-related infections | gender/sex; age; race/ethnicity; income/employment; unstable housing; overdose history; opioid use; cocaine; alcohol; other substance use; health care access; opioid agonist treatment; in-hospital addiction treatment; hospital characteristics; hospital policy; surgery during hospitalization | 10 |
| New/secondary bloodstream infections among people receiving antibiotic treatment | gender/sex; age; unstable housing and homelessness; substance use (heroin, stimulants, polysubstance use, other); substance use treatment; insertion of peripherally-inserted intravenous central catheters (PICC lines) for parenteral antimicrobial treatment | 1 |
| In-hospital death | gender/sex; age; race/ethnicity; overdose history; substance use (opioids, stimulants); health care access (insurance); hospital policies; surgery during hospital admission | 5 |
| Development of endogenous endophthalmitis | gender/sex; race/ethnicity; alcohol use; infection of central venous catheter | 1 |
| Respiratory failure among people with botulism | gender/sex; age | 1 |

Studies assessing outcomes during treatment for injecting-related infections typically had smaller sample sizes and imprecise effect estimates (compared to studies assessing incident or prevalent injecting-related infections) and findings were inconsistent between studies. Many exposures were only assessed in one study, limiting meta-analyses.

See Appendix 18 for full results including several meta-analyses for studies in the section. Among exposure-outcome pairs assessed in more than one study, we identified supporting evidence for only one association. Lacking health insurance was associated with increased risk of premature hospital discharge (aOR 2.07, 95%CI 1.09-3.91; I^2^ 85%; n=4 studies), among people hospitalized with injecting-related infections in one U.S. study. In other country settings (e.g., United Kingdom or Canada), universal public health insurance covers hospital services.

### Outcomes occurring after initial treatment of injecting-related infection

We identified 29 studies assessing outcomes after initial treatment for injecting-related infections. See Table 3 for six outcomes, and which exposures were assessed in relation to each.

**Table 3.**
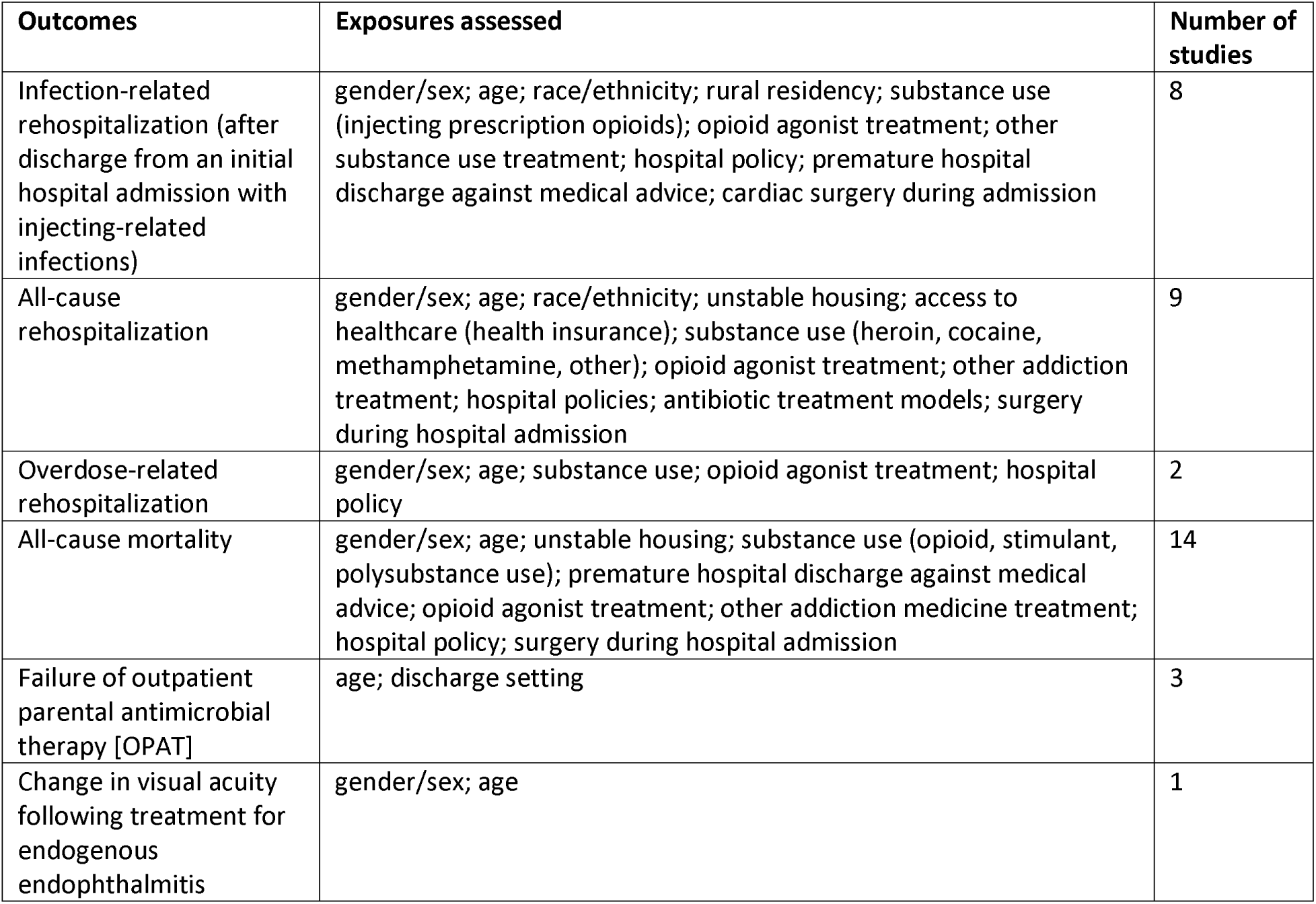
Summary of outcomes and associated exposures assessed among studies where outcome occurs after initial treatment for injecting-related infections.

Studies assessing outcomes after treatment also typically had imprecise effect estimates and inconsistent findings between studies, and opportunities for meta-analyses were limited.

See Appendix 19 for full results including several meta-analyses amongst studies in this section. Only two exposure-outcome associations were found to be significant in meta-analyses incorporating more than one study. Woman/female gender/sex was associated with increased risk of all-cause rehospitalization. Summary meta-analysis of three covariate-adjusted effect estimates was aOR 1.22 (95%CI 1.08-1.38; I^2^ 0%). One unadjusted effect estimate was nonsignificant at uOR 1.23 (95%CI 0.77-1.96). Inpatient addiction medicine consultation (during hospitalization with injecting-related infections) was associated with reduced risk of all-cause rehospitalization; summary of two unadjusted effect estimates was uOR 0.46 (95%CI 0.33-0.63; I^2^ 0%) and one fully-adjusted effect estimate was aOR 0.57 (95%CI 0.38–0.86).

### Colonisation with pathogenic bacteria

Five studies assessed factors associated with colonisation with *Staphylococcus aureus or* methicillin-resistant *S. aureus* among people who inject drugs: gender/sex; age; race/ethnicity; education; employment; relationship status; unstable housing and homelessness; incarceration; substance use (heroin, cocaine, crack, speedball, methamphetamines, prescription opioids; cannabis); public injecting; injecting in groups; opioid agonist treatment; other addiction treatment; recent hospital admission; and other (e.g. using public shower facilities). Several exposures had significant associations in single studies (e.g., public injecting, injecting frequently with three or more people, sleeping at more than one place during the prior week), but none were significant in meta-analyses of multiple studies. See Appendix 20 for full results among studies on colonization.

## Discussion

In this systematic review and meta-analysis, we identified 107 studies that assessed social determinants, substance use, and health services factors in relation to injecting-related bacterial infections and treatment outcomes. Several individual-level injecting risk practices (i.e., intramuscular or subcutaneous injecting, more frequent injecting, and lack of skin cleaning) were already known to be risk factors for these infections,^1^ and we were interested in the social contextual factors that can influence injecting practices and treatment experiences. In meta-analyses, we found evidence that risk for injecting-related infections was increased with woman/female gender/sex, less education, a history of incarceration, sex work, unstable housing and homelessness, heroin use, cocaine use, public injecting, and requiring or receiving injecting assistance. Among harm reduction and drug treatment factors, opioid agonist treatment was associated with a modest reduction in risk (i.e., ∼8% lower odds). Overall, there were many more studies where the outcome was incident or prevalent injecting-related infections than there were studies assessing health outcomes occurring during or after infection treatment (e.g., premature hospital discharge; all-cause mortality). Most studies that focused on outcomes occurring during or after infection treatment had small sample sizes and imprecise effect estimates, and most exposures assessed in this setting were only addressed in one study (so could not be meta-analysed). While this review incorporated a broad scope, there was insufficient evidence (with imprecise effect estimates) for many potential exposures, and interpreting meta-analyses was limited by high clinical and statistical heterogeneity. Nevertheless, the importance of social-structural factors on the risk of injecting-related infections and their treatment suggests that future approaches to improving prevention and treatment should look more broadly than individual-level injecting practices and engage with the social and material conditions within which people live, acquire drugs, consume them, and access health care.

The findings of this quantitative systematic review and meta-analysis complement a qualitative systematic review and thematic synthesis, published recently by our group.^10^ In the qualitative review, we identified several potential mechanisms through which social-structural factors could influence risk for injecting-related infections and poor treatment outcomes, including unregulated drug quality (e.g., poorly soluble drugs or adulterants contributing to skin and vein damage), insufficient housing (e.g. people not having access to running water to prepare drugs, or adequate lighting to find a vein), criminalization and enforcement (e.g., people compromising their drug preparation practices and rushing to inject their drugs intramuscularly to avoid police search and seizure), and operational limitations on harm reduction services (e.g. insufficient funding, or geographic restrictions). We also identified that harmful health care policies and practices lead to negative experiences of undertreated pain and withdrawal that discourage people from accessing care until infections had worsened and spread, or otherwise contribute to people leaving hospital prematurely against medical advice (before their treatment is complete). In the quantitative meta-analyses reported here, we identified consistent evidence of population-level effects for some of those exposures, but not for others. For example, here we identified evidence to support an association between incident or prevalent injecting-related infections with unstable housing or homelessness and injecting in public. A history of police contacts and arrests may have been associated with risk for infections (aOR 1.19, 95%CI 0.61-2.31), with imprecise confidence intervals that could include meaningful differences. However, this is measuring a different phenomenon than a concurrent police encounter contributing to a specific abscess for an individual.

In the qualitative review, participants highlighted the key role of harm reduction programs, like sufficient needle and syringe program coverage and access to supervised consumption sites, in enabling their ability to reduce risks for infection. Here, we did not identify evidence to support risk reduction with use of needle and syringe programs (aOR 0.75, 95%CI 0.54-1.03) or supervised consumption sites (aOR 0.59, 95%CI 0.29-1.19). Pooled effect estimates were imprecise and may include clinically meaningful reductions (or increases) in risk. These statistics from (mostly) cross-sectional observational studies may also show that needle and syringe programs and supervised consumption sites are successfully engaging people at highest risk of bacterial infections. This same phenomenon was observed in early research on HIV infection among people accessing needle and syringe programs.^51–53^ In addition, there were two ecological studies (that we could not include in meta-analyses) showing a reduction in injecting-related infections after people started accessing needle and syringe programs. A study published after our search found that people who regularly attended supervised consumption sites in France were less likely to report a recent abscess (18% vs. 22%).^54^ We identified that use of opioid agonist treatment was associated with a modest reduction in risk for injecting-related infections (aOR 0.92, 95%CI 0.89-0.95). Three studies on opioid agonist treatment and incident injecting-related infections, published after our search, estimated similar effects.^3,29,55^

In the context of the social determinants of health (and the closely related concepts of structural vulnerability and structural violence), we were interested in social identities and locations within societal power hierarchies which may enable or constrain the ability of people who inject drugs to prevent injecting-related infections and/or access treatment.^33,34^ We identified evidence to support associations between some sociodemographic characteristics and risks of injecting-related bacterial infections, including woman/female gender/sex, lower educational attainment, lower income/unemployment, incarceration, and sex work. We did not identify evidence to support associations with other characteristics, including by race/ethnicity. We conceptualized race as a proxy measure for the effects of structural racism^42,43^, and the absence of evidence identified here does not necessarily mean that racism is not an important determinant of injecting-related infections or treatment outcomes. While many studies considered sociodemographic characteristics as covariates in regression models (e.g., 17 studies included effect estimates for race/ethnicity on incident/prevalent infections), few studies were designed specifically to model the effect of these exposures (e.g., only one study modelled race/ethnicity as a primary exposure). In studies that did not specify a main exposure or estimand (instead considering all available variables in stepwise regressions), the effect of “upstream” exposures (e.g., structural racism) may be inappropriately blocked or hidden by conditioning on potential mediating variables (e.g., income, employment, or housing status that may be patterned by structural racism).^42,43^

Women may face excess risks of bacterial infections in the context of gendered power dynamics, for example that would lead them to “go second” and reuse contaminated equipment when injecting with male partners.^33,56,57^ Women may be less likely to know how to inject themself, and more likely to rely on assisted-injecting (which could reduce risks of intramuscular injection and abscesses in some people, but was associated with increased risks of bacterial infections in this review).^58,59^ Women may also be less likely to engage with harm reduction programs (which are more likely to have been designed for men); very few harm reduction programs (e.g., supervised consumptions sites) are gender-attentive or gender-specific.^33,60–62^ Some investigators have also hypothesized that excess risk of infections among women is attributable to deeper peripheral veins, due to different distributions of adipose tissue (and so women may have more difficulty accessing veins and may be more likely to inject in subcutaneous tissue).^6^ These differing risks are reflected in the greater proportion of woman/females in studies during and after treatment of injecting-related infections compared to studies assessing risk of incident or prevalent infections. Fewer studies focused on outcomes during and after treatment, which led to inconsistent findings. For example, woman/female gender/sex appeared associated with higher risks of all-cause rehospitalization but not infection-related rehospitalization, and it is unclear why this would be the case.

We also found that several substances were associated with higher risks of injecting-related bacterial infections, this including frequent or any use (vs. less or no use) of injection heroin, cocaine, and amphetamines. Studies that compared “frequent” (typically “daily or more”) use to less use did not consistently find that more frequent use of these substances was associated with greater risks of infections. Several studies also assessed specific formulations of unregulated drugs. Use of tar heroin (compared to powder heroin) was associated with nearly eight-fold increased risk of injecting-related bacterial infections in a covariate-adjusted analysis in one study^63^, and two-fold increased risk in a second (ecological) study.^64^ This may be because tar heroin is less soluble (leading to more undissolved particulate matter that can damage veins) and also that tar heroin requires the addition of acidifiers to dissolve and prepare for injection (and overuse of acidifiers contributes to vein sclerosis).^6,65,66^ “Crack” cocaine (base) formulations also require the addition of acidifiers to prepare the drug solution for injecting (while powder cocaine hydrochloride does not), but use of both crack and powder cocaine were associated with increased risks in meta-analyses of unadjusted analyses. Other specific substances were associated with increased risk of injecting-related infections in individual studies, including of ethylphenidate (a novel synthetic stimulant, associated with high frequency of injecting). In research external to this review, investigators have hypothesized that the North American drug supply transition to fentanyl has driven increasing incidence of injecting-related infections, as fentanyl has a shorter half-life than heroin and is associated with more frequent injecting.^2,67,68^ We identified no studies directly assessing illicit fentanyl use and risks of infections; two studies published after our search found injecting-related infections to be more common among people who inject fentanyl.^69,70^ Xylazine in the North American unregulated drug supply has also recently emerged as a cause of unusual wounds and infections; we identified no studies on xylazine here and we are not aware of any existing studies quantifying this risk.^71,72^

### Limitations

This review has several important limitations. First, we only included studies where the outcomes were injecting-related infections or treatment outcomes, so we did not capture studies where the outcome was risky injecting practices (e.g., lack of skin cleaning) that are associated with infections.^73–75^ Second, the inclusion of many exposures and outcomes (and, potentially, meta-analyses of unadjusted and covariate-adjusted effect estimates for each exposure) could lead to false positive findings through simply random chance (the so-called “multiple comparisons problem”). However, we wanted to take as broad a scope as possible to identify potential social determinants. Third, summary effect estimates from meta-analyses were likely not entirely accurate for several reasons: (a) we could not incorporate “negative” or “null” effect estimates from several studies that reported no statistics (only that the exposure and outcome were “not associated”) or reported unadjusted associations but dropped the variable in stepwise approaches to multivariable regression; (b) we combined effect estimates from studies with high heterogeneity (with different exposure and outcome definitions, timelines, sampling strategies, inclusion criteria, and study settings) which was often reflected in high measures of between-study statistical heterogeneity (I^2^ values); (c) most studies did not specify a hypothesis or estimand (and most did not take a causal or prespecified approach to covariate selection), which meant that most estimates did not come from studies trying to model as accurate as an effect as possible. Fourth, the observational cohort, cross-sectional, and case-control studies included in this review rarely contributed to understanding of mechanisms by which specific exposures affect the risk of infections or other treatment outcomes. This is one of the strengths of the complementary qualitative review.^10^ Future research focused on specific exposures and potential interventions could incorporate mixed-methods and critical realist methods to improve understanding of how these risks come about.^38,51–53^ Fifth, our search did not include studies during the COVID-19 era, which may have changed the risk environment. For example, a study published after our search showed rapid decline in incidence of injecting-related infections coinciding with the implementation of COVID response measures in England, comprised of decreased social mixing and temporary private accommodations for all people sleeping outside and in congregate shelters (“Everyone In” initiative).^5^

### Conclusions

Injecting-related infections, their treatment, and subsequent outcomes are shaped by multiple social determinants, substance use, and health services-related factors. Public health and clinical approaches to prevention and treatment should look more broadly than individual injecting practices, towards addressing the social and material conditions within which people live, acquire and consume drugs, and access health care.

## Data Availability

All data produced in the present work are contained in the manuscript and supplementary files

## Acknowledgments

We acknowledge that T.D.B., M.B., E.C. and I.K. live and work in Mi’kma’ki, the ancestral and unceded territory of the Mi’kmaq, and D.W. lives and works in unsurrendered and unceded territory and traditional lands of Wolastoqiyik. This territory is covered by the Treaties of Peace and Friendship which the Mi’kmaq and Wolastoqiyik Peoples first signed with the British Crown in 1725. The treaties did not deal with surrender of lands and resources but, in fact, recognized Mi’kmaq and Wolastoqiyik title and established the rules for what was to be an ongoing relationship between nations. We are Treaty people.

We thank Louise Gillis (Research Data Librarian at Dalhousie University) for helpful feedback and assistance with our search strategy. We thank Ricky Bluthenthal (University of Southern California) and Joseph Hayes (University College London) for helpful feedback on earlier drafts. We thank authors who provided correspondence and/or further information about their studies: Kasha Bornstein, Dan Ciccarone, Samantha Colledge-Frisby, Irina Ianache, Alison Ivey, Lisa Maher, David Marsh, Kristen Morin, Viktor Mravčík, Hélène Peyière.

