## Appendix for "Social determinants of injection drug use-associated bacterial infections and treatment outcomes: systematic review and meta-analysis"

### Table of Appendices

### Appendix 1. Search strategy used in mixed studies systematic review of injecting-related infections

| **Concepts** | **PubMed MEDLINE** | **EMBASE** | **Scopus** | **CINAHL** | **PsycINFO** |
| --- | --- | --- | --- | --- | --- |
| People who inject drugs, or drug preparation and injection | (“Substance-Related Disorders”[MeSH] OR | ('substance abuse'/exp OR | ( | ( |  |
|  | “Substance Abuse, Intravenous”[MeSH] OR | 'intravenous drug abuse'/exp OR |  |  |  |
|  | “Drug Users”[MeSH] OR | 'drug use'/exp OR |  |  |  |
|  | “Needle Sharing”[MeSH] OR | 'needle sharing'/exp OR |  |  |  |
|  | “people who inject drugs”[tiab] OR “persons who inject drugs”[tiab] OR PWID[tiab] OR | “people who inject drugs”:ab,ti OR “persons who inject drugs”:ab,ti OR PWID:ab,ti OR | TITLE-ABS("people who inject drugs") OR TITLE-ABS("persons who inject drugs") OR TITLE-ABS("PWID") OR | TI("people who inject drugs" OR "persons who inject drugs" OR "PWID") OR AB("people who inject drugs" OR "persons who inject drugs" OR "PWID") OR | TI("people who inject drugs" OR "persons who inject drugs" OR "PWID") OR AB("people who inject drugs" OR "persons who inject drugs" OR "PWID") OR |
|  | “people who use drugs”[tiab] OR “persons who use drugs”[tiab] OR PWUD[tiab] OR | “people who use drugs”:ab,ti OR “persons who use drugs”:ab,ti OR PWUD:ab,ti OR | TITLE-ABS("people who use drugs") OR TITLE-ABS("persons who use drugs") OR TITLE-ABS("PWUD") OR | TI("people who use drugs" OR "persons who use drugs" OR "PWUD") OR AB("people who use drugs" OR "persons who use drugs" OR "PWUD") OR | TI("people who use drugs" OR "persons who use drugs" OR "PWUD") OR AB("people who use drugs" OR "persons who use drugs" OR "PWUD") OR |
|  | “injection drug”[tiab] OR IDU[tiab] OR | “injection drug”:ab,ti OR IDU:ab,ti OR | TITLE-ABS("injection drug") OR TITLE-ABS("IDU") OR | TI("injection drug" OR "IDU") OR AB("injection drug" OR "IDU") OR | TI("injection drug" OR "IDU") OR AB("injection drug" OR "IDU") OR |
|  | “intravenous drug”[tiab] OR IVDU[tiab] OR | “intravenous drug”:ab,ti OR IVDU:ab,ti OR | TITLE-ABS("intravenous drug") OR TITLE-ABS("IVDU") OR | TI("intravenous drug" OR "IVDU") OR AB("intravenous drug" OR "IVDU") OR | TI("intravenous drug" OR "IVDU") OR AB("intravenous drug" OR "IVDU") OR |
|  | “drug abuse”[tiab] OR | “drug abuse”:ab,ti OR | TITLE-ABS("drug abuse") OR | TI("drug abuse") OR AB("drug abuse") OR | TI("drug abuse") OR AB("drug abuse") OR |
|  | “illicit drugs”[MeSH] OR “illicit drug”[tiab] OR | 'illicit drug'/exp OR “illicit drugs”:ab,ti OR | TITLE-ABS("illicit drug") OR | TI("illicit drug") OR AB("illicit drug") OR | TI("illicit drug") OR AB("illicit drug") OR |
|  | “Heroin”[MeSH] OR Heroin[tiab] OR | “heroin”:ab,ti OR | TITLE-ABS("Heroin") OR | TI("Heroin") OR AB("Heroin") OR | TI("Heroin") OR AB("Heroin") OR |
|  | “Heroin Dependence”[MeSH] OR | 'heroin dependence'/exp OR |  |  |  |
|  | “Opiate use disorder”[tiab] OR “opioid use disorder”[tiab] OR “opiate dependence”[tiab] OR “opioid dependence”[tiab] OR “opiate abuse”[tiab] OR “opioid abuse”[tiab] OR | 'narcotic dependence'/exp OR “opioid use disorder”:ab,ti OR “opiate use disorder”:ab,ti OR | TITLE-ABS("Opiate use disorder") OR TITLE-ABS("opioid use disorder") OR TITLE-ABS("opiate dependence") OR TITLE-ABS("opioid dependence") OR TITLE-ABS("opiate abuse") OR TITLE-ABS("opioid abuse") OR | TI("Opiate use disorder" OR "opioid use disorder" OR "opiate dependence" OR "opioid dependence" OR "opiate abuse" OR "opioid abuse") OR AB("Opiate use disorder" OR "opioid use disorder" OR "opiate dependence" OR "opioid dependence" OR "opiate abuse" OR "opioid abuse") OR | TI("Opiate use disorder" OR "opioid use disorder" OR "opiate dependence" OR "opioid dependence" OR "opiate abuse" OR "opioid abuse") OR AB("Opiate use disorder" OR "opioid use disorder" OR "opiate dependence" OR "opioid dependence" OR "opiate abuse" OR "opioid abuse") OR |
|  | “Cocaine”[MeSH] OR cocaine[tiab] OR | 'cocaine'/exp OR 'cocaine dependence'/exp OR cocaine:ab,ti OR | TITLE-ABS("cocaine") OR | TI("cocaine") OR AB("cocaine") OR | TI("cocaine") OR AB("cocaine") OR |
|  | “Crack Cocaine”[MeSH] OR | “crack cocaine”:ab,ti OR | TITLE-ABS("crack cocaine") OR | TI("crack cocaine") OR AB("crack cocaine") OR | TI("crack cocaine") OR AB("crack cocaine") OR |
|  | “groin injecting”[tiab] OR “femoral injecting”[tiab] OR | “groin injecting”:ab,ti OR “femoral injecting”:ab,ti OR | TITLE-ABS("groin injecting") OR TITLE-ABS("femoral injecting") OR | TI("groin injecting" OR "femoral injecting") OR AB("groin injecting" OR "femoral injecting") OR | TI("groin injecting" OR "femoral injecting") OR AB("groin injecting" OR "femoral injecting") OR |
|  | “Harm Reduction”[MeSH] OR “harm reduction”[tiab] OR | 'harm reduction'/exp OR “harm reduction”:ab,ti OR | TITLE-ABS("harm reduction") OR | TI("harm reduction") OR AB("harm reduction") OR | TI("harm reduction") OR AB("harm reduction") OR |
|  | “Needle-Exchange Programs”[MeSH] OR |  |  |  |  |
|  | “needle exchange”[tiab] OR “syringe exchange”[tiab] OR “syringe services”[tiab] OR | “needle exchange”:ab,ti OR “syringe exchange”:ab,ti OR “syringe services”:ab,ti OR | TITLE-ABS("needle exchange") OR TITLE-ABS("syringe exchange") OR TITLE-ABS("syringe services") OR | TI("needle exchange" OR "syringe exchange") OR "syringe services") OR AB("needle exchange" OR "syringe exchange") OR "syringe services") OR | TI("needle exchange" OR "syringe exchange") OR "syringe services") OR AB("needle exchange" OR "syringe exchange") OR "syringe services") OR |
|  | acidifier*[tiab] OR | acidifier*:ab,ti OR | TITLE-ABS("acidifier*") OR | TI("acidifier*") OR AB("acidifier*") OR | TI("acidifier*") OR AB("acidifier*") OR |
|  | “Opiate Substitution Treatment”[MeSH] OR ((“opiate substitution” OR “opiate agonist” OR “opioid substitution” OR “opioid agonist”) AND (treatment or therapy)) OR | 'opiate substitution treatment'/exp OR “opioid agonist”:ab,ti OR “opiate substitution”:ab,ti OR | TITLE-ABS("opiate substitution") OR TITLE-ABS("opiate agonist") OR TITLE-ABS("opioid substitution") OR TITLE-ABS("opioid agonist") OR | TI("opiate substitution") OR AB(“opiate substitution”) OR | TI("opiate substitution") OR AB(“opiate substitution”) OR |
|  | “Medications for opioid use disorder”[tiab] OR MOUD[tiab] OR | “medications for opioid use disorder”:ab,ti OR MOUD:ab,ti OR | TITLE-ABS("Medications for opioid use disorder") OR TITLE-ABS("MOUD") OR | TI("Medications for opioid use disorder" OR “MOUD) OR AB("Medications for opioid use disorder" OR "MOUD") OR | TI("Medications for opioid use disorder" OR “MOUD) OR AB("Medications for opioid use disorder" OR "MOUD") OR |
|  | Methadone[tiab] OR | 'methadone treatment'/exp OR methadone:ab,ti OR | TITLE-ABS("Methadone") OR | TI("Methadone") OR AB("Methadone") OR | TI("Methadone") OR AB("Methadone") OR |
|  | Buprenorphine[tiab]) | Buprenorphine:ab,ti) | TITLE-ABS("Buprenorphine")) | TI("Buprenorphine") OR AB("Buprenorphine")) | TI("Buprenorphine") OR AB("Buprenorphine")) |
| Injecting-related infections | AND | AND | AND | AND | AND |
|  | (“injection-related infections”[tiab] OR “injection-related infection”[tiab] OR | ('injection site abscess'/exp OR “injection-related infections”:ab,ti OR “injection-related infection”:ab,ti OR | ((TITLE-ABS("injection-related infections") OR TITLE-ABS("injection-related infection") OR | (TI("injection-related infections" OR "injection-related infection") OR AB("injection-related infections" OR "injection-related infection") OR | (TI("injection-related infections" OR "injection-related infection") OR AB("injection-related infections" OR "injection-related infection") OR |
|  | “bacterial infection”[tiab] OR “bacterial infections”[tiab] OR | “bacterial infection”:ab,ti OR | TITLE-ABS("bacterial infection") OR TITLE-ABS("bacterial infections") OR | TI("bacterial infection" OR "bacterial infections") OR AB("bacterial infection" OR "bacterial infections") OR | TI("bacterial infection" OR "bacterial infections") OR AB("bacterial infection" OR "bacterial infections") OR |
|  | Bacteremia[MeSH] OR bacteremia[tiab] OR | 'bacteremia'/exp OR bacteremia:ab,ti OR | TITLE-ABS("bacteremia") OR | TI("bacteremia") OR AB("bacteremia") OR | TI("bacteremia") OR AB("bacteremia") OR |
|  | Fungemia[MeSH] OR | 'fungemia'/exp OR |  |  |  |
|  | Cellulitis[MeSH] OR cellulitis[tiab] OR | 'cellulitis'/exp OR cellulitis:ab,ti OR | TITLE-ABS("cellulitis") OR | TI("cellulitis") OR AB("cellulitis") OR | TI("cellulitis") OR AB("cellulitis") OR |
|  | Abscess[MeSH] OR abscess*[tiab] OR | 'abscess'/exp OR abscess*:ab,ti OR | TITLE-ABS("abscess*") OR | TI("abscess*") OR AB("abscess*") OR | TI("abscess*") OR AB("abscess*") OR |
|  | “skin infection”[tiab] OR “skin infections”[tiab] OR | “skin infection”:ab,ti OR “skin infections”:ab,ti OR | TITLE-ABS("skin infection") OR TITLE-ABS("skin infections") OR | TI("skin infection" OR "skin infections") OR AB("skin infection" OR "skin infections") OR | TI("skin infection" OR "skin infections") OR AB("skin infection" OR "skin infections") OR |
|  | “skin and soft tissue”[tiab] OR SSTI*[tiab] OR | “skin and soft tisuuse”:ab,ti OR SSTI*:ab,ti OR | TITLE-ABS("skin and soft tissue") OR TITLE-ABS("SSTI*") OR | TI("skin and soft tissue" OR "SSTI*") OR AB("skin and soft tissue" OR "SSTI*") OR | TI("skin and soft tissue" OR "SSTI*") OR AB("skin and soft tissue" OR "SSTI*") OR |
|  | Endocarditis[MeSH] OR endocarditis[tiab] OR | 'endocarditis'/exp OR endocarditis:ab,ti OR | TITLE-ABS("endocarditis") OR | TI(endocarditis) OR AB(endocarditis) OR | TI(endocarditis) OR AB(endocarditis) OR |
|  | Bone Diseases, Infectious[MeSH] OR |  |  |  |  |
|  | Osteomyelitis[MeSH] OR Osteomyelitis[tiab] OR | 'osteomyelitis'/exp OR osteomyelitis:ab,ti OR | TITLE-ABS("osteomyelitis") OR | TI(“osteomyelitis") OR AB(“osteomyelitis") OR | TI(“osteomyelitis") OR AB(“osteomyelitis") OR |
|  | “septic arthritis”[tiab] OR | “septic arthritis”:ab,ti OR | TITLE-ABS("septic arthritis")) | TI("septic arthritis") OR AB("septic arthritis")) | TI("septic arthritis") OR AB("septic arthritis")) |
|  | Central Nervous System Infections[MeSH] OR |  |  |  |  |
|  | Gram-Positive Bacterial Infections[MeSH] OR |  |  |  |  |
|  | Candidiasis[MeSH]) | 'candidiasis'/exp) |  |  |  |
| Social and structural determinants, or risk environment | AND | AND | AND | AND | AND |
|  | ("risk factor"[tiab] OR “risk factors”[tiab] OR | ('risk factor'/exp OR “risk factor”:ab,ti OR | (TITLE-ABS("risk factor") OR TITLE-ABS("risk factors") OR | (TI("risk factor” OR "risk factors") OR AB("risk factor” OR "risk factors") OR | (TI("risk factor” OR "risk factors") OR AB("risk factor” OR "risk factors") OR |
|  | correlate*[tiab] OR | correlate*:ab,ti OR | TITLE-ABS(correlate*) OR | TI("correlate*") OR AB("correlate*") OR | TI("correlate*") OR AB("correlate*") OR |
|  | determinant*[tiab] OR | determinant*:ab,ti OR | TITLE-ABS("determinant*") OR | TI("determinant*") OR AB("determinant*") OR | TI("determinant*") OR AB("determinant*") OR |
|  | environment*[tiab] OR | environment*:ab,ti OR | TITLE-ABS("environment*") OR | TI("environment*") OR AB("environment*") OR | TI("environment*") OR AB("environment*") OR |
|  | “social factors”[tiab] or “structural factors”[tiab] OR | 'social determinants of health'/exp OR “social factors”:ab,ti OR “structural factors”:ab,ti OR | TITLE-ABS("social factors") OR TITLE-ABS("structural factors") OR | TI("social factors" OR "structural factors") OR AB("social factors" OR "structural factors") OR | TI("social factors" OR "structural factors") OR AB("social factors" OR "structural factors") OR |
|  | Cohort*[tiab] OR | 'cohort analysis'/exp OR cohort*:ab,ti OR | TITLE-ABS(cohort*) OR | TI("Cohort*") OR AB("Cohort*") OR | TI("Cohort*") OR AB("Cohort*") OR |
|  | Longitudinal[tiab] OR | Longitudinal:ab,ti OR | TITLE-ABS("Longitudinal") OR | TI("Longitudinal") OR AB("Longitudinal") OR | TI("Longitudinal") OR AB("Longitudinal") OR |
|  | Prospective[tiab] OR retrospective[tiab] OR | Prospective:ab,ti OR retrospective:ab,ti OR | TITLE-ABS("Prospective") OR TITLE-ABS("retrospective") OR | TI("Prospective" OR "retrospective") OR AB("Prospective" OR "retrospective") OR | TI("Prospective" OR "retrospective") OR AB("Prospective" OR "retrospective") OR |
|  | Randomized[tiab] OR randomised[tiab] OR | Randomized:ab,ti OR randomised:ab,ti OR | TITLE-ABS("Randomized") OR TITLE-ABS("randomised") OR | TI("Randomized" OR "randomised") OR AB("Randomized" OR "randomised") OR | TI("Randomized" OR "randomised") OR AB("Randomized" OR "randomised") OR |
|  | Comparative[tiab] OR | Comparative:ab,ti OR | TITLE-ABS("Comparative") OR | TI("Comparative") OR AB("Comparative") OR | TI("Comparative") OR AB("Comparative") OR |
|  | Case-control[tiab] OR | Case-control:ab,ti OR | TITLE-ABS("Case-control") OR | TI("Case-control") OR AB("Case-control") OR | TI("Case-control") OR AB("Case-control") OR |
|  | Time-series[tiab] OR | 'time series analysis'/exp OR “time-series”:ab,ti OR | TITLE-ABS("Time-series") OR | TI("Time-series") OR AB("Time-series") OR | TI("Time-series") OR AB("Time-series") OR |
|  | Survey*[tiab] OR | Survey*:ab,ti OR | TITLE-ABS("Survey*") OR | TI("Survey*") OR AB("Survey*") OR | TI("Survey*") OR AB("Survey*") OR |
|  | Epidemiolog*[tiab] OR | Epidemiolog*:ab,ti OR | TITLE-ABS("Epidemiolog*") OR | TI("Epidemiolog*") OR AB("Epidemiolog*") OR | TI("Epidemiolog*") OR AB("Epidemiolog*") OR |
|  | Qualitative[tiab] OR | Qualitative:ab,ti OR | TITLE-ABS("Qualitative") OR | TI("Qualitative") OR AB("Qualitative") OR | TI("Qualitative") OR AB("Qualitative") OR |
|  | Interview[tiab] OR | 'interview'/exp OR interview:ab,ti OR | TITLE-ABS("Interview") OR | TI("Interview") OR AB("Interview") OR | TI("Interview") OR AB("Interview") OR |
|  | Ethnograph*[tiab] OR | 'ethnography'/exp OR ethnograph*:ab,ti OR | TITLE-ABS("Ethnograph*") OR | TI("Ethnograph*") OR AB("Ethnograph*") OR | TI("Ethnograph*") OR AB("Ethnograph*") OR |
|  | Mixed-methods[tiab] OR “mixed methods”[tiab] | Mixed-methods:ab,ti OR “mixed methods”:ab,ti OR | TITLE-ABS("Mixed-methods") OR TITLE-ABS("mixed methods") OR | TI("Mixed-methods" OR "mixed methods") OR AB("Mixed-methods" OR "mixed methods") OR | TI("Mixed-methods" OR "mixed methods") OR AB("Mixed-methods" OR "mixed methods") OR |
|  | gender[tiab] OR | Gender:ab,ti OR | TITLE-ABS(gender) OR | TI(gender) OR AB(gender) OR | TI(gender) OR AB(gender) OR |
|  | homeless*[tiab] OR | homeless*:ab,ti OR | TITLE-ABS(homeless*) OR | TI(homeless*) OR AB(homeless*) OR | TI(homeless*) OR AB(homeless*) OR |
|  | race[tiab] OR racism[tiab] OR | race:ab,ti OR racism;ab,ti OR | TITLE-ABS(race OR racism) OR | TI(race or racism) OR AB(race or racism) OR | TI(race or racism) OR AB(race or racism) OR |
|  | incarcerat*[tiab] OR prison*[tiab] OR criminal*[tiab] OR | incarcerat*:ab,ti OR prison*:ab,ti OR criminal*ab,ti OR | TITLE-ABS(incarcerat* OR prison* OR criminal*) OR | TI(incarcerat* OR prison* OR criminal*) OR AB(incarcerat* OR prison* OR criminal*) OR | TI(incarcerat* OR prison* OR criminal*) OR AB(incarcerat* OR prison* OR criminal*) OR |
|  | stigma*[tiab] OR discrimination[tiab] OR exclusion[tiab]) | stigma*ab,ti OR discrimination:ab,ti OR exclusion:ab,ti) | TITLE-ABS(stigma* OR discrimination OR exclusion*)) | TI(stigma* OR discrimination OR exclusion) OR AB(stigma* OR discrimination OR exclusion)) | TI(stigma* OR discrimination OR exclusion) OR AB(stigma* OR discrimination OR exclusion)) |
|  | *NOT (“case report”[Title]) NOT (“case series”[Title])* | NOT “case report”:ti NOT “case series”:ti NOT “rare case”:ti |  |  |  |
|  | Filter: 2000-Present | Filter:2000-2021 | Filter:2000-2021 | Limit to 2000-2021 | Limit to 2000-2021 |
|  |  | Filter: AND (**'article'**/it OR **'article in press'**/it OR **'conference abstract'**/it OR **'conference paper'**/it OR **'letter'**/it OR **'short survey'**/it)  [**To remove review articles] |  |  |  |

### Appendix 2. Information on eight studies identified and included from outside of search

### Appendix 3. Studies excluded after critical appraisal.

Four potentially eligible studies that did not meet the MMAT screening questions and were therefore excluded. These were, “1. Are there clear research questions?” and “2. S2. Do the collected data allow to address the research questions?”

1. Annie FH, Bates MC, Uejio CK, Bhagat A, Kochar T, Embrey S, Uejio CK. The impact of the drug epidemic on the incidence of sepsis in West Virginia. Cureus. 2018 Oct 30;10(10).
   - No to S2: unclear unit of analysis, unclear outcomes.
2. Bates MC, Annie F, Jha A, Kerns F. Increasing incidence of IV‐drug use associated endocarditis in southern West Virginia and potential economic impact. Clinical Cardiology. 2019 Apr;42(4):432-7.
   - No to S1 and S2: unclear unit of analysis, unclear outcomes
3. Jafari S, Joe R, Elliot D, Nagji A, Hayden S, Marsh DC. A community care model of intravenous antibiotic therapy for injection drug users with deep tissue infection for “reduce leaving against medical advice”. International Journal of Mental Health and Addiction. 2015 Feb;13:49-58.
   - No to S2: Exposure is listed as community care vs. hospital, but entire sample was in the hospital.
4. Thakarar K, Rokas KE, Lucas FL, Powers S, Andrews E, DeMatteo C, Mooney D, Sorg MH, Valenti A, Cohen M. Mortality, morbidity, and cardiac surgery in Injection Drug Use (IDU)-associated versus non-IDU infective endocarditis: The need to expand substance use disorder treatment and harm reduction services. PLoS One. 2019 Nov 26;14(11):e0225460.
   - No to S2: Most of the study describes sample of patients with injection drug use-associated endocarditis and compares to people with endocarditis with no drug use. There is a subgroup analysis comparing outcomes among people with injection drug use-associated endocarditis who were prescribed opioid agonist treatment at hospital discharge vs. were not, but the proportions seem inadvertently reported as proportions of the whole sample so we cannot extract denominators.

### Appendix 4. Characteristics of included studies with outcome as incident or prevalent injecting-related infection in quantitative systematic review of social and structural determinants of injection drug use-associated bacterial and fungal infections.

| **Study** | **Included exposures in this review** | **Main exposure / estimand in study** | **Do exposures included in this review reflect study estimand** | **Infections** | **Outcomes** | **Country (City)** | **Sample size** | **Sampling method (parent study name)** | **MMAT quality rating (out of 5)** | **Data collection period** | **Inclusion criteria** | **Women/female** | **Age** | **Drugs used by  ≥50%** |
| --- | --- | --- | --- | --- | --- | --- | --- | --- | --- | --- | --- | --- | --- | --- |
| Baltes 2020^1^ | - Gender/sex - Age - Race/ethnicity - Education - Housing - Health insurance - Heroin - Cocaine - Amphetamines - Prescription opioids - Other prescription drugs | Not specified. Aimed to identify factors associated with having had an SSTI in the past year. | No estimand | - SSTI | Self-reported skin and soft-tissue infections in past year, not otherwise defined | USA (Brown, Douglas, Eau Claire, La Crosse, Outagamie, and Marathon counties, Wisconsin) | 80 | Respondent-driven sampling, starting with rural clients of needle and syringe program | 3 | May to July 2019 | 15 years or older, injected drugs within 30 days prior to survey response, and resided in a rural community | 40% | 42.5% between ages 30 and 39 years | Methamphetamine |
| Bassetti 2002^2^ | - Opioid agonist treatment | Effect of enrolling in injectable opioid agonist treatment program on risk for injecting-related bacterial infections | Yes | - Multiple - SSTI - Bacteremia | Incidence per 100 patient-years of infections requiring hospitalization, identified via electronic medical records  Skin (abscesses, phlegmonous infections, erysipelas, ulcerations, and necrosis) and bloodstream | Switzerland (Basel) | 175 | Consecutive patients enrolling in an injection opioid agonist treatment program (heroin, methadone, or morphine) | 4 | 1 November 1991 to 31 October 1998 | All participants who underwent their first evaluation from 1 November 1994 through 31 January 1997  Eligibility criteria for the opiate program were residence in the canton of Basel City, age ≥20 years, a ≥2-year history of addiction to injected heroin, failure of previous addiction treatment, and social distress and/or health problems that resulted from injection drug use. | 30% | Mean 31.4 years (range, 21-53) | Heroin |
| Bertin 2020^3^ | - Opioid agonist treatment | Effect of different opioid agonist treatment options on risk for multiple outcomes (including bacterial infections) | Yes | - Multiple - Not specified | Hospital admissions for bacterial infections, identified using ICD-10 codes.   Specific infections not defined. | France (nationwide) | 31,687 | Nationwide administrative data of linked hospital and pharmacy records  (French Nationwide Healthcare Data System, which covers 98.8% of the French population) | 4 | 1 April 2012 to 31 December 2015 | All patients aged 15 years  or older who received opioid agonist treatment with methadone, buprenorphine, or morphine sulfate from a community pharmacy at least once between 1 April 2012 and 31 December 2014, with no dispensing during the 3 months prior to inclusion, with the aim to recruit only incident patients.  Excluded patients with cancer, receiving palliative care, or in treatment for chronic pain. | Morphine: 23.4% Buprenorphine: 21% Methadone: 28.2% | Mean (SD):   Morphine: 34.7 (8.7)  Buprenorphine: 34.5 (9.1) Methadone 33.5 (8.2) | Not reported |
| Betts 2016^4^ | - Gender/sex - Age - Education - Housing - Opioid agonist treatment | Effect of opioid agonist treatment on substance use and health outcomes | Partly (1 of 5 exposures) | - SSTI | Past-month abscess, self-report (not otherwise specified) | Australia (nationwide) | 2,677 | Purposive convenience sampling, recruited through needle and syringe programs and snowball sampling in each state’s capital city.  (Illicit Drug Reporting System; IRDS) | 4 | 2011 to 2013 | Eligible participants are aged 16 years or older, report injecting an illicit drug at least monthly in the 6 months prior to interview and report living in their recruitment city for 12 months prior to interview | 34.2% | 17-35: 35.8% (957) 36-45: 38.4% (1025) 46-71: 25.7% (686) | None  (e.g., 29.8% of participants reported weekly heroin use; 17.1% reported weekly crystal methamphetamine use) |
| Bhattacharya 2006^5^ | - Needle and syringe program | Effect of implementing a needle and syringe program on the rate of SSTI among PWID | Yes | - SSTI | Monthly prevalence of skin abscess, as observed by outreach workers (not otherwise specified) | India (Tiljala slum area, Kolkata, West Bengal) | 4,736 participants observed over 27 months (unknown number of participants with multiple observations) | Sampling method not described  (“Observations were made in a cohort of street IDUs… observed daily through peer outreach workers supervised by field supervisors”) | 2 | January 2000 to March 2002 | Not described | Not described | Not described | Not described |
| Binswanger 2000^6^ | - Gender/sex - Age - Other substance use - Housing - Needle and syringe program | Not specified | No estimand | - SSTI | Current “pain, swelling, redness, hardness under their skin, heat, pus, or oozing” at injection site, self-reported and confirmed via physical examination | USA (San Francisco, California) | 169 | Targeted and snowball sampling through outreach, health and social services, and word-of-mouth  (Urban Health Study) | 4 | May 1997 | Age >18 years and physical evidence of drug injection (e.g., track marks) or previous participation in the Urban Health Study. | 25% | 54% were aged 40-49 years | Heroin |
| Buchanan 2006^7^ | - Cocaine | Effect of “crack” cocaine injection (vs. injecting other drugs) on multiple outcomes | Yes, but timeline mismatch | - SSTI | Ever hard “abscess” (not otherwise specified), self-reported | USA (New Haven and Hartford, Connecticut; Springfield, Massachusetts) | 989 | Targeted sampling via street outreach  (Syringe Access, Use and Discard research project) | 4 | January 2000 to May 2002 | 18 years of age or older, not currently in drug treatment, resident in targeted neighborhood and injection drug use within the past 30 days; current use was confirmed by physical examination of site of injections | Recent crack cocaine injection: 19.0%  Lifetime/ever: 25.8%  Never: 29.8% | Mean age  Recent crack cocaine injection: 37.8  Lifetime/ever: 38.4  Never: 38.8 | Cocaine, heroin |
| Cedarbaum 2016^8^ | - Age | Effect of being under 30 years old (vs. being older) on multiple outcomes | Yes | - SSTI | Past year “abscess” (not otherwise specified), self-report | USA (Seattle, Washington) | 389 | Convenience sample of needle and syringe programs clients | 5 | July 2013 | Needle and syringe program clients; recent heroin use and who had adequate data recorded for the injection and age-related questions | 29.3%  (“Transgender” coded as separate category from “Male” or “Female” | Under 30 years: 32.9%  30 years and older: 67.1% | Heroin, methamphetamines, benzodiazepines |
| Ciccarone 2016^9^ | - Class - Heroin type | Effect of geographic area and heroin type on risk of SSTI | Partly (1 of 2 exposures) | - SSTI | Hospitalization for opioid injection-related skin and soft tissue infections | USA (nationwide) | Not reported | 20% stratified national random sample of United States Community Hospitals   (Nationwide Inpatient Sample) | 5 | 1993 to 2010 | Hospital admission with ICD-9 codes 681.1–682.9, were between the ages of 15 and 65 and did not have a diagnosis of diabetes type 1 or type 2. | Not reported | Not reported | Not reported |
| Cooper 2005^10^ | - Age - Race/ethnicity - Class - Police contacts/arrests | Effect of police crackdowns on risk of hospitalization with injecting-related infections | Partly (1 of 4 exposures) | - Multiple - SSTI - Endocarditis | Monthly rate of hospitalisation for abscess/cellulitis or endocarditis | USA (New York, New York) | 27 police precincts (2,727,000 population; unknown number of people who inject drugs) | Hospital records from New York State Statewide Planning and Research  Cooperative System (SPARCS) database, which covers all individuals  admitted to community-based hospital facilities within  NYC | 5 | 1995 to 1999 | Hospital discharge diagnosis codes for endocarditis, abscess, or cellulitis, and also between 18 and 64 years old without diabetes; had an accompanying illicit-drug-related co-diagnosis or procedure mentioned  in their medical record; infection was not iatrogenic | Not reported | Precinct age structure (mean, standard deviation)  0-17 years: 23.1% (7.8) 18-64 years: 64.7% (6.7) >65 years 12.2% (4.0) | Not reported |
| Dahlman 2015^11^ | - Gender/sex - Age - Heroin - Prescription opioids - Prescription stimulants - Other prescription drug | Not specified. Aimed to identify factors associated with having ever had an SSTI. | No estimand | - SSTI | Ever had “an abscess or symptoms of skin and soft tissue infection (redness, swelling, pain, pus)”, self-report  Participants were asked to distinguish between signs of infection, and irritation caused by extravasal injection | Sweden (Malmö) | 80 | Consecutive clients attending needle and syringe program | 4 | 2012 | Reporting current or previous injection drug use, age ≥ 20 years, consent to HIV testing.  Exclusion criteria were inability to understand the informed consent or perform the interview due to Swedish language difficulties, psychiatric disability, or intoxication. | 30% | Median (range) 44.5 (23–64) | None |
| Dahlman 2017^12^ | - Gender/sex - Age - Race/ethnicity - Heroin - Cocaine - Amphetamine - Speedball - Other prescription drug - Housing - Inject in public - Received injecting assistance | Not specified. Aimed to identify factors associated with recently having had an SSTI. | No estimand | - SSTI | Past-30 days “abscess or symptoms for skin and soft tissue infection (redness,  swelling, pain, pus)” at injection site, self-report | USA (San Francisco, California) | 201 | Recruited from community settings using targeted sampling methods | 4 | November 2011 through April 2014 | injection drug use in the past 30 days, being 18 years of age or older, and the  ability to provide informed consent | 22.9% | 18–29: 15.4%  30–44: 36.3%  45–54: 27.9%  55 and older: 20.4% | Cocaine, methamphetamine, heroin |
| DiGiorgio 2019^13^ | - Drug policy change | Effect of opioid prescribing prescription policy on risk of injecting-related epidural abscess | Yes | - Epidural abscess | Monthly rate of hospitalisation with injection drug use-associated spinal epidural abscess | USA (New Orleans, Louisiana) | 45 | Hospital patients admitted to tertiary care center with billing codes or imaging tests indicating epidural abscess | 2 | July 2013 through July 2018 | Recent “intravenous drug use” recorded in medical charts. Not otherwise specified. | 24% | Mean (range) 47.1 (25–71) | Not reported |
| Doran 2020^14^ (UAM) | - Gender/sex - Age - Migration - Sex work - Incarceration - Amphetamine - Housing | Not specified. Aimed to identify factors associated with risk of SSTI. | No estimand | - SSTI | Past year “swelling containing pus (abscess), sore or open wound” at injection site, self-report | England, Wales, and Northern Ireland (87% outside of London) | 2,874 | Recruited from clients attending needle and syringe programs and addiction treatment programs (Unlinked Anonymous Monitoring Survey) | 4 | 2017 and 2018 annual surveys | Age 18 and older, injected drugs in the past year. | 29% | 68% aged 35 years and older | Not reported. “Majority… report injecting heroin or a heroin/crack cocaine combination.” |
| Doran 2020^14^ (Care & Prevent) | - Gender/sex - Age - Race/ethnicity - Class - Amphetamine - Housing | Not specified. Aimed to identify factors associated with risk of SSTI. | No estimand | - SSTI | Ever had SSTI, self-report.  “Participants were provided with photographs of mild,  moderate and severe abscesses, cellulitis and leg ulcers to aid their  recall, ensure correct SSTI identification and provide a comparative measure to assess SSTI severity.” | England (London) | 455 | Recruited from clients of drug treatment programs, homeless hostels, and outreach services (Care and Prevent Study) | 4 | October 2017 through March 2019 | Aged 18 and older, ever injected drugs | 25% | 57% aged 35 years and older | Not reported. “Majority… report injecting heroin or a heroin/crack cocaine combination.” |
| Dunleavy 2017^15^ | - Gender/sex - Age - Incarceration - Other polysubstance - Housing - Needle and syringe program - Opioid agonist treatment - Alcohol | Effect of needle and syringe program uptake, and of opioid agonist treatment uptake, on risk of SSTI | Partly (2 of 8 exposures) | - SSTI | “In the last year, have you had a swelling containing pus (abscess), a  sore or open wound at an injection site?”, self-report | Scotland | 1,876 | Recruited from clients at pharmacies and needle and syringe programs (National Exchange Surveillance Initiative Scotland study) | 4 | February 2013 through February 2014 | Injected drugs in the past 6 months | 29% | Age (years)  25 or younger: 8% 26-30: 15% 31-35: 25% 36 or older: 52% | Heroin |
| Fink 2013^16^ | - Gender/sex - Age - Race/ethnicity - Education - Heroin - Cocaine - Amphetamine - Speedball - Housing - Needle and syringe program | Not specified. Aimed to identify factors associated with risk of SSTI. | No estimand | - SSTI | “During the last six months, did you have an  abscess related to injection drug use? (Including any enduring lumps, even if they did not “come to a head,” drain, or require treatment of any kind”, self-report | USA (Los Angeles, Oakland, and Berkeley, California) | 858 | Recruited through community outreach and clients of needle and syringe programs | 4 | 2003 through 2005 | Aged 18 years or older, and injected drugs in the past 30 days | 29% | Age (years)  29 or younger: 5% 30-39: 16%  40-49: 37%  50+: 41% | Heroin |
| Hope 2014^17^ | - Gender/sex - Age - Class - Incarceration - Overdose history - Heroin - Cocaine - Amphetamine - Housing - Police contacts/arrests | Not specified. Aimed to identify factors associated with SSTI. | No estimand | - SSTI | Past 28-days, “redness,  swelling and tenderness; an abscess (a swelling containing pus);  or a sore/open wound” at an injection site, self-report | England (Bristol, Leeds, and Birmingham) | 855 | Respondent-driven sampling, with seed participants identified via street outreach and key informant referrals | 5 | 2006 (Bristol), 2008 (Leeds), and 2009 (Birmingham) | Aged 15 and older, injected drugs in the past four weeks, living within survey area | 25% | Mean 32, median 31 years | Heroin, crack cocaine |
| Hope 2015^18^ | - Gender/sex - Age - Class - Migration - Sex work - Overdose - Heroin - Cocaine - Amphetamine - Housing - Police contacts/arrests | Not specified. Aimed to identify factors associated with SSTI. | No estimand | - SSTI | Past year, “redness, swelling and tenderness; an abscess; or a sore/open wound” at an injection site, self-report | England (Bristol, Leeds, and Birmingham) | 855 | Respondent-driven sampling, with seed participants identified via street outreach and key informant referrals  Same sample as Hope 2014 | 5 | 2006 (Bristol), 2008 (Leeds), and 2009 (Birmingham) | Aged 15 and older, injected drugs in the past four weeks, living within survey area | 25% | Mean 32, median 31 years | Heroin, crack cocaine |
| Hope 2010^19^ | - Gender/sex - Age - Incarceration - Other stimulant - Other polysubstance - Housing - Needle and syringe program - Opioid agonist treatment | Not specified. Aimed to identify factors associated with SSTI. | No estimand | - SSTI | Past year “swelling containing pus (abscess), sore, or  open wound at an injection site”,  self-report | England, Wales, and Northern Ireland | 5,209 | Recruited from clients attending needle and syringe programs and addiction treatment programs (Unlinked Anonymous Monitoring Survey) | 4 | 2006 through 2008 | Injected drugs in the past year. | 25% | Mean 32.5, median 32 years | Heroin, stimulants (not otherwise specified) |
| Hope 2008^20^ | - Gender/sex - Age - Incarceration - Cocaine - Amphetamine - Housing - Opioid agonist treatment | Not specified. Aimed to identify factors associated with SSTI. | No estimand | - SSTI | Past year “abscess (pus filled swelling)” or “open wound/sore” at injection site, self-reported | England, Wales, and Northern Ireland | 1,058 | Recruited from clients attending needle and syringe programs and addiction treatment programs (Unlinked Anonymous Monitoring Survey) | 4 | Autumn 2003 through summer 2005 | Injected drugs in past 28 days | 23% | Median 30 years, range 16 to 72 years | Opiates |
| Islam 2019^21^ | - Gender/sex - Age | Effect of reducing or stopping injection use on injecting-elated infections | No | - Multiple - Endocarditis - Sepsis/bacteremia - Pneumonia | Invasive bacterial infection (pneumonia, sepsis, endocarditis) at 9 months and 12 months following baseline study visit, self-report & confirmed with medical chart review  Conceptualized as injecting-related, despite including pneumonia | USA (Baltimore, Maryland) | 2,247 | Recruitment through community-based outreach and through clients of health and social services (AIDS Linked to the Intravenous Experience; ALIVE)  Similar sample as Safaeian 2000 and Wilson 2002 | 4 | December 1988 through June 2012 | Age 18 years and older, history of injection drug use and high-frequency injection drug use (defined as >1 time daily) | 27.8% | Not reported | Heroin, cocaine |
| Lee 2013^22^ | - Received injecting assistance | Not specified. Aimed to identify factors associated with receiving injecting assistance. | No estimand. | - SSTI | Past 6 month soft-tissue infections (not otherwise specified), self-reported | Thailand (Bangkok) | 430 | Recruited through community outreach (Mitsampan Community Research Project) | 5 | July 2011 to October 2011 | Adults, residing in Bangkok or adjacent provinces, injected drugs in past six months | 19.3% | Median 38 years (interquartile range 34 - 48 years) | Heroin, midazolam |
| Lewer 2020^23^ | - Gender/sex | Effect of gender/sex on injecting-related infections | Yes | - Multiple - SSTI - Sepsis/bacteremia - Endocarditis - Septic arthritis - Osteomyelitis | Rate of hospital admissions for heroin-injection associated bacterial infections | England (London) | 2,335 | Electronic health records of people entering community-based substance use treatment, with reported use of heroin and drug injection (Clinical Records Interactive Search resource at the South London and Maudsley NHS Foundation Trust Biomedical Research Centre) | 5 | 1 January 2006 and 31 March 2017 | Age 18-64 who were entering community-based substance use treatment, reported heroin use and drug injection | 26% | Mean 36.3 (SD 8.4) years | Heroin, crack cocaine, alcohol |
| Lloyd-Smith 2005^24^ | - Gender/sex - Sex work - Incarceration - Heroin - Cocaine - Housing | Not specified. Aimed to identify factors associated with SSTI. | No estimand | - SSTI | Past 6-months abscess (“lasting for more than 3 days”), self-report | Canada (Vancouver, British Columbia) | 1,585 | Recruitment through community-based outreach (Vancouver Injection Drug Users Study; VIDUS) | 5 | 1 May 1996 to May 31 2004 | Lived in Vancouver area, injected drugs in past month | 36% | Not reported | Not reported |
| Lloyd-Smith 2012^25^ | - Gender/sex - Age - Heroin - Cocaine - Speedball - Housing - Received injecting assistance | Not specified. Aimed to identify factors associated with SSTI. | No estimand | - SSTI | ED visit for cutaneous injecting-related infection, via administrative data | Canada (Vancouver, British Columbia) | 1,083 | Randomly recruited clients at supervised injecting facility  (Scientific Evaluation of Supervised Injection; SEOSI)  Similar sample to Lloyd-Smith 2008 and Milloy 2010, and same sample as Lloyd-Smith 2009 and Lloyd-Smith 2010 | 4 | 1 January 2004 to 31 January 2008 | Not reported | 29% | Median (interquartile range) was 35.1 (28.7 to 41.5) years among females and 39.7 (33.7 to 45.3) years among males | Not reported |
| Lloyd-Smith 2008^26^ | - Gender/sex - Age - Sex work - Heroin - Amphetamine - Housing - Received injecting assistance - Supervised consumption site | Not specified. Aimed to identify factors associated with SSTI. | No estimand | - SSTI | Current “any sores or abscesses from where you have been injecting”, self-report and confirmed by study nurse | Canada (Vancouver, British Columbia) | 1,065 | Randomly recruited clients at supervised injecting facility  (Scientific Evaluation of Supervised Injection; SEOSI)  Similar sample to Lloyd-Smith 2012, Lloyd-Smith 2009, Lloyd-Smith 2010, and Milloy 2010 | 4 | 1 January 2004 to 31 December 2005 | Age 19 years and older, performed at least two injections at the supervised injection facility | 29% | Median (interquartile range) was 36 (31-43) among participants with current SSTI at baseline interview and 39 (33-45) among participants without | Not reported |
| Lloyd-Smith 2009^27^ | - Gender/sex - Age - Heroin - Housing - Received injecting assistance - Cocaine - Speedball | Not specified. Aimed to identify factors associated with incidence of SSTIsupervised injecting facility. | No estimand | - SSTI | Current SSTI cared for at supervised injecting facility, from nursing notes | Canada (Vancouver, British Columbia) | 1,080 | Randomly recruited clients at supervised injecting facility  (Scientific Evaluation of Supervised Injection; SEOSI)  Similar sample to Lloyd-Smith 2008 and Milloy 2010, and same sample as Lloyd-Smith 2012 and Lloyd-Smith 2010 | 4 | 1 December 2003 and 31 January 2008 | Second visit to supervised injecting facility | 29% | Median (interquartile range) 38.4 (32.7 - 44.3) years | Not reported |
| Lloyd-Smith 2010^28^ | - Gender/sex - Age - Speedball - Housing | Not specified. Aimed to identify factors associated with SSTI hospitalization. | No estimand | - Multiple - SSTI - Osteomyelitis - Endocarditis - Septic arthritis | Hospitalization for injecting-related infection (cellulitis, abscess, osteomyelitis, Staph infection, endocarditis, septic arthritis, ulcer, thrombophlebitis, myositis), identified via administrative data | Canada (Vancouver, British Columbia) | 1,083 | Randomly recruited clients at supervised injecting facility  (Scientific Evaluation of Supervised Injection; SEOSI)  Similar sample to Lloyd-Smith 2008 and Milloy 2010, same sample as Lloyd-Smith 2009 and Lloyd-Smith 2012 | 4 | 1 January 2004 to 31 January 2008 | Not reported | 29% | Median (interquartile range), 38.4 (32.7 - 44.3) | Not reported |
| McMahan 2020^29^ | - Amphetamine | Not specified. Aimed to identify factors associated with interest in reducing/stopping substance use | No estimand | - Multiple - SSTI - Sepsis/bacteremia - Endocarditis | Past-year injecting-related infection (“an abscess, skin infection such as cellulitis, blood clot or blood infection like sepsis, or endocarditis”), self-report | USA (Washington state) | 583 | Attempted census of all needle and syringe program clients (Washington State Syringe Exchange Survey) | 4 | June 2019-August 2019 | Needle and syringe program clients who reported that methamphetamine or opioids were their main drug, and they were not currently receiving addiction treatment.  Excluded participants whose main drug was “goofball” (i.e., heroin and methamphetamine mixed together) | 45% | Median 35 (IQR 30-45) years | Heroin |
| Milloy 2010^30^ | - Gender/sex - Age - Race/ethnicity - Sex work - Incarceration - Heroin - Cocaine - Speedball - Housing - Inject in public - Supervised consumption site - Opioid agonist treatment | Effect of recent incarceration on risk of SSTI | Partly (1 of 12 exposures) | - SSTI | ED visit for abscess or cellulitis, identified via administrative codes | Canada (Vancouver, British Columbia) | 901 | Randomly recruited clients at supervised injecting facility  (Scientific Evaluation of Supervised Injection; SEOSI)  Similar sample to Lloyd-Smith 2008 and Lloyd-Smith 2010, and same sample as Lloyd-Smith 2009 and Lloyd-Smith 2012 | 4 | June 2004 to December 2006 | Not reported | 29.5% | Median (IQR) was 37.5 (32.8-42.3) years among people reporting recent ED visit for SSTI at baseline interview, and 39.9 (33.7-46.1) among people not reporting this | Not reported |
| Morin 2020^31^ | - Gender/sex - Age - Class - Opioid agonist treatment | Not specified. Aimed to identify factors associated with injecting-related infections. | No estimand | - Multiple - Endocarditis - Osteomyelitis - Septic arthritis | Diagnostic codes in medical records (outpatient/ambulatory, emergency department, or hospitalization). Timing unclear. | Canada (Ontario) | 55,924 | Patients with claims in public health insurance databases for (a) any billing code associated with opioid agonist treatment, or (b) opioid agonist treatment medications via drug identification numbers (not specified) | 3 | 1 January 2011 to 31 December 2016 | Age 15 years and older, resident in Ontario | 35% | 15 to 24 years: 18% 25 to 34 years: 34% 35 to 44 years: 22% 45 to 54 years: 18% 54 to 65 years: 7% 66 years and older: 2% | Not reported |
| Murphy 2001^32^ | - Gender/sex - Age - Race/ethnicity - Education - Class - Heroin - Cocaine - Amphetamine - Speedball - Alcohol - Smoking | Effect of HTLV-II infection on risk of SSTI. Also aimed to identify factors associated with SSTI. | No | - SSTI | ED visit or hospitalization for injecting-related abscess (case-control study) | USA (San Francisco, California) | 424 | Cases were hospital or ED patients with abscess who had injected drugs within the past 6 months. Sampling approach not specified.  Controls were hospital or ED patients who were also enrolled in a local community-based cohort study of people who inject drugs, matched 2:1 on age, sex, and race. Parent study not specified. | 3 | Not reported | Age 18 years and older, spoke English | 32% | 29 years and younger: 12%  30 to 39 years: 27% 40 to 49 years: 45% 50 years and older: 17% | Heroin, cocaine, speedball, amphetamine |
| Nagar 2015^33^ | - Drug policy change | Effect of opioid prescribing prescription policy on risk of injecting-related epidural abscess | Yes | - Epidural abscess | Annual frequency of hospitalisation with spinal epidural abscess and substance use | USA (Lexington, Kentucky) | 172 | Hospital patients admitted with billing codes indicating epidural abscess | 3 | 1 July 2010 to 30 June 2014 | Patients with ICD-9 codes indicating substance dependence, abuse, or withdrawal | Not reported | Not reported | Not reported |
| Noroozi 2019^34^ | - Age - Education - Class - Relationships - Amphetamine - Other polysubstance - Housing - Needle and syringe program | Not specified. Aimed to identify factors associated with SSTI. | No estimand | - SSTI | Participants were asked “Have you ever had skin infections (such abscess *[sic]* or cellulitis) at your injection sites?” | Iran (Tehran) | 500 | Convenience and snowball sampling from drop-in centres | 4 | March 2016 to August 2016 | Age 18 years and older, injection drug use at least once during the last month. | 0% | Mean 31.2 (SD 7.2) years | Heroin, methamphetamine |
| Oviedo-Joekes 2017^35^ | - Opioid agonist treatment | Effect of hydromorphone vs. diacetylmorphine injectable opioid agonist treatment on risk of SSTI. | Yes | - SSTI | Cellulitis or abscess identified during clinical trial via MedDRA codes | Canada (Vancouver, British Columbia) | 202 | Not reported (Study to Assess Longer-term Opioid Medication Effectiveness; SALOME) | 5 | December 2011 to December 2013 | Age 19 years and older; at least 5 years of “illicit opioid dependence”, regular injection of illicit opioids in the prior year; at least one prior episode of opioid agonist treatment; no severe medical conditions contraindicated for treatment with diacetylmorphine or hydromorphone | 30.7% | Mean 44.3 (SD 9.63) years | Not reported |
| Phillips 2017^36^ | - Gender/sex - Age - Race/ethnicity - Education - Cocaine - Housing - Alcohol | Not specified. Aimed to identify factors associated with SSTI. | No estimand | - SSTI | Past year “skin abscesses  (defined as red, hard infected lumps that contain pockets of  pus), ulcers (defined as open, infected sores), or cellulitis (defined as a more widespread skin infection)”, self-reported | USA (Boston, Massachusetts) | 143 | Consecutive inpatients in medical units at an academic hospital | 4 | January 2014 to October 2015 | Age 18 years and older; injected drugs at least 3 days during the week before hospital admission; without psychosis or homicidal/suicidal ideation | 40.6% | Mean 38.7 (SD 10.7) years | Heroin, cocaine |
| Phillips 2008^37^ | - Gender/sex - Age - Race/ethnicity - Heroin - Cocaine - Housing - Shooting gallery - Alcohol | Not specified. Aimed to identify factors associated with injecting-related infections. | No estimand | - SSTI - Osteomyelitis - Endocarditis | ED visit or hospitalization for skin abscess, cellulitis, osteomyelitis, or endocarditis in 6 months before study visit; self-report | USA (Providence, Rhode Island) | 109 | Recruited through placing fliers at community health and social services | 4 | April 2001 to December 2004 | Injection drug use; hepatitis C virus negative; heroin or cocaine use in past month; not experiencing psychotic symptoms | 25.7% | Mean 38.7 (SD 8.8) years | Not reported |
| Phillips 2010^38^ | - Age - Race/ethnicity - Heroin - Speedball - Housing | Not specified. Aimed to identify factors associated with SSTI | No estimand | - SSTI | Past year “abscess or other skin infection (such as an ulcer or cellulitis) at a place where you injected drugs—that is, any pain, swelling, redness, hardness under your skin, heat, pus, or oozing anywhere you inject?”, self-reported | USA (Denver, Colorado) | 51 | Recruited through drop-in center, drug treatment center, and newspaper advertisement | 4 | November 2007 to August 2008 | Age 18 years and older; not experiencing psychotic symptoms; injection drug use in past month | 33.3% | Mean 39.2 (SD 9.7) years | Heroin |
| Pollini 2010^39^ | - Gender/sex - Age - Race/ethnicity - Class - Sex work - Incarceration - Heroin - Heroin type - Amphetamine - Speedball - Housing - Inject with others - Inject in public - Received injecting assistance - Police contacts/arrests - Shooting gallery - Smoking | Not specified. Aimed to identify factors associated with SSTI. | No estimand | - SSTI | Past 6-months abscess (“Have you ever had an abscess?’’, ‘‘When was the last time you had an  abscess?’’), self-report | Mexico (Tijuana, Baja California) | 623 | Respondent-driven sampling (El Cuete Phase III)  Similar sample to Pollini 2010b and Robertson 2010. Unclear overlap. | 5 | April 2006 to April 2007 | Age 18 years or older; injected drugs in past month; no plans to move out of the city in the next 18 months | 18% | Median 37 (IQR 32-43) | Heroin; Methamphetamine |
| Pollini 2010b^40^ | - Barriers to needle/syringe access | Effect of being refused/overcharged when trying to purchase syringes at pharmacy on SSTI. | Yes | - SSTI | Past 6-months abscess and lifetime history of abscess, self-report | Mexico (Tijuana, Baja California) | 649 | Respondent-driven sampling (El Cuete Phase III)  Similar sample to Pollini 2010a and Robertson 2010. Unclear overlap. | 5 | April 2006 to April 2007 | 18 years or older; injected drugs in the past month | 18% | Median 38 (IQR 33-44) years | Heroin; methamphetamine |
| Robertson 2010^41^ | - Received injecting assistance | Not specified. Aimed to identify factors associated with “seeking injection assistance”. | No estimand | - SSTI | Past 6-months abscess, self-report | Mexico (Tijuana, Baja California) | 1056 | Respondent-driven sampling (El Cuete Phase III)  Similar sample to Pollini 2010a and Pollini 2010b. Unclear overlap. | 5 | April 2006 to April 2007 | Age 18 years or older; injected drugs in past month; no plans to move out of the city in the next 18 months | 15% | Median 37 (IQR 31-42) years | Heroin; otherwise not reported |
| Roux 2020^42^ | - Gender/sex - Age - Class - Relationships - Heroin - Other polysubstance - Prescription opioids - Housing - Inject with others - Inject in public - Received injecting assistance - Opioid agonist treatment - Alcohol | Effect of an educational intervention on risk of SSTI | No | - SSTI | At least one cutaneous abscess in the previous six months, self-reported | Bulgaria; Greece; Portugal; Romania | 307 | Recruited from clients of harm reduction programs | 4 | 1 December 2017 to 30 November 2019 | Age 18 years or older; injected drugs during the previous week | 17% | Median 38 (IQR 34-43) years | Heroin |
| Saeland 2014^43^ | - Age - Education - Sex work - Incarceration - Food insecurity - Overdose history - Heroin - Other prescription drug - Smoking | Effect of malnutrition on risk of SSTI | Partly (1 of 9 exposures) | - SSTI | Current abscess. Self-report and confirmed by physical examination. | Norway (Oslo) | 188 | Recruited via street outreach and through health and social services | 3 | November 2001 to April 2003 | Not reported | Not reported | With abscess: Mean 36.9 (SD 7.7)  Without abscess: Mean 35.1 (SD 7.6) | Heroin; flunitrazepam |
| Safaeian 2000^44^ | - Gender/sex - Age - Race/ethnicity - Education - Class - Alcohol - Smoking | Effect of HTLV-II virus infection on risk of SSTI and endocarditis. Also aimed to identify factors associated with SSI and endocarditis. | No | - SSTI - Endocarditis | Infective endocarditis, self-report confirmed through medical chart review  Abscess, self-report | USA (Baltimore, Maryland) | 86 cases with endocarditis and 567 controls  356 cases with abscess and 1436 controls  Unclear overlap | RecruIted through street outreach and snowball sampling (AIDS Link to Intravenous Experience; ALIVE)  Similar sample as Islam 2019 and Wilson 2002 | 4 | 1988 to 1982 | Age 18 years and older | Endocarditis analysis: 50%  Abscess analysis: 77% | Percentage older than 34 years  Endocarditis analysis: cases 48%, controls 45%  Abscess analysis: Cases 50%, controls 46% | Not reported |
| Scherbaum 2010^45^ | - Supervised consumption site | Effect of supervised consumption site attendance on risk of SSTI | Yes | - SSTI | “During the last month, did you visit a physician because of an abscess?”, self -reported | Germany (Essen, Ruhr zone) | 129 | Invited consecutive clients at supervised consumption site | 4 | November 2002 to December 21 2003 | New attendance (first time or at least 6 weeks since last visit) at supervised consumption site | 25% | Mean 31 (SD 6) years | Opiates; cocaine; cannabis; alcohol |
| Shah 2020^46^ | - Gender/sex - Age - Race/ethnicity - Education - Class - Heroin - Fentanyl - Cocaine - Amphetamine - Prescription opioids - Prescription stimulant - Housing | Effect of injecting hydromorphone controlled release formulation on risk of endocarditis. Also aimed to identify factors associated with endocarditis. | Partly (1 of 12 exposures) | - Endocarditis | Current hospital admission with diagnosis of “definite infective endocarditis” according to the Modified Duke Criteria | Canada (London, Ontario) | 135 (33 cases with endocarditis, 102 controls) | Cases were recruited from among inpatients or recently discharged outpatients with endocarditis among three hospitals  Controls were recruited from community-based health and social services, addiction treatment programs, and outpatient infectious diseases clinic  Matching approach not reported | 4 | 11 August 2016 to 27 July 2018 | Age 18 years or older; injected drugs within past 4 months | 27% | Cases: Mean 30.0 (SD 11.0) years  Controls: 35.5 (SD 8.4) years | Hydromorphone controlled-release capsules; methamphetamine; hydromorphone tablets |
| Sierra 2006^47^ | - Gender/sex - Age - Cocaine - Speedball - Supply network - Housing - Opioid agonist treatment | Not specified. Aimed to identify factors associated with SSTI (specifically, invasive S. pyogenes) | No estimand | - SSTI | Hospital admission with invasive soft-tissue Group A Strep (S. pyogenes) infections | Spain (Barcelona) | 73 (15 cases, 58 locally recruited controls) | Cases recruited from among hospitalized patients with S. pyogenes identified in bacterial cultures (sites not specified)  Controls recruited among clients of a local needle and syringe program who had attended the same hospital | 3 | Fall 2002 | Age 18 year and older, injected drugs (timeline not defined), did not live in an institution (not defined) | 26% | Cases: Mean 30.1 years  Controls: 27.5 years | Heroin; cocaine |
| Silverman 2020^48^ | - Prescription opioids | Effect of recently being prescribed controlled-release hydromorphone on risk of endocarditis | Yes | - Endocarditis | Hospital admission for endocarditis and injection drug use, identified via administrative codes | Canada (Ontario province) | 46,505 (for ecological analysis)  13,823 (for individual-level analysis) | Province-wide hospital and prescription administrative data | 5 | 1 April 2006 to 30 September 2015 | Age between 18 and 55 years old; at least one hospital admission with evidence of injection drug use; at least one opioid prescription through public insurance | Matched cohort for any hydromorphone exposure analysis: 43.9%  Matched cohort for controlled-release hydromorphone exposure analysis: 43.5% | Matched cohort for any hydromorphone exposure analysis: mean 44.4 (SD 8.4) years  Matched cohort for controlled-release hydromorphone exposure analysis: 44.6 (SD 8.4) years | Not reported. |
| Smith 2015^49^ | - Gender/sex - Race/ethnicity - Cocaine - Speedball - Housing - Inject with others | Not specified. Aimed to identify factors associated with SSTI. | No estimand | - SSTI | Current abscess, “defined as swollen, red, painful lumps under the skin that may or may not be open and that have lasted <8 weeks”. Self-reported and visually confirmed by researcher. | USA (Baltimore, Maryland) | 152 | Recruited from clients at Baltimore City Needle Exchange Program | 4 | May 2012 to November 2013 | Age 18 years or older | 36.8% | Median 45 (IQR 35-52) years | Heroin |
| Stein 2020^50^ | - Gender/sex - Age - Race/ethnicity - Opioid agonist treatment | Effect of an educational intervention on risk of injecting-related infections | No | - Multiple - SSTI - Endocarditis - Sepsis - Not specified | Number of ED visits for injecting-related infections in 12 months following educational intervention  Number of hospitalizations visits for injecting-related infections in 12 months following educational intervention | USA (Boston, Massachusetts) | 252 | Recruited from among hospital inpatients with “an indication” of current or past injection drug use or a current skin abscess or cellulitis in electronic medical record | 4 | January 2014 to August 2019 | Injected drugs at least 3 days out of the week prior to hospital admission; ability to return to hospital for follow-up; at least two additional contacts with active telephone numbers; no planned move from the region | 41.7% | Mean 37.9 (SD 10.7) years | Not reported |
| Summers 2017^51^ | - Age - Race/ethnicity - Class - Heroin type - Housing | Effect of tar heroin on risk of SSTI. Also aimed to identify factors associated with SSTI. | Partly (1 of 5 exposures) | - SSTI | Past year abscess, defined as, “a painful, hot, swollen skin infection with pus inside”, self-report | USA (Sacramento, California; Boston, Massachusetts) | 145 | Recruited from clients attending harm reduction programs | 4 | December 2014 to February 2015 | Age 18 years or older; self-reported heroin injection in preceding month | 29% | Mean 40 (95% CI 38.09, 41.90) | Heroin; otherwise not reported |
| Thønnings 2020^52^ | - Age - Housing - Opioid agonist treatment | Not specified. Aimed to identify factors associated with bacteremia | No estimand | - Sepsis/bacteremia | Bacteraemia, among hospitalised PWID | Denmark (Hvivdovre, Region Hovedstaden) | 257 | Retrospectively identified via hospital administrative codes | 3 | 2000 to 2006 | Not reported | Not reported | Median 39 (IQR 34-45) years | Not reported |
| Tomolillo 2007^53^ (Ecological time series study) | - Needle and syringe program | Effect of needle and syringe program use on risk of SSTI.  Effect of policy restricting needle and syringe program effectiveness on risk of SSTI | Yes | - SSTI | Number of abscesses treated at clinic associated with needle and syringe program, per week | USA (Eureka, California) | 2942 visits (participant count unknown) | Administrative data (number of needles exchanged) and health records (number of abscesses treated) | 4 | January 1, 2002 to February 28, 2004 | All client visits | 35.5% | Not reported | Not reported |
| Tomolillo 2007^53^ (Cross-sectional study) | - Needle and syringe program | Effect of needle and syringe program use on risk of SSTI | Yes | - SSTI | Self-report occurrences of abscesses (timeline not specified) | USA (Eureka, California) | 62 | Recruited “former intravenous drug users from local 12-step meetings” | 3 | January 1, 2002 to February 28, 2004 | “former intravenous drug users” | 41.9% | Not reported | Not reported |
| Trayner 2020^54^ | - Inject in public | Effect of public injecting on risk of SSTI | Yes | - SSTI | Past year SSTI, self-report | Scotland | 1469 | Recruited through clients at harm reduction programs (Needle Exchange Surveillance Initiative) | 4 | 2017 to 2018 | Injected drugs within past 6 months | 25% | Mean 39.6 years | Heroin |
| Weir 2019^55^ | - Prescription opioids - Drug policy change | Effect of removal of controlled-release oxycodone from market on risk of endocarditis. Also, effect of population rate of hydromorphone prescribing on risk of endocarditis. | Yes | - Endocarditis | Quarterly trend in proportion of hospital admissions with evidence of injection drug use that include endocarditis diagnosis | Canada (Ontario) | 60,529 hospital admissions | Hospital admissions with evidence of injection drug use, identified via administrative data | 3 | 2006 to 2015 | Age 18 to 55 years; hospital admissions with diagnostic codes indicating opioid or stimulant use, or hepatitis C | Endocarditis admissions: 53%  Other admissions: 44% | Endocarditis admissions:  Mean 36.3 (SD 9.5) years  Other admissions: Mean 40.2 (SD 10.8) years | Not reported |
| Wilson 2002^56^ | - Gender/sex - Age - Education - Housing - Alcohol | Not specified. Aimed to identify factors associated with endocarditis. | No estimand | - Endocarditis | Infective endocarditis, self-report and/or medical chart review | USA (Baltimore, Maryland) | 470 in nested case-control study (79 cases with endocarditis, 391 controls matched on date and duration follow-up) | Not reported (AIDS Link to Intravenous Experiences; ALIVE)  Similar sample as Islam 2019 and Safaeian 2000 | 4 | February 1988 to December 1998 | For nested case-control study: person with HIV, age 18 years or older, injected drugs at least once after 1977, no AIDS diagnosis at study entry, returned for at least one follow-up study visit | 26% | Age 39 or older  Endocarditis cases: 48.1%  Controls: 45.8% | Not reported |
| Wright 2020^57^ | - Gender/sex - Age - Housing | Not specified. Aimed to identify factors associated with SSTI. | No estimand | - SSTI | Lifetime SSTI, self-report  Question supported by pictures of abscesses and/or cellulitis at different stages | England (London) | 455 | Recruitment through street outreach and drug treatment services (Care & Prevent study) | 4 | October 2017 to March 2019 | Age 18 years or older; any prior injection drug use | 25% | Median 46 (IQR 39 to 52) years | Not reported |
| Wurcel 2016^58^ | - Gender/sex - Age - Race/ethnicity | Effect of age, race/ethnicity, and gender on trends in incidence of endocarditis | Yes | - Endocarditis | Percentage of hospital admissions for endocarditis that also have diagnostic codes suggestive of injection drug use | USA (national) | Not reported | Nationwide hospital administrative database (Nationwide Inpatient Sample) | 4 | 2000 to 2013 | Age 15 to 64 years; Hospitalization with ICD-9 codes consistent with endocarditis and also substance use or hepatitis C | 40.9% | Not reported | Not reported |
| Wurcel 2018^59^ | - Gender/sex - Age - Race/ethnicity - Education - Sex work - Heroin - Cocaine - Housing - Needle and syringe program | Effect of sex work on risk of SSTI, depending on gender | Partly (1 of 9 exposures) | - SSTI | Lifetime abscess, “Has a medical professional ever told you that you had an  abscess?”, self-reported  Past 30 days abscess, self-reported | USA (Boston and Worcester, Massachusetts) | 298 | Recruited via street outreach and clients at needle and syringe programs and local health services  (Responding to the Epidemic of Addiction and Hepatitis C Virus Together; REACTS; and HCV and HIV-HCV Hotspots Study) | 4 | 2015 to 2016 | Age 18-45 years; injected drugs in past 30 days | 30% | Median 33 (IQR 30 to 39) years | Heroin |
| Yeung 2017^60^ | - Other stimulant - Drug policy change | Effect of drug policy change (temporary class drug order on ethylphenidate) on STTI | Partly (1 of 2 exposures) | - Not specified | Weekly rate of *S. pyogenes* or *S. aureus* infections | Scotland (Lothian) | Estimated 3000 people who inject drugs in Lothian (211 cases) | Microbiology samples that grew *S. pyogenes* or *S. aureus* were investigated for injection drug use and use of ethylphenidate | 5 | February 2014 to December 2015 | Injection drug use | Among cases: 27.5% | Among cases:  20 and under: 0.5%  21-25: 4.3%  26-30: 13.7%  31-35: 28.0%  36-40: 23.2%  41-45: 17.1%  46-50: 10.9%  51+: 2.4% | Novel psychoactive substances (not otherwise specified) |

### Appendix 5. Characteristics of included studies with outcome during infection treatment in quantitative systematic review of social and structural determinants of injection drug use-associated bacterial and fungal infections.

| **Study** | **Included exposures in this review** | **Main exposure / estimand in study** | **Do exposure and outcome pairs included in this review reflect study estimand** | **Infections** | **Outcomes** | **Country (City)** | **Sample size** | **Sampling method (parent study name)** | **MMAT quality rating (out of 5)** | **Data collection period** | **Inclusion criteria** | **% women/female** | **Age** | **Drugs used by ≥50%** |
| --- | --- | --- | --- | --- | --- | --- | --- | --- | --- | --- | --- | --- | --- | --- |
| Cooksey 2020^61^ | - Housing - Hospital policy | Effect of hospital policy on all-cause readmission, after hospital discharge | Partly (1 of 2) | - Endocarditis | - Hospital discharge against medical advice - In-hospital mortality | USA (Knoxville, Tennessee) | 168 | Retrospectively identified admitted to tertiary care hospital with endocarditis and injection drug use, via electronic medical records | 5 | January 2013 to January 2019 | Age 19 years and older, diagnosis of definite infective endocarditis by modified Duke criteria, and active injection drug use (defined as self-reported in past 30 days, “positive urine drug screen for illicit substances”, or reported in infectious diseases consultant’s note)  Excluded patients who underwent invasive cardiac procedure in prior 30 days, had history of congenital or rheumatic heart disease, or “were missing finalized diagnostic test and/or culture results from an outside facility were excluded” | Pre-Intervention group: 54%  Post-Intervention group: 69% | Pre-Intervention group: Median 32 (IQR 26-41) years  Post-Intervention group: Median 36 (IQR 28-43) years | Opioids |
| Eaton 2020^62^ | - Gender/sex - Opioid agonist treatment | Not specified. Aimed to identify factors associated with in-hospital illicit drug use and with premature hospital discharge. | No estimand | - Multiple - SSTI - Endocarditis - Osteomyelitis - Septic arthritis - Sepsis/bacteremia | - Hospital discharge against medical advice | USA (Birmingham, Alabama) | 83 | Retrospectively identified patients admitted to tertiary care hospital and referred to a specialized “Intravenous Antibiotic and Addiction Team” | 2 | October 2016 to December 2017 | First hospitalization during the study period | 43% | Median 36.3 years | Opioids |
| Fink 2013^16^ | - Gender/sex - Age - Race/ethnicity - Housing - Needle and syringe program - Access to healthcare | Not specified. Aimed to identify factors associated with abscess self-treatment. | No estimand | - SSTI | - Self-treatment. “Thinking about the last abscess you had, how did you deal with it?” | USA (Los Angeles, Oakland, and Berkeley, California) | 858 | Recruited through street outreach and from clients at four large, government-sanctioned needle and syringe programs. | **4** | 2003 to 2005 | Age 18 and older, self-reported injection drug use in past 30 days | 29% | Less than 30 years: 5%  30-39 years: 16%  40-49 years: 37%  50+ years: 40% | Heroin |
| Hope 2008^20^ | - Gender/sex - Age - Housing - Incarceration - Overdose history - Cocaine - Amphetamines - Opioid agonist treatment | Not specified. Aimed to identify factors associated with seeking health care for abscess. | No estimand | - SSTI | - Healthcare seeking, self-reported | England (nationwide) | 1,058 | Recruited through street outreach and from clients of health and social services (Unlinked Anonymous Monitoring Survey) | 4 | Fall 2003 to Summer 2005 | Injected drugs in past 28 days | 23% | Median 30 (range 16 to 72) years | Opiates |
| Hope 2015^18^ | - Gender/sex - Age - Income/employment - Sex work - Housing - Incarceration - Overdose history - Migration status - Heroin - Cocaine - Amphetamines | Not specified. Aimed to identify factors associated with seeking health care for abscess and with hospital admission | No estimand | - SSTI | - Healthcare seeking, “sought medical advice (i.e. from a doctor or nurse) about that symptom.” - Hospital admission, “if they had then been admitted to hospital as a result of that symptom” | England (Bristol, Leeds, and Birmingham) | 855 | Respondent-driven sampling | 5 | 2006 (Bristol), 2008 (Leeds), 2009 (Birmingham) | Age 16 years or older, injected drugs in preceding 4 weeks, and live within the survey area | 25% | Median 31 years, mean 32 years | Heroin; crack cocaine |
| Jo 2021^63^ | - Gender/sex - Age - Race/ethnicity - Access to healthcare - Opioid agonist treatment - Stimulants | Effect of opioid agonist treatment on multiple outcomes | Partly (1 of 6 exposures) | - Multiple - Endocarditis - Osteomyelitis | - Hospital discharge against medical advice | USA (143 hospitals across 21 states) | 1407 | Patients admitted to one large nationwide hospital system with concurrent diagnostic codes for opioid use disorder and endocarditis or osteomyelitis | 5 | 1 January 2014 to 31 December 2018 | Age 18 to 65 | 44% | Mean 42.7 years | Opioids |
| Kimmel 2020^64^ | - Gender/sex - Age - Income/employment - Access to healthcare - Unhealthy alcohol use - Other substance use - Surgery in-hospital - Hospital characteristics | Not specified. Aimed to identify factors associated with hospital discharge against medical advice | No estimand | - Endocarditis | - Hospital discharge against medical advice | USA (nationwide) | 7,259 | Patients admitted to hospital with diagnostic codes for native valve endocarditis and opioids, stimulants, and/or hepatitis C virus (Nationwide Inpatient Sample) | 5 | January 2010 to September 2015 | Age 18-64 years | 43.3% | 18-24 years: 10.8% 25-34 years: 31.8% 35-44 years: 21.1% 45-55 years: 22.6% 56-65 years: 13.8% | Not reported |
| Kimmel 2020^65^ | - Hospital policy | Effect of initiating public outcomes reporting for aortic valve surgery on multiple outcomes | Yes | - Endocarditis | - In-hospital mortality | USA (nationwide) | 7,322 | Patients admitted to hospital with diagnostic codes for native valve endocarditis and opioids, stimulants, and/or hepatitis C virus (Nationwide Inpatient Sample) | 5 | 1 January 2010 to 31 August 2015 | Age 18-65 years | Pre-intervention: 39.2%  Post-intervention: 45.5% | Pre-intervention: 41.2 years  Post-intervention: 38.5 years | Not reported |
| Lloyd-Smith 2012^25^ | - Supervised consumption site | Not specified. Aimed to identify factors associated with emergency department visit for SSTI | No estimand | - SSTI | - Healthcare seeking, defined by emergency department visit identified via administrative data | Canada (Vancouver, British Columbia) | 1,083 | Randomly recruited clients at supervised injecting facility  (Scientific Evaluation of Supervised Injection; SEOSI)  Similar sample to Lloyd-Smith 2008 and Milloy 2010, and same sample as Lloyd-Smith 2009 and Lloyd-Smith 2010 | 4 | 1 January 2004 to 31 January 2008 | Not reported | 29% | Median (interquartile range) was 35.1 (28.7 to 41.5) years among females and 39.7 (33.7 to 45.3) years among males | Not reported |
| Lloyd-Smith 2010^28^ | - Supervised consumption site | Not specified. Aimed to identify factors associated with hospital admission for SSTI | No estimand | - Multiple - SSTI - Osteomyelitis - Endocarditis - Septic arthritis | - Healthcare seeking, defined as hospital admission identified via administrative data | Canada (Vancouver, British Columbia) | 1,083 | Randomly recruited clients at supervised injecting facility  (Scientific Evaluation of Supervised Injection; SEOSI)  Similar sample to Lloyd-Smith 2008 and Milloy 2010, same sample as Lloyd-Smith 2009 and Lloyd-Smith 2012 | 4 | 1 January 2004 to 31 January 2008 | Not reported | 29% | Median (interquartile range), 38.4 (32.7 - 44.3) | Not reported |
| Marks 2020a^66^ | - Addiction treatment | Effect of hospital inpatient addiction medicine consultation on multiple outcomes | Yes | - Multiple - Endocarditis - Fungemia - Bacteremia - SSTI - Septic arthritis - Epidural abscess - Osteomyelitis | - Hospital discharge against medical advice | USA (St. Louis, Missouri) | 125 | All hospital admissions with diagnoses of injecting-related infection and opioid use disorder at at one tertiary care hospital who received infectious diseases consultation , identified via electronic medical records | 4 | January 2016 to January 2018 | Infection attributable to injection drug use by the infectious diseases consultant; greater than 2 weeks of intravenous antibiotics treatment was recommended; patient was not eligible to receive outpatient treatment | Consultation group: 55%  No consultation group: 52% | Consultation group: Median 36 (range 19-63) years  No consultation group: Median 35 (range 19-67) years | Heroin |
| Marks 2020b^67^ | - Opioid agonist treatment | Effect of opioid agonist treatment on multiple outcomes | Yes | - Endocarditis | - Hospital discharge against medical advice | USA (St. Louis, Missouri) | 123 | Consecutive patients referred for infectious diseases consultation with opioid injection-associated infections, identified prospectively | 4 | 1 July 2017 to 1 May 2020 | Confirmed as injection opioid use-associated endocarditis by study physician on retrospective review of medical records | 47% | Median 34 (IQR 25-48) years | Opioids |
| Martín-Dávila 2005^68^ | - Gender/sex | Not specified. Aimed to identify factors associated with in-hospital mortality | No estimand | - Endocarditis | - In-hospital mortality | Spain (Madrid) | 220 | All patients with diagnosis of endocarditis, identified retrospectively via electronic health records | 5 | 1 January 1985 to 31 December 1999 | “Injection drug users”. Operational definition not reported. | 14% | Median 27.8 (range 18-44) years | Not reported |
| Meel 2018^69^ | - Age | Not specified. Aimed to identify factors associated with in-hospital mortality | No estimand | - Endocarditis | - In-hospital mortality | South Africa (Johannesburg) | 68 | All patients seen at cardiology clinic with endocarditis “secondary to IV nyaope use”, identified retrospectively. | 4 | December 2014 to February 2017 | Age 18 years and older; definite or possible infective endocarditis by modified Duke criteria; “history of IV nyaope use” (not otherwise specified) | 2.9% | Mean 25.8 (SD 4.5) years | Nyaope |
| Mertz 2008^70^ | - Gender/sex - Age - Unhealthy alcohol use | Not specified. Aimed to identify factors associated with multiple outcomes. | No estimand | - Multiple - SSTI - Endocarditis - Osteomyelitis - Septic arthritis - Sepsis/bacteraemia - Pneumonia | - Hospital discharge against medical advice - In-hospital mortality | Switzerland (Basel) | 216 | Among all patients admitted to tertiary care hospital, identified “intravenous drug users” (not defined) and those referred for infectious diseases consultation | 4 | January 2001 to December 2006 | Not reported | 33% | Median 38 (range 18-58) years | Opioids |
| Monteiro 2020^71^ | - Gender/sex - Age - Race/ethnicity - Access to healthcare - Heroin - Cocaine | Not specified. Aimed to identify factors associated with self-treatment of SSTI | No estimand | - SSTI | - Self-treatment, self-reported | USA (Boston, Massachusetts) | 162 | Recruited from among hospital inpatients with “an indication” of current or past injection drug use or a current skin abscess or cellulitis in electronic medical record  Similar sample as Stein 2020 | 4 | January 2014 to June 2018 | Age 18 years or older; self-reported injection drug use at least three times during week prior to hospitalization | 40.7% | Mean 38 (SD 10.5) years | Not reported |
| Nolan 2020^72^ | - Gender/sex - Housing - Heroin/fentanyl - Polysubstance use - Access to healthcare - Opioid agonist treatment | Effect of OAT on hospital discharges against medical advice.  Also aimed to identify factors associated with hospital discharges against medical advice. | Partly (1 of 6 exposures) | - Multiple - Endocarditis - Osteomyelitis Septic arthritis - Epidural abscess - Sepsis/bacteraemia | - Hospital discharge against medical advice | USA (St. Louis, Missouri) | 262 | All hospital admissions with diagnoses of injecting-related infection and opioid use disorder at at one tertiary care hospital who received infectious diseases consultation , identified via electronic medical records  Similar sample as Marks 2020a (10.1093/cid/ciy924.) | 4 | January 2016 to July 2019 | Not reported | Inpatient OAT group: 60.1%  No OAT group: 41.1% | Inpatient OAT group: mean 38 (SD 9) years  No OAT group: mean 41 (SD 12) years | Opioids (fentanyl or heroin) |
| Rudasill 2019^73^ | - Surgery in-hospital | Effect of valve surgery for endocarditis on multiple outcomes | Yes | - Endocarditis | - Hospital discharge against medical advice - In-hospital mortality | USA (nationwide) | 27,432 | All hospitalized patients with endocarditis in nationwide hospital admissions database  (National Readmissions Database) | 4 | January 2010 to September 2015 | Age 16 to 64; diagnostic codes for illicit drug use; no congenital or rheumatic heart disease; no cardiac procedures | 45.3% | Mean 38.3 (SD 0.1) years | Not reported |
| Sandrock 2001^74^ | - Gender - Age | Not specified. Aimed to identify factors associated with respiratory failure. | No estimand | - Botulism | - Respiratory failure | USA (Sacramento, California) | 20 | Consecutive patients with a discharge diagnosis of botulism | 4 | 1990 to 1999 | Injection drug use “within the months preceding hospitalization”, or “a positive result on toxicology screen” | 25% | Median 47 years | Heroin |
| Saydain 2010^75^ | - Gender/sex - Age - Race/ethnicity | Not specified. Aimed to identify factors associated with in-hospital mortality. | No estimand | - Endocarditis | - In-hospital mortality | USA (Detroit, Michigan) | 33 | Patients admitted to teaching hospital intensive care unit with diagnosis of endocarditis by modified Duke criteria and “were injection drug users” | 4 | January 2001 to December 2006 | Not reported | 45% | Mean 47.2 (SD 10.5) years | Heroin |
| Serota 2021^76^ | - Gender/sex - Age - Race/ethnicity - Overdose history - Opioids - Stimulants - Polysubstance - Access to healthcare | Effect of stimulant use and stimulant/opioid co-use vs. opioid use-only on multiple outcomes | Partly (3 of 8 exposures) | - Multiple - SSTI - Sepsis/bacteraemia - Endocarditis - Osteomyelitis | - Hospital discharge against medical advice - In-hospital mortality | USA (Florida, statewide) | 31,964 | Census of all patients admitted to all hospitals in Florida   (Agency for Health Care Administration Hospital Inpatient Limited Data Set) | 5 | 1 January 2016 to 31 December 2017 | Hospital admissions with ICD-10 code for injecting-related infections and opioid- or stimulant-related diagnostic codes | 46% | Median 44 (IQR 33-56) years | Opioids |
| Suzuki 2020^77^ | - Opioid agonist treatment | Effect of opioid agonist treatment on hospital discharge against medical advice | Yes | - Endocarditis | - Hospital discharge against medical advice | USA (Boston, Massachusetts) | 84 | All patients admitted to tertiary care hospital with discharge diagnosis including endocarditis and opioid/heroin or injection drug use, identified retrospectively via electronic medical records | 5 | 1 January 2016 to 31 December 2018 | Diagnosis of opioid use disorder and hospital admission with endocarditis attributed to injection drug use; recent injection drug use (not otherwise specified) | 46.4% | Mean 36.2 (SD 10.3) years | Opioids; cocaine; tobacco |
| Takahashi 2007^78^ | - Age - Gender/sex - Race/ethnicity - Education - Income/employment - Housing - Heroin - Needle and syringe program - Unhealthy alcohol use - Access to healthcare | Not specified. Aimed to identify factors associated with hospital admission | No estimand | - SSTI | - Hospital admission | USA (Seattle, Washington) | 136 | Prospectively recruited emergency department patients who inject drugs with SSTI | 4 | May 2001 to March 2002 | English-speaking; provided informed consent | 38% | Mean 43 (SD 8) years | Not reported |
| Tan 2020^79^ | - Gender/sex - Age - Housing - Opiates - Stimulants - Polysubstance - Other prescription medications - Addiction treatment - PICC line | Not specified. Aimed to identify factors associated with new bloodstream infections | No estimand | - Endocarditis | - New bloodstream infection during treatment | Canada (London, Ontario) | 309 | Patients admitted to three urban hospitals with discharge diagnosis codes for endocarditis, identified via administrative data | 4 | 1 April 2007 to 31 March 2018 | Diagnosis of definite endocarditis by modified Duke criteria; injected drugs in prior 3 months | 49.3% | Mean 35.7 (SD 9.7) years | Opiates; stimulants |
| Uppuluri 2021^80^ | - Gender/sex - Age - Race/ethnicity - Cocaine - Amphetamines - Unhealthy alcohol use - Other substance use - PICC line | Not specified. Aimed to identify factors associated with endogenous endophthalmitis | No estimand | - Multiple - SSTI - Sepsis/bacteraemia - Endocarditis - Osteomyelitis - Candidemia - Endophthalmitis | - Development of endogenous endophthalmitis | USA (nationwide) | 605,859 | Hospital admissions (at a hospital contributing to nationwide database) with diagnosis codes for opioid use disorder or overdose, and injecting-related infections  (National Inpatient Sample) | 5 | 2002 to 2014 | Age 21-65 years; | 42.7% | “Average” 42.7 years | Not reported |
| Wang 2020^81^ | - Opioid agonist treatment - Hospital policy | Effect of hospital policy and of opioid agonist treatment on premature hospital discharge | Yes | - Multiple - SSTI - Sepsis/bacteraemia - Endocarditis - Osteomyelitis - Septic arthritis | - Hospital discharge against medical advice | USA (Concord, New Hampshire) | 147 | Patients admitted to a suburban hospital with diagnoses of “intravenous drug use” or “opioid use disorder” and injecting-related infections, identified retrospectively via electronic medical records search | 4 | 1 January 2018 to 1 October 2019 | Infection related to “intravenous opioid use” (excluding people only injecting stimulants); not currently in law enforcement custody; not critically ill | 48.3% | Average 35.9 years | Not reported |

### **Appendix 6. Characteristics of included studies with outcome after initial treatment in quantitative systematic review of social and structural determinants of injection drug use-associated bacterial and fungal infections**.

| **Study** | **Included exposures in this review** | **Main exposure / estimand in study** | **Do exposures included in this review reflect study estimand** | **Infections** | **Outcomes** | **Country (City)** | **Sample size** | **Sampling method (parent study name)** | **MMAT quality rating (out of 5)** | **Data collection period** | **Inclusion criteria** | **Women/female** | **Age** | **Drugs used by** ≥**50%** |
| --- | --- | --- | --- | --- | --- | --- | --- | --- | --- | --- | --- | --- | --- | --- |
| Barocas 2020^82^ | - Age - Gender/sex - Opioid agonist treatment - Other substance use | Effect of opioid agonist treatment on multiple outcomes | Partly (1 of 4) | - SSTI | - Infection-related rehospitalization - All-cause rehospitalization - Overdose-related rehospitalization | USA (Nationwide) | 6,538 | Private/commercial health insurance claims database, with hospital admissions for injecting-related infections identified via discharge diagnosis codes. | 4 | 2010 to 2017 | Age 18-64 years, hospital admission for SSTI, minimum 30-day follow-up after hospital discharge, diagnostic codes for opioid use disorder within 6 months before or after the index SSTI hospitalization.  Excluded people who had a pharmacy claim for opioid agonist treatment in three months preceding hospitalization. | 48% | Mean 40 (SD 14.5) years | Opioids |
| Barocas 2021^83^ | - Age - Gender/sex - Opioid agonist treatment - Other substance use | Effect of opioid agonist treatment on multiple outcomes | Partly (1 of 4) | - Endocarditis | - All-cause rehospitalization | USA (Nationwide) | 768 | Private/commercial health insurance claims database, , with hospital admissions for injecting-related infections identified via discharge diagnosis codes. | 4 | 1 July 2020 to 30 June 2016 | Age 18 years and older, hospital admission for endocarditis, minimum of 30-day follow-up after hospital discharge, | 48.7% | Mean 39 (SD 15.5) years | Opioids |
| Buehrle 2017^84^ | - Age - Discharge location | Not specified. Aimed to identify factors associated with OPAT failure. | No estimand | - Multiple infections - Endocarditis - Epidural abscess - Sepsis/bacteremia - SSTI - Osteomyelitis | - OPAT complications | USA (Pittsburgh, Pennsylvania) | 118 | Retrospective chart review of hospital records. Sampling approach not specified. | 4 | December 2013 to January 2015 | Self-reported injection drug use in 4 weeks preceding hospitalization, or “a positive urine drug screen plus suspicion of” injection drug use | Not reported | Median 34.5 years | Not reported |
| Clarelin 2021^85^ | - Age - Gender/sex | Not specified. Aimed to identify factors associated with all-cause mortality | No estimand | - Endocarditis | - All-cause mortality | Sweden (nationwide) | 586 | Registry of patients admitted to hospital with endocarditis, with voluntary reporting by physicians (Swedish Registry on Infective Endocarditis) | 5 | 2008 to 2019 | Assessed by physician to be person who injects drugs | Left-sided endocarditis: 23%  Right-sided endocarditis: 40% | Left-sided endocarditis:  Mean 46 (SD 12) years  Right-sided endocarditis: Mean 35 (SD 9) years | Not reported |
| Connell 2010^86^ | - Age - Gender/sex | Effect of age and gender on change in visual acuity after treatment of endogenous endophthalmitis | Yes | - Endophthalmitis | - Visual acuity after treatment | Australia (Melbourne, Victoria) | 19 | Consecutive patients with endogenous fungal endophthalmitis admitted to a specialized, quaternary care hospital for eye and ear disorder | 4 | 2001 to 2007 | A history of injection drug use (not otherwise specified) | 58% | Mean 32.7 (SD 8) years | Not reported |
| Cooksey 2020^61^ | - Health care access - Hospital policy | Effect of hospital policy on all-cause readmission, after hospital discharge | Partly (1 of 2) | - Endocarditis | - All-cause rehospitalization - All-cause mortality | USA (Knoxville, Tennessee) | 168 | Retrospectively identified admitted to tertiary care hospital with endocarditis and injection drug use, via electronic medical records | 5 | January 2013 to January 2019 | Age 19 years and older, diagnosis of definite infective endocarditis by modified Duke criteria, and active injection drug use (defined as self-reported in past 30 days, “positive urine drug screen for illicit substances”, or reported in infectious diseases consultant’s note)  Excluded patients who underwent invasive cardiac procedure in prior 30 days, had history of congenital or rheumatic heart disease, or “were missing finalized diagnostic test and/or culture results from an outside facility were excluded” | Pre-Intervention group: 54%  Post-Intervention group: 69% | Pre-Intervention group: Median 32 (IQR 26-41) years  Post-Intervention group: Median 36 (IQR 28-43) years | Opioids |
| D’Couto 2018^87^ | - Discharge location | Effect of discharge location (home vs. skilled nursing facility) on OPAT failure | Yes | - Multiple infections - Endocarditis - Osteomyelitis - Septic arthritis | - OPAT complications | USA (Boston, Massachusetts) | 52 | All patients enrolled in OPAT program at tertiary care hospital | 3 | 1 January 2010 to 31 December 2015 | Recent or active injection drug use (as documented in the medical record) | Discharged home: 29%  Discharged to skilled nursing facility: 32% | Not reported | Not reported |
| Fanucchi 2020^88^ | - Discharge location | Effect of discharge location (outpatient with integrated care vs. remaining in hospital). | Yes | - Multiple - Endocarditis - Osteomyelitis | - OPAT complications | USA (Lexington, Kentucky) | 20 | Recruited patients hospitalized at a tertiary care hospital with injecting-related infections | 3 | 1 March 2017 to 2 October 2018 | Age 18 to 65 years; moderate to severe opioid use disorder; injecting-related infection requiring 2 or more weeks of intravenous antibiotics; accepting buprenorphine opioid agonist treatment; living within 45 minutes of the hospital; home discharge expected  Excluded people with central nervous system complications of infection (e.g. embolic stroke), end-stage renal disease, Class III or IV congestive heart failure, decompensated cirrhosis, prosthetic valve or fungal endocarditis, concurrent dependence on sedative-hypnotics, homelessness, current pregnancy, or incarceration | Outpatient: 70%  Inpatient: 70% | Outpatient:  Mean 32.9 (range 26-38) years  Inpatient:  Mean 31.3 (range 21-48) years | Opioids, stimulants |
| Hilbig 2020^89^ | - Opioid agonist treatment | Not specified. Aimed to identify factors associated with multiple outcomes. | No estimand | - Endocarditis | - Infection-related rehospitalization | Australia (Melbourne, Victoria) | 46 | Patients hospitalized with endocarditis at a tertiary care hospital, identified retrospectively using discharge diagnosis codes | 3 | 2008 to 2015 | Diagnosis of definite or possible infective endocarditis, by Duke criteria; documentation of reporting injection drug use within prior 3 months; age younger than 70 years | 41% | Median 39 (IQR 34-47.5) years | None |
| Huang 2018^90^ | - Age - Gender/sex - Race/ethnicity - Rural/urban - Prescription opioids | Not specified. Aimed to identify factors associated with recurrence of endocarditis | No estimand | - Endocarditis | - Infection-related rehospitalization | USA (Winston-Salem, North Carolina) | 87 | Patients hospitalized with endocarditis at a tertiary care hospital, identified retrospectively using discharge diagnosis codes | 5 | January 2004 to January 2017 | Age 18 years or older; no intracardiac device; diagnosis of definite or possible infective endocarditis by modified Duke criteria; documentation of reporting injection drug use within prior 3 months | 50% | Median 28.5 years | Prescription opioids |
| Jo 2021^63^ | - Age - Race/ethnicity - Health care access - Cocaine/amphetamines - Opioid agonist treatment | Effect of initiating opioid agonist treatment in-hospital on multiple outcomes | Partly (1 of 5) | - Multiple infections - Endocarditis - Osteomyelitis | - All-cause rehospitalization | USA (nationwide) | 1407 | Patients hospitalized with (a) endocarditis or osteomyelitis and (b) opioid use disorder at a large health system, identified retrospectively using discharge diagnosis codes | 4 | 1 January 2014 to 31 December 2018 | Age 18 to 65 years | 44% | Mean 42.7 | Opioids |
| Kimmel 2020^91^ | - Age - Gender/sex - Unstable housing - Opioid agonist treatment | Effect of opioid agonist treatment after hospital discharge on all-cause mortality | Partly (1 of 4) | - Endocarditis | - All-cause mortality | USA (Massachusetts) | 679 | People in a state-wide hospital medical claims database with (a) endocarditis-related hospital admission and (b) opioid, cocaine, or amphetamine use, or hepatitis C virus infection, identified retrospectively using discharge diagnosis codes | 5 | 1 January 2011 to 31 December 2015 | Age 18 to 65 years | 39.2% | Mean 39.2 (SD 12.1) years | Opioids |
| Marks 2020a^92^ | - Age - Gender/sex - Race/ethnicity - Unstable housing - Heroin/fentanyl - Cocaine - Amphetamines - Other substance use - Inpatient addiction medicine consultation service - Antibiotic treatment mode - Surgery during hospitalization | Effect of antibiotic treatment mode on rehospitalization | Partly (1 of 11 exposures) | - Multiple infections - Endocarditis - Septic arthritis - Sepsis/bacteremia - Osteomyelitis - Epidural abscess | - All-cause rehospitalization | USA (St. Louis, Missouri) | 293 | Patients hospitalized with bacterial infections potentially consistent with injection drug use, identified retrospectively using discharge diagnosis codes | 5 | 1 January 2016 to 30 July 2019 | Infectious diseases consultation occurred; infection related to injection drug use, as determined by Infectious Diseases consultant physician  May be overlapping sample with Marks 2020b^66^ and Marks 2020c^67^ | Complete IV: 45.5%  Partial IV, no oral: 59.7%  Partial IV, partial oral: 48.2% | Complete IV: Mean 40 (range 20-71) years  Partial IV, no oral: Mean 38 (range 20-71) years  Partial IV, partial oral:  Mean 39 (range 26-61) years | Heroin or fentanyl |
| Marks 2020b^66^ | - Inpatient addiction medicine consultation | Effect of inpatient addiction medicine consultation on multiple outcomes | Yes | - Multiple infections - Osteomyelitis - Epidural abscess - Septic arthritis SSTI | - All-cause rehospitalization | USA (St. Louis, Missouri) | 125 | Patients hospitalized with bacterial infections potentially consistent with injection drug use, identified retrospectively using discharge diagnosis codes | 4 | January 2016 to January 2018 | Infectious diseases consultation occurred; infection related to injection drug use, as determined by Infectious Diseases consultant physician; recommended >2 weeks antibiotic course; patient ineligible for OPAT  May be overlapping sample with Marks 2020a^92^ and Marks 2020c^67^ | Addiction medicine consultation: 55%  No addiction medicine consultation: 52% | Addiction medicine consultation: Median 36 (range 19-63 years)  No addiction medicine consultation:  Median 35 (range 19-67 years) | Heroin |
| Marks 2020c^67^ | - Opioid agonist treatment | Effect of opioid agonist treatment on multiple outcomes | Yes | - Endocarditis | - All-cause mortality | USA (St. Louis, Missouri) | 123 | Consecutive patients hospitalized with injecting related infections and opioid use disorder, who were referred for infectious diseases consultation. Enrolled in a prospective registry. | 4 | 1 July 2017 to 1 May 2020 | Infective endocarditis caused by injection drug use, as determined by infectious diseases consultant  May be overlapping sample with Marks 2020a^92^ and Marks 2020b^66^ | 47% | Median 37 (IQR 25-48) years | Opioids |
| Nguemeni Tiako 2020^93^ | - Inpatient addiction medicine consultation | Effect of hospital inpatient addiction on all-cause mortality | Yes | - Endocarditis | - Drug rehabilitation program attendance | USA (New Haven, Connecticut) | 56 (subgroup of 42 with “active drug use” used for mortality analysis) | Consecutive patients undergoing cardiac surgery for injecting-related endocarditis. Sampling approach not described. | 3 | 2011 to 2016 | Infective endocarditis diagnosis by USA Center for Disease Control and Prevention (CDC) criteria; history of injection drug use | 12.5% | Mean 44 (SD 13) years | Heroin |
| Pericàs 2021^94^ | - Gender/sex - Surgery during hospitalization | Not specified. Aimed to identify factors associated with multiple outcomes | No estimand | - Endocarditis | - Infection-related rehospitalization - All-cause mortality | International (30 countries) | 591 | Patients with endocarditis enrolled in one of two prospective cohort studies. Sampling approach not described.  (International Collaboration on Endocarditis [ICE] Prospective Cohort Study and ICE-Plus study) | 3 | 1 January 2000 to 31 December 2006 and 1 September 2008 to 31 December 2012 | “People who inject drugs”, not otherwise defined | 27.5% | Median 37.0 (IQR 29.5-44.2) years | Not reported |
| Ray 2020^95^ | - Hospital policy | Effect of change in hospital policy on multiple outcomes | Yes | - Endocarditis | - Infection-related rehospitalization - All-cause rehospitalization | USA (Milwaukee, Wisconsin) | 70 | Patients admitted to tertiary care hospital with endocarditis and opioid use, identified retrospectively via discharge diagnosis codes | 4 | 1 January 2015 to 31 December 2016 and 1 April 2017 to 31 March 2018 | Self-reported “intravenous drug use”, as documented in the medical record | Pre-intervention: 56.8%  Post-intervention: 57.6% | Pre-intervention: Median 31 (range 18-54) years  Post-intervention:  Median 31 (range 25 to 52) years | Tobacco; otherwise, not reported |
| Rodger 2018^96^ | - Age - Gender/sex - Opioids - Stimulants - Polysubstance use - Opioid agonist treatment - Hospital discharge against medical advice - Other substance use/addiction treatment - Surgery during hospitalization | Not specified. Aimed to identify factors associated with all-cause mortality. | No estimand | - Endocarditis | - All-cause mortality | Canada (London, Ontario) | 202 | All patients admitted to three hospitals with discharge diagnosis of infective endocarditis, identified retrospectively  Sample largely overlaps with Rodger 2019 and Tan 2020 | 3 | 1 April 2007 to November 2017 | Definite infective endocarditis by modified Duke criteria; self-reported injection drug use, as per medical record | 48% | Median 34 (IQR 28-42) years | “Polysubstance” |
| Rodger 2019^97^ | - Other substance use/addiction treatment - Surgery during hospitalization - Hospital discharge against medical advice | Not specified. Aimed to identify factors associated with recurrent endocarditis | No estimand | - Endocarditis | - Infection-related rehospitalization | Canada (London, Ontario) | 212 | All patients admitted to three hospitals with discharge diagnosis of infective endocarditis, identified retrospectively  Sample largely overlaps with Rodger 2018 and Tan 2020 | 5 | February 2007 to March 2016 | Age 18 years and older; Definite infective endocarditis by modified Duke criteria; self-reported injection drug use in preceding three months, as per medical record | 48.6% | Median 34 (IQR 28-42 years) | “Polysubstance” |
| Rohn 2020^98^ | - Age - Gender/sex | Not specified. Aimed to identify factors associated with all-cause mortality | No estimand | - Endocarditis | - All-cause mortality | Czech Republic (Prague, Motol) | 72 | All patients undergoing cardiac surgery for endocarditis. Sampling approach not described. | 3 | March 2006 to December 2015 | “Active intravenous drug use was confirmed by both the patient and the attending physician” | 38.9% | Mean 29.4 (SD 5.8) years | Not reported |
| Rudasill 2019^73^ | - Age - Surgery during hospitalization | Not specified. Aimed to identify factors associated with all-cause rehospitalization | No estimand | - Endocarditis | - All-cause rehospitalization | USA (nationwide) | 27,432 (survey-weighted) | All patients admitted to hospitals reporting to nationwide administrative database, identified retrospectively via discharge diagnosis codes for (a) endocarditis and (b) use of cocaine, heroin, or methamphetamine  (National Readmissions Database) | 5 | January 2010 to September 2015 | Age 16 to 64 years and older;  Excluded patients with diagnosis codes for congenital or rheumatic heart disease | 45.3% | Mean 38.3 (SD 0.1) years | Not reported |
| Slaughter 2019^99^ | - Age - Surgery during hospitalization | Effect of type of surgery on multiple surgery | Partly (1 of 2 exposures) | - Endocarditis | - All-cause rehospitalization - All-cause mortality | USA (nationwide) | 1,613 | Patients enrolled in a national cardiac surgery registry. Sampling approach not described.  (Society of Thoracic Surgeon Adult Cardiac Surgical Database) | 2 | July 2011 to December 2016 | Undergoing cardiac surgery for tricuspid valve endocarditis  Excluded patients with severe aortic or mitral valve insufficiency | 60% | Median 30 (IQR 26-36) years | Not reported |
| Straw 2020^100^ | - Age - Gender/sex - Surgery during hospitalization | Effect of cardiac surgery on all-cause mortality | Partly (1 of 3 exposures) | - Endocarditis | - All-cause mortality | England (Leeds, Yorkshire) | 92 | Prospectively collected data on consecutive patients admitted to hospital with endocarditis | 5 | 1 January 2006 to 31 December 2016 | Definite or possible endocarditis by modified Duke criteria; “IVDU within 90 days” (not otherwise defined) | 29% | Mean 36.7 (SD 8.4) years | Not reported |
| Suzuki 2020^101^ | - Opioid agonist treatment | Effect of opioid agonist treatment on multiple outcomes | Yes | - Endocarditis | - Infection-related rehospitalization - All-cause mortality | USA (Boston, Massachusetts) | 26 | Consecutive patients admitted to a tertiary care hospital with infective endocarditis, and referred for in-hospital addiction consultation | 2 | 2013 to 2015 | None specified | 50% | Mean 33.8 (SD 12.0) years | Heroin; cocaine |
| Tan 2020^79^ | - Other substance use/addiction treatment | Not specified. Aimed to identify factors associated with multiple outcomes. | No | - Endocarditis | - All-cause mortality | Canada (London, Ontario) | 309 | All patients admitted to three hospitals with discharge diagnosis of infective endocarditis, identified retrospectively  Sample largely overlaps with Rodger 2018 and Rodger 2019 | 4 | 1 April 2007 to 31 March 2018 | Definite infective endocarditis by modified Duke criteria; self-reported injection drug use in preceding three months, as per medical record | 49.3% | Mean 35.7 (SD 9.7) years | Opiate; stimulant; “polysubstance” |
| Thønnings 2020^52^ | - Age - Opioid agonist treatment - Other substance use/addiction treatment | Not specified. Aimed to identify factors associated with recurrent bacteremia. | No estimand | - Sepsis/bacteraemia | - Infection-related rehospitalization | Denmark (Hvidovre) | 58 | Patients admitted to one tertiary care hospital with electronic medical record entries consistent with injection drug use | 2 | 1 January 2000 to 31 December 2006 | Age 19 years or older; “drug abuse including injection of drugs”; bacteraemia during hospital admission | Not reported | Median 40 (SD 35-45) years | Not reported |
| Wang 2020^81^ | - Opioid agonist treatment - Hospital policy | Effect of opioid agonist treatment and of hospital policy change on multiple outcomes | Yes | - Multiple infections - Osteomyelitis - Endocarditis - SSTI - Septic arthritis | - All-cause rehospitalization - Overdose-related rehospitalization - All-cause mortality | USA (Concord, New Hampshire) | 147 (146 with bacterial infections, 1 with acute hepatitis C) | Patients admitted to one suburban community hospital with medical records describing (a) intravenous drug use or opioid use disorder and (b) injecting-related infections. Identified retrospectively via electronic medical record search. | 4 | 1 January 2018 to 1 October 2019 | Primary indication for hospital admission is injecting-related infection from injection opioid use.  Excluded patients injecting only stimulants; in law enforcement custody; critical illness | 48.3% | Average 35.9 (summary statistic not defined) | Opioids |
| Weymann 2014^102^ | - Age - Gender/sex | Not specified. Aimed to identify factors associated with all-cause mortality. | No estimand | - Endocarditis | - All-cause mortality | Germany (Heidelberg) | 20 | Consecutive patients undergoing cardiac surgery for endocarditis at one tertiary care hospital | 5 | January 1993 to July 2013 | Definite infective endocarditis by modified Duke criteria; medical record documentation of “acknowledgment by the patient of active intravenous drug abuse (heroin) until the day of admission” | 35% | Mean 35 (SD 7.7) years | Heroin |

### Appendix 7. Characteristics of included studies where outcome is colonization with pathogenic bacteria in quantitative systematic review of social and structural determinants of injection drug use-associated bacterial and fungal infections.

| **Study** | **Included exposures in this review** | **Main exposure / estimand in study** | **Do exposures included in this review reflect study estimand** | **Infections** | **Outcomes** | **Country (City)** | **Sample size** | **Sampling method (parent study name)** | **MMAT quality rating (out of 5)** | **Data collection period** | **Inclusion criteria** | **% women/female** | **Age** | **Drugs used by** ≥**50%** |
| --- | --- | --- | --- | --- | --- | --- | --- | --- | --- | --- | --- | --- | --- | --- |
| Colombo 2012^103^ | - Gender/sex - Age | Not specified. Aimed to identify factors associated with MRSA colonisation | No estimand | - Colonisation | - MRSA colonisation (nasal/throat or wound swab) | Switzerland (Zurich) | 497 | Clients of harm reduction programs and other health and social services. All clients during given day invited to participate. | 4 | November 2008 to September 2009 | History of intravenous drug use | 21.1% | Median 41 (range 18-60) years | Not reported |
| Leibler 2019^104^ | - Unstable housing | Not specified. Aimed to identify factors associated with MRSA colonisation | No estimand | - Colonisation | - MRSA colonisation (nasal swab) | USA (Boston, Massachusetts) | 78 | Patients recruited from hospital inpatient units at an urban, “safety net” tertiary care hospital | 4 | October 2016 to April 2018 | Self-reported injection drug use in at least 3 days out of the week prior to hospital admission; spoken English language proficiency; ability to return for follow-up; at least two additional contacts with valid phone numbers; no known upcoming prison sentences or planned move away | 36% | Mean 38.7 (SD 11) years | “nearly 90%” |
| Leung 2015^105^ | - Gender/sex - Age - Race/ethnicity - Education - Income/employment - Relationship status - Unstable housing - Incarceration - Heroin - Cocaine - Amphetamines - Speedball - Prescription opioids - Other substance use - Recent hospitalization - Other substance use/addiction treatment | Not specified. Aimed to identify factors associated with S. aureus colonisation | No estimand | - Colonisation | - MRSA and MSSA colonisation (nasal swab) | USA (Houston, Texas) | 440 | Respondent driven sampling | 5 | September 2012 to December 2012 | Having a valid recruitment coupon (for RDS); not already enrolled in the study; age 18 years or older; lived in the local area; injected drugs in the past 12 months; completed the interview in English/Spanish; visible evidence of recent injection (e.g., “track marks”); knowledge of drug preparation, injection, and needles/syringes  Additional inclusion criteria for “seed” participants (at beginning of RDS): recruited by study staff; not transgender (not otherwise explained) | 19% | Colonized:  Mean 43.7 (SD) 12.6) years  Not colonized: Mean 47.6 (SD 11.1) years | Heroin |
| Miller 2007^106^ | - Gender/sex - Age - Race/ethnicity - Income/employment - Unstable housing - Incarceration - Recent hospitalization - Opioid agonist treatment | Not specified. Aimed to identify factors associated with S. aureus colonisation | No estimand | - Colonisation | - MRSA and MSSA colonisation (nasal swab) | USA (Bronx, New York) | 282 | Not reported (though cites prior publication of potential parent study that describes recruiting methadone clinic patients) | 4 | February 1999 to September 2000 | Not reported | 41% | 30 years and younger: 7%  31-45 years: 67%  46 years and older: 26% | None |
| Packer 2019^107^ | - Unstable housing - Public injecting - Recent hospitalization | Not specified. Aimed to identify factors associated with MRSA colonisation | No estimand | - Colonisation | - MRSA colonisation (nasal and groin swabs) | England (Bristol) | 149 | Recruited needle and syringe program clients, using non-probability quota sampling | 4 | 2012 to 2017 | Reported injecting drugs in the past year | 16% | Men:  Median 39 (IQR 34.5-46) years  Women:  Median 40 (IQR 31-45) years | None |

### Appendix 8. Critical appraisal of studies using the Mixed Methods Appraisal Tool (MMAT) for studies where outcome is incident or prevalent injecting-related bacterial infections, included in quantitative systematic review

|  | **SCREENING QUESTIONS** | | **3. NON-RANDOMIZED STUDIES (observational or interventional)** | | | | |
| --- | --- | --- | --- | --- | --- | --- | --- |
| Study | S1. Are there clear research questions? | S2. Do the collected data allow to address the research questions? | 3.1. Are the participants representative of the target population? | 3.2. Are measurements appropriate regarding both the outcome and intervention (or exposure)? | 3.3. Are there complete outcome data? | 3.4. Are the confounders accounted for in the design and analysis? | 3.5. During the study period, is the intervention administered (or exposure occurred) as intended? |
| Baltes 2020 | Yes | Yes | Yes | Yes | Yes | No | No |
| Bassetti 2002 | Yes | Yes | Yes | Yes | Yes | No | Yes |
| Bertin 2020 | Yes | Yes | Yes | Can't tell | Yes | Yes | Yes |
| Betts 2016 | Yes | Yes | Yes | Yes | Yes | Can't tell | Yes |
| Bhattacharya 2006 | Yes | Yes | Can't tell | Can't tell | Yes | No | Yes |
| Binswanger 2000 | Yes | Yes | Yes | Yes | Yes | No | Yes |
| Buchanan 2006 | Yes | Yes | Yes | Yes | Yes | Yes | No |
| Cedarbaum 2016 | Yes | Yes | Yes | Yes | Yes | Yes | Yes |
| Ciccarone 2016 | Yes | Yes | Yes | Yes | Yes | Yes | Yes |
| Cooper 2005 | Yes | Yes | Yes | Yes | Yes | Yes | Yes |
| Dahlman 2015 | Yes | Yes | Yes | Yes | Yes | Yes | No |
| Dahlman 2017 | Yes | Yes | Yes | Yes | Yes | Yes | No |
| DiGiorgio 2019 | Yes | Yes | Can't tell | Yes | Can't tell | No | Yes |
| Doran 2020 (UAM) | Yes | Yes | Yes | Yes | Yes | Yes | No |
| Doran 2020 (C&P) | Yes | Yes | Yes | Yes | Yes | Yes | No |
| Dunleavy 2017 | Yes | Yes | Yes | Yes | Yes | Yes | No |
| Fink 2013 | Yes | Yes | Yes | Yes | Yes | Yes | No |
| Hope 2014 | Yes | Yes | Yes | Yes | Yes | Yes | Yes |
| Hope 2015 | Yes | Yes | Yes | Yes | Yes | Yes | Yes |
| Hope 2010 | Yes | Yes | Yes | Yes | Yes | Yes | No |
| Hope 2008 | Yes | Yes | Yes | Yes | Yes | Yes | No |
| Islam 2019 | Yes | Yes | Yes | Yes | Yes | Yes | No |
| Lee 2013 | Yes | Yes | Yes | Yes | Yes | Yes | Yes |
| Lewer 2020 | Yes | Yes | Yes | Yes | Yes | Yes | Yes |
| Lloyd-Smith 2005 | Yes | Yes | Yes | Yes | Yes | Yes | Yes |
| Lloyd-Smith 2012 | Yes | Yes | Yes | Yes | Yes | Yes | No |
| Lloyd-Smith 2008 | Yes | Yes | Yes | Yes | Yes | Yes | No |
| Lloyd-Smith 2009 | Yes | Yes | Yes | Yes | Yes | Yes | No |
| Lloyd-Smith 2010 | Yes | Yes | Yes | Yes | Yes | Yes | No |
| McMahan 2020 | Yes | Yes | Yes | Yes | Yes | Yes | No |
| Milloy 2010 | Yes | Yes | Yes | Yes | Yes | Yes | No |
| Morin 2020 | Yes | Yes | Yes | No | Yes | Yes | No |
| Murphy 2001 | Yes | Yes | Can't tell | Yes | Yes | No | Yes |
| Nagar 2015 | Yes | Yes | Can't tell | Yes | Yes | No | Yes |
| Noroozi 2019 | Yes | Yes | Yes | Yes | Yes | Yes | No |
| Phillips 2017 | Yes | Yes | Yes | Yes | Yes | Yes | No |
| Phillips 2008 | Yes | Yes | Yes | Yes | Yes | Yes | No |
| Phillips 2010 | Yes | Yes | Yes | Yes | Yes | Yes | No |
| Pollini 2010 | Yes | Yes | Yes | Yes | Yes | Yes | Yes |
| Pollini 2010b | Yes | Yes | Yes | Yes | Yes | Yes | Yes |
| Robertson 2010 | Yes | Yes | Yes | Yes | Yes | Yes | Yes |
| Roux 2020 | Yes | Yes | Yes | Yes | Yes | Yes | No |
| Saeland 2014 | Yes | Yes | Can't tell | Yes | Yes | No | Yes |
| Safaeian 2000 | Yes | Yes | Can't tell | Yes | Yes | Yes | Yes |
| Scherbaum 2010 | Yes | Yes | Yes | Yes | Yes | No | Yes |
| Shah 2020 | Yes | Yes | Can't tell | Yes | Yes | Yes | Yes |
| Sierra 2006 | Yes | Yes | Can't tell | Yes | Yes | No | Yes |
| Silverman 2020 | Yes | Yes | Yes | Yes | Yes | Yes | Yes |
| Smith 2015 | Yes | Yes | Yes | Yes | Yes | Yes | Can't tell |
| Stein 2020 | Yes | Yes | Yes | Yes | Yes | Yes | No |
| Summers 2017 | Yes | Yes | Yes | Yes | Yes | Yes | No |
| Thønnings 2020 | Yes | Yes | Can't tell | Yes | Yes | Yes | Can't tell |
| Tomolillo 2007 (Ecological time series study) | Yes | Yes | Yes | Yes | Yes | No | Yes |
| Tomolillo 2007 (Cross-sectional study) | Yes | Yes | No | Yes | Yes | No | Yes |
| Trayner 2020 | Yes | Yes | Yes | Yes | Yes | Yes | No |
| Weir 2019 | Yes | Yes | Yes | No | Yes | No | Yes |
| Wilson 2002 | Yes | Yes | Yes | Yes | Yes | Yes | No |
| Wright 2020 | Yes | Yes | Yes | Yes | Yes | Yes | No |
| Wurcel 2016 | Yes | Yes | Yes | Yes | Yes | No | Yes |
| Wurcel 2018 | Yes | Yes | Yes | Yes | Yes | Yes | No |
| Yeung 2017 | Yes | Yes | Yes | Yes | Yes | Yes | Yes |
|  | **SCREENING QUESTIONS** | | **2. RANDOMIZED CONTROLLED TRIALS** | | | | |
| **Study** | **S1. Are there clear research questions?** | **S2. Do the collected data allow to address the research questions?** | **2.1. Is randomization appropriately performed?** | **2.2. Are the groups comparable at baseline?** | **2.3. Are there complete outcome data?** | **2.4. Are outcome assessors blinded to the intervention provided?** | **2.5 Did the participants adhere to the assigned intervention?** |
| Oviedo-Joekes 2017 | Yes | Yes | Yes | Yes | Yes | Yes | Yes |

### Appendix 9. Critical appraisal of studies using the Mixed Methods Appraisal Tool (MMAT) for studies where outcome occurs during treatment for injecting-related bacterial infection, included in quantitative systematic review

|  | **SCREENING QUESTIONS** | | **3. NON-RANDOMIZED STUDIES (observational or interventional)** | | | | |
| --- | --- | --- | --- | --- | --- | --- | --- |
| First author, year | S1. Are there clear research questions? | S2. Do the collected data allow to address the research questions? | 3.1. Are the participants representative of the target population? | 3.2. Are measurements appropriate regarding both the outcome and intervention (or exposure)? | 3.3. Are there complete outcome data? | 3.4. Are the confounders accounted for in the design and analysis? | 3.5. During the study period, is the intervention administered (or exposure occurred) as intended? |
| Cooksey 2020 | Yes | No | Yes | Yes | Yes | Yes | Yes |
| Eaton 2020 | Yes | Yes | Can't tell | Can't tell | Can't tell | Yes | Yes |
| Fink 2013 | Yes | Yes | Yes | Yes | Yes | Yes | Can't tell |
| Hope 2008 | Yes | Yes | Yes | Yes | Yes | Yes | No |
| Hope 2015 | Yes | Yes | Yes | Yes | Yes | Yes | Yes |
| Jo 2021 | Yes | Yes | Yes | Yes | Yes | Yes | Yes |
| Kimmel 2020 | Yes | Yes | Yes | Yes | Yes | Yes | Yes |
| Kimmel 2020 | Yes | Yes | Yes | Yes | Yes | Yes | Yes |
| Lloyd-Smith 2012 | Yes | Yes | Yes | Yes | Yes | Yes | No |
| Lloyd-Smith 2010 | Yes | Yes | Yes | Yes | Yes | Yes | No |
| Marks 2020 | Yes | Yes | Yes | Yes | Yes | No | Yes |
| Marks 2020 | Yes | Yes | Yes | Yes | Yes | No | Yes |
| Martín-Dávila 2005 | Yes | Yes | Yes | Yes | Yes | Yes | Yes |
| Meel 2018 | Yes | Yes | Can't tell | Yes | Yes | Yes | Yes |
| Mertz 2008 | Yes | Yes | Can't tell | Yes | Yes | Yes | Yes |
| Monteiro 2020 | Yes | Yes | Yes | Yes | Yes | Yes | No |
| Nolan 2020 | Yes | Yes | Yes | Yes | Yes | Yes | Can't tell |
| Rudasill 2019 | Yes | Yes | Yes | Yes | Yes | No | Yes |
| Sandrock 2001 | Yes | Yes | Yes | Yes | Yes | No | Yes |
| Saydain 2010 | Yes | Yes | No | Yes | Yes | Yes | Yes |
| Serota 2021 | Yes | Yes | Yes | Yes | Yes | Yes | Yes |
| Suzuki 2020 | Yes | Yes | Yes | Yes | Yes | Yes | Yes |
| Takahashi 2007 | Yes | Yes | Can't tell | Yes | Yes | Yes | Yes |
| Tan 2020 | Yes | Yes | Yes | Yes | Yes | Yes | No |
| Uppuluri 2021 | Yes | Yes | Yes | Yes | Yes | Yes | Yes |
| Wang 2020 | Yes | Yes | Yes | Yes | Yes | No | Yes |

### Appendix 10. Critical appraisal of studies using the Mixed Methods Appraisal Tool (MMAT) for studies where outcome occurs after initial treatment for injecting-related bacterial infection, included in quantitative systematic review

|  | **SCREENING QUESTIONS** | | **3. NON-RANDOMIZED STUDIES (observational or interventional)** | | | | |
| --- | --- | --- | --- | --- | --- | --- | --- |
| **Study** | **S1. Are there clear research questions?** | **S2. Do the collected data allow to address the research questions?** | **3.1. Are the participants representative of the target population?** | **3.2. Are measurements appropriate regarding both the outcome and intervention (or exposure)?** | **3.3. Are there complete outcome data?** | **3.4. Are the confounders accounted for in the design and analysis?** | **3.5. During the study period, is the intervention administered (or exposure occurred) as intended?** |
| Barocas 2020 | Yes | Yes | Can't tell | Yes | Yes | Yes | Yes |
| Barocas 2021 | Yes | Yes | Can't tell | Yes | Yes | Yes | Yes |
| Buehrle 2017 | Yes | Yes | Can't tell | Yes | Yes | Yes | Yes |
| Clarelin 2021 | Yes | Yes | Yes | Yes | Yes | Yes | Yes |
| Connell 2010 | Yes | Yes | Yes | Yes | Yes | No | Yes |
| Cooksey 2020 | Yes | Yes | Yes | Yes | Yes | Yes | Yes |
| D’Couto 2018 | Yes | Yes | Yes | Yes | No | No | Yes |
| Hilbig 2020 | Yes | Yes | Yes | Yes | Yes | No | Can't tell |
| Huang 2018 | Yes | Yes | Yes | Yes | Yes | Yes | Yes |
| Jo 2021 | Yes | Yes | Yes | Yes | Yes | Yes | No |
| Kimmel 2020 | Yes | Yes | Yes | Yes | Yes | Yes | Yes |
| Marks 2020a | Yes | Yes | Yes | Yes | Yes | Yes | Yes |
| Marks 2020b | Yes | Yes | Yes | Yes | Yes | No | Yes |
| Marks 2020c | Yes | Yes | Yes | Yes | Yes | No | Yes |
| Nguemeni Tiako 2020 | Yes | Yes | Can't tell | Yes | Yes | No | Yes |
| Pericàs 2021 | Yes | Yes | Can't tell | Yes | Can't tell | Yes | Yes |
| Ray 2020 | Yes | Yes | Yes | Yes | Yes | No | Yes |
| Rodger 2018 | Yes | Yes | Yes | Yes | No | Yes | No |
| Rodger 2019 | Yes | Yes | Yes | Yes | Yes | Yes | Yes |
| Rohn 2020 | Yes | Yes | Can't tell | Yes | Yes | No | Yes |
| Rudasill 2019 | Yes | Yes | Yes | Yes | Yes | Yes | Yes |
| Slaughter 2019 | Yes | Yes | Can't tell | Yes | Can't tell | No | Yes |
| Straw 2020 | Yes | Yes | Yes | Yes | Yes | Yes | Yes |
| Suzuki 2020 | Yes | Yes | Yes | Yes | Can't tell | No | Yes |
| Tan 2020 | Yes | Yes | Yes | Yes | Can't tell | Yes | Yes |
| Thønnings 2020 | Yes | Yes | Can't tell | No | Yes | No | Yes |
| Wang 2020 | Yes | Yes | Yes | Yes | Yes | No | Yes |
| Weymann 2014 | Yes | Yes | Yes | Yes | Yes | Yes | Yes |
|  | **SCREENING QUESTIONS** | | **2. RANDOMIZED CONTROLLED TRIALS** | | | | |
| **Study** | **S1. Are there clear research questions?** | **S2. Do the collected data allow to address the research questions?** | **2.2. Are the groups comparable at baseline?** | **2.3. Are there complete outcome data?** | **2.4. Are outcome assessors blinded to the intervention provided?** | **2.5 Did the participants adhere to the assigned intervention?** | **2.2. Are the groups comparable at baseline?** |
| Fanucchi 2020 | Yes | Yes | Yes | Yes | Yes | Can't tell | Yes |

### Appendix 11. Critical appraisal of studies using the Mixed Methods Appraisal Tool (MMAT) for studies where outcome is colonization with pathogenic bacteria among people who inject drugs, included in quantitative systematic review

|  | **SCREENING QUESTIONS** | | **3. NON-RANDOMIZED STUDIES (observational or interventional)** | | | | |
| --- | --- | --- | --- | --- | --- | --- | --- |
| **Study** | **S1. Are there clear research questions?** | **S2. Do the collected data allow to address the research questions?** | **3.1. Are the participants representative of the target population?** | **3.2. Are measurements appropriate regarding both the outcome and intervention (or exposure)?** | **3.3. Are there complete outcome data?** | **3.4. Are the confounders accounted for in the design and analysis?** | **3.5. During the study period, is the intervention administered (or exposure occurred) as intended?** |
| Colombo 2012 | Yes | Yes | Yes | Yes | Yes | No | Yes |
| Leibler 2019 | Yes | Yes | Yes | Yes | Yes | No | Yes |
| Leung 2015 | Yes | Yes | Yes | Yes | Yes | Yes | Yes |
| Miller 2007 | Yes | Yes | Can't tell | Yes | Yes | Yes | Yes |
| Packer 2019 | Yes | Yes | Yes | Yes | Yes | No | Yes |

### Appendix 12. List of exposure-outcome pair effect estimates for studies where outcome is incident or prevalent injecting-related bacterial infections drug use-associated bacterial and fungal infections, included in quantitative systematic review. Blank cells represent where effect estimate (or frequencies) were not reported in the paper.

| **Study** |  | **Exposures** | **Outcomes** | **Unadjusted effect estimate** | **Adjusted effect estimate** |  |
| --- | --- | --- | --- | --- | --- | --- |
| **Sociodemogaphic factors** | | | | | | |
| Gender/Sex |  |  |  |  |  |  |
| Baltes 2020^1^ |  | Female sex | Past-year SSTI, self report | Female: 11/32 Male: 7/48 |  |  |
| Betts 2016^4^ |  | Female gender | Past-month abscess, self-report |  | aOR 1.47 (1.07 – 2.02) |  |
| Binswanger 2000^6^ |  | Sex (not defined) | Current abscess or cellulitis on physical examination | “did not differ significantly” |  |  |
| Dahlman 2015^11^ |  | Female sex (vs. male) | Ever had an SSTI | OR 4.08 (1.34 – 12.46) | aOR 6.75 (1.40-32.47) |  |
| Dahlman 2017^12^ |  | Female sex (vs. male) | Past-30 day skin and soft-tissue infections | Female: 17/46 (36.9%)  Male: 5/155 (3.2%)  Reported in paper:  OR 0.99 (0.34 – 2.85)  Calculated by me:  17.59 (6.01 – 51.45) |  |  |
| Doran 2020^14^ (UAM) |  | Female gender | Past year SSTI | OR 1.3 (1.1 – 1.5) | aOR 1.4 (1.1 – 1.7) |  |
| Doran 2020^14^ |  | Female gender | Ever SSTI | OR 1.1 (0.7 – 1.7) | aOR 1.4 (0.8 – 2.6) |  |
| Dunleavy 2017^15^ |  | Male vs. female | SSTI in past year | Male: 374/1306 (29%)  Female: 156/553 (28%), p=0.852 |  |  |
| Fink 2013^16^ |  | Gender | Abscess in past 6 months, self-report | Male: 206/612 (33.6%)  Female: 114/245 (46.5%) p=0.0004 | aOR 1.42 (1.01 – 2.00) |  |
| Hope 2014^17^ |  | Gender | Self-reported abscess “(a swelling containing pus)”, past 28 days | “No associations found” |  |  |
| Hope 2014^17^ |  | Gender | Self-reported “redness, swelling, and tenderness”, past 28 days | “No associations found” |  |  |
| Hope 2010^19^ |  | Gender | Self-report, ‘swelling containing pus (abscess), sore, or open wound’ at an injection site during the previous 12 months | Male: 1338/3896 (34%) Female: 525/1313 (40%) p<0.001 | aOR Male: 1.00 Female: 1.43 (1.25-1.64) |  |
| Hope 2010^19^ |  | Gender | Self-report, ‘swelling containing pus (abscess), sore, or open wound’ at an injection site during the previous 12 months  (among subgroup of patients reporting injecting in past 4 weeks) | Male: 989/2820 (35%) Female: 386/913 (42%) p<0.001 | aOR Male: 1.00 Female: 1.41 (1.19-1.66) |  |
| Hope 2008^20^ |  | Gender | Self-reported symptoms of injection site infections (abscess or open wound) in last year | ORs Male: 1.0 Female: 1.4 (1.1 – 1.9) | aORs Male: 1.0 Female: 1.7 (1.2 – 2.4) |  |
| Islam 2019^421^ |  | Gender (female) | Invasive bacterial infection (pneumonia, sepsis, endocarditis) at 9 months (self-report & confirmed with medical chart review)  (among high-frequency injectors [>1 injection per day]) | ORs 1.26 (0.86–1.84) | aORs 1.74 (1.17–2.57) |  |
| Islam 2019^21^ |  | Gender (female) | Invasive bacterial infection (pneumonia, sepsis, endocarditis) at 12 months (self-report & confirmed with medical chart review)  (among high-frequency injectors [>1 injection per day]) | ORs 1.50 (1.14–1.97) | aORs 1.76 (1.34–2.32) |  |
| Lewer 2020^23^ |  | Sex | Rate of hospital admissions for heroin-injection associated bacterial infections | IRR 1.50 (1.32 – 1.69) |  |  |
| Lloyd-Smith 2005^24^ |  | Gender | Self-report abscess (“lasting for more than 3 days”), past 6 months | OR 2.4 (1.9 – 3.0) | aOR 1.7 (1.4 – 2.4) |  |
| Lloyd-Smith 2012^25^ |  | Female | ED visit for cutaneous injecting-related infection (admin data) from SIF/community cohort | OR 1.71 (1.18 – 2.47) |  |  |
| Lloyd-Smith 2008^26^ |  | Sex | Current injecting-related skin infection (self-report & confirmed by study nurse) | OR  Male: Ref  Female: 1.90 (1.39 - 2.58) | aOR 1.68 (1.16 – 2.43) |  |
| Lloyd-Smith 2009^27^ |  | Sex (female) | Injecting-related infection cared for at supervised consumption site (from nursing notes) | OR 1.89 (1.35 – 2.63)  HR 2.08 (1.49 – 2.92) | aHR 1.87 (1.32 – 2.64) |  |
| Lloyd-Smith 2010^28^ |  | Sex (female) | Hospitalization for injecting-related infection (cellulitis, abscess, osteomyelitis, Staph infection, endocarditis, septic arthritis, ulcer, thrombophlebitis, myositis) | HR 1.59 (1.07 – 2.39) | aHR 1.36 (0.90 – 2.05) |  |
| Milloy 2010^30^ |  | Gender | ED visit for cutaneous injecting-related infection (admin data) from SIF/community cohort | At baseline, ED visit in previous 6 month:  Male: 49/380 Female: 23/165  P=0.741  RR for number of ED visits for CIRI during follow-up:  1.08 (0.77–1.53)  (female vs. male) | aRR for number of ED visits for CIRI during follow-up: 1.04 (0.71–1.51) |  |
| Murphy 2001^32^ |  | Sex | ED visit or hospitalization for injecting-related abscess, recruited for this case-control study | Male: 85/289  Female: 66/135  P<0.001 | Not tested |  |
| Roux 2020^42^ |  | Gender | At least one cutaneous abscess in the previous six months | OR Male: Ref  Female or transgender: 0.87 [0.38,2.00] |  |  |
| Phillips 2017^36^ |  | Female | Past year SSTI, self-report | OR 0.80 (0.40, 1.62) | aOR 0.77 (0.29, 2.05) |  |
| Phillips 2008^37^ |  | Gender (male) | ED visit or hospitalization for skin abscess, cellulitis, osteomyelitis, or endocarditis; self-report | OR 1.42 (0.28–7.20) | aOR 1.86; 0.23–15.03 |  |
| Pollini 2010^39^ |  | Gender | Past 6-months abscess | Male: 91/513  Females: 36/110 p<0.001 |  |  |
| Safaeian 2000^44^ |  | Gender | Infective endocarditis, self-report confirmed through medical chart review | OR Male: Ref  Female: 2.8 (1.7 – 4.6) | aOR  3.4 (1.9-6.2) |  |
| Safaeian 2000^44^ |  | Gender | Abscess, self-report | OR Male: Ref  Female: 2.2 (1.6-2.8) | aOR  2.0 (1.5-2.8) |  |
| Shah 2020^46^ |  | Female | Endocarditis | Female: 16/37  Male: 17/98 p=0.002  OR 3.85 (1.68-8.96) | aOR 4.65 (1.85-12.28) |  |
| Sierra 2006^47^ |  | Sex | Invasive soft-tissue Group A Strep (S. pyogenes) infections in Barcelona | Female: Reference  Male: 2.82 (0.4-23.4)  Female: 3/19  Male: 12/54  P=0.27 |  |  |
| Smith 2015^49^ |  | Gender | Current abscess | OR  Male: Reference  Female: 2.56 (1.10 – 5.97) | aOR  2.35 (0.72 – 7.64) |  |
| Stein 2020^50^ |  | Sex (male) | Number of ED visits for injecting-related infections in 12 months following educational intervention |  | IRR 1.18 (0.70, 1.99) |  |
| Stein 2020^50^ |  | Sex (male) | Number of hospitalizations visits for injecting-related infections in 12 months following educational intervention |  | IRR 0.91 (0.55, 1.51) |  |
| Wilson 2002^56^ |  | Female sex | Infective endocarditis, self-report + medical chart review | OR 2.56 (1.54–4.24) | aOR 3.26 (1.73–6.14) |  |
| Wright 2020^57^ |  | Sex | Lifetime SSTI, self-report | Male: 216/341  Female: 75/114 p=0.64 |  |  |
| Wurcel 2016^58^0/0/00 0:00:00 AM |  | Gender | Percentage of hospital admissions for IE that are IDU | “There were no significant changes in IDU-IE hospitalizations by gender over time (data not shown).” |  |  |
| Wurcel 2018^59^ |  | Gender | Abscess ever | Female: 48/88  Male: 77/210 p=0.004 |  |  |
| Wurcel 2018^59^ |  | Gender | Abscess past 30 days | Female: 3/88  Male: 2/210  P=0.044 |  |  |
| Morin 2020^31^ |  | Sex | Infective Endocarditis (diagnostic code in administrative data, but date/timing unclear) | OR  Female: 1.2 (1.1–1.3)  Male: Ref | aOR  Female: 1.2 (1.2–1.3)  Male: Ref |  |
| Morin 2020^31^ |  | Sex | Osteomyelitis (diagnostic code in administrative data, but date/timing unclear) | OR  Female: 1.0 (0.9–1.2)  Male: Ref | aOR  Female: 1.1 (1.0–1.2)  Male: Ref |  |
| Morin 2020^31^ |  | Sex | Septic Arthritis (administrative data, but date/timing unclear) | OR  Female: 1.0 (0.9–1.2)  Male: Ref | aOR  Female: 0.8 (0.7–1.0)  Male: Ref |  |
| Hope 2015^18^ |  | Gender | Abscess, past 12 months, self-reported | “not associated” |  |  |
| Hope 2015^18^ |  | Gender | Cellulitis (redness, swelling, or tenderness), past 12 months, self-reported | “not associated” |  |  |
| **Study** |  | **Exposures** | **Outcomes** | **Unadjusted effect estimate** | **Adjusted effect estimate** |  |
| Age |  |  |  |  |  |  |
| Baltes 2020^1^ |  | Age | Past-year SSTI, self report | 18-29: 4/21  30-39: 7/34  40-49: 6/15  50-59: 1/9  60+: 0/1 |  |  |
| Betts 2016^4^ |  | Age 36-45 (Ref 17-35) | Past-month abscess, self-report |  | aOR  Age 17-35: Reference  Age 36-45: 0.66 (0.44, 0.98) Age 46-71: 0.84 (0.58, 1.22) |  |
| Binswanger 2000^6^ |  | Age (not defined) | Current abscess or cellulitis on physical examination | “did not differ significantly” |  |  |
| Cedarbaum 2016^8^ |  | Age <30 years old (vs. 30 or older) | Abscess in past year, self-report | 22% vs. 48% (p<0.001) | aOR 0.32 (0.16 – 0.65) |  |
| Cooper 2005^10^ |  | Percent neighborhood residents aged 18-64 (vs. <18 or >64) | Monthly rate of hospitalisation for abscess/cellulitis | IRR 0.94 (0.91–0.97) | aIRR 0.96 (0.95–0.98) |  |
| Cooper 2005^10^ |  | Percent neighborhood residents aged 18-64 (vs. <18 or >64) | Monthly rate of hospitalisation for endocarditis | IRR 0.94 (0.92–0.97) | aIRR 0.98 (0.95–1.01) |  |
| Dahlman 2015^11^ |  | Age (years), continuous | Ever had an SSTI | OR 1.00 (0.96 – 1.05) | aOR 1.09 (1.01 – 1.18) |  |
| Dahlman 2017^12^ |  | Age, grouped into 4 groupings (18-29, 30-44, 45-54, 55+) | Past 30-day SSTI | OR not calculated, p = 0.58 |  |  |
| Doran 2020^14^ (UAM) |  | Age (25-34 and 35+ vs. <25 as ref) | Past year SSTI | 25-34 years: OR 1.9 (1.2 – 2.9)  35+ years: OR 2.0 (1.3 – 3.1) | 25-34 years: aOR 3.9 (1.7 – 8.9)  35+ years: aOR 4.4 (2.0 – 10.0) |  |
| Doran 2020^14^ (C&P) |  | Age (25-34 and 35+ vs. <25 as ref) | Ever SSTI | 25-34 years: OR 2.1 (1.1-3.8)  35+ years: OR 3.1 (1.7-5.6) | 25-34 years: aOR 2.2 (1.0 – 5.2)  35+ years: aOR 3.2 (1.4 – 7.1) |  |
| Dunleavy 2017^15^ |  | Age (years) | Past year SSTI | <25: 40/149 (27%) 26-30: 75/277 (27%)  31-35: 116/462 (25%)  >35: 302/980 (31%)  P=0.129 |  |  |
| Fink 2013^16^ |  | Age (years) | Past 6 months abscess, self-reported | <30 years: 15/42 (36%) 30-39 years: 42/139 (30%)  40-49 years: 124/318 (39%)  50+ years: 135/351(38%)  P=0.31 | aOR  <30 years: 1  30-39 years: 0.74 (0.34-1.63)  40-49 years: 1.05 (0.51-2.17)  50+ years: 1.02 (0.49-2.14) |  |
| Hope 2014^17^ |  | Age | Self-reported abscess “(a swelling containing pus)”, past 28 days | “No associations found” |  |  |
| Hope 2014^17^ |  | Age | Self-reported “redness, swelling, and tenderness”, past 28 days | “No associations found” |  |  |
| Hope 2010^19^ |  | Age | Self-report, ‘swelling containing pus (abscess), sore, or open wound’ at an injection site during the previous 12 months | <25: 249/821 (30%) 25-29: 397/1214 (33%) 30-34: 410/1174 (35%) 35-39: 419/1045 (40%) >=40: 388/955 (41% p<0.001 | “Not in final multivariable model” |  |
| Hope 2008^20^ |  | Age in years | Self-reported symptoms of injection site infections (abscess or open wound), past year | ORs <25: 1.0 25-29: 1.8 (1.2 – 2.7) 30-34: 2.1 (1.4 – 3.2) 35+: 1.9 (1.3 – 2.9) | aORs <25: 1.0 25-29: 1.6 (1.0 – 2.6) 30-34: 2.0 (1.3 – 3.2) 35+: 1.9 (1.2 – 3.0) |  |
| Islam 2019^21^ |  | Age (per 10 years) | Invasive bacterial infection (pneumonia, sepsis, endocarditis) at 9 months (self-report & confirmed with medical chart review)  (among high-frequency injectors [>1 injection per day]) | ORs 1.18 (0.95–1.46) | aORs 1.30 (1.03–1.66) |  |
| Islam 2019^21^ |  | Age (per 10 years) | Invasive bacterial infection (pneumonia, sepsis, endocarditis) at 12 months (self-report & confirmed with medical chart review)  (among high-frequency injectors [>1 injection per day]) | ORs 1.21 (1.04–1.42) | aORs 1.34 (1.13–1.59) |  |
| Lloyd-Smith 2012^25^ |  | Age, per year older | ED visit for cutaneous injecting-related infection (admin data) from SIF/community cohort | HR, only separate models by sex/gender  Among females:  HR 1.00 (0.97 – 1.02)  Among males:  HR 0.99 (0.97 – 1.01) |  |  |
| Lloyd-Smith 2008^26^ |  | Age, per year older | Current injecting-related skin infection (self-report & confirmed by study nurse) | OR 0.98 (0.96 – 1.00) | aOR 1.00 (0.98 – 1.02) |  |
| Lloyd-Smith 2009^27^ |  | Age, per year older | Injecting-related infection cared for at supervised consumption site (from nursing notes) | OR 0.99 (0.97 – 1.01)  HR 0.99 (0.97 – 1.00) |  |  |
| Lloyd-Smith 2010^28^ |  | Age, per year older | Hospitalization for injecting-related infection (cellulitis, abscess, osteomyelitis, Staph infection, endocarditis, septic arthritis, ulcer, thrombophlebitis, myositis) | HR 0.98 (0.96 – 1.01) |  |  |
| Milloy 2010^30^ |  | Age, per year older | ED visit for cutaneous injecting-related infection (admin data) from SIF/community cohort | At baseline, ED visit in previous 6 month:  Median (IQR):  CIRI: 37.5 (32.8-42.3)  No CIRI: 39.9 (33.7–46.1)  p=0.194  RR for number of ED visits for CIRI during follow-up:  1.00 (0.98–1.02) | aRR for number of ED visits for CIRI during follow-up: 1.01 (0.99–1.03) |  |
| Murphy 2001^32^ |  | Age | ED visit or hospitalization for injecting-related abscess, recruited for this case-control study | <30: 16/51  30-39: 47/111  40-49: 67/189  50+: 21/73 p=0.73 |  |  |
| Noroozi 2019^34^ |  | Age | Lifetime injection site infection | ORs  <30: 1 30-39: 1.3 (0.1-5.3)  40+: 1.4 (0.2-4.8) | aORs  <30: 1 30-39: 1.2 (0.56-2.5)  40+: 1.6 (0.47-4.5) |  |
| Roux 2020^42^ |  | Age, years | At least one cutaneous abscess in the previous six months | OR 1.01 [0.97,1.05] |  |  |
| Phillips 2017^36^ |  | Age, years | Past year SSTI, self-report | OR 0.99 (0.96, 1.03) | aOR 1.03 (0.97, 1.08) |  |
| Phillips 2008^37^ |  | Age, years | ED visit or hospitalization for skin abscess, cellulitis, osteomyelitis, or endocarditis; self-report | OR 1.03 (0.96–1.11) | aOR 1.03 (0.96–1.11) |  |
| Phillips 2010^38^ |  | Age, years | Past year SSTI, self-report | OR 0.98; 0.92–1.04 |  |  |
| Pollini 2010^39^ |  | Age, years | Past 6-months abscess | Median age (IQR) Abscess: 39 (33-43)  No abscess: 37 (32-43) p=0.14 |  |  |
| Saeland 2014^43^ |  | Age, years | Current abscess. Self-report and confirmed by physical examination. | Mean age (SD)  Abscess: 36.9 (7.7)  No abscess: 35.1 (7.6) p=0.181 |  |  |
| Safaeian 2000^44^ |  | Age | Infective endocarditis, self-report confirmed through medical chart review | OR <34 years: Ref  >34 years: 1.1 (0.6-2.0)  [unclear what happens to people who are exactly 34] |  |  |
| Safaeian 2000^44^ |  | Age | Abscess, self-report | OR <34 years: Ref  >34 years: 1.4 (1.0-1.8)  [unclear what happens to people who are exactly 34] |  |  |
| Shah 2020^46^ |  | Age, years | Endocarditis | Mean (SD) Endocarditis: 35.5 (8.4)  No endocarditis: 40.0 (11.0) p=0.034 |  |  |
| Sierra 2006^47^ |  | Age, years | Invasive soft-tissue Group A Strep (S. pyogenes) infections in Barcelona | Mean (range)  Cases: 30.1 (22-41) Controls: 27.5 (20-43)  P=0.9 |  |  |
| Stein 2020^50^ |  | Age | Number of ED visits for injecting-related infections in 12 months following educational intervention |  | IRR 1.10 (0.97, 1.02) |  |
| Stein 2020^50^ |  | Age | Number of hospitalizations visits for injecting-related infections in 12 months following educational intervention |  | IRR 1.01 (0.99, 1.03) |  |
| Summers 2017^51^ |  | Age (categories undefined) | Past year abscess, self-report | OR 1.05 [1.02, 1.09] | aOR 1.06 [0.97, 1.15] |  |
| Thønnings 2020^52^ |  | Age (years) | Bacteraemia, among hospitalised PWID | OR 1.02 (0.99-1.05) | aOR 3.46 (1.30 – 9.23) |  |
| Wilson 2002^56^ |  | Age, >38 years | Infective endocarditis, self-report + medical chart review | OR 1.11 (0.67–1.83) |  |  |
| Wright 2020^57^ |  | Age | Lifetime SSTI, self-report | OR <35 years: Ref  35-44 years: 1.92 (1.05 – 3.51)  45+ years: 3.32 (1.90 – 5.82) | aOR <35 years: Ref  35-44 years: 2.03 (1.11–3.71)  45+ years: 3.68 (2.09–6.50) |  |
| Wurcel 2016^58^ |  | Age groups, over time | Percentage of hospital admissions for IE that are IDU | “…the percentage of IDU-IE hospitalizations among young adults (15–34 years) steadily increased from 2000 to 2013, with a steep increase from 2008 to 2013 (27.7%–42.0%; P < .001 using χ2 test for trend in proportions). In contrast, IDU-IE rates among middle-aged adults (ages 35–54) steadily decreased between 2000 and 2013 (67.2%–39.9%; P < .001).” |  |  |
| Wurcel 2018^59^ |  | Age, years | Abscess ever | OR  Among females: 0.97 (0.90, 1.04)  Among males: 0.96 (0.92, 1.01) | aOR  Among females: 0.89 (0.77, 1.02)  Among males: 0.96 (0.88, 1.04) |  |
| Morin 2020^31^ |  | Age, years | Infective Endocarditis (diagnostic code in administrative data, but date/timing unclear) | OR  15-24: 0.9 (0.8–0.9)  25-34: 1.4 (1.3–1.6)  35-44: 2.4 (2.2–2.7)  45-54: 4.2 (3.8–4.7)  55-65: 10.0 (8.7–11.6)  65+: Reference | aOR  15-24: 0.9 (0.8–0.9)  25-34: 1.4 (1.3–1.6)  35-44: 2.3 (2.1–2.6)  45-54: 3.9 (3.5–4.3)  55-65: 8.6 (7.4-10.0)  65+: Reference |  |
| Morin 2020^31^ |  | Age, years | Osteomyelitis (diagnostic code in administrative data, but date/timing unclear) | OR  15-24: 1.4 (1.1–1.7)  25-34: 2.9 (2.3–3.6)  35-44: 4.4 (3.5–5.5)  45-54: 7.0 (5.4–9.0)  55-65: 8.3 (5.8–11.8)  65+: Reference | aOR  15-24: 1.4 (1.1–1.7)  25-34: 2.8 (2.2–3.5)  35-44: 3.9 (3.1–5.0)  45-54: 6.0 (4.7–7.8)  55-65: 6.6 (4.6-9.4)  65+: Reference |  |
| Morin 2020^31^ |  | Age, years | Septic Arthritis (administrative data, but date/timing unclear) | OR  15-24: 1.3 (0.9–1.8)  25-34: 3.1 (2.2–4.3)  35-44: 3.9 (2.8–5.4)  45-54: 5.1 (3.5-7.4)  55-65: 10.2 (6.4-16.1)  65+: Ref | aOR  15-24: 1.3 (0.9–1.8)  25-34: 2.7 (2.0–3.8)  35-44: 3.2 (2.3–4.4)  45-54: 4.1 (2.8–6.1)  55-65: 8.3 (5.2–13.3)  65+: Ref |  |
| Hope 2015^18^ |  | Age, years | Abscess, past 12 months, self-reported | <25: 8/113  25-29: 46/263  30-34: 35/186  >=35: 68/293 |  |  |
| Hope 2015^18^ |  | Age, years | Cellulitis (redness, swelling, or tenderness), past 12 months, self-reported | “not associated” |  |  |
| **Study** |  | **Exposures** | **Outcomes** | **Unadjusted effect estimate** | **Adjusted effect estimate** |  |
| Race/Ethnicity | | | | | | |
| Baltes 2020^1^ |  | Race | Past-year SSTI, self report | Caucasian: 14/62  African American: 2/4  American Indian: 1/11  Mixed race: 1/3 |  |  |
| Cooper 2005^10^ |  | Percent residents non-Hispanic white (continuous variable) | Monthly rate of hospitalisation for abscess/cellulitis | IRR 0.99 (0.98–1.00) | aIRR 0.99 (0.98–1.00) |  |
| Cooper 2005^10^ |  | Percent residents non-Hispanic white (continuous variable) | Monthly rate of hospitalisation for endocarditis | IRR 0.98 (0.98–0.99) | aIRR 0.99 (0.98–1.00) |  |
| Dahlman 2017^12^ |  | Race, in multiple groups (White, Black, Hispanic, Other, Refused to answer) | Past 30 day SSTI | No effect size calculated, p=0.44 for distribution?  Calculating Non-Hispanic White vs. else: OR 0.70 (0.28 – 1.76) |  |  |
| Doran 2020^14^ (C&P) |  | Race (White/White British vs. other as ref) | Ever SSTI | OR 1.5 (0.9 – 2.4) |  |  |
| Fink 2013^16^ |  | Race | Past 6 months SSTI, self-report | Black: 116/311 (37%)  White: 66/182 (36%) Latino: 110/302 (36%) Other: 20/48 (42%)  P=0.91  Calculating Non-Hispanic White vs. else: OR 0.96 (0.68-1.35) | Black: 0.95 (0.62-1.45)  White: 1  Latino: 0.87 (0.57-1.31)  Other: 1.32 (0.66-2.64) |  |
| Milloy 2010^30^ |  | Ethnicity (Aboriginal vs. other) | ED visit for cutaneous injecting-related infection (admin data) from SIF/community cohort | At baseline, ED visit in previous 6 month:  Non-aboriginal: 60/440  Aboriginal: 12/105 p=0.548  RR for number of ED visits for CIRI during follow-up:  0.72 (0.47–1.09)  (aboriginal vs. other) | aRR for number of ED visits for CIRI during follow-up: 0.71 (0.47–1.07) |  |
| Murphy 2001^32^ |  | Race | ED visit or hospitalization for injecting-related abscess, recruited for this case-control study | White: 86/228 Black: 42/140  Hispanic: 14/31 Asian/other: 9/25 p=0.86 | Not tested |  |
| Phillips 2017^36^ |  | Non-Latino Caucasian (vs. other) | Past year SSTI, self-report | OR 1.29 (0.64, 2.58) | aOR 0.97 (0.39, 2.41) |  |
| Phillips 2008^37^ |  | Race (Caucasian vs. other) | ED visit or hospitalization for skin abscess, cellulitis, osteomyelitis, or endocarditis; self-report | OR 6.91; 0.83–57.20 | aOR 6.71; 0.81–55.82 |  |
| Phillips 2010^38^ |  | Race (Caucasian vs. other) | Past year SSTI, self-report | OR 0.36; 0.06–2.05 |  |  |
| Safaeian 2000^44^ |  | Race | Infective endocarditis, self-report confirmed through medical chart review | Other: 0/34  Black: 86/533  [Study did not calculate/report OR because 100% of cases were Black people] |  |  |
| Safaeian 2000^44^ |  | Race | Abscess, self-report | Other: Ref  Black: 1.8 (1.0-3.3) |  |  |
| Shah 2020^46^ |  | Caucasian | Endocarditis | Caucasian: 28/102  Other: 5/33 p=0.17 |  |  |
| Smith 2015^49^ |  | Race | Current abscess | OR  African American: Reference  Caucasian: 2.20 (0.88 – 5.49)  Native American: 7.50 (0.92 – 60.90) | OR  African American: Reference  Caucasian: 2.21 (0.64–7.58)  Native American: 7.35 (0.48–113.36) |  |
| Stein 2020^50^ |  | Non-Latinx white | Number of ED visits for injecting-related infections in 12 months following educational intervention |  | IRR 0.73 (0.44, 1.21) |  |
| Stein 2020^50^ |  | Non-Latinx white | Number of hospitalizations visits for injecting-related infections in 12 months following educational intervention |  | IRR 0.91 (0.56, 1.49) |  |
| Summers 2017^51^ |  | White race | Past year abscess, self-report | OR 0.62 [0.30, 1.28] | aOR 1.34 [0.39, 4.61] |  |
| Wurcel 2016^58^ |  | Race | Percentage of hospital admissions for IE that are IDU | “Injection drug use-related IE increased in whites from 40.2% in 2000 to 68.9% in 2013 (P < .001) (Figure 1C).”  “Non-white” category appeared stable, but missing data category hugely dropped off so that may explain part of increase in white people? |  |  |
| Wurcel 2018^59^ |  | White | Abscess ever | OR  Among females: 1.82 (0.53, 6.27)  Among males: 1.61 (0.89, 2.90) | aOR  Among females: 2.04 (0.29, 14.22)  Among males: 1.35 (0.65, 2.78) |  |
| Education |  |  |  |  |  |  |
| Baltes 2020^1^ |  | Education | Past-year SSTI, self report | Less than high school: 3/15  High school equivalent: 8/31  Some college: 7/29  Associate’s degree: 0/4  Bachelor’s degree or higher: 0/1 |  |  |
| Betts 2016^4^ |  | No tertiary education (vs. tertiary education) | Past-month abscess, self-report |  | aOR 0.74 (0.55, 1.01) |  |
| Fink 2013^16^ |  | High school education or greater (vs. less than high school) | Past 6-month abscess, self-report | Yes: 186/512 No: 140/384, p=0.46 |  |  |
| Murphy 2001^32^ |  | Education | ED visit or hospitalization for injecting-related abscess, recruited for this case-control study | Less than high school: 66/178 High school graduate: 85/246 p=0.62 |  |  |
| Noroozi 2019^34^ |  | Education | Lifetime injection site infection | High school diploma or less: 90/210  More than high school: 110/290  P=0.02 |  |  |
| Roux 2020^42^ |  | Education | At least one cutaneous abscess in the previous six months | Less than high school diploma: Ref  High school diploma or more: 1.43 [0.75,2.71]  OR |  |  |
| Phillips 2017^36^ |  | Education | Past year SSTI, self-report | OR  High school diploma or more: 2.43 (1.20, 4.90) | aOR 4.81 (1.89, 12.3) |  |
| Saeland 2014^43^ |  | Years in school | Current abscess. Self-report and confirmed by physical examination. | Median (IQR)  Abscess: 11.0 (9.0, 13.0)  No abscess: 11.0 (9.0, 12.0)  P=0.617 |  |  |
| Safaeian 2000^44^ |  | Education | Infective endocarditis, self-report confirmed through medical chart review | OR No HS diploma: Ref  HS diploma: 0.9 (0.6-1.4) |  |  |
| Safaeian 2000^44^ |  | Education | Abscess, self-report | OR No HS diploma: Ref  HS diploma: 0.8 (0.6-1.0) |  |  |
| Shah 2020^46^ |  | Completion of secondary | Endocarditis | Completion: 19/50  No completion: 12/73 p=0.010 |  |  |
| Wilson 2002^56^ |  | Education >= 12 years | Infective endocarditis, self-report + medical chart review | OR 0.87 (0.53–1.43) |  |  |
| Wurcel 2018^59^ |  | High school education or greater | Abscess ever | OR  Among females: 0.46 (0.17, 1.21)  Among males: 0.75 (0.42, 1.36) | aOR  Among females: 0.28 (0.08, 1.08)  Among males: 0.48 (0.23, 0.99) |  |
| **Study** |  | **Exposures** | **Outcomes** | **Unadjusted effect estimate** | **Adjusted effect estimate** |  |
| Poverty/income/employment | | | | | | |
| Ciccarone 2016^9^ |  | Percent unemployment & percent povery  (Not otherwise defined) | Proportion of opiate-related hospital admissions comprised of skin and soft-tissue infections | Presented only in figure and does not list numbers.  It seems like “Percent unemployment” had point estimate of OR 1.00 and was not statistically significant, and “Percent poverty” had point estimate of OR 0.98 with p<0.05 | Unclear / uninterpretable |  |
| Cooper 2005^10^ |  | >/=20% of neighbourhood residents living below poverty level | Monthly rate of hospitalisation for abscess/cellulitis | IRR 1.88 (1.54–2.29) | aIRR 0.67 (0.41–1.09) |  |
| Cooper 2005^10^ |  | >/=20% of neighbourhood residents living below poverty level | Monthly rate of hospitalisation for endocarditis | IRR 1.83 (1.51–2.23) | aIRR 0.93 (0.50–1.74) |  |
| Doran 2020^14^ (C&P) |  | Main Income source (Social Welfare/Illicit Activities/Other vs. Regular/Temporary Job/Family Support as ref) | Ever SSTI | OR 2.0 (1.1-3.7) | aOR 2.2 (1.0-4.9) |  |
| Hope 2014^17^ |  | Main source of income (licit vs. illicit) | Self-reported abscess “(a swelling containing pus)”, past 28 days | Licit: 28/588 (4.8%)  Illicit: 24/267 (9.0%) |  |  |
| Hope 2014^17^ |  | Main source of income (licit vs. illicit) | Self-reported “redness, swelling, and tenderness”, past 28 days | Licit: 109/588 (19%) Illicit: 68/267 (25%) p=0.023 |  |  |
| Hope 2015^18^ |  | Main source of income (licit vs. illicit) | Abscess, past 12 months, self-reported | Licit: 92/588  Illicit: 68/267 |  |  |
| Hope 2015^18^ |  | Main source of income (licit vs. illicit) | Cellulitis (redness, swelling, or tenderness), past 12 months, self-reported | “Not associated” |  |  |
| Murphy 2001^32^ |  | Annual family income | ED visit or hospitalization for injecting-related abscess, recruited for this case-control study | <$10,000: 125/334  $10,000 to $19,999: 20/70  >=$20,000: 6/20 p=0.24 |  |  |
| Noroozi 2019^34^ |  | Socioeconomic status | Lifetime injection site infection | ORs  Low: 3.3 (1.6- 6.4)  Moderate: 1.8 (1.2- 4.7)  High: 1 | aORs  Low: 2.4 (1.4- 3.8) Moderate: 1.3 (1.14- 2.16)  High: 1 |  |
| Noroozi 2019^34^ |  | Employment status | Lifetime injection site infection | Employed: 94/250  Uneployed: 106/250  P=0.02 |  |  |
| Noroozi 2019^34^ |  | Monthly income | Lifetime injection site infection | Less than USD$150: 114/228  USD$150+: 86/272  p=0.01 |  |  |
| Pollini 2010^39^ |  | Income through informal work/odd jobs (vs. not?) | Past 6-months abscess | Yes: 75/377  No: 52/246  P=0.74 |  |  |
| Pollini 2010^39^ |  | Income through legal job with pau (vs. not?) | Past 6-months abscess | Yes: 24/99  No: 103/524 P=0.33 |  |  |
| Roux 2020^42^ |  | Employment | At least one cutaneous abscess in the previous six months | OR  No: Ref  Yes: 0.84 [0.42,1.66] |  |  |
| Safaeian 2000^44^ |  | “Socioeconomic status” (NOS) | Infective endocarditis, self-report confirmed through medical chart review | OR <$5,000 U.S.: Ref  >=$5,000 U.S.: 0.7 (0.4 – 1.3) |  |  |
| Safaeian 2000^44^ |  | “Socioeconomic status” (NOS) | Abscess, self-report | OR <$5,000 U.S.: Ref  >=$5,000 U.S.: 0.7 (0.5 – 0.9) |  |  |
| Shah 2020^46^ |  | Employed or seasonally employed | Endocarditis | Employed: 3/9  Not employed: 27/121 p=0.43 |  |  |
| Summers 2017^51^ |  | Reported income (categories undefined)… could literally be continuous dollars? | Past year abscess, self-report | OR 1.00 (1.00, 1.00)  P=0.61 | aOR 1.00 (1.00, 1.00)  p=0.71 |  |
| Morin 2020^31^ |  | Neighbourhood Income | Infective Endocarditis (diagnostic code in administrative data, but date/timing unclear) | OR  5 (highest): Ref  4: 0.9 (0.8–1.0)  3: 0.9 (0.8–1.0)  2: 0.9 (0.8–1.0)  1 (lowest): 0.9 (0.9–1.0) | aOR  5 (highest): Ref  4: 0.9 (0.8–1.0)  3: 1.0 (0.9–1.1)  2: 0.9 (0.9–1.1)  1 (lowest): 1.0 (0.9-1.1) |  |
| Morin 2020^31^ |  | Neighbourhood Income | Osteomyelitis (diagnostic code in administrative data, but date/timing unclear) | OR  5 (highest): Ref  4: 1.1 (0.9–1.4)  3: 1.0 (0.8–1.3)  2: 1.2 (0.9–1.5)  1 (lowest): 1.3 (1.1–1.6) | aOR  5 (highest): Ref  4: 4.2 (3.0–5.8)  3: 1.0 (0.8–1.3)  2: 1.2 (0.9–1.4)  1 (lowest): 1.3 (1.0–1.6) |  |
| Morin 2020^31^ |  | Neighbourhood Income | Septic Arthritis (administrative data, but date/timing unclear) | OR  5 (highest): Ref  4: 1.0 (0.7–1.4)  3: 1.1 (0.8–1.5)  2: 1.3 (0.9–1.7)  1 (lowest): 1.2 (0.9–1.6) | aOR  5 (highest): Ref  4: 1.0 (0.7–1.4)  3: 1.0 (0.7–1.4)  2: 1.1 (0.8–1.6)  1 (lowest): 1.0 (0.8–1.4) |  |
| Relationship status | | | | | | |
| Noroozi 2019^34^ |  | Marital status | Lifetime injection site infection | Single: 106/262  Married: 94/238  P=0.4 |  |  |
| Roux 2020^42^ |  | Living in a couple | At least one cutaneous abscess in the previous six months | OR  No: Ref  Yes: 0.35 [0.17,0.71] | aOR 0.38 [0.17,0.85] |  |
| Migration history | | | | | | |
| Doran 2020^14^ (UAM) |  | Born in UK (yes vs. no) | Past year SSTI | OR 1.2 (0.8 0 1.6) |  |  |
| Hope 2015^18^ |  | Migration, years lived in current area | Abscess, past 12 months, self-reported | <= 1: 11/101 (10.9%)  2-10: 50/198 (25%)  11-20: 19/99 (19%)  >=21: 80/457 (18.7%) p=0.019 |  |  |
| Hope 2015^18^ |  | Migration, years lived in current area | Cellulitis (redness, swelling, or tenderness), past 12 months, self-reported | “Not associated” |  |  |
| **Social support and housing characteristics** | | | | | | |
| Sex work | | | | | | |
| Doran 2020^14^ |  | Taken part in transactional sex (Yes in past year or Yes but not in past year, vs. never as ref) | Past year SSTI | Yes, in past year: OR 1.2 (1.0 – 1.1)  Yest but not in past year: OR 1.4 (1.0-2.0) |  |  |
| Hope 2015^18^ |  | Sex, preceding year (paid) | Abscess, past 12 months, self-reported | No: 45/175  Yes, but not paid: 100/626  Yes, but paid: 15/54 | No: Ref  Yes, but not paid: 0.59 (0.39 – 0.90)  Yes, but paid: 1.08 (0.53 – 2.20) |  |
| Hope 2015^18^ |  | Sex, preceding year (paid | Cellulitis (redness, swelling, or tenderness), past 12 months, self-reported | No: 72/175  Yes, but not paid: 311/626  Yes, but paid: 31/54 p=0.054 |  |  |
| Lloyd-Smith 2005^24^ |  | Sex trade involved, past 6 months | Self-report abscess (“lasting for more than 3 days”), past 6 months | OR 2.4 (1.9 – 3.1) | aOR 1.5 (1.1 – 2.1) |  |
| Lloyd-Smith 2008^26^ |  | Sex trade, past 6 months | Current injecting-related skin infection (self-report & confirmed by study nurse) | OR 1.74 (1.24 – 2.45) | aOR 1.02 (0.67 – 1.56) |  |
| Milloy 2010^30^ |  | Sex-trade participation, past 6 months | ED visit for cutaneous injecting-related infection (admin data) from SIF/community cohort | At baseline, ED visit in previous 6 month:  No: 60/447 Yes: 12/98 p=0.755  RR for number of ED visits for CIRI during follow-up:  1.40 (1.08–1.84) | aRR for number of ED visits for CIRI during follow-up: 1.48 (1.10–1.98) |  |
| Pollini 2010^39^ |  | Principal source of income was through sex work | Past 6-months abscess | Yes: 17/31  No: 110/592 P<0.01 | aOR 4.56 (2.08 – 10.00) |  |
| Pollini 2010^39^ |  | Traded sex for money or drugs, past 6 mos. | Past 6-months abscess | Yes: 18/38  No: 109/585  P<0.01 |  |  |
| Saeland 2014^43^ |  | Sex trade involvement (NOS) | Current abscess. Self-report and confirmed by physical examination. | “did not differ” (data not shown) |  |  |
| Wurcel 2018^59^ |  | Sex work | Abscess ever | OR  Among females: 2.19 (0.83, 5.81)  Among males: 0.60 (0.26, 1.37) | aOR  Among females: 5.42 (1.27, 23.10)  Among males: 0.49 (0.20, 1.21) |  |
| **Study** |  | **Exposures** | **Outcomes** | **Unadjusted effect estimate** | **Adjusted effect estimate** |  |
| Incarceration history | | | | | | |
| Doran 2020^14^ (UAM) |  | Ever imprisoned | Past year SSTI | OR 1.2 (1.1 – 1.5) |  |  |
| Dunleavy 2017^15^ |  | Ever in prison | Past year SSTI | Yes: 356/1238 (29%) No: 175/622 (28%), p=0.780 |  |  |
| Hope 2014^17^ |  | Imprisonment last year | Self-reported abscess “(a swelling containing pus)”, past 28 days | “No associations found” |  |  |
| Hope 2014^17^ |  | Imprisonment last year | Self-reported “redness, swelling, and tenderness”, past 28 days | “No associations found” |  |  |
| Hope 2015^18^ |  | Imprisonment (Never; Yes, not preceding year; Yes, preceding year) | Abscess, past 12 months, self-reported | “not associated” |  |  |
| Hope 2015^18^ |  | Imprisonment (Never; Yes, not preceding year; Yes, preceding year) | Cellulitis (redness, swelling, or tenderness), past 12 months, self-reported | “not associated” |  |  |
| Hope 2010^19^ |  | Ever imprisoned | Self-report, ‘swelling containing pus (abscess), sore, or open wound’ at an injection site during the previous 12 months | No: 536/1570 (34%) Yes, not last year: 844/2289 (37%) Yes, in last year: 469/1315 (36%) p=0.22 |  |  |
| Hope 2008^20^ |  | Having been imprisoned | Self-reported symptoms of injection site infections (abscess or open wound), past year | “…was not associated with…” |  |  |
| Lloyd-Smith 2005^24^ |  | Recent incarceration, past 6 months | Self-report abscess (“lasting for more than 3 days”), past 6 months | OR 1.7 (1.3 – 2.1) | aOR 1.7 (1.3 – 2.2) |  |
| Milloy 2010^30^ |  | Recent incarceration, past 6 months | ED visit for cutaneous injecting-related infection (admin data) from SIF/community cohort | At baseline: Yes: 31/187  No: 41/358 p=0.093  RR for number of ED visits for CIRI during follow-up:  1.56 (1.32 – 1.85) | aRR for number of ED visits for CIRI during follow-up: 1.56 (1.31-1.85) |  |
| Pollini 2010^39^ |  | Incarcerated, past 6 mos. | Past 6-months abscess | Yes: 72/320  No: 55/303  P=0.14 |  |  |
| Saeland 2014^43^ |  | Imprisonment (NOS) | Current abscess. Self-report and confirmed by physical examination. | “did not differ” (data not shown) |  |  |
| Injected during incarceration | | | | | | |
| Pollini 2010^39^ |  | Injected during incarceration, past 6 mos. | Past 6-months abscess | Yes: 22/86  No: 105/537  P=0.18 |  |  |
| **Study** |  | **Exposures** | **Outcomes** | **Unadjusted effect estimate** | **Adjusted effect estimate** |  |
| Unstable housing or homelessness | | | | | | |
| Baltes 2020^1^ |  | Homelessness (past 6 months) | Past-year SSTI, self report | Yes: 11/52  No: 7/28 |  |  |
| Betts 2016^4^ |  | Unstable housing (vs. stable housing), self-report | Past-month abscess, self-report |  | aOR 1.39 (0.97, 1.99) |  |
| Binswanger 2000^6^ |  | Homelessness (not otherwise specified) | Current abscess or cellulitis on physical examination | “did not differ significantly” |  |  |
| Dahlman 2017^12^ |  | Homeless | Past 30-day skin and soft-tissue infections | OR 1.25 (0.49 – 3.23) |  |  |
| Doran 2020^14^ |  | Homeless (Street or Hostels) | Past year SSTI | Yes, in past year: OR 1.2 (1.0 – 1.1)  *Note that OR is not in between CI…  Yes, but not in past year: OR 1.2 (0.9 – 1.4) |  |  |
| Doran 2020^14^ |  | Ever Street Homeless | Ever SSTI | OR 1.2 (0.8 – 1.9) |  |  |
| Dunleavy 2017^15^ |  | Ever homeless | SSTI in past year | Yes: 414/1435 (29%) No: 118/430 (27%), p=0.57 |  |  |
| Dunleavy 2017^15^ |  | Homeless in past 6 months | SSTI in past year | Yes: 143/529 (27%) No: 388/1333 (29%), p=0.371 |  |  |
| Fink 2013^16^ |  | Homeless | Past 6-month abscess, self-report | Yes: 175/460 (38%) No: 140/384 (36%) |  |  |
| Hope 2014^17^ |  | Homelessness | Self-reported abscess “(a swelling containing pus)”, past 28 days | “No associations found” |  |  |
| Hope 2014^17^ |  | Homelessness | Self-reported “redness, swelling, and tenderness”, past 28 days | Never: 13/106 (12%) Yes, not last year: 64/319 (20%) Yes, last year: 101/430 (23%) p=0.036 |  |  |
| Hope 2015^18^ |  | Homelessness | Abscess, past 12 months, self-reported | “not associated” |  |  |
| Hope 2015^18^ |  | Homelessness | Cellulitis (redness, swelling, or tenderness), past 12 months, self-reported | Never: 33/106  Yes, not preceding year: 156/319  Yes, preceding year: 224/430  P=0.001 | Never: 1.00  Yes, not preceding year: 2.01 (1.24 – 3.23)  Yes, preceding year: 2.16 (1.36 – 3.45) |  |
| Hope 2010^19^ |  | Homeless last year | Self-report, ‘swelling containing pus (abscess), sore, or open wound’ at an injection site during the previous 12 months | No: 1022/3015 (34%) Yes: 841/2194 (38%) p value not reported | No: 1.00 Yes: 1.18 (1.05-1.33) |  |
| Hope 2010^19^ |  | Homeless last year | Self-report, ‘swelling containing pus (abscess), sore, or open wound’ at an injection site during the previous 12 months  (among subgroup of participants reporting injecting in past 4 weeks) | Yes: 629/1588 (40%)  No: 746/2145 (35%) p= 0.002 |  |  |
| Hope 2008^20^ |  | Homeless | Self-reported symptoms of injection site infections (abscess or open wound), past year | ORs Never: 1.0 Over a year ago: 1.7 (1.1 – 2.7)  In last year: 1.9 (1.2 – 2.9) |  |  |
| Lloyd-Smith 2005^24^ |  | Unstable housing, past 6 months | Self-report abscess (“lasting for more than 3 days”), past 6 months | OR 1.3 (1.1 – 1.8) |  |  |
| Lloyd-Smith 2012^25^ |  | Unstable housing, past 6 months | ED visit for cutaneous injecting-related infection (admin data) from SIF/community cohort | Only separate models by sex/gender  Among females:  HR 1.68 (1.09 – 2.61)  Among males:  HR 1.60 (1.18 – 2.17) | Only separate models by sex/gender  Among females:  aHR 1.12 (0.69 – 1.82)  Among males: aHR 1.37 (0.98 – 1.92) |  |
| Lloyd-Smith 2008^26^ |  | Unstable housing, current | Current injecting-related skin infection (self-report & confirmed by study nurse) | OR 1.56 (1.15 – 2.12) | aOR 1.49 (1.10 – 2.03) |  |
| Lloyd-Smith 2009^27^ |  | Unstable housing, past 6 months | Injecting-related infection cared for at supervised consumption site (from nursing notes) | OR 1.19 (0.86 – 1.65)  HR 1.61 (1.17 – 2.22) | aHR 1.39 (1.02 – 1.88) |  |
| Lloyd-Smith 2010^28^ |  | Unstable housing, current visit | Hospitalization for injecting-related infection (cellulitis, abscess, osteomyelitis, Staph infection, endocarditis, septic arthritis, ulcer, thrombophlebitis, myositis) | HR 1.65 (1.08 – 2.53) | aHR 1.26 (0.79 – 2.02) |  |
| Milloy 2010^30^ |  | Unstable housing, current visit | ED visit for cutaneous injecting-related infection (admin data) from SIF/community cohort | At baseline, ED visit in previous 6 month:  No: 12/168 Yes: 18/109 p=0.259  RR for number of ED visits for CIRI during follow-up:  1.23 (0.98–1.56) | aRR for number of ED visits for CIRI during follow-up: 1.30 (1.01–1.67) |  |
| Noroozi 2019^34^ |  | Housing | Lifetime injection site infection | Homeless: 150/300 Stable housing: 50/245 p=0.02 |  |  |
| Phillips 2017^36^ |  | Homelessness (past 90 d) | Past year SSTI, self-report | OR 0.78 (0.39, 1.56) | aOR 0.52 (0.22, 1.27) |  |
| Phillips 2008^37^ |  | Homeless (how many nights they had spent on the street or in a shelter in the 6-months prior to baseline) | ED visit or hospitalization for skin abscess, cellulitis, osteomyelitis, or endocarditis; self-report | OR 1.00; 0.43–2.32 | aOR 1.00; 0.43–2.33 |  |
| Phillips 2010^38^ |  | Homeless | Past year SSTI, self-report | OR 1.22; 0.35–4.27 |  |  |
| Pollini 2010^39^ |  | Homeless, past 6 months | Past 6-months abscess | Yes: 8/22  No: 119/482 p=0.07 |  |  |
| Roux 2020^42^ |  | Slept in the street at least once in prior month | At least one cutaneous abscess in the previous six months | OR  No: Ref  Yes: 2.22 [1.19,4.15] |  |  |
| Shah 2020^46^ |  | Stable housing | Endocarditis | Stable housing: 12/57  No stable housing: 21/78 p=0.17 |  |  |
| Sierra 2006^47^ |  | Homeless | Invasive soft-tissue Group A Strep (S. pyogenes) infections in Barcelona | 4.22 (1.5-12.5) |  |  |
| Smith 2015^49^ |  | Housing | Current abscess | Unstable housing: 6/39  Stable housing: 21/113 |  |  |
| Summers 2017^51^ |  | Stably housed | Past year abscess, self-report | OR 3.09 [1.53, 6.24] | aOR 1.28 [0.33, 4.94] |  |
| Thønnings 2020^52^ |  | Homeless | Bacteraemia, among hospitalised PWID | OR 0.74 (0.38–1.45) |  |  |
| Wilson 2002^56^ |  | Homeless | Infective endocarditis, self-report + medical chart review | OR 0.78 (0.46–1.32) |  |  |
| Wright 2020^57^ |  | Ever homeless | Lifetime SSTI, self-report | Yes: 231/355  No: 60/100 p=0.35 |  |  |
| Wurcel 2018^59^ |  | Homeless (“Do you consider yourself homeless?”) | Abscess ever | OR  Among females: 2.33 (0.77, 7.12)  Among males: 0.69 (0.30, 1.56) | aOR  Among females: 1.99 (0.48, 8.27)  Among males: 0.40 (0.16, 1.00) |  |
| Food insecurity | | | | | | |
| Saeland 2014^43^ |  | “Limited access to food” (NOS) | Current abscess. Self-report and confirmed by physical examination. | Limited access: 35/123  No limited access: 12/65  P=0.10 |  |  |
| Saeland 2014^43^ |  | Number of meals last 24 hours | Current abscess. Self-report and confirmed by physical examination. | Median (IQR) Abscess: 2 (1,3)  No abscess: 3 (2,4)  P=0.01 |  |  |
| Health insurance | | | | | | |
| Baltes 2020^1^ |  | Health insurance | Past-year SSTI, self report | Private: 0/5  Medicaid: 9/43  Medicare: 2/2  Other: 1/2  Calculated OR (exposure is Medicaid)  0.79 (0.18-3.56) |  |  |
| **Study** |  | **Exposures** | **Outcomes** | **Unadjusted effect estimate** | **Adjusted effect estimate** |  |
| Substance use | | | | | | |
| Overdose history | | | | | | |
| Hope 2014^17^ |  | Overdose history | Self-reported abscess “(a swelling containing pus)”, past 28 days | Never: 20/479 (4.2%)  Yes, not last year: 13/200 (6.5%) Yes, last year: 19.175 (11%) p=0.007 | aOR Never: 1.00 Yes, not last year: 1.39 (0.65 – 2.99)  Yes, last year: 2.41 (1.20 – 4.82) |  |
| Hope 2014^17^ |  | Overdose | Self-reported “redness, swelling, and tenderness”, past 28 days | Never: 83/479 (17%)  Yes, not last year: 43/200 (21%) Yes, last year: 52/175 (30%) p=0.003 | Never: 1.00  Yes, not last year: 1.20 (0.76 – 1.88) Yes, last year: 1.84 (1.19 – 2.87) |  |
| Hope 2015^18^ |  | Overdose | Abscess, past 12 months, self-reported | Never: 68/479  Yes, not preceding year: 47/200  Yes, preceding year: 45/175  P=0.001 | Never: Ref  Yes, not preceding year: 1.50 (0.97 – 2.31)  Yes, preceding year: 1.69 (1.09 – 2.63) |  |
| Hope 2015^18^ |  | Overdose | Cellulitis (redness, swelling, or tenderness), past 12 months, self-reported | Never: 198/479  Yes, not preceding year: 105/200  Yes, preceding year: 110/175  P<0.001 | Never: Ref  Yes, not preceding year: 1.37 (0.98 – 1.93)  Yes, preceding year: 2.00 (1.39 – 2.89) |  |
| Saeland 2014^43^ |  | Number of overdoses (NOS) | Current abscess. Self-report and confirmed by physical examination. | “did not differ” (data not shown) |  |  |
| **Study** |  | **Exposures** | **Outcomes** | **Unadjusted effect estimate** | **Adjusted/final effect estimate** |  |
| Heroin use | | | | | | |
| Baltes 2020^1^ |  | Drug of choice: heroin | Past-year SSTI, self report | Heroin: 6/20  Other: 12/59 |  |  |
| Dahlman 2015^11^ |  | Heroin as “main drug” vs. not” | Ever had an SSTI | OR 2.18 (0.88–5.43) |  |  |
| Dahlman 2017^12^ |  | Injected heroin, past 6 months | Past 30 day SSTI | OR 1.28 (0.48, 3.44) |  |  |
| Fink 2013^16^ |  | Type of drug injected: Heroin | Past 6 months SSTI, self-report | Heroin: 312/807 (39%),  Not heroin: 8/51  p=0.001 |  |  |
| Hope 2014^17^ |  | Injected heroin last 28 days | Self-reported abscess “(a swelling containing pus)”, past 28 days | No: 0/48 (0%)  Yes: 52/807 (6.4%) p=0.07 |  |  |
| Hope 2014^17^ |  | Injected heroin last 28 days | Self-reported “redness, swelling, and tenderness”, past 28 days | “No associations found” |  |  |
| Hope 2015^18^ |  | Injected heroin preceding year | Abscess, past 12 months, self-reported | “Not associated” |  |  |
| Hope 2015^18^ |  | Injected heroin preceding year | Cellulitis (redness, swelling, or tenderness), past 12 months, self-reported | “Not associated” |  |  |
| Hope 2010^19^ |  | Opiate use in past year | Self-report, ‘swelling containing pus (abscess), sore, or open wound’ at an injection site during the previous 12 months | Opiate, no stimulant: 688/2105 (33%) Stimulant, no opiate: 39/206 (19%) Stimulant and opiate: 1136/2898 (39%) p<0.001 | aOR Opiate, no stimulant: 1.00 Stimulant, no opiate: 0.49 (0.34-0.71) Stimulant and opiate: 1.24 (1.09-1.40) |  |
| Hope 2010^19^ |  | Opiate use in past year | Self-report, ‘swelling containing pus (abscess), sore, or open wound’ at an injection site during the previous 12 months  (among subgroup reporting injecting in past 4 weeks) | Opiate, no stimulant: 484/1416 (34%) Stimulant, no opiate: 30/144 (21%) Stimulant and opiate: 861/2173 (40%) p<0.001 | aOR Opiate, no stimulant: 1.00 Stimulant, no opiate: 0.47 (0.30-0.75) Stimulant and opiate: 1.06 (0.91-1.24) |  |
| Lloyd-Smith 2005^24^ |  | Heroin use, past 6 months | Self-report abscess (“lasting for more than 3 days”), past 6 months | OR  Less than daily: Ref  Daily use: 1.4 (1.1 – 1.8) |  |  |
| Lloyd-Smith 2012^25^ |  | Heroin injecting, past 6 months | ED visit for cutaneous injecting-related infection (admin data) from SIF/community cohort | OR  Less than daily: Ref Daily use: 1.44 (1.00 – 2.07)  Among females:  HR 1.55 (1.01 – 2.37)  Among males:  HR 1.13 (0.85 – 1.51) | aHR, only separate models by sex/gender  Among females: aHR 1.22 (0.79 – 1.90)  Among males: Not reported (because not included in stepwise regression) |  |
| Lloyd-Smith 2008^26^ |  | Heroin injection, past 6 months | Current injecting-related skin infection (self-report & confirmed by study nurse) | OR  Less than daily: Ref  Daily use: 1.53 (1.14 – 2.04) | aOR  Less than daily: Ref  Daily use: 1.26 (0.93 – 1.72) |  |
| Lloyd-Smith 2009^27^ |  | Heroin injection, past six months | Injecting-related infection cared for at supervised consumption site (from nursing notes) | OR  Less than daily: Ref  Daily use: 1.41 (1.02 – 1.95)  HR  Daily use: 1.82 (1.37 – 2.42) | aHR 1.52 (1.13 – 2.04) |  |
| Milloy 2010^30^ |  | >= Daily heroin use, past 6 months | ED visit for cutaneous injecting-related infection (admin data) from SIF/community cohort | At baseline, ED visit in previous 6 month:  No: 40/302  Yes: 32/243 p=0.979  RR for number of ED visits for CIRI during follow-up:  0.82 (0.69–0.98) |  |  |
| Murphy 2001^32^ |  | Ever injected heroin | ED visit or hospitalization for injecting-related abscess, recruited for this case-control study | Yes: 146/401 No: 5/18 p=0.06 |  |  |
| Phillips 2008^37^ |  | Heroin use days in past month | ED visit or hospitalization for skin abscess, cellulitis, osteomyelitis, or endocarditis; self-report | OR 1.00; 0.94–1.06 | aOR 1.00; 0.94–1.06 |  |
| Phillips 2010^38^ |  | Heroin injection days past month | Past year SSTI, self-report | OR 1.11; 1.04–1.18 |  |  |
| Pollini 2010^39^ |  | Injected heroin (alone) past 6 months | Past 6-months abscess | Yes: 105/502  No: 22/121  P=0.35 |  |  |
| Roux 2020^42^ |  | Heroin injection at least once in previous month | At least one cutaneous abscess in the previous six months | OR  No: Ref  Yes: 2.21 [1.14,4.30] |  |  |
| Saeland 2014^43^ |  | Heroin on a regular basis | Current abscess. Self-report and confirmed by physical examination. | Heroin: 44/155  No heroin: 3/36  P=0.012 |  |  |
| Shah 2020^46^ |  | Heroin injected | Endocarditis | Heroin: 4/18  No heroin: 29/117 p=1.00 |  |  |
| Sierra 2006^47^ |  | Heroin alone | Invasive soft-tissue Group A Strep (S. pyogenes) infections in Barcelona | Cases: 0 (n=15 total)  Controls: 20 (n=58 total)  P<0.001  Heroin alone: 0/20  Other: 15/53 |  |  |
| Smith 2015^49^ |  | Heroin use | Current abscess | Other: 2/31  Every day: 25/121 |  |  |
| Wurcel 2018^59^ |  | High heroin injection use in the past 30 days | Abscess ever | OR  Among females: 1.50 (0.64, 3.51)  Among males: 0.85 (0.49, 1.50) | aOR  Among females: 2.33 (0.70, 7.78)  Among males: 0.74 (0.37, 1.47) |  |
| Type of heroin / heroin formulation | | | | | | |
| Ciccarone 2016^9^ |  | Mexican “tar” heroin-dominant cities vs. Colombian powder heroin-dominant cities | Proportion of opiate-related hospital admissions comprised of skin and soft-tissue infections | 10.7% in MHD cities vs. 5.2% in CHD cities (p<0.001) | Unclear / uninterpretable  Figure 3 shows adjusted odds ratio of 2.05 (p<0.001) for “Percent Mexican-sourced dominant”, but units and increments are not provided.  Visually extracting from figure, 95%CI appears to be 1.75-2.40. |  |
| Pollini 2010^39^ |  | Form of heroin usually injected (Black tar) | Past 6-months abscess | Black tar: 127/618  Other: 0/5 p=0.25 |  |  |
| Summers 2017^51^ |  | Form of heroin, black tar vs. powder | Past year abscess, self-report | OR 7.93 [3.73, 16.88] | aOR 7.68 [3.01,19.60]  This is from “Final model” which was stepwise, after an intermediate model that included all covariates |  |
| Cocaine | | | | | | |
| Baltes 2020^1^ |  | Drug of choice: Cocaine | Past-year SSTI, self report | Cocaine: 2/5  Other: 16/74 |  |  |
| Buchanan 2006^7^ |  | Ever injected “crack” cocaine (vs. never injected crack cocaine) | Ever had abscess | OR 1.66 (1.05 – 2.63) | aOR 0.91 (0.53 – 1.57) |  |
| Dahlman 2017^12^ |  | Injected crack or rock cocaine, past 6 months | Past 30 day SSTI | OR 0.95 (0.37, 2.46) |  |  |
| Dahlman 2017^12^ |  | Injected powder cocaine, past 6 months | Past 30 day SSTI | OR 1.09 (0.38, 3.15) |  |  |
| Fink 2013^16^ |  | Type of drug injected: Cocaine (powder) | Past 6 months SSTI, self-report | Cocaine (powder): 40/108 (37%), Not cocaine (powder): 280/750  p=0.94 |  |  |
| Fink 2013^16^ |  | Type of drug injected: Cocaine (rock) | Past 6 months SSTI, self-report | Cocaine (rock): 20/53 (38%)  Not cocaine (rock): 280/785 p=0.95 |  |  |
| Hope 2014^17^ |  | Injected cocaine last 28 days | Self-reported abscess “(a swelling containing pus)”, past 28 days | No: 44/796 (5.5%)  Yes: 8/59 (14%) p=0.013 |  |  |
| Hope 2014^17^ |  | Injected crack last 28 days | Self-reported abscess “(a swelling containing pus)”, past 28 days | No: 18/425 (4.2%)  Yes: 33/430 (7.7%) p=0.034 |  |  |
| Hope 2014^17^ |  | Injected cocaine last 28 days | Self-reported “redness, swelling, and tenderness”, past 28 days | “No associations found” |  |  |
| Hope 2014^17^ |  | Injected crack last 28 days | Self-reported “redness, swelling, and tenderness”, past 28 days | No: 78/425 (18%) Yes: 99/430 (23%) p=0.092 |  |  |
| Hope 2015^18^ |  | Injected cocaine preceding year | Abscess, past 12 months, self-reported | No: 123/732  Yes: 37/123 | 1.78 (1.14 – 2.78) |  |
| Hope 2015^18^ |  | Injected cocaine preceding year | Cellulitis (redness, swelling, or tenderness), past 12 months, self-reported | No: 334/732  Yes: 70/123 |  |  |
| Hope 2015^18^ |  | Injected crack preceding year | Abscess, past 12 months, self-reported | No: 43/331  Yes: 117/524 |  |  |
| Hope 2015^18^ |  | Injected crack preceding year | Cellulitis (redness, swelling, or tenderness), past 12 months, self-reported | “not associated” |  |  |
| Hope 2008^20^ |  | Inject crack last 4 weeks | Self-reported symptoms of injection site infections (abscess or open wound), past year | ORs No: 1.0 Yes: 1.7 (1.3 – 2.2) | aORs  No: 1.0 Yes: 1.5 (1.1 – 2.0) |  |
| Lloyd-Smith 2005^24^ |  | Cocaine use, past 6 months | Self-report abscess (“lasting for more than 3 days”), past 6 months | OR  Less than daily: Ref  Daily use: 1.9 (1.5 – 2.5) | aOR  Less than daily: Ref  Daily use: 1.5 (1.2 – 2.0) |  |
| Lloyd-Smith 2012^25^ |  | Cocaine injecting, past 6 months | ED visit for cutaneous injecting-related infection (admin data) from SIF/community cohort | HR, only separate models by sex/gender  Among females:  Less than daily: Ref  Daily: 1.37 (0.88 – 2.11)  Among males:  Less than daily: Ref  Daily: 1.18 (0.87 – 1.60) |  |  |
| Lloyd-Smith 2012^25^ |  | Crack use, past 6 months | ED visit for cutaneous injecting-related infection (admin data) from SIF/community cohort | HR, only separate models by sex/gender  Among females:  Less than daily: Ref  Daily: 1.42 (0.90 – 2.24)  Among males:  Less than daily: Ref  Daily: 1.46 (1.10 – 1.94) | aHR, only separate models by sex/gender  Among females:  Not reported  Among males: 1.30 (0.97 – 1.74) |  |
| Lloyd-Smith 2008^26^ |  | Cocaine injection, past 6 months | Current injecting-related skin infection (self-report & confirmed by study nurse) | OR  Less than daily: Ref  Daily use: 1.66 (1.23 – 2.25) | aOR  Less than daily: Ref  Daily use: 1.41 (1.02 – 1.95) |  |
| Lloyd-Smith 2008^26^ |  | Crack injection, past 6 months | Current injecting-related skin infection (self-report & confirmed by study nurse) | OR  Less than daily: Ref  Daily use: 1.54 (0.96 – 2.46) |  |  |
| Lloyd-Smith 2009^27^ |  | Cocaine injection, past six months | Injecting-related infection cared for at supervised consumption site (from nursing notes) | OR  Less than daily: Ref  Daily use: 1.57 (1.13 – 2.19)  HR  Daily use: 1.14 (0.82 - 1.58) |  |  |
| Lloyd-Smith 2010^28^ |  | Cocaine injection, past six months | Hospitalization for injecting-related infection (cellulitis, abscess, osteomyelitis, Staph infection, endocarditis, septic arthritis, ulcer, thrombophlebitis, myositis) | HR  Daily use: 1.75 (1.17 – 2.62) | aHR  Daily use: 1.46 (0.94 – 2.25) |  |
| Milloy 2010^30^ |  | >= Daily cocaine use, past 6 months | ED visit for cutaneous injecting-related infection (admin data) from SIF/community cohort | At baseline, ED visit in previous 6 month:  No: 49/381  Yes: 23/164 p=0.713  RR for number of ED visits for CIRI during follow-up:  1.05 (0.85–1.29) |  |  |
| Murphy 2001^32^ |  | Ever injected cocaine | ED visit or hospitalization for injecting-related abscess, recruited for this case-control study | Yes: 113/305 No: 18/82 p=0.31 |  |  |
| Phillips 2017^36^ |  | Cocaine use, past 90 days | Past year SSTI, self-report | OR 1.77 (0.83, 3.78) | aOR 1.39 (0.50, 3.91) |  |
| Phillips 2008^37^ |  | Cocaine use days in past month | ED visit or hospitalization for skin abscess, cellulitis, osteomyelitis, or endocarditis; self-report | OR 0.97; 0.90–1.04 | aOR 0.97; 0.90–1.04 |  |
| Roux 2020^42^ |  | Stimulant injection at least once in previous month | At least one cutaneous abscess in the previous six months | OR  No: Ref  Yes: 1.60 [0.88,2.91] |  |  |
| Shah 2020^46^ |  | Cocaine injected | Endocarditis | Cocaine: 3/19  No cocaine: 30/116 p=0.41 |  |  |
| Shah 2020^46^ |  | Crack injected | Endocarditis | Crack: 0/8  No cocaine: 33/127 p=0.20 |  |  |
| Sierra 2006^47^ |  | Cocaine alone | Invasive soft-tissue Group A Strep (S. pyogenes) infections in Barcelona | OR 0 (0-1.26)  P=0.94 |  |  |
| Smith 2015^49^ |  | Cocaine use | Current abscess | Other: 14/98  Every day: 13/54 |  |  |
| Wurcel 2018^59^ |  | High cocaine injection use in the past 30 days | Abscess ever | OR  Among females: 0.47 (0.17, 1.29)  Among males: 1.77 (0.89, 3.51) | aOR  Among females: 0.27 (0.06, 1.23)  Among males: 2.50 (1.06, 5.91) |  |
| Illicit amphetamines / Methamphetamines | | | | | | |
| Baltes 2020^1^ |  | Drug of choice: Methamphetamine | Past-year SSTI, self report | Methamphetamine: 7/40  Other: 11/39 |  |  |
| Dahlman 2017^12^ |  | Injected methamphetamine, past 6 months | Past 30 day SSTI | OR 0.70 (0.27, 1.76) |  |  |
| Doran 2020^14^ |  | Main drug injected in past year (opioids, cocaine, crack, and combinations vs. amphetamine-like drugs as ref) | Past year SSTI | OR 2.0 (1.3 – 3.0) | aOR 1.7 (1.1 – 2.8) |  |
| Doran 2020^14^ |  | Main drug injected in past year (opioids, cocaine, crack, and combinations vs. amphetamine-like drugs as ref) | Ever SSTI | OR 2.5 (0.9 – 6.8) |  |  |
| Fink 2013^16^ |  | Type of drug injected: Methamphetamines | Past 6 months SSTI, self-report | Methamphetamines: 14/69 (20%),  Not methamphetamines: 306/789 (39%)  p=0.002 |  |  |
| Hope 2014^17^ |  | Injected amphetamine last 28 days | Self-reported abscess “(a swelling containing pus)”, past 28 days | “No associations found” |  |  |
| Hope 2014^17^ |  | Injected amphetamine last 28 days | Self-reported “redness, swelling, and tenderness”, past 28 days | “No associations found” |  |  |
| Hope 2015^18^ |  | Injected amphetamine preceding year | Abscess, past 12 months, self-reported | “Not associated” |  |  |
| Hope 2015^18^ |  | Injected amphetamine preceding year | Cellulitis (redness, swelling, or tenderness), past 12 months, self-reported | “Not associated” |  |  |
| Hope 2008^20^ |  | Injecting amphetamines | Self-reported symptoms of injection site infections (abscess or open wound), past year | “…was not associated with…” |  |  |
| Lloyd-Smith 2008^26^ |  | Crystal meth injection, past 6 months | Current injecting-related skin infection (self-report & confirmed by study nurse) | OR  Less than daily: Ref  Daily use: 1.48 (0.73 – 3.02) |  |  |
| Lloyd-Smith 2008^26^  #530 |  | Crystal meth injection, past 6 months | Current injecting-related skin infection (self-report & confirmed by study nurse) | OR  Less than daily: Ref  Daily use: 1.48 (0.73 – 3.02) |  |  |
| McMahan 2020^29^ |  | Main drug is methamphetamine (vs. main drug is opioids) | Past-year injecting-related infection (“an abscess, skin infection such as cellulitis, blood clot or blood infection like sepsis, or endocarditis”) | “A smaller proportion of participants whose main drug was methamphetamine… had an infection that was likely related to injection in the past 12 months (26 % vs 48 %, p < .001) compared to participants whose main drug was an opioid.”  Methamphetamine: 36/140 Opioids: 214/443 |  |  |
| Murphy 2001^32^ |  | Ever injected amphetamine | ED visit or hospitalization for injecting-related abscess, recruited for this case-control study | Yes: 81/243  No: 70/111 p=0.27 |  |  |
| Noroozi 2019^34^ |  | Methamphetamine injection during last 6 mo. | Lifetime injection site infection | ORs  Yes: 1.7 (1.1-4.52)  No: 1 | aORs  1.6 (0.48-5.7) |  |
| Pollini 2010^39^ |  | Injected methamphetamine (alone) past 6 months | Past 6-months abscess | Yes: 19/88  No: 108/535  P=0.72 |  |  |
| Saeland 2014^43^ |  | Amphetamine on a regular basis | Current abscess. Self-report and confirmed by physical examination. | Amphetamine: 21/88  No amphetamine: 26/104  P=0.890 |  |  |
| Shah 2020^46^ |  | Crystal methamphetamine | Endocarditis | Crystal meth: 18/98  No: 15/37 p=0.07 |  |  |
| Pollini 2010^39^ |  | Color of meth usually injected | Past 6-months abscess | Clear (crystal): 100/462  Other: 27/161 p=0.26 |  |  |
| Other/Combined stimulant use | | | | | | |
| Hope 2010^19^ |  | Stimulant use in past year | Self-report, ‘swelling containing pus (abscess), sore, or open wound’ at an injection site during the previous 12 months | Opiate, no stimulant: 688/2105 (33%) Stimulant, no opiate: 39/206 (19%) Stimulant and opiate: 1136/2898 (39%) p<0.001 | aOR Opiate, no stimulant: 1.00 Stimulant, no opiate: 0.49 (0.34-0.71) Stimulant and opiate: 1.24 (1.09-1.40) |  |
| Hope 2010^19^ |  | Stimulant use in past year | Self-report, ‘swelling containing pus (abscess), sore, or open wound’ at an injection site during the previous 12 months  (among subgroup reporting injecting in past 4 weeks) | Opiate, no stimulant: 484/1416 (34%) Stimulant, no opiate: 30/144 (21%) Stimulant and opiate: 861/2173 (40%) p<0.001 | aOR Opiate, no stimulant: 1.00 Stimulant, no opiate: 0.47 (0.30-0.75) Stimulant and opiate: 1.06 (0.91-1.24) |  |
| Yeung 2017^60^ |  | Self-reported use of ethylphenidate (a novel psychoactive substance / stimulant associated with high frequency of injecting) | Weekly rate of S. pyogenes or S. aureus infections among people who inject drugs |  | aRR 1.81 (1.12-2.93) |  |
| Speedball | | | | | | |
| Dahlman 2017^12^ |  | Injected speedball, past 6 months | Past 30 day SSTI | OR 0.78 (0.30, 2.01) |  |  |
| Dahlman 2017^12^ |  | Injected goofball, past 6 months | Past 30 day SSTI | OR 1.17 (0.45, 3.04) |  |  |
| Fink 2013^16^ |  | Type of drug injected: Speedball | Past 6 months SSTI, self-report | Speedball (heroin+cocaine): 88/211 (42% ),  Not speedball: 232/647 p=0.13 |  |  |
| Lloyd-Smith 2012^25^ |  | Speedball injecting, past 6 months | ED visit for cutaneous injecting-related infection (admin data) from SIF/community cohort | OR  Less than daily: Ref  Daily use: 1.77 (1.12 – 2.79)  Among females:  HR 1.61 (0.97 - 2.65)  Among males:  HR 1.28 (0.82 – 2.02) |  |  |
| Lloyd-Smith 2009^27^ |  | Speedball injection, past six months | Injecting-related infection cared for at supervised consumption site (from nursing notes) | OR  Less than daily: Ref  Daily use: 1.96 (1.30 – 2.95)  HR  Daily use: 1.92 (1.21 – 3.05) | aHR 1.47 (0.95 – 2.26) |  |
| Lloyd-Smith 2010^28^ |  | Speedball injection, past six months | Hospitalization for injecting-related infection (cellulitis, abscess, osteomyelitis, Staph infection, endocarditis, septic arthritis, ulcer, thrombophlebitis, myositis) | HR 1.90 (1.15 – 3.14) | aHR 1.19 (0.69 – 2.07) |  |
| Milloy 2010^30^ |  | >= Daily speedball use, past 6 months | ED visit for cutaneous injecting-related infection (admin data) from SIF/community cohort | At baseline, ED visit in previous 6 month:  No speedball use: 63/483  Yes speedball use: 9/62 p=0.747  RR for number of ED visits for CIRI during follow-up:  1.24 (0.92–1.67) |  |  |
| Murphy 2001^32^ |  | Ever injected speedball | ED visit or hospitalization for injecting-related abscess, recruited for this case-control study | Yes: 133/331  No: 18/75 p<0.001 |  |  |
| Phillips 2010^38^ |  | Speedball injection days past month | Past year SSTI, self-report | OR 1.06; 1.00–1.12 |  |  |
| Pollini 2010^39^ |  | Injected heroin and methamphetamine together, past 6 months | Past 6-months abscess | Yes: 70/318  No: 57/305  P=0.38 |  |  |
| Smith 2015^49^ |  | Speedball | Current abscess | Other: 16/103  Every day: 11/49 |  |  |
| Other or unspecified polydrug use | | | | | | |
| Binswanger 2000^6^ |  | “Type of drug injected” (not defined) | Current abscess or cellulitis on physical examination | “did not differ significantly” |  |  |
| Dunleavy 2017^15^ |  | Poly-drug injection (Defined as use of more than one drug type in past 6 months (where drug type is Opiate, Stimulant or Other including Legal Highs). | Past year SSTI | Yes: 133/300 (44%) No: 399/1567 (26%), p<0.001 |  |  |
| Hope 2010^19^ |  | Stimulant and opiate use, past year | Self-report, ‘swelling containing pus (abscess), sore, or open wound’ at an injection site during the previous 12 months | Opiate, no stimulant: 688/2105 (33%) Stimulant, no opiate: 39/206 (19%) Stimulant and opiate: 1136/2898 (39%) p<0.001 | aOR Opiate, no stimulant: 1.00 Stimulant, no opiate: 0.49 (0.34-0.71) Stimulant and opiate: 1.24 (1.09-1.40) |  |
| Hope 2010^19^ |  | Stimulant and opiate use, past year | Self-report, ‘swelling containing pus (abscess), sore, or open wound’ at an injection site during the previous 12 months  (among subgroup reporting injecting in past 4 weeks) | Opiate, no stimulant: 484/1416 (34%) Stimulant, no opiate: 30/144 (21%) Stimulant and opiate: 861/2173 (40%) p<0.001 | aOR Opiate, no stimulant: 1.00 Stimulant, no opiate: 0.47 (0.30-0.75) Stimulant and opiate: 1.06 (0.91-1.24) |  |
| Noroozi 2019^34^ |  | “Poly-drug use” | Lifetime injection site infection | Yes: 110/236 No: 90/264  P=0.01 |  |  |
| Roux 2020^42^ |  | Polydrug use in past month (excluding cannabis) | At least one cutaneous abscess in the previous six months | OR  No: Ref  Yes: 1.53 [1.23,1.89] | aOR 1.41 [1.10,1.81] |  |
| Prescription opioids - Injected methadone | | | | | | |
| Baltes 2020^1^ |  | Drug of choice: Methadone | Past-year SSTI, self report | Methadone: 1/2  Other: 17/77  Calculated OR 3.52 (0.21-59.43) |  |  |
| Dahlman 2015^11^ |  | Injected methadone liquid, ever | Ever had an SSTI | OR 1.41 (0.57–3.52) |  |  |
| Dahlman 2015^11^ |  | Injected methadone tablets, ever | Ever had an SSTI | OR 1.30 (0.42–4.00) |  |  |
| Roux 2020^42^ |  | Methadone injection at least once in previous month | At least one cutaneous abscess in the previous six months | OR  No: Ref  Yes: 0.32 [0.15,0.68] |  |  |
| Prescription opioids - Injected buprenorphine | | | | | | |
| Dahlman 2015^11^ |  | Injected buprenorphine, ever | Ever had an SSTI | OR 0.95 (0.38–2.36) |  |  |
| Dahlman 2015^11^ |  | Injected buprenorphine-naloxone, ever | Ever had an SSTI | OR 1.21 (0.42–3.54) |  |  |
| Prescription opioids (not OAT) | | | | | | |
| Baltes 2020^1^ |  | Drug of choice: Opiate pain killers | Past-year SSTI, self report | “Opiate pain killers”: 0/2  Other: 18/77 |  |  |
| Shah 2020^46^ |  | Oxycodone hydrochloride tablets (Oxycontin) | Endocarditis | Oxy: 10/53 No Oxy: 59/82 p=0.22  Calculated OR  0.83 (0.36-1.90) |  |  |
| Shah 2020^46^ |  | Hydromorphone controlled-release capsules (HydromorphContin) | Endocarditis | Hydromorph: 30/113 No Hydromorph: 3/22 p=0.20  Calculated OR  2.29 (0.63-8.29) |  |  |
| Shah 2020^46^ |  | Hydromorphone tablets (Dilaudid) | Endocarditis | Dilaudid: 21/97  No dilaudid: 12/38 p=0.43  Calculated OR 0.59 (0.26-1.38) |  |  |
| Shah 2020^46^ |  | Fentanyl patch | Endocarditis | Fentanyl patch: 3/4  No patch: 30/131  p=0.45  Calculated:  OR 10.1 (1.01 - 100.70) |  |  |
| Shah 2020^46^ |  | Fentanyl tablet | Endocarditis | Fentanyl tablet: 1/5  No fentanyl: 32 /130  p=1.00  Calculated:  OR 0.76 (0.08-7.10) | Not tested |  |
| Silverman 2020^48^ |  | Living in regions with high vs. low hydromorphone prescription rates | Endocarditis | “Within the matched cohort, we observed 254 (1·6%) of 16288 admissions with infective endocarditis related to injection drug use in sectors with high hydromorphone prescription rates and 113 (0·7%) of 16288 admissions in sectors with low prescription rates (adjusted OR 2·2, 95% CI 1·8–2·8, p<0·0001). |  |  |
| Silverman 2020^48^ |  | People who filled a prescription for hydromorphone vs. other opioids | Endocarditis | “Among the matched cohort, we observed 109 (2·8%) admissions with infective endocarditis among patients who filled prescriptions for hydromorphone compared with 41 (1·1%) admissions among those who filled prescriptions for non-hydromorphone opioids (adjusted OR 2·5, 95% CI 1·8–3·7, p<0·0001).” |  |  |
| Silverman 2020^48^ |  | People who filled a prescription for hydromorphone vs. other opioids | Endocarditis | “Among the matched cohort, we observed 109 (2·8%) admissions with infective endocarditis among patients who filled prescriptions for hydromorphone compared with 41 (1·1%) admissions among those who filled prescriptions for non-hydromorphone opioids (adjusted OR 2·5, 95% CI 1·8–3·7, p<0·0001).”  “We observed 36 (1·8%) admissions with infective endocarditis among 1989 patients who filled prescriptions for immediate-release hydromorphone and 21 (1·1%) admissions among 1989 matched patients who filled prescriptions of non-hydromorphone opioids (adjusted OR 1·7, 95% CI 0·9–3·6, p=0·072).  For controlled-release hydromorphone, we observed 73 (3·9%) admissions compared with 20 (1·1%) admissions among 1895 matched patients who filled prescriptions for nonhydromorphone opioids (adjusted OR 3·3, 95% CI 2·1–5·6, p <0·0001). |  |  |
| Weir 2019^55^ |  | Trend in proportion of prescription opioids that are hydromorphone | Trend in proportion of endocarditis admissions attributable to injection drug use | Visually compared.  Hydromorphone was 16% of outpatient opioid prescriptions at the start of the study period and 53% by the end. |  |  |
| Prescription stimulants | | | | | | |
| Dahlman 2015^11^ |  | Injected methylphenidate, ever | Ever had an SSTI | OR 0.80 (0.31–2.06) |  |  |
| Shah 2020^46^ |  | Bupropion (Wellbutrin) | Endocarditis | Bupropion: 1/1  No: 32/134 p=0.24  Calculated OR is Infinity. |  |  |
| Shah 2020^46^ |  | Methylphenidate (Ritalin) | Endocarditis | Methylphenidate: 4/22  No: 29/113 p=0.43  Calculated OR  0.64 (0.20-2.06) |  |  |
| Other/multiple prescription drugs | | | | | | |
| Baltes 2020^1^ |  | Drug of choice: Prescription anxiety drugs | Past-year SSTI, self report | Prescription anxiety drugs: 1/5  Other: 17/74 |  |  |
| Baltes 2020^1^ |  | Drug of choice: “Synthetics” | Past-year SSTI, self report | Synthetics: 0/2  Other: 18/79 |  |  |
| Baltes 2020^1^ |  | Drug of choice: Other | Past-year SSTI, self report | Other: 1/3  Other-other: 17/76 |  |  |
| Dahlman 2015^11^ |  | Injected prescribed drugs (crushed tablets/liquid), ever | Ever had an SSTI | OR 7.50 (2.52–22.32) | aOR 52.15 (5.17–525.67) |  |
| Dahlman 2015^11^ |  | Injected benzodiazepines, ever | Ever had an SSTI | OR 2.44 (0.98–6.04) |  |  |
| Dahlman 2017^12^ |  | Injected “nonpowder drugs”, past 6 months  “Nonpowder drugs” meant any of: “prescription pain relievers, prescription tranquilizers or sedatives, prescription stimulants, methadone, buprenorphine, or Suboxone” | Past 30 day SSTI | OR 3.57 (1.23, 10.35) | aOR 2.18 (0.66–7.18) |  |
| Saeland 2014^43^ |  | Flunitrazepam on a regular basis | Current abscess. Self-report and confirmed by physical examination. | Flunitrazepam: 39/130  No flunitrazepam: 8/61  P=0.009 |  |  |
| Saeland 2014^43^ |  | Benzodiazepines (?other than flunitrazepam) | Current abscess. Self-report and confirmed by physical examination. | Benzos: 14/58  No benzos: 33/135  P=0.965 |  |  |
| Unhealthy alcohol use | | | | | | |
| Dunleavy 2017^15^ |  | Excessive alcohol use (>14units/week for women and >21units per week for men) | Past year SSTI | Yes: 145/475 (30%) No: 385/1388 (28%), p=0.245 |  |  |
| Murphy 2001^32^ |  | Had 1+ alcoholic drink in the past month | ED visit or hospitalization for injecting-related abscess, recruited for this case-control study | No: 64/161  Yes: 87/263 p=0.17 |  |  |
| Phillips 2017^36^ |  | AUDIT-C positive  “The 3-item Alcohol Use Disorders Identification Test Consumption (AUDIT-C) (Bradley et al., 2007) was used to assess hazardous alcohol use. Scores range from 0 to 12 and a total score above 3 for females” | Past year SSTI, self-report | OR 0.72 (0.36, 1.47) | aOR 0.47 (0.19, 1.17) |  |
| Phillips 2008^37^ |  | AUDIT (>=8) | ED visit or hospitalization for skin abscess, cellulitis, osteomyelitis, or endocarditis; self-report | OR 0.83; 0.23–3.08 | aOR 0.78; 0.20–3.08 |  |
| Phillips 2008^37^ |  | Alcohol intoxication days in past month | ED visit or hospitalization for skin abscess, cellulitis, osteomyelitis, or endocarditis; self-report | OR 1.01; 0.94–1.08 | aOR 1.01; 0.94–1.08 |  |
| Roux 2020^42^ |  | Harmful alcohol consumption (“AUDIT C score ≥3 for women or ≥4 for women”) | At least one cutaneous abscess in the previous six months | OR  No: Ref  Yes: 0.89 [0.45,1.77] |  |  |
| Safaeian 2000^44^  #936 |  | Alcohol use | Infective endocarditis, self-report confirmed through medical chart review | OR No: Ref  Yes: 0.6 (0.4 – 1.2) |  |  |
| Wilson 2002^56^ |  | Alcohol consumption | Infective endocarditis, self-report + medical chart review | None: Ref  1-21 drinks/week: 0.61 (0.35–1.06)  >21 drinks/week: 0.53 (0.26–1.09) | None: Ref  1-21 drinks/week: 0.43 (0.22–0.83)  >21 drinks/week: 0.32 (0.13–0.78) |  |
| Smoking | | | | | | |
| Murphy 2001^32^ |  | Current cigarette smoker | ED visit or hospitalization for injecting-related abscess, recruited for this case-control study | Yes: 142/388  No: 9/36 p=0.16 |  |  |
| Pollini 2010^39^ |  | Smoked methamphetamine, past 6 months | Past 6-months abscess | Yes: 39/134  No: 88/489 p<0.01 | aOR 1.65 (1.05–2.62) |  |
| Saeland 2014^43^ |  | Hashish/cannabis | Current abscess. Self-report and confirmed by physical examination. | Cannabis: 24/92  No cannabis: 23/99  P=0.647 |  |  |
| Safaeian 2000^44^ |  | Cigarette smoking | Infective endocarditis, self-report confirmed through medical chart review | OR No: Ref  Yes: 0.6 (0.3 – 1.2) |  |  |
| Safaeian 2000^44^ |  | Cigarette smoking | Abscess, self-report | OR No: Ref  Yes: 2.4 (1.5-3.9) | aOR 1.8 (1.1-3.2) |  |
| **Study** |  | **Exposures** | **Outcomes** | **Unadjusted effect estimate** | **Adjusted/final effect estimate** |  |
| Drug policy and injecting contexts | | | | | | |
| Drug policy change - Policy changes on unregulated drug supply | | | | | | |
| DiGiorgio 2019^13^ |  | Implementation of state-wide restrictions on opioid prescribing in Louisiana, USA | Monthly rate of injection drug use-associated spinal epidural abscess | 0.54 cases per month to 1.15 cases per month (p = 0.017) |  |  |
| Weir 2019^55^ |  | Implementation of province-wide delisting of extended-release oxycodone | Quarterly proportion of endocarditis hospital admissions attributed to injection drug use | Non-significant step (p = 0.4) and slope (p = 0.8) change |  |  |
| Yeung 2017^60^ |  | Implementation of temporary class order on ethylphenidate (a novel psychoactive substance / stimulant associated with high frequency of injecting) | Weekly rate of S. pyogenes or S. aureus infections among people who inject drugs |  | level change: aRR 1.11 (95% CI 0.46 – 2.70)  trend change: aRR 0.88 (95% CI 0.82–0.94) |  |
| Nagar 2015^33^ |  | Implementation of House Bill 1 (July 2012) in Kentucky, USA, which restricted prescription opioids prescribing and dispensing | Annual rate of hospital admissions for spinal epidural abscess at one teaching hospital in Lexington, Kentucky | “The incidence of intraspinal abscess in subjects with drug abuse diagnosis remained constant between 2010 (n = 3) and 2012 (n = 3). However, it increased twofold (n = 7) in 2013 and then ninefold (n = 27) in 2014. |  |  |
| Supply network - Person / place of purchase | | | | | | |
| Sierra 2006^47^ |  | Purchasing from one particular drug seller who was identified as high risk (likely colonized) during outbreak | Invasive soft-tissue Group A Strep (S. pyogenes) infections in Barcelona | OR 72 (8 – 3090) |  |  |
| Sierra 2006^47^ |  | Purchasing from one particular drug selling site, where suspected drug seller worked | Invasive soft-tissue Group A Strep (S. pyogenes) infections in Barcelona | OR 33.92 (7.44 – 174.93) |  |  |
| **Study** |  | **Exposures** | **Outcomes** | **Unadjusted effect estimate** | **Adjusted effect estimate** |  |
| Social context of injecting | | | | | | |
| Pollini 2010^39^ |  | Injected drugs alone, past 6 mos. | Past 6-months abscess | Yes: 71/349  No: 56/274  p=0.99 |  |  |
| Pollini 2010^39^ |  | Injected drugs with friends, past 6 mos. | Past 6-months abscess | Yes: 58/282  No: 69/341  p=0.79 |  |  |
| Pollini 2010^39^ |  | Injected drugs with family member/spouse, past 6 mos. | Past 6-months abscess | Yes: 20/55  No: 107/568  P<0.01 |  |  |
| Roux 2020^42^ |  | Context of injecting | At least one cutaneous abscess in the previous six months | OR  Alone or did not inject: Ref  With someone else: 0.90 [0.48,1.68]  In group: 2.28 [0.90,5.80] 0.083 |  |  |
| Smith 2015^49^ |  | With whom do you inject | Current abscess | OR  Alone: Reference  Friends: 1.37 (0.45–4.18)  Family member or partner: 3.78 (1.38–10.31) | OR  Alone: Reference  Friends: 1.66 (0.42–6.47)  Family member or partner: 4.06 (0.99–16.58) |  |
| Shooting gallery | | | | | | |
| Phillips 2008^37^ |  | Shooting gallery use (item on the Risk Assessment Battery [RAB]) | ED visit or hospitalization for skin abscess, cellulitis, osteomyelitis, or endocarditis; self-report | OR 1.32; 0.31–5.58 | aOR 1.33; 0.31–5.73 |  |
| Pollini 2010^39^ |  | Locations injected drugs: Shooting gallery | Past 6-months abscess | Yes: 53/286  No: 74/337  P=0.33 |  |  |
| Public injecting | | | | | | |
| Dahlman 2017^12^ |  | Never injecting publicly vs. other response | Past 30 day SSTI | OR  Never: Ref Other: 1.15 (0.44 – 2.96) |  |  |
| Milloy 2010^30^ |  | Public injection, past 6 months | ED visit for cutaneous injecting-related infection (admin data) from SIF/community cohort | At baseline, ED visit in previous 6 month:  No: 62/506  Yes: 10/39 p=0.0.017  RR for number of ED visits for CIRI during follow-up:  1.40 (1.01–1.95) | aRR for number of ED visits for CIRI during follow-up: 1.35 (0.97–1.88) |  |
| Roux 2020^42^ |  | Injected in public settings | At least one cutaneous abscess in the previous six months | OR  No: Ref Yes: 1.67 [0.94,2.97] |  |  |
| Trayner 2020^54^ |  | Public injecting | Past year SSTI, self-report | OR 1.67 (1.24 to 2.23) | aOR 1.42 (1.17 to 1.73) |  |
| Assisted injecting / require help to inject | | | | | | |
| Dahlman 2017^12^ |  | Injected by another person, past 30 days | Past 30 day SSTI | OR 2.63 (1.02, 6.78) | aOR 2.08 (0.72–5.65) |  |
| Lee 2013^22^ |  | Requiring help injecting, past 6 moths | Past 6 month soft-tissue infections | OR 3.51 (1.43 – 8.64) | aOR 3.02 (1.14 – 7.72) |  |
| Lloyd-Smith 2012^25^ |  | Require assistance with injecting, past 6 months | ED visit for cutaneous injecting-related infection (admin data) from SIF/community cohort | OR  No: Ref  Yes: 2.01 (1.40 – 2.89)  Among females:  HR 1.56 (1.03 – 2.37)  Among males:  HR 1.59 (1.16 – 2.17) | aOR, only separate models by sex/gender  Among females:  aHR 1.40 (0.92 – 2.13)  Among males: 1.38 (1.01 – 1.90) |  |
| Lloyd-Smith 2008^26^ |  | Requiring help injecting, past 6 months | Current injecting-related skin infection (self-report & confirmed by study nurse) | OR 1.85 (1.37 – 2.50) | aOR 1.42 (1.03 – 1.94) |  |
| Lloyd-Smith 2009^27^ |  | Require help to inject | Injecting-related infection cared for at supervised consumption site (from nursing notes) | OR 1.27 (0.91 – 1.77) |  |  |
| Pollini 2010^39^ |  | Sought someone to help you inject, past 6 mos. | Past 6-months abscess | Yes: 25/70  No: 102/553  P<0.01 | aOR 2.06 (1.18 – 3.61) |  |
| Robertson 2010^41^ |  | Sought someone to help you inject, past 6 mos. | Past 6-months abscess | Yes: 123/260  No: 188/796 OR 2.90 2.17–3.89 | aOR 2.59 (1.93–3.47) |  |
| Roux 2020^42^ |  | Injection by someone else | At least one cutaneous abscess in the previous six months | OR  No: Ref Yes: 1.96 [1.08,3.57] | aOR 1.94 [0.96,3.92] |  |
| **Study** |  | **Exposures** | **Outcomes** | **Unadjusted effect estimate** | **Adjusted effect estimate** | **Notes** |
| Police contacts and arrests | | | | | | |
| Cooper 2005^10^ |  | Policing crackdowns on drug use and possession | Monthly rate of hospitalisation for abscess/cellulitis | IRR  Preparation: 0.96 (0.90 – 1.02)  Initiation month: aRR 0.99 (0.89 – 1.09)  First crackdown quarter: 1.06 (0.98 – 1.15)  Second crackdown quarter: 1.09 (0.98–1.22)  Third crackdown quarter: 0.94 (0.87–1.02)  Fourth crackdown quarter: 0.79 (0.71–1.15) | aIRR  Preparation: aRR 0.94 (0.85 – 1.04)  Initiation month: aRR 0.89 (0.78-1.00)  First crackdown quarter: 0.88 (0.77–1.00)  Second crackdown quarter: 0.86 (0.74–0.99)  Third crackdown quarter: 0.80 (0.67–0.94)  Fourth crackdown quarter: 0.72 (0.57–0.91) |  |
| Cooper 2005^10^ |  | Policing crackdowns on drug use and possession | Monthly rate of hospitalisation for endocarditis | Preparation: aRR 1.11 (0.88–1.40)  Initiation month: aRR 0.92 (0.68–1.26)  First crackdown quarter: 0.92 (0.73–1.16)  Second crackdown quarter: 0.89 (0.73–1.09)  Third crackdown quarter: 0.87 (0.66–1.15)  Fourth crackdown quarter: 0.62 (0.44–0.89) | Preparation: aRR 1.21 (0.96 – 1.54)  Initiation month: aRR 0.96 (0.66–1.41)  First crackdown quarter: 0.91 (0.66–1.25)  Second crackdown quarter: 0.81 (0.62–1.06)  Third crackdown quarter: 0.74 (0.53–1.05)  Fourth crackdown quarter: 0.57 (0.35–0.92) |  |
| Hope 2014^17^ |  | Arrested last year | Self-reported abscess “(a swelling containing pus)”, past 28 days | “No associations found” |  |  |
| Hope 2014^17^ |  | Arrested last year | Self-reported “redness, swelling, and tenderness”, past 28 days | Not in last year: 44/281 (16%) In last year: 133/574 (23%) p=0.011 | Not in last year: 1.00  In last year: 1.61 (1.07 – 2.43) |  |
| Hope 2015^18^ |  | Arrested | Abscess, past 12 months, self-reported | Not in preceding year: 42/281  Yes in preceding year: 118/574 |  |  |
| Hope 2015^18^ |  | Arrested | Cellulitis (redness, swelling, or tenderness), past 12 months, self-reported | “Not associated” |  |  |
| Pollini 2010^39^ |  | Arrested for sterile syringes, past 6 mos. | Past 6-months abscess | Yes: 14/59  No: 113/564  P=0.50 |  |  |
| Pollini 2010^39^ |  | Arrested for used syringes, past 6 mos. | Past 6-months abscess | Yes: 11/71  No: 116/552  P=0.44 |  |  |
| Pollini 2010^39^ |  | Arrested for track marks, past 6 mos. | Past 6-months abscess | Yes: 29/104  No: 98/519  P=0.03 |  |  |
| Pollini 2010^39^ |  | Police asked you for money, past 6 mos. | Past 6-months abscess | Yes: 50/188  No: 77/435  P=0.02 |  |  |
| Pollini 2010^39^ |  | Police affected where you use drugs, past 6 mos. | Past 6-months abscess | Yes: 19/54  No: 108/569  P=0.02 | aOR 2.14 (1.15–3.96) |  |
| **Study** |  | **Exposures** | **Outcomes** | **Unadjusted effect estimate** | **Adjusted effect estimate** |  |
| Harm reduction and drug treatment | | | | | | |
| Needle and syringe programs | | | | | | |
| Bhattacharya 2006^5^ |  | Implementation of needle and syringe program in October 2000 | Monthly prevalence of skin abscess in community-based cohort |  | Abscess rates were as high as 22.6% in September 2000, whereas they declined to very low levels (at times 0%; e.g. during October 2001), after the intervention started. |  |
| Binswanger 2000^6^ |  | Used syringe exchange program, past 30 days | Current abscess or cellulitis on physical examination | 1.0 (0.4 – 2.20) |  |  |
| Dunleavy 2017^15^ |  | High needle and syringe uptake (>/=200% uptake) vs. low | SSTI past year | OR 0.65 (0.53-0.80), p<0.001  High: 270/1085 (25%)  Low: 258/767 (34%) | aOR 0.72 (0.58-0.89), p=0.002 |  |
| Dunleavy 2017^15^ |  | High paraphernalia uptake (>/=200% uptake) vs. low | SSTI past year | OR 0.71 (0.58–0.87), p=0.001  High: 270/1060 (25%)  Low: 258/792 (33%) | aOR 0.77 (0.63–0.95), p=0.015 |  |
| Dunleavy 2017^15^ |  | High injecting equipment uptake (>/=200% uptake) vs. low | SSTI past year | OR 0.706 (0.576–0.865), p=0.001  High: 246/979 (25%) Low: 280/869 (32%) | aOR 0.775 (0.628–0.956), p=0.017 |  |
| Fink 2013^16^ |  | Syringe exchange program client | SSTI past six months, self-report | Yes: 220/587 (37%)  No: 100/270 (37%), p=0.90 | aOR 0.91 (0.65–1.27) |  |
| Hope 2010^19^ |  | Used needle exchange last year | Self-report, ‘swelling containing pus (abscess), sore, or open wound’ at an injection site during the previous 12 months | No: 150/566 (27%) Yes: 1713/4643 (37%) P<0.001 | No: 1.00 Yes: 1.65 (1.35-2.01) |  |
| Hope 2010^19^ |  | Used needle exchange last year | Self-report, ‘swelling containing pus (abscess), sore, or open wound’ at an injection site during the previous 12 months  (among subgroup who report injecting in past 4 weeks) | No: 75/270 (28%) Yes: 1300/3463 (38%) P<0.001 | No: 1.00 Yes: 1.44 (1.07-1.93) |  |
| Noroozi 2019^34^ |  | NSP utilization during last 6 mo. | Lifetime injection site infection | ORs  Yes: 0.4 (0.30-0.71)  No: 1 | aORs  0.5 (0.32-0.78) |  |
| Tomolillo 2007^53^ |  | Number of needle exchanged, per week | Number of abscesses treated at clinic associated with NSP, per week | “Analyses with ARIMA (*p* = 1) yielded a significant neg- ative relationship between number of needles exchanged and number of abscesses treated, *b* = −.001, *t* (109) = −3.1, *p* = .002” |  |  |
| Tomolillo 2007^53^ |  | Number of visits to needle exchange program, per week | Number of abscesses treated at clinic associated with NSP, per week | “ARIMA (*p* = 1) yielded a significant negative relationship be- tween number of needle exchange program visits and the number of abscesses treated in the community, *b* = −.12, *t* (109) = −3.7, *p* < .001 ” |  |  |
| Tomolillo 2007^53^ |  | Self-report number of needles exchanged (timeline not specified) | Self-report occurrences of abscesses (timeline not specified) | “Regression analysis yielded a significant negative relationship between number of needles exchanged and occurrences of abscesses, *R*2 = .10, *F* (1, 60) = 6.41, *p* = .01, *B* = −.68 ” |  |  |
| Tomolillo 2007^53^ |  | Self-reported “more use of sterile equipment” (timeline not specified) | Self-report occurrences of abscesses (timeline not specified) | “Use of sterile injecting equipment was not related to abscesses reported, *R*2 = .10, *F* (3, 58) = 2.23, *p* = .10 ” |  |  |
| Wurcel 2018^59^ |  | Obtains needles at Needle Exchange Program | Abscess ever | OR  Among females: 0.48 (0.19, 1.24)  Among males: 1.43 (0.80, 2.56) | aOR  Among females: 0.27 (0.07, 1.01)  Among males: 1.31 (0.65, 2.63) |  |
| **Study** |  | **Exposures** | **Outcomes** | **Unadjusted effect estimate** | **Adjusted effect estimate** |  |
| Barriers to needle/syringe access - Access to needles/syringes outside of NSP harm reduction programs | | | | | | |
| Pollini 2010b ^40^ |  | Refused/overcharged syringes when trying to purchase at pharmacy | Abscess last 6 months | Refused/overcharged: 20/100  No: 108/527  P=0.91 |  |  |
| Pollini 2010b ^40^ |  | Refused/overcharged syringes when trying to purchase at pharmacy | Abscess ever | Refused/overcharged: 48/100  No: 240/527  P=0.65 |  |  |
| Pollini 2010b ^40^ |  | Refused/overcharged syringes when trying to purchase at pharmacy | Median # abscesses (lifetime) | Refused/overcharged: 0 (0-3)  No: 0 (0-2)  P=0.195 | aOR 1.02 (1.00, 1.03) |  |
| Tomolillo 2007^53^ |  | Implementation of policy restricting the number of needles distributed per person and requiring appointment for access | Abscesses treated at clinic associated with needle exchange | Mean (SD) abscesses treated each week - Before policy change: 8.51 (3.18)  - After policy change: 14.34 (5.95)  Mean (SD) needles distributed each week  - Before policy change: 3268.32 (965.25)  - After policy change: 470.53 (320.75)  No statistical test/p-value |  |  |
| Supervised consumption site / overdose prevention site / SIF | | | | | | |
| **Study** |  | **Exposures** | **Outcomes** | **Unadjusted effect estimate** | **Adjusted effect estimate** |  |
| Scherbaum 2010^45^ |  | Attendance at SCS | Past-month skin abscess, self -reported during follow-up interview at SCS | Among 71 people with 1 month follow-up:   - 3/71 (4.2%) at baseline vs. 6/71 (8.5%) at 1 month   Among 38 people with 2 month follow-up:   - 3/38 (7.9%) at baseline vs. 2/38 (5.3%) at 2 months   Among 26 people with 3 month follow-up:   - 2/26 (7.7%) at baseline vs. 3/26 (1.5%) at 3 months   “There were no statistically significant intra-individual changes from M0 to M1, M2, or M3; McNemar tests, all at p > 0.3 or greater” |  |  |
| Lloyd-Smith 2008^26^ |  | SIF use | Current injecting-related skin infection (self-report & confirmed by study nurse) | OR  Less than always: Ref  Always use: 0.47 (0.23 – 0.94) | aOR  0.58 (0.29 – 1.19) |  |
| Milloy 2010^30^ |  | >=75% of injections at supervised injection facility | ED visit for cutaneous injecting-related infection (admin data) from SIF/community cohort | At baseline, ED visit in previous 6 month:  No: 49/367  Yes: 23/178  P=0.889  RR for number of ED visits for CIRI during follow-up:  1.25 (1.06–1.48)  (yes vs. no) |  |  |
| **Study** |  | **Exposures** | **Outcomes** | **Unadjusted effect estimate** | **Adjusted effect estimate** |  |
| Opioid agonist treatment | | | | | | |
| Bassetti 2002^2^ |  | Enrolling in injection opiate maintenance program | Skin infections requiring hospitalisation (Abscesses, phlegmonous infections, erysipelas, ulcerations, and necrosis); frequency and incidence per 100 patient-years before and after | Before admission into program: 20 (3.8/100 patient-years[PY])  During program: 21 (4.6/100PY) |  |  |
| Bassetti 2002^2^ |  | Enrolling in injection opiate maintenance program | Bloodstream infections requiring hospitalisation; frequency and incidence per 100 patient-years, before and after | Before admission into program: 5 (0.9/100PY)  During program: 3 (0.7/100PY) |  |  |
| Bertin 2020^3^ |  | Prescribed morphine sulfate as opioid maintenance, vs. buprenorphine and vs. methadone | 1-year risk of hospitalizations for bacterial infections associated with intravenous injecting | Crude incidence per 100,000 PY, by cohort  MS: 7.0 (4.7 – 10.6)  Bupe: 2.2 (1.8 – 2.5)  Methadone: 1.6 (1.2 – 2.0) | aHR 2.8 (1.8-4.4) vs. patients starting buprenorphine  aHR 3.6 (2.2-5.9) vs. patients starting methadone |  |
| Betts 2016^4^ |  | NOT receiving OST treatment (vs. yes receiving) | Past-month abscess, self-report |  | aOR 0.97 (0.71 – 1.33)  so inverse is 1.03 (0.75 - 1.41) |  |
| Dunleavy 2017^15^ |  | Opiate substitution treatment | Past year SSTI, self-report | Past: 139/380 (36%) Never: 42/162 (26%)  Current: 352/1320 (27%)  OR  Past: 1  Never: 0.622 (0.413–0.936), p=0.023  Current 0.646 (0.508–0.822), p<0.001 | aOR  Past: 1  Never: 0.593 (0.386–0.910),p=0.017  Current: 0.672 (0.524–0.862), p=0.002 |  |
| Hope 2010^19^ |  | “Prescribed treatment for their drug use” | Self-report, ‘swelling containing pus (abscess), sore, or open wound’ at an injection site during the previous 12 months | Never in treatment: 206/706 (29%) Currently scripted: 1321/3570 (37%) Previously scripted: 336/933 (36%) p<0.001 |  |  |
| Hope 2008^20^ |  | Received prescribed substitute drug | Self-reported symptoms of injection site infections (abscess or open wound), past year | ORs Currently: 1.0 Previously: 1.6 (1.2 – 2.2)  Never: 0.6 (0.4 – 0.9) | aORs Currently: 1.0 Previously: 1.7 (1.3 – 2.4) Never: 0.9 (0.5 – 1.3) |  |
| Milloy 2010^30^ |  | Current MMT | ED visit for cutaneous injecting-related infection (admin data) from SIF/community cohort | At baseline, ED visit in previous 6 month:  No: 53/383  Yes: 19/159 p=0.577  RR for number of ED visits for CIRI during follow-up:  0.88 (0.72–1.08)  (yes vs. no) | aRR for number of ED visits for CIRI during follow-up: 0.92 (0.75–1.13) |  |
| Morin 2020^31^ |  | Receiving OAT | Infective Endocarditis (diagnostic code in administrative data, but date/timing unclear) | OR 0.6 (0.5–0.9) | aOR 0.71 (0.55-0.93) |  |
| Morin 2020^31^ |  | Receiving OAT | Osteomyelitis (diagnostic code in administrative data, but date/timing unclear) | OR 0.7 (0.6–0.8) | aOR 0.94 (0.91–0.93) |  |
| Morin 2020^31^ |  | Receiving OAT | Septic Arthritis (administrative data, but date/timing unclear) | OR 0.6 (0.4–0.9) | aOR 0.93 (0.92–0.94) |  |
| Oviedo-Joekes 2017^35^ |  | Injectable OAT with hydromorphone vs. diacetylmorphine | Cellulitis or abscess | HDM: 7 episodes among 100 patients  DAM: 17 episodes among 100 patients |  |  |
| Roux 2020^42^ |  | On opioid agonist treatment | At least one cutaneous abscess in the previous six months | OR  No: Ref  Yes: 0.73 [0.39,1.37] |  |  |
| Sierra 2006^47^  #482 |  | Methadone program | Invasive soft-tissue Group A Strep (S. pyogenes) infections in Barcelona | OR 0.03 (0-0.19)  Methadone: 0/12  No methadone: 15/46 |  |  |
| Stein 2020^50^ |  | MOUD, past 3 months | Number of ED visits for injecting-related infections in 12 months following educational intervention |  | IRR 0.98 (0.61, 1.59) |  |
| Stein 2020^50^ |  | MOUD, past 3 months | Number of hospitalizations visits for injecting-related infections in 12 months following educational intervention |  | IRR 0.95 (0.58, 1.56) |  |
| Thønnings 2020^52^ |  | Opioid substitution treatment | Bacteraemia, among hospitalised PWID | OR 2.25 (0.90–5.60) |  |  |
| Combined harm reduction interventions | | | | | | |
| Dunleavy 2017^15^ |  | Combined Injecting Equipment uptake & current OST  Low = Low IE (<200% uptake), no OST; Medium = Low IE + OST, or High IE + no OST; High = High IE + OST (where No OST = never and in the past; OST = currently prescribed). | Past year SSTI, self-report | Low: 117/316 (37%) Medium: 225/777 (28%)  High: 184/754 (24%)  OR Low: 1 (ref) Medium: 0.693 (0.526–0.914), p=0.022  High: 0.549 (0.414–0.728), p=0.000 | aOR Low: 1 (ref) Medium: 0.732 (0.551–0.973), p=0.032 High: 0.622 (0.463–0.834), p=0.002 |  |
| Other substance use treatment | | | | | | |
| Binswanger 2000^6^ |  | “Substance abuse treatment” (not defined) | Current abscess or cellulitis on physical examination | “did not differ significantly” |  |  |
| Thønnings 2020^52^ |  | “Contact to an addiction treatment center” (not defined) | Bacteraemia, among hospitalised PWID | OR 1.21 (0.64–2.26) |  |  |
| **Study** |  | **Exposures** | **Outcomes** | **Unadjusted effect estimate** | **Adjusted effect estimate** |  |

### Appendix 13. List of exposure-outcome pair effect estimates for studies where outcome occurs during treatment of injecting-related bacterial infections drug use-associated bacterial and fungal infections, included in quantitative systematic review. Blank cells represent where effect estimate (or frequencies) were not reported in the paper.

| **Study** | **Exposures** | **Outcomes** | **Unadjusted effect estimate** | **Adjusted effect estimate** |
| --- | --- | --- | --- | --- |
| **Outcome: healthcare seeking for injecting-related infections** | | | |  |
| *Exposures: Gender/sex* | | | | |
| Hope 2008^20^ | Gender | Seeking health care among those having had injection site infections in the previous year | “…was not associated with…” |  |
| Hope 2015^18^ | Gender | Seeking medical advice about an injection-site abscess, past 12 months | Male: 79/121  Female: 33/39  P=0.022 | Male: Ref  Female: 3.04 (1.14 – 8.13) |
| Hope 2015^18^ | Gender | Seeking medical advice about injection-site cellulitis (“redness, swelling, and tenderness”), past 12 months | Male: 105/306  Female: 54/109  P=0.005 | Male: Ref  Female: 2.41 (1.49 – 3.91) |
| *Age* |  |  |  |  |
| Hope 2008^20^ | Age | Seeking health care among those having had injection site infections in the previous year | “…was not associated with…” |  |
| Hope 2015^18^ | Age | Seeking medical advice about an injection-site abscess, past 12 months | “not associated” |  |
| Hope 2015^18^ | Age | Seeking medical advice about injection-site cellulitis (“redness, swelling, and tenderness”), past 12 months | “not associated” |  |
| Income/employment | | | | |
| Hope 2015^18^ | Main source of income | Seeking medical advice about an injection-site abscess, past 12 months | “not associated” |  |
| Hope 2015^18^ | Main source of income | Seeking medical advice about injection-site cellulitis (“redness, swelling, and tenderness”), past 12 months | Licit: 95/272  Illicit: 63/141  P=0.053 |  |
| *Sex work* |  |  |  |  |
| Hope 2015^18^ | Sex preceding year (paid / sex work) | Seeking medical advice about an injection-site abscess, past 12 months | “not associated” |  |
| Hope 2015^18^ | Sex preceding year (paid / sex work) | Seeking medical advice about injection-site cellulitis (“redness, swelling, and tenderness”), past 12 months | No: 29/72  Yes, but not paid: 111/311  Yes, but paid: 18/30  P=0.03 |  |
| *Housing* |  |  |  |  |
| Hope 2008^20^ | Homelessness | Seeking health care among those having had injection site infections in the previous year | “…was not associated with…” |  |
| Hope 2015^18^ | Homelessness | Seeking medical advice about an injection-site abscess, past 12 months | “not associated” |  |
| Hope 2015^18^ | Homelessness | Seeking medical advice about injection-site cellulitis (“redness, swelling, and tenderness”), past 12 months | “not associated” |  |
| *Incarceration* |  |  |  |  |
| Hope 2008^20^ | Having been imprisoned | Seeking health care among those having had injection site infections in the previous year | “…was not associated with…” |  |
| Hope 2015^18^ | Imprisonment | Seeking medical advice about an injection-site abscess, past 12 months | “not associated” |  |
| Hope 2015^18^ | Imprisonment | Seeking medical advice about injection-site cellulitis (“redness, swelling, and tenderness”), past 12 months | Never: 31/81  Yes, not preceding year: 88/202  Yes, preceding year: 39/131  P=0.04 |  |
| *Overdose history* | | | | |
| Hope 2008^20^ | Having had an overdose | Seeking health care among those having had injection site infections in the previous year | “…was not associated with…” |  |
| *Migration status* | | | | |
| Hope 2015^18^ | Migration, years lived in current area | Seeking medical advice about an injection-site abscess, past 12 months | “not associated” |  |
| Hope 2015^18^ | Migration, years lived in current area | Seeking medical advice about injection-site cellulitis (“redness, swelling, and tenderness”), past 12 months | “not associated” |  |
| *Heroin* |  |  |  |  |
| Hope 2015^18^ | Injecting heroin, predecing year | Seeking medical advice about an injection-site abscess, past 12 months | “not associated” |  |
| Hope 2015^18^ | Injecting heroin, predecing year | Seeking medical advice about injection-site cellulitis (“redness, swelling, and tenderness”), past 12 months | “not associated” |  |
| *Cocaine* |  |  |  |  |
| Hope 2008^20^ | Injecting crack-cocaine | Seeking health care among those having had injection site infections in the previous year | “…was not associated with…” |  |
| Hope 2015^18^ | Injected cocaine preceding year | Seeking medical advice about an injection-site abscess, past 12 months | “not associated” |  |
| Hope 2015^18^ | Injected cocaine preceding year | Seeking medical advice about injection-site cellulitis (“redness, swelling, and tenderness”), past 12 months | No: 119/344  Yes: 40/71  P<0.001 | No: Ref  Yes: 2.37 (1.36 – 4.14) |
| Hope 2015^18^ | Injected crack preceding year | Seeking medical advice about an injection-site abscess, past 12 months | “not associated” |  |
| Hope 2015^18^ | Injected crack preceding year | Seeking medical advice about injection-site cellulitis (“redness, swelling, and tenderness”), past 12 months | No: 45/149  Yes: 113/265  P=0.012  1.71 (1.12-2.63) |  |
| *Amphetamines* | | | | |
| Hope 2008^20^ | Injecting amphetamines | Seeking health care among those having had injection site infections in the previous year | “…was not associated with…” |  |
| Hope 2015^18^ | Injected amphetamines preceding year | Seeking medical advice about an injection-site abscess, past 12 months | “not associated” |  |
| Hope 2015^18^ | Injected amphetamines preceding year | Seeking medical advice about injection-site cellulitis (“redness, swelling, and tenderness”), past 12 months | “not associated” |  |
| *Opioid agonist treatment* | | | | |
| Hope 2008^20^ | Received prescribed substitute drug | Seeking health care among those having had injection site infections in the previous year | ORs Currently: 1.0 Previously: 0.6 (0.4 – 0.9)  Never: 0.2 (0.1 – 0.5) | aORs Currently: 1.0 Previously: 0.5 (0.3 – 0.9)  Never: 0.3 (0.1 – 0.7) |
| *Supervised consumption site* | | | | |
| Lloyd-Smith 2012^25^ | Referral from nurse at supervised consumption site | ED visit for cutaneous injecting-related infection (admin data) from SIF/community cohort | OR 4.69 (2.76 – 7.97)  Among females:  HR 5.06 (3.14 – 8.17)  Among males: HR 3.28 (2.14 – 5.04) | Among females:  aHR 4.48 (2.76 – 7.30)  Among males: aHR 2.97 (1.93 – 4.57) |
| Lloyd-Smith 2010^28^ | Referral from nurse at supervised consumption site | Hospitalization for injecting-related infection (cellulitis, abscess, osteomyelitis, Staph infection, endocarditis, septic arthritis, ulcer, thrombophlebitis, myositis) | HR 2.41 (1.55 – 3.77) | aHR 5.38 (3.39 – 8.55) |
| *Outcome: self-treated abscess* | | | | |
| *Gender/sex* | | | | |
| *Fink 2013*^16^ | Age (years) | Self-treated last abscess, among people ever having an abscess requiring treatment | <30: 14/22 (64%)  30-39: 33/74 (45%)  40-49: 108/209 (52%)  50+: 105/234 (45%)  P=0.21 | aOR  <30: 1  30-39: 0.91 (0.29 – 2.90)  40-49: 1.31 (0.44 – 3.90)  50+: 1.49 (0.49 – 4.56) |
| Monteiro 2020^71^ | Age (units unknown) | Self-treatment of SSTI among PWID who had SSTI in past year | OR 0.98 (0.95; 1.01) | aOR 0.95 (0.94; 1.01) |
| *Gender/sex* |  |  |  |  |
| *Fink 2013*^16^ | Gender | Self-treated last abscess, among people ever having an abscess requiring treatment | Male: 177/380 (47%)  Female: 84/164 (51%)  P=0.32 | Male: 1  Female: 1.21 (0.79 – 1.85) |
| Monteiro 2020^71^ | Sex (male) | Self-treatment of SSTI among PWID who had SSTI in past year | OR 0.60 (0.31; 1.70) | aOR 1.05 (0.40; 2.83) |
| *Race/ethnicity* | | | | |
| *Fink 2013*^16^ | Race | Self-treated last abscess, among people ever having an abscess requiring treatment | White: 55/109 (50%) Black: 70/186 (38%) Latino: 116/208 (56%) Other: 15/32 (47%)  P=0.004 | White: 1  Black: 0.60 (0.34 – 1.04) Latino: 1.57 (0.95 – 2.60)  Other: Not reported  “Latino vs. Black” “Odds Ratio 2.62 (1.66-4.13)” (from table)  “AOR 2.83, 1.6-5.1” (from text) |
| Monteiro 2020^71^ | Race (white) | Self-treatment of SSTI among PWID who had SSTI in past year | OR 1.16 (0.59; 2.28) | aOR 1.06 (0.34; 3.31) |
| Monteiro 2020^71^ | Hispanic (yes) | Self-treatment of SSTI among PWID who had SSTI | OR 0.92 (0.47; 1.80) | aOR 1.32 (0.55; 3.17) |
| *Housing* |  |  |  |  |
| *Fink 2013*^16^ | Homelessness | Self-treated last abscess, among people ever having an abscess requiring treatment | Yes: 144/294 (49%)  No: 114/242 (47%)  P=0.67 |  |
| *Needle and syringe program* | | | | |
| *Fink 2013*^16^ | Syringe exchange program client | Self-treated last abscess, among people ever having an abscess requiring treatment | Yes: 174/362 (48%)  No: 87/182 (48%)  P=0.95 | Yes: 0.88 (0.58 – 1.32)  No: 1 |
| *Access to healthcare* | | | | |
| *Fink 2013*^16^ | Access to medical care | Self-treated last abscess, among people ever having an abscess requiring treatment | No usual place for care: 110/190 (58%)  Have a usual place for care: 151/354 (43%)  P=0.0007 | No: 1  Yes: 0.61 (0.40 – 0.92) |
| *Fink 2013*^16^ | Usual place of care | Self-treated last abscess, among people ever having an abscess requiring treatment | Private doctor’s office: 24/36 (67%) Community clinic: 42/97 (43%)  Hospital outpatient clinic: 24/84 (29%)  Emergency room: 44/93 (47%) Syringe exchange program: 4/4 (100%) Other: 19/42 (45%)  Refuse to answer: 6/10 (60%)  P<0.0001 |  |
| Monteiro 2020^71^ | Has a primary care provider (yes) | Self-treatment of SSTI among PWID who had SSTI | OR 0.57 (2.29; 1.12) | aOR 1.14 (0.44; 2.95) |
| Monteiro 2020^71^ | Trust in the medical profession | Self-treatment of SSTI among PWID who had SSTI | OR 0.89 (0.59; 1.36) | aOR 0.96 (0.53; 1.71) |
| *Fink 2013*^16^ | Insurance | Self-treated last abscess, among people ever having an abscess requiring treatment | No insurance: 162/323 (50%)  Insured: 98/220 (45%)  P=0.20 | No: 1  Yes: 0.93 (0.62 – 1.39) |
| *Heroin* |  |  |  |  |
| Monteiro 2020^71^ | Days injecting heroin (past 3 months) | Self-treatment of SSTI among PWID who had SSTI | OR 1.00 (0.99; 1.01) | aOR 1.00 (0.99; 1.01) |
| *Cocaine* |  |  |  |  |
| Monteiro 2020^71^ | Days injecting cocaine (past 3 months) | Self-treatment of SSTI among PWID who had SSTI | OR 1.00 (0.99; 1.01) | aOR 0.99 (0.97; 1.01) |
| **Study** | **Exposures** | **Outcomes** | **Unadjusted effect estimate** | **Adjusted effect estimate** |
| **Outcome: hospital admission for SSTI, among people reporting SSTI** | | | | |
| *Age* | | | | |
| Hope 2015^18^ | Age | Hospital admission for infection when had injection site abscess, past 12 months | <25: 0/5  25-29: 9/30  30-34: 15/31  >=35: 27/47  P=0.019 |  |
| Hope 2015^18^ | Age | Hospital admission for infection when had injection site cellulitis (redness, swelling, tenderness), past 12 months | <25: 1/16  25-29: 18/49  30-34: 10/42  >=35: 18/51  P=0.077 |  |
| Takahashi 2007^78^ | Age | Hospitalized for SSTI, among ED patients with injecting-related infection | Mean age, (SD), yrs Hospitalized: 41 (8)  Discharged from ED: 43 (8)  P=0.19 |  |
| *Gender* | | | | |
| Hope 2015^18^ | Gender | Hospital admission for infection when had injection site abscess, past 12 months | Male: 41/79  Female: 10/33  P=0.036 |  |
| Hope 2015^18^ | Gender | Hospital admission for infection when had injection site cellulitis (redness, swelling, tenderness), past 12 months | Male: 36/105  Female: 11/53  P=0.079 |  |
| Takahashi 2007^78^ | Female | Hospitalized for SSTI, among ED patients with injecting-related infection | Female: 24/52  Male: 31/84 |  |
| *Race/ethnicity* | | | | |
| Takahashi 2007^78^ | Non-Hispanic White | Hospitalized for SSTI, among ED patients with injecting-related infection | Non-Hispanic White: 36/85  Other: 19/51 |  |
| *Housing* | | | | |
| Takahashi 2007^78^ | Living situation | Hospitalized for SSTI, among ED patients with injecting-related infection | OR (95% CI)  Owns or rents a home: Ref  Lives in a shelter: 5.6 (1.6–19.0) | Owns or rents a home: Reference  Lives in a shelter: 4.2 (1.2–15.1)  Lives on the streets: 1.4 (0.5–4.1)  Other: 1.1 (0.5–2.8) |
| Hope 2015^18^ | Homelessness (Never; Yes, not preceding year; Yes, preceding year) | Hospital admission for infection when had injection site abscess, past 12 months | “Not associated” |  |
| Hope 2015^18^ | Homelessness (Never; Yes, not preceding year; Yes, preceding year) | Hospital admission for infection when had injection site cellulitis (redness, swelling, tenderness), past 12 months | “Not associated” |  |
| *Education* |  |  |  |  |
| Takahashi 2007^78^ | Graduated from high school/GED | Hospitalized for SSTI, among ED patients with injecting-related infection | Graduated: 42/104  Did not: 13/32  p=0.97 |  |
| Income/employment |  |  |  |  |
| Takahashi 2007^78^ | Currently employed | Hospitalized for SSTI, among ED patients with injecting-related infection | Employed: 6/15  Unemployed: 49/121  p=1.00 |  |
| Hope 2015^18^ | Main source of income (licit vs. illicit) | Hospital admission for infection when had injection site abscess, past 12 months | “Not associated” |  |
| Hope 2015^18^ | Main source of income (licit vs. illicit) | Hospital admission for infection when had injection site cellulitis (redness, swelling, tenderness), past 12 months | “Not associated” |  |
| Sex work |  |  |  |  |
| Hope 2015^18^ | Sex preceding year (sex work / paid) | Hospital admission for infection when had injection site abscess, past 12 months | “Not associated” |  |
| Hope 2015^18^ | Sex preceding year (sex work / paid) | Hospital admission for infection when had injection site cellulitis (redness, swelling, tenderness), past 12 months | “Not associated” |  |
| Migration |  |  |  |  |
| Hope 2015^18^ | Migration, years lived in current area | Hospital admission for infection when had injection site abscess, past 12 months | “Not associated” |  |
| Hope 2015^18^ | Migration, years lived in current area | Hospital admission for infection when had injection site cellulitis (redness, swelling, tenderness), past 12 months | “Not associated” |  |
| Incarceration | | | | |
| Hope 2015^18^ | Imprisonment | Hospital admission for infection when had injection site abscess, past 12 months | “Not associated” |  |
| Hope 2015^18^ | Imprisonment | Hospital admission for infection when had injection site cellulitis (redness, swelling, tenderness), past 12 months | Never: 4/31  Yes, not preceding year: 29/89  Yes, preceding year: 15/39  P=0.052 |  |
| Takahashi 2007^78^ | Reported time in jail, prison, or juvenile detention | Hospitalized for SSTI, among ED patients with injecting-related infection | History of incarceration: 41/112  Not: 14/24  p=0.18 |  |
| Health care access |  |  |  |  |
| Takahashi 2007^78^ | Two or more hospitalizations past year | Hospitalized for SSTI, among ED patients with injecting-related infection | OR 2.7 (0.9–7.7) | aOR 4.4 (1.6–11.8) |
| Takahashi 2007^78^ | Insurance | Hospitalized for SSTI, among ED patients with injecting-related infection | Medicare/Medicaid: 39/79  Self pay: 13/29  Other/unknown: 3/31 p=0.96 |  |
| Takahashi 2007^78^ | Has a primary care provider | Hospitalized for SSTI, among ED patients with injecting-related infection | Yes: 19/54  No: 36/80  p=0.26 |  |
| Takahashi 2007^78^ | Considered coming in for the infection. Before today | Hospitalized for SSTI, among ED patients with injecting-related infection | Yes: 43/106  No: 12/30  p=0.80 |  |
| Takahashi 2007^78^ | One or more days of ANY symptoms | Hospitalized for SSTI, among ED patients with injecting-related infection | Yes: 42/101 No: 13/35  p=0.30 |  |
| Takahashi 2007^78^ | One or more days of SYSTEMIC symptoms | Hospitalized for SSTI, among ED patients with injecting-related infection | Yes: 50/114  No: 5/22  p=0.30 |  |
| Takahashi 2007^7^ | Self-treatment with oral antibiotics | Hospitalized for SSTI, among ED patients with injecting-related infection | Yes: 14/35 No: 41/101  p=1.00 |  |
| Takahashi 2007^7^ | Self-treatment with incision and drainage | Hospitalized for SSTI, among ED patients with injecting-related infection | Yes: 22/55  No: 33/81  p=1.00 |  |
| Overdose history |  |  |  |  |
| Hope 2015^18^ | Overdose history (No; Yes, not preceding year; Yes, preceding year) | Hospital admission for infection when had injection site abscess, past 12 months | “Not associated” |  |
| Hope 2015^18^ | Overdose history (No; Yes, not preceding year; Yes, preceding year) | Hospital admission for infection when had injection site cellulitis (redness, swelling, tenderness), past 12 months | “Not associated” |  |
| *Heroin* | | | | |
| Takahashi 2007^78^ | Most frequently used drug is heroin only | Hospitalized for SSTI, among ED patients with injecting-related infection | Yes: 51/119  No: 4/17  p=0.07 |  |
| Hope 2015^18^ | Injected heroin preceding year | Hospital admission for infection when had injection site abscess, past 12 months | “Not associated” |  |
| Hope 2015^18^ | Injected heroin preceding year | Hospital admission for infection when had injection site cellulitis (redness, swelling, tenderness), past 12 months | “Not associated” |  |
| *Cocaine* |  |  |  |  |
| Hope 2015^18^ | Injected cocaine preceding year | Hospital admission for infection when had injection site abscess, past 12 months | “Not associated” |  |
| Hope 2015^18^ | Injected cocaine preceding year | Hospital admission for infection when had injection site cellulitis (redness, swelling, tenderness), past 12 months | “Not associated” |  |
| Hope 2015^18^ | Injected crack preceding year | Hospital admission for infection when had injection site abscess, past 12 months | No: 7/33  Yes: 44/80  P=0.001 | No: Ref  Yes: 7.49 (2.50 – 22.50) |
| Hope 2015^18^ | Injected crack preceding year | Hospital admission for infection when had injection site cellulitis (redness, swelling, tenderness), past 12 months | No: 9/45  Yes: 39/114  P=0.079 |  |
| *Amphetamines* | | | | |
| Hope 2015^18^ | Injected amphetamine preceding year | Hospital admission for infection when had injection site abscess, past 12 months | “Not associated” |  |
| Hope 2015^18^ | Injected amphetamine preceding year | Hospital admission for infection when had injection site cellulitis (redness, swelling, tenderness), past 12 months | “Not associated” |  |
| Needle and syringe program | | | | |
| Takahashi 2007^78^ | Reported using a needle exchange program | Hospitalized for SSTI, among ED patients with injecting-related infection | Yes: 51/121 No: 4/15 p=0.14 |  |
| Alcohol |  |  |  |  |
| Takahashi 2007^78^ | Hazardous drinking (AUDIT-C score >4) | Hospitalized for SSTI, among ED patients with injecting-related infection | Yes: 13/38 No: 42/98 p=0.34 |  |
| **Outcome: Against medical advice discharge** | | | |  |
| *Gender/sex* |  |  |  |  |
| Eaton 2020^62^ | Gender (female) | Patient-directed discharge  (among patients being treated for injecting-related infections) | OR 4.89 (1.22-19.65) | aOR 3.31 (0.64-17.19) |
| Jo 2021^63^ | Biological sex | Patient-directed discharge  (among patients with untreated OUD and either endocarditis or osteomyelitis) |  | aOR Male: 0.83 (0.62 – 1.11)  Female: Ref |
| Kimmel 2020^64^ | Sex | Discharge AMA  (Among patients with IDU-IE) |  | aOR  Female: 1.21 (1.04, 1.41)  Male: Ref |
| Mertz 2008^70^ | Male gender | “Patient’s non-compliance… “if the patient did not comply with diagnostic measures or adhere to therapeutic measures, left the hospital against medical advice, continued intravenous drug use during hospitalization, smoked in the room, or assaulted hospital staff.”” |  | aOR 1.2 (0.6 – 2.2) |
| Nolan 2020^72^ | Female | AMA Discharge | OR 1.69 (1.03–2.79) | aOR 2.37 (1.34– 4.20) |
| Serota 2021^76^ | Biological sex | AMA Discharge |  | Female: Ref  Male: 0.86 (.80, .92) |
| *Age* |  |  |  |  |
| Jo 2021^63^ | Age (?continuous) | Patient-directed discharge  (among patients with untreated OUD and either endocarditis or osteomyelitis) |  | aOR 0.96 (0.94, 0.97) |
| Kimmel 2020^64^ | Age | Discharge AMA  (Among patients with IDU-IE) |  | aOR  18-24: 3.02 (2.10, 4.34)  25-34: 2.87 (2.08, 3.95)  35-44: 2.31 (1.66, 3.21)  45-55: 1.61 (1.16, 1.41)  56-65: Ref |
| Mertz 2008^70^ | Age (per 10 years older | “Patient’s non-compliance… “if the patient did not comply with diagnostic measures or adhere to therapeutic measures, left the hospital against medical advice, continued intravenous drug use during hospitalization, smoked in the room, or assaulted hospital staff.”” |  | aOR 0.8 (0.5 – 1.2) |
| Serota 2021^76^ | Age | AMA Discharge |  | aRR  18–34: 4.87 (3.41, 6.96)  35–54: 3.97 (2.79, 5.63)  55–64: 2.20 (1.53, 3.15)  65–75: REF |
| *Race/ethnicity* | |  |  |  |
| Jo 2021^63^ | Ethnicity | Patient-directed discharge  (among patients with untreated OUD and either endocarditis or osteomyelitis) |  | aOR White: 1.24 (0.82 – 1.87)  Other: Ref |
| Kimmel 2020^64^ | Race/Ethnicity | Discharge AMA  (Among patients with IDU-IE) |  | aOR  White: Ref Black: 1.13 (0.89 – 1.45)  Hispanic: 1.32 (1.03 – 1.69)  Asian: 0.59 (0.18 – 1.94)  Native American: 1.01 (0.42 – 2.46)  Other: 0.99 (0.62 – 1.59) |
| Serota 2021^76^ | Race | AMA Discharge |  | aRR  Hispanic: 0.97 (.85, 1.10)  Non-Hispanic Black: 0.75 (.65, .86)  Non-Hispanic White: Ref |
| *Unstable housing* |  |  |  |  |
| Cooksey 2020^61^ | Homeless (not otherwise defined) | Hospital discharge against medical advice  (among patients being treated for IDU-IE) | OR 4.6 (1.4-15.0) |  |
| Nolan 2020^72^ | Unstable housing (not otherwise defined) | AMA Discharge | OR 0.85 (0.54–1.32) | aOR 1.39 (0.62–3.12) |
| *Health insurance (?same as “access to health care” larger category)* | | | | |
| Jo 2021^63^ | Insurance status | Patient-directed discharge  (among patients with untreated OUD and either endocarditis or osteomyelitis) |  | aOR Govt funded insurance: 0.45 (0.33, 0.61) Commercial insurance: 0.28 (0.16 – 0.48)  Uninsured: Ref |
| Kimmel 2020^64^ | Payor | Discharge AMA  (Among patients with IDU-IE) |  | aOR  Medicaid: Ref Medicare: 0.75 (0.58 – 0.96)  Commercial: 0.57 (0.44 – 0.74)  Self: 1.37 (1.13 – 1.66)  No charge: 1.24 (0.79 – 1.94)  Other: 0.70 (.046 – 1.05) |
| Nolan 2020^72^ | Uninsured | AMA Discharge | OR 3.93 (2.17–7.13) | aOR 4.10 (2.22–7.58) |
| Serota 2021^76^ | Insurance | AMA Discharge |  | aRR  Private: Ref  Medicare: 1.09 (.93, 1.29)  Medicaid 1.49 (1.29, 1.72)  Uninsured 2.07 (1.81, 2.38) |
| *Income/employment* | | | | |
| Kimmel 2020^64^ | Zip Code Income Quartile | Discharge AMA  (Among patients with IDU-IE) |  | aOR  Quartile 1 (lowest): 1.56 (1.21, 1.99)  Quartile 2: 1.24 (0.96, 1.60)  Quartile 3: 1.16 (0.89, 1.52)  Quartile 4 (highest): Ref |
| *Inpatient addiction medicine consultation* | | | | |
| Marks 2020^66^ | Inpatient addiction medicine consultation  (during hospitalization with endocarditis, fungemia, bacteremia, necrotizing fasciitis or myositis, septic joint, epidural abscess, osteomyelitis) | “Elopement or discharged AMA” | 0.19 (0.08 – 0.48) |  |
| *Opioid agonist treatment* | | | | |
| Eaton 2020^62^ | MOUD | Patient-directed discharge  (among patients being treated for injecting-related infections) | OR 1.63 (0.48-5.57) | aOR 0.83 (0.16-4.31) |
| Jo 2021^63^ | Initiation of OAT (but maybe just for treatment of withdrawal, and/or at insufficient dose…) | Patient-directed discharge  (among patients with untreated OUD and either endocarditis or osteomyelitis) | MOUD: 49 out of 269 No MOUD: 209 out of 1138  p value not tested | aOR 0.85 (0.59 – 1.22) |
| Marks 2020^67^ | MOUD prescription at discharge | AMA discharge | RR 0.49 (0.19 – 1.22)  (Cannot find frequencies to generate 2X2 table, but 15% of sample experienced AMA discharge |  |
| Nolan 2020^72^ | Received MOUD in hospital | AMA Discharge | OR 0.55 (0.34–0.91) | 0.49 (0.28–0.84) |
| Suzuki 2020^77^ | Initiated on MOUD | AMA Discharge  (among patients with IDU-IE) | OR 0.98, 95%CI 0.26 to 3.7 |  |
| Wang 2020^81^ | Any form of medication-assisted therapy (continuation, initiation and linkage, or detox/taper) | AMA discharge  (among patients admitted to hospital with injecting-related infections) | No MAT: 59.7% AMA (out of n = 57)  MAT: 30.0% (out of n = 90)  relative risk [RR] 0.50; 95% confidence interval [CI], 0.34-0.74). |  |
| *Overdose history* | | | | |
| Serota 2021^76^ | Overdose (NOS)… potentially during same admission? | AMA Discharge |  | aRR  0.87 (.74, 1.01) |
| *Substance use* | | | | |
| Jo 2021^63^ | Cocaine/amphetamine use | Patient-directed discharge  (among patients with untreated OUD and either endocarditis or osteomyelitis) |  | aOR Yes: 1.23 (0.85 – 1.77)  No: Ref |
| Nolan 2020^72^ | Opioid use alone (fentanyl or heroin) without stimulants  vs. people who use opioids and stimulants | AMA Discharge | OR 0.54 (0.34–0.87) | aOR 0.84 (0.17–4.06) |
| Nolan 2020^72^ | Opioid use + methamphetamines  (vs. patients using opioid alone or opioid + cocaine) | AMA Discharge | OR 1.86 (1.05–3.29) | aOR 1.83 (0.99–3.41) |
| Nolan 2020^72^ | Opioid use + cocaine  (vs. patients using opioid alone or opioid + methamphetamines) | AMA Discharge | OR 1.35 (0.77–2.36) | aOR 1.38 (0.73–2.59) |
| Kimmel 2020^64^ | Alcohol | Discharge AMA  (Among patients with IDU-IE) |  | aOR 0.92 (0.73, 1.16) |
| Kimmel 2020^64^ | Cannabis | Discharge AMA  (Among patients with IDU-IE) |  | aOR 1.26 (0.96, 1.65) |
| Kimmel 2020^64^ | Sedative | Discharge AMA  (Among patients with IDU-IE) |  | aOR 1.05 (0.64, 1.71) |
| Mertz 2008^70^ | Alcohol addiction | “Patient’s non-compliance… “if the patient did not comply with diagnostic measures or adhere to therapeutic measures, left the hospital against medical advice, continued intravenous drug use during hospitalization, smoked in the room, or assaulted hospital staff.”” |  | aOR 1.2 (0.4 – 1.7) |
| Serota 2021^76^ | Opioids, stimulants, or opioids+stimulants | AMA Discharge |  | Opioid only: Ref  Opioid+stimulant: 1.28 (1.17, 1.40)  Stimulant-only: 1.09 (1.00, 1.19) |
| *Valve surgery for IE* | | | | |
| Kimmel 2020^64^ | CT Surgery | Discharge AMA  (Among patients with IDU-IE) |  | aOR 0.23 (0.16, 0.33) |
| Rudasill 2019^73^ | Surgery  (among patients with IDU-IE) | Discharge AMA | Medical treatment: 4,048/24,314  Surgical treatment: 131/3,073 p<0.001  Calculated OR  0.22 (0.19-0.27) |  |
| **Study** | **Exposures** | **Outcomes** | **Unadjusted effect estimate** | **Adjusted effect estimate** |
| *Hospital characteristics* | | | | |
| Kimmel 2020^64^ | Hospital location/teaching status | Discharge AMA  (Among patients with IDU-IE) |  | aOR  Rural: Ref  Urban, non-teaching: 1.12 (0.82 – 1.53) Urban, teaching: 0.91 (0.67, 1.24) |
| Kimmel 2020^64^ | Hospital bed size | Discharge AMA  (Among patients with IDU-IE) |  | aOR  Small: Ref  Medium: 0.96 (0.76, 1.22)  Large: 0.76 (0.61, 0.94) |
| *Hospital policy* | | | | |
| Cooksey 2020^61^ | Implementation of hospital-wide policy: search of patient’s belongings, supervised and limited visitation, restricted cell phone access, analgesics and sedatives provided only in liquid formulation. Patients’ wear self-identifying gowns, medical chart is flagged. Coerced (patient’s must agree) | Hospital discharge against medical advice  (among patients being treated for IDU-IE) | 6% pre-intervention vs. 35% post-intervention (p<0.001) |  |
| Wang 2020^81^ | Implementation of a hospital-wide policy to identify OUD and facilitate MOUD | AMA discharge  (among patients admitted to hospital with injecting-related infections) | 42.2% (out of 71 hospitalizations) prior to protocol rollout and 40.8% (out of 76 hospitalizations) after protocol (p=0.85) |  |
| Outcome: New bloodstream infection during treatment | | | | |
| Tan 2020^79^ | Age | New bloodstream infection during treatment for IDU-IE | Mean (SD)  New BSI: 34.5 (8.1)  No new BSI: 36.0 (10.0) |  |
| Tan 2020^79^ | Male | New bloodstream infection during treatment for IDU-IE | Male: 38/213  Female: 44/207  Calculated OR 1.24 (0.76-2.02) |  |
| Tan 2020^79^ | No fixed address | New bloodstream infection during treatment for IDU-IE | NFA: 20/72  Other: 62/348  Calculated OR  1.77 (0.99-3.18) |  |
| Tan 2020^79^ | Injecting opiates | New bloodstream infection during treatment for IDU-IE | Opiates: 80/365 No opiates: 2/55  Calculated OR  7.44 (1.77-31.19) |  |
| Tan 2020^79^ | Injecting stimulants | New bloodstream infection during treatment for IDU-IE | Stimulants: 55/272 No stimulants: 27/148  Calculated OR 1.14 (0.68-1.89) |  |
| Tan 2020^79^ | Injecting antidepressant | New bloodstream infection during treatment for IDU-IE | Antidepressants: 6/46 No antidepressants: 76/374  Calculated OR  0.59 (0.24-1.44) |  |
| Tan 2020^79^ | Polysubstance injection | New bloodstream infection during treatment for IDU-IE | Polysubstance: 72/321 Mono-substance: 10/99  Calculated OR  2.57 (1.27-5.21) |  |
| Tan 2020^79^ | Inpatient prescription for opiates | New bloodstream infection during treatment for IDU-IE | Rx for opiates: 82/402  No rx: 0/18  Unable to calculate OR.  With continuity correction, 0.11 (0.01-1.76) |  |
| Tan 2020^79^ | Consultation with inpatient addictions treatment | New bloodstream infection during treatment for IDU-IE | Consult: 35/156  No consult: 47/264  Calculated OR  1.34 (0.82-2.18) | aHR 0.53 (0.32 – 0.88) |
| Tan 2020^79^ | Referral to outpatient addictions treatment | New bloodstream infection during treatment for IDU-IE | Referral: 35/151  No consult: 47/269  Calculated OR 1.43 (0.87-2.33) |  |
| Tan 2020^79^ | PICC insertion | New bloodstream infection during treatment for IDU-IE |  | aHR 0.60 (0.14 – 2.56) |
| **Study** | **Exposures** | **Outcomes** | **Unadjusted effect estimate** | **Adjusted effect estimate** |
| In-hospital mortality | | | | |
| Gender/Sex |  |  |  |  |
| Martín-Dávila 2005^68^ | Sex (male) | In-hospital mortality | OR 0.90 (0.10 – 7.70) |  |
| Mertz 2008^70^ | Male gender | In-hospital mortality |  | aOR 1.0 (0.4 – 2.6) |
| Saydain 2010^75^ | Sex (female) | In-hospital mortality  (among patients with IDU-IE who were admitted to intensive care unit) | OR 0.50 (0.10 – 2.47) |  |
| Serota 2021^76^ | Biological sex | In-hospital mortality |  | aRR  Male: 1.09 (0.95, 1.27)  Female: REF |
| Age |  |  |  |  |
| Mertz 2008^70^ | Age (per 10 years older) | In-hospital mortality |  | aOR 2.0 (1.0 – 3.8) |
| Saydain 2010^75^ | Age, per year | In-hospital mortality  (among patients with IDU-IE who were admitted to intensive care unit) | OR 0.99 (0.92 – 1.07) | aOR  0.94 (0.83 – 1.07) |
| Serota 2021^76^ | Age | In-hospital mortality |  | aRR  18–34: 0.41 (.30, .56)  35–54: 0.62 (.48, .81)  55–64: 0.90 (.70, 1.15)  65–75: REF |
| Meel 2018^69^ | Age >/= 30 years | In-hospital mortality  (among patients hospitalized with IE from IV nyaope use in South Africa) | OR 3.7 (0.95 – 14.7) | aOR 4.13 (0.89 – 19.17) |
| Race |  |  |  |  |
| Saydain 2010^75^ | Race: “White/African American” | In-hospital mortality  (among patients with IDU-IE who were admitted to intensive care unit) | OR 1.33 (0.28 - 6.30)  Unclear which is ref… |  |
| Serota 2021^76^ | Race | In-hospital mortality |  | aRR  Hispanic: 1.27 (1.01, 1.61)  Non-Hispanic Black: 0.85 (.68, 1.07)  Non-Hispanic White: REF |
| *Substance use* | | | | |
| Serota 2021^76^ | Substance use | In-hospital mortality |  | aRR  Opioid+stimulant: 0.99 (.78, 1.25)  Stimulant-only: 1.26 (1.03, 1.46)  Opioid-only: Ref |
| *Overdose history* | | | | |
| Serota 2021^76^ | “Overdose” (NOS, unclear if diagnostic codes for overdose during the same hospitalization) | In-hospital mortality |  | aRR 1.26 (1.01, 1.59) |
| *Insurance* |  |  |  |  |
| Serota 2021^76^ | Insurance | In-hospital mortality |  | aRR  Medicare: 0.96 (.74, 1.25)  Medicaid: 1.41 (1.09, 1.82)  Uninsured: 0.74 (.55, .98)  Private: REF |
| Surgery in-patient |  |  |  |  |
| Martín-Dávila 2005^68^ | Surgery  (among patients with IDU-IE) | In-hospital mortality | OR 0.95 (0.92 – 0.98) |  |
| Rudasill 2019^73^ | Surgery  (among patients with IDU-IE) | In-hospital mortality | Medical treatment: 1,730/24,314  Surgical treatment: 145/3,073 p=0.007  Calculated OR  0.64 (95%CI 0.54-0.77) |  |
| Hospital policies |  |  |  |  |
| Cooksey 2020^61^ | Implementation of hospital-wide policy: search of patient’s belongings, supervised and limited visitation, restricted cell phone access, analgesics and sedatives provided only in liquid formulation. Patients’ wear self-identifying gowns, medical chart is flagged. Coerced (patient’s must agree) | In-hospital mortality rate  (among patients being treated for IDU-IE) | 11% pre-intervention vs. 0% post-intervention (p=0.003) |  |
| Other policies |  |  |  |  |
| Kimmel 2020^65^ | to investigate the effect of initiating public reporting of AVR outcomes in January 2013 on rates of valve surgery and in-hospital mortality in IDU and non–IDU-IE cases. | Inpatient mortality | “In the preintervention period, the in-hospital mortality rates were 7.9% (95% CI 6.8%–9.0%) for IDU-IE…  In the postintervention period, the proportions of hospitalizations resulting in death were 7.8% (95% CI 7.0%–8.6%) of IDU-IE…” | “In adjusted segmented regression models following backwards selection… the odds of in-hospital mortality during the preintervention period decreased by 2% per quarter for both IDU-IE and non–IDU-IE cases (AOR 0.97, 95% CI 0.97–0.9)…  Compared to projected preintervention trends, 2 years after the implementation of public reporting, the in-hospital mortality rate for IDU-IE cases changed from 3.7% (95% CI 2.2%–6.2%) to 3.2% (95% CI 1.8%–5.4%)… a decrease of 16%” |
| **Study** | **Exposures** | **Outcomes** | **Unadjusted effect estimate** | **Adjusted effect estimate** |
| Outcome: Respiratory failure in botulism | | | | |
| Sandrock 2001^74^ | Gender | Respiratory failure | Male: 13/15  Female: 2/5  “p=NS” |  |
| Sandrock 2001^74^ | Age, yr | Respiratory failure | Average (unclear if mean or median)  With RF: 47 Without RF: 40  “p=NS” | q |
| Outcome: Injecting-related infection with endogenous endophthalmitis vs. without EE | | | | |
| Uppuluri 2021^80^ | Sex | EE (vs. infection without EE) | OR Women: Ref  Men: 1.74 (1.39–2.18) | aOR 1.84 (1.44–2.34) |
| Uppuluri 2021^80^ | Age group | EE (vs. infection without EE) | OR 21-45: Ref  46-65: 1.23 (1.00–1.51) | aOR 1.12 (0.88–1.42) |
| Uppuluri 2021^80^ | Race | EE (vs. infection without EE) | OR White: Ref  Black: 1.15 (0.86–1.55)  Hispanic: 1.34 (1.00–1.81)  Asian/Pacific Islander: 4.31 (1.93–9.66)  Native American 0.27 (0.02–4.3)  Other 0.59 (0.25–1.41) | aOR White: Ref  Black: Not tested  Hispanic: Not tested  Asian/Pacific Islander: 4.41 (1.99–9.77)  Native American: Not tested  Other: Not tested |
| Uppuluri 2021^80^ | Cocaine use | EE (vs. infection without EE) | OR 0.57 (0.41–0.79) | aOR 0.80 (0.57–1.12) |
| Uppuluri 2021^1,280^ | Amphetamine use | EE (vs. infection without EE) | OR 0.34 (0.15–0.79) | aOR 0.49 (0.21–1.13) |
| Uppuluri 2021^80^ | Alcohol use disorder | EE (vs. infection without EE) | OR 0.34 (0.22–0.53) | aOR 0.35 (0.22–0.56) |
| Uppuluri 2021^80^ | Marijuana use | EE (vs. infection without EE) | OR 0.50 (0.27–0.92) | aOR 0.80 (0.43–1.48) |
| Uppuluri 2021^80^ | Tobacco use | EE (vs. infection without EE) | OR 0.82 (0.66–1.01) | aOR Not tested |
| Uppuluri 2021^80^ | Infection of central venous line | EE (vs. infection without EE) | OR 4.46 (2.67–7.44) | aOR 1.90 (1.09–3.29) |

### Appendix 14. List of exposure-outcome pair effect estimates for studies where outcome occurs after treatment of injecting-related bacterial infections drug use-associated bacterial and fungal infections, included in quantitative systematic review. Blank cells represent where effect estimate (or frequencies) were not reported in the paper.

| **Study** | **Exposures** | **Outcomes** | **Unadjusted effect estimate** | **Adjusted effect estimate** |  |
| --- | --- | --- | --- | --- | --- |
| **Infection-related rehospitalization** | | | | | |
| *Age* |  |  |  |  |  |
| Barocas 2020^82^ | Age (years)  (after hospitalization for injecting-related SSTI) | 1-year rehospitalization for SSTI |  | aHR 1.01 (1.01 – 1.01) |  |
| *Huang 2018*^90^ | Age, median (IQR) | Repeat episode of endocarditis (comparing features during first episode among people with single episode vs. repeat episode) | Single episode: 29 (24-38.5) Repeat episode: 28.5 (23-37.3) p=0.63 |  |  |
| *Thønnings 2020*^52^ | Age, years | Recurrent bacteraemia after initial bacteremia (vs. no recurrence) | OR 0.93 (0.86–1.01) |  |  |
| **Study** | **Exposures** | **Outcomes** | **Unadjusted effect estimate** | **Adjusted effect estimate** |  |
| *Gender/sex* |  |  |  |  |  |
| Barocas 2020^82^ | Sex (female)  (after hospitalization for injecting-related SSTI) | 1-year rehospitalization for SSTI |  | aHR 1.10 (0.96 – 1.25) |  |
| Huang 2018^90^ | Male | Repeat episode of endocarditis (comparing features during first episode among people with single episode vs. repeat episode) | Single episode: 32 (49% male)  Repeat episode: 11 (50% male) p=0.95 |  |  |
| Pericàs 2021^94^ | Male  (among PWID with IE) | 6-month “relapse”  (Readmission with same microbiology) | OR  Female: Ref Male: 0.83 (0.37–1.88) |  |  |
| *Race* |  |  |  |  |  |
| Huang 2018^90^ | Caucasian race | Repeat episode of endocarditis (comparing features during first episode among people with single episode vs. repeat episode) | Single episode: 62 (95.4%)  Repeat episode: 22 (100%) p=0.57 |  |  |
| **Study** | **Exposures** | **Outcomes** | **Unadjusted effect estimate** | **Adjusted effect estimate** |  |
| Rural/urban |  |  |  |  |  |
| Huang 2018^90^ | Nonmetro residency | Repeat episode of endocarditis (comparing features during first episode among people with single episode vs. repeat episode) | Single episode: 46 (71%)  Repeat episode: 15 (68%) p=0.82 |  |  |
| Substances injected |  |  |  |  |  |
| Huang 2018^90^ | Prescription opioid injecting | Repeat episode of endocarditis (comparing features during first episode among people with single episode vs. repeat episode) | Single episode: 44 (68%)  Repeat episode: 21 (95.4%%) p=0.01 |  |  |
| *Opioid agonist treatment or naltrexone* |  |  |  |  |  |
| Barocas 2020^82^ | Prescription for naltrexone or buprenorphine within 30 days of hospital discharge (after hospitalization for injecting-related SSTI) | 30-day rehospitalization for SSTI | No MOUD group: 2.8 per 100 person-months (95% CI, 2.73-2.81)  MOUD group: 1.13 per 100 person-months (95% CI, 1.02-1.24) | aHR = 0.49, 95% CI 0.18-1.23 |  |
| Barocas 2020^82^ | Prescription for naltrexone or buprenorphine within 30 days of hospital discharge (after hospitalization for injecting-related SSTI) | 1-year rehospitalization for SSTI | No MOUD group: 18.7 per 100 person-years (95% CI, 18.53-18.78)  MOUD group: 10.3 per 100 person-years (95% CI, 9.87-10.64) | aHR 0.41 (0.42 – 0.91) |  |
| Hilbig 2020^89^ | Methadone vs. no methadone  (among patient’s hospitalized with first episode IDU-IE) | “Recurrence” | “No patients who were receiving methadone therapy on admission had a recurrence, compared with 25% of patients not receiving methadone therapy, but this difference was not statistically significant (p = 0.06).” |  |  |
| Hilbig 2020^89^ | Methadone vs. no methadone  (among patient’s hospitalized with first OR second episode IDU-IE) | “Recurrence” | “When analysis was extended to all primary and secondary episodes, there were some differences… The association with methadone therapy was no longer significant.” |  |  |
| Suzuki 2020^101^ | MOUD | Repeat episode of endocarditis  (among PWID with IE) | Buprenorphine: 4/8  Methadone taper and referral: 2/8  Declined MOUD: 4/10  P=NS |  |  |
| *Thønnings 2020*^52^ | Opioid substitution treatment | Recurrent bacteraemia after initial bacteremia (vs. no recurrence) | OR 5.88 (0.64–50.00) |  |  |
| *Other substance use treatment* | | | | | |
| *Thønnings 2020*^52^ | Contact to an addiction treatment center (not otherwise defined) | Recurrent bacteraemia after initial bacteremia (vs. no recurrence) | OR 1.39 (0.45–4.17) |  |  |
| Rodger 2019^97^ | Referral to addiction treatment | Recurrent endocarditis | OR 0.63; 0.32–1.24 | aOR 0.54; 0.26–1.14 |  |
| *Cardiac surgery during admission* | | | | | |
| Pericàs 2021^94^ | Cardiac surgery during first admission | 6-month “relapse”  (Readmission with same microbiology) | OR 1.73 (0.79–3.78) | aOR 1.96 (0.88–4.37) |  |
| Rodger 2019^97^ | Surgery during first admission | Recurrent endocarditis | OR 1.06; 0.50–2.22 | aOR 0.90; 0.38–2.14 |  |
| *AMA discharge* |  |  |  |  |  |
| Rodger 2019^97^ | AMA discharge during first admission | Recurrent endocarditis | OR 0.56; 0.24–1.28 | aOR 0.50; 0.21–1.20 |  |
| *Hospital factors/policy* |  |  |  |  |  |
| Ray 2020^95^ | Hospital initiative to improve pain and OUD management for patients with IDU-IE, including pain and addiction medicine consultation and new care pathway | 90-day endocarditis readmission | Preintervention: 8/37 (22%)  Postintervention: 9/33 (27%) p=0.58 |  |  |
| **Study** | **Exposures** | **Outcomes** | **Unadjusted effect estimate** | **Adjusted effect estimate** |  |
| All-cause rehospitalization | | | | | |
| **Study** | **Exposures** | **Outcomes** | **Unadjusted effect estimate** | **Adjusted effect estimate** |  |
| *Age* |  |  |  |  |  |
| Barocas 2020^82^ | Age (years)  (after hospitalization for injecting-related SSTI) | 30-day all-cause rehospitalization |  | aHR 1.0 (0.99 – 1.00) |  |
| Jo 2021^63^ | Age (?continuous)  (among patients with untreated OUD and either endocarditis or osteomyelitis) | 30-day all-cause rehospitalization |  | aOR 1.00 (0.99, 1.01) |  |
| Barocas 2021^83^ | Age (years)  (after hospitalization for injecting-related endocarditis) | 1-year all-cause rehospitalization |  | aHR 1.01 (1.00 – 1.01) |  |
| Marks 2020^92^ | Age (>50 years)  (after hospitalization with invasive infections: Endocarditis, septic arthritis, bacteremia, osteomyelitis, epidural abscess) | 90-day all-cause readmission | OR 1.09 (0.59 – 2.02) |  |  |
| *Sex/Gender* |  |  |  |  |  |
| Barocas 2020^82^ | Sex (female)  (after hospitalization for injecting-related SSTI) | 30-day all-cause rehospitalization |  | aHR 1.25 (1.13 – 1.39) |  |
| Jo 2021^63^ | Biological sex  (among patients with untreated OUD and either endocarditis or osteomyelitis) | 30-day all-cause rehospitalization |  | aOR Male: 0.89 (0.71 – 1.11)  Female: Ref |  |
| Barocas 2021^83^ | Sex (female)  (after hospitalization for injecting-related endocarditis) | 1-year all-cause rehospitalization |  | aHR 1.18 (0.98 – 1.43) |  |
| Marks 2020^92^ | Sex (female)  (after hospitalization with invasive infections) | 90-day all-cause readmission | OR 1.23 (0.77 – 1.96) |  |  |
| *Race/Ethnicity* |  |  |  |  |  |
| Jo 2021^63^ | Ethnicity  (among patients with untreated OUD and either endocarditis or osteomyelitis) | 30-day all-cause rehospitalization |  | aOR White: 0.69 (0.51 – 0.93)  Other: Ref |  |
| Marks 2020^92^ | African American (extracted from medical chart review)  (after hospitalization with invasive infections) | 90-day all-cause readmission | OR 0.79 (0.49 – 1.27) |  |  |
| *Unstable housing* |  |  |  |  |  |
| Marks 2020^92^ | Homeless (extracted from medical chart review)  (after hospitalization with invasive infections) | 90-day all-cause readmission | OR 0.92 (0.46 – 1.83) |  |  |
| *Insurance* |  |  |  |  |  |
| Cooksey 2020^61^ | Lacking insurance on hospital admission for IDU-IE | 90-day all-cause rehospitalization  (after hospitalization for IDU-IE) | OR 1.3 (0.6 – 2.6) |  |  |
| Jo 2021^63^ | Insurance status  (among patients with untreated OUD and either endocarditis or osteomyelitis) | 30-day all-cause rehospitalization |  | aOR Govt funded insurance: 1.18 (0.90, 1.56)  Commercial insurance: 1.08 (0.73 – 1.61)  Uninsured: Ref |  |
| *Substances injected - heroin* | | | | | |
| Marks 2020^92^ | Heroin or fentanyl use  (after hospitalization with invasive infections) | 90-day all-cause readmission | OR 0.69 (0.32 – 1.46) |  |  |
| Jo 2021^63^ | Cocaine/amphetamine use  (among patients with untreated OUD and either endocarditis or osteomyelitis) | 30-day all-cause rehospitalization |  | aOR Yes: 0.86 (0.62 0 1.18)  No: Ref |  |
| *Cocaine use* |  |  |  |  |  |
| Marks 2020^92^ | Cocaine use  (after hospitalization with invasive infections) | 90-day all-cause readmission | OR 1.53 (0.90 – 2.60) |  |  |
| *Methamphetamine* | | | | | |
| Marks 2020^92^ | Methamphetamine use  (after hospitalization with invasive infections) | 90-day all-cause readmission | OR 1.32 (0.78 – 2.25) |  |  |
| *Other substance use* | | | | | |
| Barocas 2020^82^ | Other substance use disorder (ICD codes related to alcohol, cannabis, hallucinogens, or sedatives from hospital admin data)  (after hospitalization for injecting-related SSTI) | 30-day all-cause rehospitalization |  | aHR 1.29 (1.11 – 1.50) |  |
| Marks 2020^92^ | Benzodiazepine use  (after hospitalization with invasive infections) | 90-day all-cause readmission | OR 0.49 (0.09 – 2.45) |  |  |
| *Addiction medicine consultation* | | | | | |
| Marks 2020^92^ | Inpatient addiction medicine consultation  (after hospitalization with invasive infections) | 90-day all-cause readmission | OR 0.39 (0.24 – 0.64)  HR 0.49 (0.34 – 0.72) | aHR 0.57 (0.38 – 0.86) |  |
| Marks 2019^66^ | Inpatient addiction medicine consultation  (after hospitalization with invasive infections) | 90-day all-cause readmission | HR 0.378 (0.21 – 0.69) |  |  |
| *OAT + naltrexone* |  |  |  |  |  |
| Barocas 2020^82^ | Prescription for naltrexone or buprenorphine within 30 days of hospital discharge (after hospitalization for injecting-related SSTI) | 30-day all-cause rehospitalization | no MOUD group: 27.5 per 100 person-months (95% CI 27.4-27.7)  MOUD group: 35.9 (95% CI 35.3-36.6) | aHR 1.29 (1.05-1.59) |  |
| Barocas 2020^82^ | Prescription for naltrexone or buprenorphine within 30 days of hospital discharge (after hospitalization for injecting-related SSTI) | 1-year all-cause rehospitalization | no MOUD group: 192.9 per 100 person-years (95% CI, 192.5-193.3)  MOUD group: 169.4 per 100 person-years (95% CI, 167.9-171.0) | aHR = 0.96, 95% CI 0.83-1.11 |  |
| Barocas 2021^83^ | Prescription for naltrexone or buprenorphine within 30 days of hospital discharge (after hospitalization for injecting-related endocarditis | 1-year all-cause rehospitalization |  | aHR 0.81 (0.51 – 1.28) |  |
| Jo 2021^63^ | OAT initiation  (among patients with untreated OUD admitted with endocarditis or osteomyelitis) | 30 days all-cause readmission | MOUD: 106/269 No MOUD: 421/1138  P value not tested | aOR 1.14 (0.87 – 1.50) |  |
| Wang 2020^81^ | MOUD in hospital AND continued on discharge (vs. no MOUD on discharge) | 30-day all-cause readmission  (among patients admitted to hospital with injecting-related infections) | RR 0.54; 95% CI, 0.32-0.96 |  |  |
| Wang 2020^81^ | MOUD in hospital AND continued on discharge (vs. no MOUD on discharge) | 90-day all-cause readmission  (among patients admitted to hospital with injecting-related infections) | RR 0.64; 95% CI, 0.40-1.03; |  |  |
| **Study** | **Exposures** | **Outcomes** | **Unadjusted effect estimate** | **Adjusted effect estimate** |  |
| *Hospital policy* |  |  |  |  |  |
| Cooksey 2020^61^ | Implementation of hospital-wide policy: search of patient’s belongings, supervised and limited visitation, restricted cell phone access, analgesics and sedatives provided only in liquid formulation. Patients’ wear self-identifying gowns, medical chart is flagged. Coerced (patient’s must agree) | 90-day all-cause rehospitalization  (after hospitalization for IDU-IE) | 48% pre-intervention vs. 34% post-intervention (p=0.068) | aOR 0.2 (95% CI 0.08-0.6) |  |
| Cooksey 2020^61^ | Implementation of hospital-wide policy: search of patient’s belongings, supervised and limited visitation, restricted cell phone access, analgesics and sedatives provided only in liquid formulation. Patients’ wear self-identifying gowns, medical chart is flagged. Coerced (patient’s must agree) | 12-month all-cause rehospitalization  (after hospitalization for IDU-IE) | 70% pre-intervention vs. 38% post-intervention (p<0.001) |  |  |
| Ray 2020^95^ | Hospital initiative to improve pain and OUD management for patients with IDU-IE, including pain and addiction medicine consultation and new care pathway | 90-day all-cause readmission | Preintervention: 16/37 (43%)  Postintervention: 13/33 (39%) p=0.74 |  |  |
| Wang 2020^81^ | Implementation of a hospital-wide policy to identify OUD and facilitate MOUD | 30-day all-cause readmission  (among patients admitted to hospital with injecting-related infections) | The rate of 30-day all-cause readmissions was 29.6% prior to protocol rollout and 25.3% afterward (RR 0.86; 95% CI, 0.51-1.45; P = .56) |  |  |
| Wang 2020^81^ | Implementation of a hospital-wide policy to identify OUD and facilitate MOUD | 90-day all-cause readmission  (among patients admitted to hospital with injecting-related infections) | The rate of 90-day all-cause readmissions was 38.0% prior to protocol rollout and 32.9% afterward (RR 0.86; 95% CI, 0.55-1.35; P = .52) |  |  |
| **Study** | **Exposures** | **Outcomes** | **Unadjusted effect estimate** | **Adjusted effect estimate** |  |
| *Antibiotic treatment mode* | | | | | |
| Marks 2020^92^ | Antibiotic treatment mode  (after hospitalization with invasive infections) | 90-day all-cause readmission | Completed inpatient IV antibiotics: Ref Partial IV, partial oral: OR 1.05 (0.58 – 1.83)  Partial IV, no oral: OR 4.77 (2.55 – 8.92)  Partial IV, partial OR: HR 0.92 (0.57 – 1.48)  Partial IV, no oral: HR 3.17 (0.197 – 5.12) | Partial IV, partial oral: aHR 0.99 (0.62 – 1.62)  Partial IV, no oral: aHR 2.32 (1.41 – 3.82) |  |
| *Surgery* |  |  |  |  |  |
| Marks 2020^92^ | Received surgical procedure  (after hospitalization with invasive infections) | 90-day all-cause readmission | OR 0.38 (0.23 – 0.62)  HR 0.44 (0.29 – 0.66) | aHR 0.57 (0.37 – 0.87) |  |
| Rudasill 2019^73^ | Surgery  (among patients with IDU-IE) | Readmission within 30 days | Medical treatment: 5,507/24,314 (22.6%)  Surgical treatment: 584/3,073 (19%) p=0.007 |  |  |
| Rudasill 2019^73^ | Surgery  (among patients with IDU-IE) | Readmission from 30 to 180 days | Medical treatment: 2,681/24,314  Surgical treatment: 405/3,073 p=0.044 |  |  |
| Slaughter 2019^99^ | Type of cardiac surgery in tricuspid valve endocarditis | 30 day readmission | Valvectomy: 10/119 (9.9%) Repair: 31/532 (5.9%)  Replacement: 84/962 (8.9%) p=0.34 | aOR Repair: Reference Replacement: 1.04 (0.47 – 2.27)  Valvectomy: 5.42 (2.33 – 12.57) |  |
| **Study** | **Exposures** | **Outcomes** | **Unadjusted effect estimate** | **Adjusted effect estimate** |  |
| **Overdose Hospitalisation** | | | | | |
| Barocas 2021^83^ | Age (years)  (after hospitalization for injecting-related endocarditis) | Opioid-related overdose hospitalization |  | aHR 0.97 (0.95 – 0.99) |  |
| Barocas 2021^83^ | Sex (female)  (after hospitalization for injecting-related endocarditis) | Opioid-related overdose hospitalization |  | aHR 0.89 (0.47 – 1.63) |  |
| Barocas 2021^83^ | Other substance use disorder (ICD codes related to alcohol, cannabis, hallucinogens, or sedatives from hospital admin data)  (after hospitalization for injecting-related endocarditis) | Opioid-related overdose hospitalization |  | aHR 0.86 (0.26 – 2.84) |  |
| Barocas 2021^83^ | Prescription for naltrexone or buprenorphine within 30 days of hospital discharge (after hospitalization for injecting-related endocarditis | Opioid-related overdose |  | aHR 0.86 (0.26 – 2.91) |  |
| Wang 2020^81^ | MOUD in hospital and continued on discharge (vs. no MOUD on discharge) | 30-day opioid-related readmission  (among patients admitted to hospital with injecting-related infections) | The respective 30-day opioid-related readmission rate was 10.1% vs 29.9% (RR 0.34; 95% CI, 0.16-0.74; P = .003). |  |  |
| Wang 2020^81^ | MOUD in hospital and continued on discharge (vs. no MOUD on discharge) | 90-day opioid-related readmission  (among patients admitted to hospital with injecting-related infections) | The respective 90-day opioid-related readmission rate was 15.2% vs 33.3% (RR 0.46; 95% CI, 0.24-0.88; P = .01). |  |  |
| Wang 2020^81^ | Implementation of a hospital-wide policy to identify OUD and facilitate MOUD | 30-day opioid-related readmission  (among patients admitted to hospital with injecting-related infections) | The rate of 30-day opioid-related readmissions was 22.5% prior to protocol rollout and 18.7% afterward (RR 0.83; 95% CI, 0.437-1.57, P = .56). |  |  |
| Wang 2020^81^ | Implementation of a hospital-wide policy to identify OUD and facilitate MOUD | 90-day opioid-related readmission  (among patients admitted to hospital with injecting-related infections) | The rate of 90- day opioid-related readmissions was 26.7% prior to protocol rollout and 22.9% afterward (RR 0.95; 95% CI, 0.48-1.52; P = .59). |  |  |
| All-cause mortality |  |  |  |  |  |
| *Sex/gender* |  |  |  |  |  |
| Clarelin 2021^85^ | Sex (female) | Mortality, survival analysis  After hospitalization with IDU-IE | HR 0.72 (0.50 – 1.03) | aHR 1.05 (0.71 – 1.56) |  |
| Kimmel 2020^91^ | Female | Mortality, survival analysis  After hospitalization with IDU-IE |  | aHR 0.80 (0.45-1.43) |  |
| Pericàs 2021^94^ | Male | 6-month mortality  (among PWID with IE) | OR  Female: Ref Male: 1.92 (1.06–3.46) | aOR 1.75 (0.89–3.48) |  |
| Rodger 2018^96^ | Sex | All-cause mortality (time undefined) | Unclear numerators and denominators and no reference category for relative risk statistics? RR estimates for both categories… “p=0.12” |  |  |
| Rohn 2020^98^ | Gender | 30-day mortality | Female: 2/28  Male: 4/34 p=>0.999 |  |  |
| Straw 2020^100^ | Sex (male vs. female) | Survival |  | aHR 0.67 (0.30 – 1.5) |  |
| Weymann 2014^102^ | Female  (among patients with IDU-IE undergoing surgery) | 90-day mortality | Female: 1/7  Male: 1/13 p=0.639 |  |  |
| *Age* |  |  |  |  |  |
| Clarelin 2021^85^ | Age (per year) | Mortality, survival analysis  After hospitalization with IDU-IE | OR 1.05 (1.04-1.07) | aOR 1.03 (1.01 – 1.05) |  |
| Kimmel 2020^91^ | Age | Mortality, survival analysis  After hospitalization with IDU-IE |  | aHR  18-34: 0.26 (0.12-0.55)  35-49: 0.62 (0.35 – 1.10)  50-64: Ref |  |
| Rodger 2018^96^ | Age | All-cause mortality | Only presents age median (IQR) for those who died, but “p=0.04” |  |  |
| Rohn 2020^98^ | Age | 30-day mortality | Mean (SD),  Survivors: 29 (5.97)  Non-survivors: 33.3 (26.5)  P=0.0525 |  |  |
| Slaughter 2019^99^ | Age, per year | “Operative mortality” which was death in hospital OR within 30 days of discharge |  | aOR 1.01 (0.98 – 1.04) |  |
| Straw 2020^100^ | Age, per year | Survival |  | aHR 1.1 (1.0-1.1) |  |
| Weymann 2014^102^ | Age, years  (among patients with IDU-IE undergoing surgery) | 90-day mortality | Survivors: 35.1 (7.7)  Non-survivors: 34.5 (10.6)  P=0.926  (I think mean, SD, but doesn’t say?) |  |  |
| *Unstable housing* |  |  |  |  |  |
| Kimmel 2020^91^ | Homelessness | Mortality, survival analysis  After hospitalization with IDU-IE |  | aHR 0.60 (0.31 – 1.14) |  |
| Specific substances |  |  |  |  |  |
| Rodger 2018^96^ | Opioid (alone) | All-cause mortality | RR 1.72 (1.06 – 2.80)  Unclear reference category -- is it everyone else? |  |  |
| Rodger 2018^96^ | Stimulant (alone) | All-cause mortality | RR 0.79 0.30 – 2.11)  Unclear reference category -- is it everyone else? |  |  |
| Rodger 2018^96^ | Polysubstance use | All-cause mortality | RR 0.81 (0.62 – 1.06)  Unclear reference category -- is it everyone else? |  |  |
| *Against medical advice AMA discharge* | | | | | |
| Rodger 2018^96^ | Left against medical advice | All-cause mortality | RR 0.34 (0.14 – 0.84)  HR 0.34 (0.14 – 0.85)  But outcome included many (most?) people who died during hospitalization (e.g. sepsis, etc.) so really this is just an indicator that they survived the hospital admission… | aHR 0.47 (0.18 – 1.19)  (adjusted for age and sex) |  |
| *Opioid agonist treatment at/after discharge* | | | | | |
| Kimmel 2020^91^ | Receipt of MOUD (buprenorphine, methadone, naltrexone) | Mortality, survival analysis  After hospitalization with IDU-IE |  | aHR 0.30; 95% CI, 0.10-0.89 |  |
| Marks 2020^67^ | MOUD prescription at discharge | All-cause mortality at 1 year | RR 0.26 (0.09 – 0.75)  (Cannot find frequencies to generate 2X2 table, but 15% of sample died) |  |  |
| Rodger 2018^96^ | Opioid substitution treatment prescription at discharge | All-cause mortality  (among PWID with IE) | RR 0.34 (0.43 – 1.33)  Unclear if it includes OST received during hospitalization or only at discharge. Outcome seems to include in-hospital deaths, so may just be an indicator of who survived to discharge |  |  |
| Suzuki 2020^101^ | MOUD given in hospital | Mortality  (among PWID with IE) | Buprenorphine: 0/8  Methadone taper and referral: 3/8  Declined MOUD: 1/10  P=NS |  |  |
| *Addiction medicine consultation/referral* | | | | | |
| Nguemeni Tiako 2020^93^ | “Comprehensive addiction treatment” as inpatient | 24-month survival | Yes addiction treatment: 0/20 died  No/partial addiction treatment 7/22 died | Not tested |  |
| Rodger 2018^96^ | Referral to addiction treatment (on discharge) | All-cause mortality  (among PWID with IE) | RR 0.28 (0.12 – 0.69)  HR 0.28 (0.12 – 0.69)  Outcome seems to have included in-hospital deaths, so this is likely just an indicator of who survived to hospital discharge… | aHR 0.29 (0.12 – 0.73)  (adjusted for age and sex) |  |
| Tan 2020^79^ | Consultation with inpatient addictions treatment | 90 day all-cause mortality |  | aHR 0.64 (0.32 – 1.29) |  |
| *Hospital policy* |  |  |  |  |  |
| Cooksey 2020^61^ | Implementation of hospital-wide policy: search of patient’s belongings, supervised and limited visitation, restricted cell phone access, analgesics and sedatives provided only in liquid formulation. Patients’ wear self-identifying gowns, medical chart is flagged. Coerced (patient’s must agree) | 12-month all-cause mortality  (after hospitalization for IDU-IE) | 7% pre-intervention vs. 4% post-intervention (p=0.73) | aOR 0.25 (95% CI 0.07 – 0.89) |  |
| Wang 2020^81^ | Implementation of a hospital-wide policy to identify OUD and facilitate MOUD | Mortality in 3 months after discharge  (among patients admitted to hospital with injecting-related infections) | In the pre-protocol cohort, 2 of 71 patients died within 3 months. In the post-protocol cohort, 3 of 76 patients died within 3 months. The overall 3-month mortality rate was 3.4%. These numbers were too small to compare | Not tested |  |
| Surgery during hospitalization | | | | | |
| Pericàs 2021^94^ | Cardiac surgery during initial admissions  (among PWID with IE) | 6-month mortality | OR  No: Ref Yes: 0.57 (0.34–0.95) | aOR 0.32 (0.16–0.61) |  |
| Rodger 2018^96^ | Cardiac surgery | All-cause mortality | Unclear numerators and denominators and no reference category for relative risk statistics? RR estimates for both categories… “p>0.99”  HR 0.89 (0.49 – 1.64) | aHR 0.44 (0.23 – 0.84)  (adjusted for age and sex) |  |
| Slaughter 2019^99^ | Type of cardiac surgery in tricuspid valve endocarditis | “Operative mortality” which was death in hospital OR within 30 days of discharge | Valvectomy: 19/119 (16%) Repair: 12/532 (2.3%)  Replacement: 29/962 (3.0%) p<0.01 | aOR Repair: Reference Replacement: 1.04 (0.47 – 2.27)  Valvectomy: 5.42 (2.33 – 12.57) |  |
| Straw 2020^100^ | Surgery | Survival | HR 1.8 (0.95 – 3.3) | aHR 17 (0.75 – 3.8) |  |
| Tan 2020^79^ | Cardiac surgery | New bloodstream infection during treatment for IDU-IE |  | aHR 0.66 (0.27 – 1.61) |  |
| **Study** | **Exposures** | **Outcomes** | **Unadjusted effect estimate** | **Adjusted effect estimate** | **Notes** |
| Other post-hospital/outpatient outcomes | | | | | |
| Buehrle 2017^84^ | Age (years)  (while on OPAT after hospitalization for injecting-related infection) | OPAT failure as any of the following: worsening or ongoing infection  requiring hospital readmission within 30 days, worsening or  ongoing infection resulting in prolonged antibiotic therapy,  antibiotic noncompliance, noncompliance with follow-up clinic  appointments, or death during treatment course. | Median age in years (range)  Success: 34 (25–62)  Failure: 35 (19–63)  P=0.82 |  |  |
| Connell 2010^86^ | Male vs. female | Change in visual acuity after treatment for endogenous Candida endophthalmitis | Among males: mean visual acuity pre-treatment 6/48 vs. post-treatment 6/24 (p=0.04)  Among females: mean visual acuity pre-treatment 6/60 vs. post-treatment 6/48 (p=0.58) |  |  |
| Connell 2010^86^ | Age <= 35 years vs. >35 years | Change in visual acuity after treatment for endogenous Candida endophthalmitis | Among age <= 35: mean visual acuity pre-treatment 6/48 vs. post-treatment 6/36 (p=0.29)  Among age >5: mean visual acuity pre-treatment 6/60 vs. post-treatment 6/36 (p=0.45) |  |  |
| **Study** | **Exposures** | **Outcomes** | **Unadjusted effect estimate** | **Adjusted effect estimate** | **Notes** |
| *Discharge location* |  |  |  |  |  |
| D’Couto 2018^87^ | Discharge home vs. to skilled nursing facility  (while on OPAT, after hospitalization for injecting-related infeciton | Any complication (including line complications, IDU relapse, loss to follow-up, death) | Discharged home: 4/21  Discharged to SNF: 11/31  P = 0.23 |  |  |
| Fanucchi 2020^88^ | Discharge home (OPAT) vs. remaining in hospital [pilot randomized trial]  (once hospitalized for injecting-related infection) | Antibiotic completion | All participants (100%) completed the recommended course of IV antibiotic therapy.  OPAT participants completed 20.1 (SD ± 11.1) days of outpatient IV antibiotics compared 1.8 (SD ± 5.3) days for UC participants (t(17) = –4.5, P < .001). |  |  |
| Buehrle 2017^84^ | Discharged from hospital to residential treatment facility  (while on OPAT after hospitalization for injecting-related infection) | OPAT  failure as any of the following: worsening or ongoing infection  requiring hospital readmission within 30 days, worsening or  ongoing infection resulting in prolonged antibiotic therapy,  antibiotic noncompliance, noncompliance with follow-up clinic  appointments, or death during treatment course. | Success: 1 (4%) out of 26  Failure: 0 (0%) out of 41  P = NA |  |  |

### Appendix 15. List of exposure-outcome pair effect estimates for studies where outcome is colonization with pathogenic bacteria among people who inject drugs, included in quantitative systematic review. Blank cells represent where effect estimate (or frequencies) were not reported in the paper.

| **Study** | **Exposures** | **Outcomes** | **Unadjusted effect estimate** | **Adjusted effect estimate** |
| --- | --- | --- | --- | --- |
| **Sociodemogaphic factors** | | | | |
| *Gender/Sex* |  |  |  |  |
| Colombo 2012^103^ | Sex (not defined) | MRSA-positive nose/throat or wound swabs (vs. MRSA-negative) | “no statistical differences” |  |
| Leung 2015^105^ | Sex | *S. aureus* nasal colonization *(76% MSSA, 24% MRSA)* | Male: 78/347 (22%)  Female: 27/ 90 (30%)  P value not reported | Male: Ref  Female: 1.07 (0.98 – 1.18) |
| Miller 2007^106^ | Sex | *S. aureus* nasal colonization *(14% MRSA)* | Male: 36/166  Female: 29/116  P=0.52 |  |
| *Age* |  |  |  |  |
| Colombo 2012^103^ | Age (not defined) | MRSA-positive nose/throat or wound swabs (vs. MRSA-negative) | “no statistical differences” |  |
| Leung 2015^105^ | Age (mean, SD) | *S. aureus* nasal colonization | Colonized: 43.7, 12.6 Not colonized: 47.6, 11.1  “p<0.05” | aOR 0.99 (0.99-1.00) |
| Miller 2007^106^ | Age | *S. aureus* nasal colonization *(14% MRSA)* | <=30 years: 2/20  21-45 years: 44/188  >45 years: 19/74 p=0.3 |  |
| *Race/Ethnicity* |  |  |  |  |
| Leung 2015^105^ | Race | *S. aureus* nasal colonization | Hispanic/Latino: 18/72  White: 26/90  Black: 59/258  Other: 3/20  P value not reported |  |
| Miller 2007^106^ | Race/ethnicity | *S. aureus* nasal colonization *(14% MRSA)* | Hispanic: 43/200  Black: 11/53  Other: 11/29  P=0.13 |  |
| *Education* |  |  |  |  |
| Leung 2015^105^ | Education | *S. aureus* nasal colonization | Grades 1-11: 36/139  Grade 12 or GED: 44/185  Some college or more: 26/116  P value not reported |  |
| *Employment* |  |  |  |  |
| Leung 2015^105^ | Employment | *S. aureus* nasal colonization | Employed: 24/104  Unemployed: 82/318  “Not currently in the workforce”: 0/18  P<0.05 | Employed: 0.96 (0.88 – 1.06)  Unemployed: Ref  “Not currently in the workforce”: 0.78 (0.64 – 0.95) |
| Miller 2007^106^ | Employed | *S. aureus* nasal colonization *(14% MRSA)* | Employed: 15/73  Unemployed: 50/209  P=0.56 |  |
| *Marital status* |  |  |  |  |
| Leung 2015^105^ | Marital status | *S. aureus* nasal colonization | Married/common-law: 16/54  Separated: 11/54  Divorced: 27/122  Widowed: 2/23  Never married: 50/187  P value not reported | Married/common-law: 1.04 (0.99 – 1.00)  *double-checked this one and must be a typo  Separated: 0.94 (0.83 – 1.07)  Divorced: 0.96 (0.87 – 1.05)  Widowed: 0.83 (0.70 – 1.00)  Never married: Ref |
| *Unstable housing* | |  |  |  |
| Leung 2015^105^ | History of homelessness | *S. aureus* nasal colonization | Yes homelessness: 63 colonized out of 278 homelessness  No homelessness: 43 colonized out of 162 not homeless  P value not reported  OR 0.81 (0.52-1.27) |  |
| Leibler 2019^104^ | Sleeping in a homeless shelter in the last 3 months | MRSA-positive nasal swab | OR 3.0 (1.2, 7.6) |  |
| Leibler 2019^104^ | Sleeping on the street | MRSA-positive nasal swab | “No associations were observed…” |  |
| Leibler 2019^104^ | Sleeping at >1 place during the last week | MRSA-positive nasal swab | OR 3.1 (1.3, 7.6) |  |
| Leibler 2019^104^ | Use of public shower facilities in the last week  (“Public restrooms are exclusive of restrooms in homeless shelters, day centers, or hospitals”) | MRSA-positive nasal swab | OR 13.7 (1.4, 132.8) |  |
| Leibler 2019^104^ | Sharing bedding with other people | MRSA-positive nasal swab | OR 2.2 (1.0 – 4.7) |  |
| Miller 2007^106^ | Homeless during the past six months | *S. aureus* nasal colonization *(14% MRSA)* | Yes: 5/13  No: 60/269 p=0.19 |  |
| Miller 2007^106^ | Spent time in a shelter during the past 6 months | *S. aureus* nasal colonization *(14% MRSA)* | Yes: 2/65  No: 63/275  P=0.66 |  |
| Packer 2019^107^ | Homeless past year | *MRSA* colonization | OR 3.2 (0.94 – 10.96) |  |
| *Public injecting* |  |  |  |  |
| Packer 2019^107^ | Most frequent injecting location | *MRSA* colonization | ORs House own/friend: Ref  Hostel, squat, other: 1.7 (0.41 – 7.16)  Public places: 5.5 (1.34 – 22.73) |  |
| *Inject in groups* |  |  |  |  |
| Packer 2019^107^ | Frequently inject in groups | *MRSA* colonization | ORs Own: Ref  Less than three people: 1.5 (0.4 – 5.3)  Three or more people: 15.8 (2.51 – 99.28) |  |
| *Incarceration* | |  |  |  |
| Leung 2015^105^ | Incarceration, past 12 months | *S. aureus* nasal colonization | Yes incarcerated: 37 colonized out of 165 incarceration  Not incarcerated: 69 colonized out of 275 not incarcerated  P value not reported.  OR 0.86 (0.55-1.36) |  |
| Miller 2007^106^ | Spent time in prison during the past 6 months | *S. aureus* nasal colonization *(14% MRSA)* | Yes: 9/36  No: 56/246 p=0.77 |  |
| *Recent hospitalization* |  |  |  |  |
| Leung 2015^105^ | Hospitalized, past 6 months | *S. aureus* nasal colonization | Yes hospitalized: 20 colonized out of 79 hospitalized  Not hospitalized: 86 colonized out of 361 not hospitalized  P value not reported  OR 1.08 (0.62-1.90) |  |
| Miller 2007^106^ | Hospitalized during the past 6 months | *S. aureus* nasal colonization *(14% MRSA)* | Yes: 6/39  No: 59/243  P=0.22 |  |
| Packer 2019^107^ | Hospital contact past month | *MRSA* colonization | OR 4.3 (1.34 – 13.8) |  |
| *Drug treatment* |  |  |  |  |
| Leung 2015^105^ | Drug treatment program, past 12 months | *S. aureus* nasal colonization | Yes drug treatment: 25 colonized out of 82 in drug treatment  No drug treatment: 81 colonized out of 358 not in drug treatment  P value not reported | aOR 1.08 (0.97 – 1.19) |
| Miller 2007^106^ | Enrolled in a methadone program  (but not limited to opioid users?) | *S. aureus* nasal colonization *(14% MRSA)* | Yes: 60/254  No: 5/28  P=0.49 |  |
| *Substances injected* |  |  |  |  |
| Leung 2015^105^ | Speedball, injection | *S. aureus* nasal colonization |  | aOR 1.02 (0.94 – 1.10) |
| Leung 2015^105^ | Heroin (alone), injection | *S. aureus* nasal colonization |  | aOR 1.10 (0.98 – 1.24) in full “Factors associated with” vs. QIC best-fit model  aOR 1.13 (1.01-1.27) in second model specifically controlling for gender, employment status, HIV status, and ARV use, +/- other variables? (unclear) |
| Leung 2015^105^ | Cocaine (powder or crack), injection | *S. aureus* nasal colonization |  | aOR 0.98 (0.89 – 1.09) |
| Leung 2015^105^ | Crystal meth, injection | *S. aureus* nasal colonization |  | aOR 0.99 (0.89 – 1.10) |
| Leung 2015^105^ | Oxycontin, injection | *S. aureus* nasal colonization |  | aOR 1.01 (0.90–1.13) |
| *Other substance use* |  |  |  |  |
| Leung 2015^105^ | Marijuana | *S. aureus* nasal colonization |  | aOR 0.98 (0.90 – 1.06) |
| Leung 2015^105^ | Heroin, non-injection use | *S. aureus* nasal colonization |  | aOR 0.93 (0.86-1.12) in full “Factors associated with” vs. QIC best-fit model  aOR 0.90 (0.83 – 0.98) in second model specifically controlling for gender, employment status, HIV status, and ARV use, +/- other variables? (unclear) |
| Leung 2015^105^ | Cocaine (powder or crack), non-injection use | *S. aureus* nasal colonization |  | aOR 0.98 (0.90 – 1.06) |
| Leung 2015^105^ | Crystal meth, non-injection use | *S. aureus* nasal colonization |  | aOR 1.06 (0.96 – 1.18) |

### Appendix 16. Details on handling of variables for meta-analysis of social determinants of injection drug use-associated bacterial and fungal infections

#### Studies where outcome is incident or prevalent injecting-related infection.

###### Demographic factors

###### Gender

Multiple related outcomes:

- Islam 2019^21^ had two related sets of unadjusted and adjusted effect estimates, one at 3 months follow-up and one at 6 months follow-up. We included only the 6 month follow-up to avoid double-counting.
- Morin 2020^31^ had three separate infectious outcomes (based on different discharge diagnosis codes), and provided only 1 decimal point in ORs and confidence intervals. We could not include these in meta-analysis when 95% confidence interval differs by 0.1 or less (because this implies the standard error is 0). The authors provided more detailed ORs by email follow-up, but only for one compound outcome variable. We used this one outcome variable here.
- Stein 2020^50^ had OR for hospitalizations and for ED visits (same participants). Kept ED visits and dropped hospitalizations. Also, reference group for ORs was female, so took inverse of OR and 95% CI.
- Wurcel 2018^59^: Kept OR for abscess “ever” (because this was aligned with more of the exposures). Dropped OR for “abscess past 30 days”.

Subsamples:

- Islam 2019^21^ had separate effect estimates for whole sample and for subset of “high frequency injectors” (>1 injection per day). Kept estimate for whole sample only.
- Hope 2010^19^ had one OR for whole sample (for abscess in past year) and one OR for subsample reporting injecting in past 4 weeks. Since exposure is time-fixed, took whole sample OR.

Studies with “not significant” findings (but no statistics presented):

- Binswanger 2000^6^: “Did not significantly differ”
- Hope 2014^17^: “No associations found” for abscess outcome or cellulitis outcome
- Wurcel 2016^58^: “No significant changes”
- Hope 2015^18^: “Not associated” for abscess outcome or cellulitis outcome

###### Age

Multiple related outcomes:

- Stein 2020^50^, has highly related outcomes for ED visits for injecting-related infections and hospitalizations for injecting-related infections in same sample. Kept only ED visits.

Collapsed more than two levels of exposure:

- Baltes 2020^1^ (combined <40 and 40 or older)
- Betts 2016^4^ (kept only age 36-45 vs. 35 or less, and excluded OR for age 46-71
- Doran 2020^14^ (UAM and for C&P): (kept 35+ years vs. less than 25, excluded 25-34 years)
- Dunleavy 2017^15^ (combined >30 vs. 30 years or less for uOR, because this was assessed in aOR
- Fink 2013^16^ kept 50+ years vs. under 30 years
- Hope 2010^19^ combined 35 years or older vs. under 35 for uOR; aOR not reported (because not included in stepwise regression)
- Hope 2008^20^ kept uOR and aOR for 35 years and older vs. under 25 years
- Hope 2015^18^ combined frequencies for uOR 30 and olver vs. under 30
- Murphy 2001^32^ Combined 40 years and older vs. and less than 40 for uOR frequencies, for two comparable sized groups
- Noroozi 2019^34^ for uOR and aOR kept 40 years and older vs. under 30
- Wright 2020^57^ kept only 45+ years vs. less than 35
- Morin 2020^31^ reference was 65+ and broken into six levels. Kept 25-34 vs. 65+ (because CI did not have same values as point estimate, so could include in meta-analysis) and inverted it.

Inverted effect estimates (study treated “older” as reference group, so took inverse for meta-analysis):

- Cedarbaum 2016^8^ was aOR 0.32 (0.16-0.65), now aOR 3.13 (1.54 - 6.25)
- Morin 2020^31^ reference was 65+ and broken into six levels. Kept 25-34 vs. 65+ (because CI did not have same values as point estimate, so could include in meta-analysis) and inverted it.

Studies with “not significant” findings (but no statistics presented):

- Binswanger 2000^6^; Dahlman 2017^12^; Hope 2014^17^; Hope 2015^18^ (cellulitis)

Other:

- Cooper 2005^10^, in an ecological study, assessed effect of percent of neighborhood residents aged 18-64 years, vs. less than 18 OR older than 64 years (so not younger or older).
- Thønnings 2020^52^: reported as p=0.21 but confidence interval does not cross one. This is impossible. So excluded both uOR and aOR
- Wurcel 2016^58^ “…the percentage of IDU-IE hospitalizations among young adults (15–34 years) steadily increased from 2000 to 2013, with a steep increase from 2008 to 2013 (27.7%–42.0%; P < .001 using χ2 test for trend in proportions). In contrast, IDU-IE rates among middle-aged adults (ages 35–54) steadily decreased between 2000 and 2013 (67.2%–39.9%; P < .001).”

###### Race/ethnicity

Inverted effect estimates

- Fink 2013^16^: had White as referent for adjusted odds ratios, and separate categories for Black, Latino, Other. Inverted Black category from aOR 0.95 (0.62-1.45) to 1.05 (0.69-1.61). Inverted Latino category from aOR 0.87 (0.57-1.31) to aOR 1.15 (0.76-1.75)
- Milloy 2010^30^: had White as referent and Aboriginal (ndigenous) for incident rate ratios. Inverted uRR from 0.72 (0.47-1.09) to 1.39 (0.92-2.13) and aRR from 0.71 (0.47–1.07) to 1.41 (0.93-2.13)
- Safaeian 2000^44^ Black as exposure and other (?white) as referent. Inverted from uOR 1.8 (1.0-3.3) to 0.56 (0.30-1.00)

Multiple related outcomes

- Stein 2020^50^ included as separate outcomes for ED visit and hospitalization with injecting-related infections. We included only ED visits.

Other:

- Smith 2015^49^ includes African American as referent group, so could include OR vs. “Caucasian” but could not include OR vs. Native American. With Native American as the exposure (uOR 7.50; 95%CI 0.92-60.90 and aOR 7.35; 95%CI 0.48-113.36). The study also included a race category for “Other” including Hispanic, Asian, or “multiple” that had two few participants for regression.
- Wurcel 2016^58^ was a USA nationwide ecological study of hospital records, and reported that the percentage of hospital admissions for endocarditis that were attributable to injection drug use increased among white people from 40.2% in 2000 to 68.9% in 2013 (P < .001). The proportion of hospital admissions for endocarditis that were attributable to injection drug use for “non-white” people appeared stable, but a missing data category decreased substantially over time.

###### Education

Inverted effect estimates:

- Betts 2016^4^ had tertiary education as reference group. Was aOR 0.74 (0.55-1.01), now 1.35 (0.99 – 1.82)

Outliers:

- Shah 2020^46^ was outlier (uOR 3.12, 95%CI 1.34-7.23) for summary uOR including it (0.98, 95% 0.80-1.21). Removing Shah 2020 changed summary uOR to 0.92 (95%CI 0.77-1.09).
- After removing Shah 2020, then Phillips 2017^36^ (uOR 2.4249 [1.2000; 4.9000]) became an outlier. Removing Phillips 2017 changed summary uOR to 0.8711 [0.7642; 0.9929]

###### Income/employment

Could not include in meta-analysis:

- Ciccarone 2016^9^ included “percent unemployment” and “percent poverty” in multivariable regression but did not define these and we were unable to extract the confidence intervals from the figure where they were presented. It seems like “Percent unemployment” had point estimate of OR 1.00 and was not statistically significant, and “Percent poverty” had point estimate of OR 0.98 with p<0.05.
- Summers 2017^51^: “Reported income” was exposure but categories undefined. uOR and aOR were both 1.00 (1.00-1.00) so could not include in inverse-weight meta-analysis. So excluded.

Multiple related outcomes:

- Hope 2014^17^ reported highly related outcomes of abscess and cellulitis (redness, swelling, tenderness). Included only cellulitis as this was more common.
- Noroozi 2019^34^ it seems like employment status and monthly income were combined into “socioeconomic status”, so only used the latter

Collapsed more than two levels:

- Murphy 2001^32^ collapsed three levels into annual family income less than 10,000 or 10,000+
- Morin 2020^31^ for all outcomes, took OR of lowest neighbourhood income vs. highest (quintiles).

Inverted effect estimates

- Roux 2020^42^ had “employed” as exposed and unemployed as referent. Inverted from uOR 0.84 (0.42-1.66) to 1.19 (0.60 – 2.38)
- Safaeian 2000^44^ for both Endocarditis and Abscess outcomes has lower income as reference and higher income has exposures. Inverted from uOR 0.7 (0.4-1.3) to 1.42 (0.77-2.5) and from uOR 0.7 (0.5-0.9) to 1.43 (1.11-2.00)

###### Social and housing support characteristics

###### Incarceration history

Multiple related outcomes:

- Two studies [Hope 2014; Hope 2015]^17,18^ provided separate effect estimates for two related outcomes measures (abscess and cellulitis, both of which we categorized as SSTI), but reported only “no associations found” so we could not include them in meta-analyses.

Reporting no difference without statistics:

- Two further studies [Hope 2008; Saeland 2014]^20,43^ reported no significant association but did not provide data.

Other:

- One study [Pollini 2010]^39^ found no evidence of an association between self-reported injecting during incarceration in past 6 months and self-reported abscess in past 6 months.

###### Sex work

Multiple related outcomes:

- One study [Hope 2015]^18^ included two separate analyses for related (but different) outcomes in the sample, self-reported abscess and self-reported “redness, swelling, or tenderness” (we have labelled as cellulitis); there is a unadjusted analyses for both and an adjusted analysis only for abscess. We included both analyses in the main meta-analysis and then performed a sensitivity analysis including only the abscess outcome analysis. In sensitivity analysis omitting Hope 2015 cellulitis outcome from unadjusted analysis (including only abscess outcome), the summary effect estimate was generally the same: OR 1.68 (1.04 – 2.75).

Multiple related exposures:

- One study [Pollini 2010]^39^ provided effect estimates for two related exposure measures of sex work (“Principal source of income was through sex work” and “Traded sex or money for drugs”); we included only the “Principal source of income” measure.

Reporting no difference without statistics:

- In one study [Saeland 2014]^43^, the association between sex work and injecting-related infection was found to be not statistically significant but the data was not reported, so this could not be included in meta-analysis.

###### Unstable housing and homelessness

Multiple related exposures:

- One study [Dunleavy 2017]^15^ provided effect estimates for two related exposure measures of homelessness (lifetime history [OR 0.90, 95% CI 0.72-1.13] vs. past 6 months [OR 0.90, 95% CI 0.72-1.13]) in relation to SSTI in the past year, so only the past 6 months analyses was included in meta-analysis.

Collapsing more than two levels:

- Two studies [Hope 2014; Hope 2015]^17,18^ categorized their homelessness exposure into three levels as “Never”; “Yes but not in last year”; and “Yes in last year”. For unadjusted odds ratios we treated this as “Yes in last year” vs. other, but in adjusted odds ratios (provided only in Hope 2015) the pre-calculated odds ratio provided was for “Yes in last year” vs. “Never” as the reference group.

Reporting no difference without statistics:

- Across three studies [Biswanger 2000; Hope 2014; Hope 2015],^6,17,18^ three tests of association were found to be not statistically significant but the data was not reported, so we could not include them in the meta-analyses.

###### Substance use-related factors

###### Overdose history

Reported no difference, without providing statistics:

- Saeland 2014^43^ reports only that groups with and without history of overdose, “did not differ”, so was not included in meta-analysis.

Multiple related exposures and outcomes

- Hope 2014^17^ and Hope 2015^18^ had separate exposure categories for (a) history of overdose in preceding year and (b) history of overdose, but not in preceding year, and compared these to reference category of “never” overdosed. We kept only history of overdose in preceding year, noting that the effect estimate is only in reference to people who never overdosed and excludes people that overdosed prior to the past year. Also both papers had separate effect estimates for abscess and for cellulitis (“redness, swelling, and tenderness”. All were statistically significantly increased. For meta-analysis, we only include estimates of abscess.

###### Heroin use

Multiple related outcomes:

- Hope 2014^17^ and Hope 2015^18^ had separate analyses for abscess outcome and cellulitis (“redness, swelling, or tenderness”) outcome. In Hope 2014, only statistics for abscess outcome were reported, and for cellulitis was only “no associations found”. For Hope 2015, only reported as “not associated” and so not included in meta-analysis.

Collapsed more than two levels of exposures:

- Hope 2010^19^ separated out “Opiate, no stimulant”; “Stimulant, no opiate”, and “Stimulant and opiate”. We compared only “Opiate, no stimulant” and “Stimulant, no opiate” (interpreted as opiate/heroin only vs. no opiate use). Since “Opiate, no stimulant” was reference group for multivariable/adjusted odds ratio, took inverse; transformed aOR 0.47 (0.30-0.75) (for stimulant use) to 2.12 (1.33 - 3.33). Also had separate estimates for whole sample and sample injecting in past 4 weeks (but outcome was any SSTI in past year), so kept estimate for whole sample.

Other:

- Lloyd-Smith 2009^27^ and Milloy 2010^30^ reported odds ratios for statistics assessed during baseline study visits, and hazard ratios or rate ratios for incidence studies during follow-up. We included the hazard ratios and rate ratios rather than the baseline odds ratios.

Reported no difference, without providing statistics:

- Other studies reporting “no associations found” (and therefore providing no statistics to include in meta-analysis) include Lloyd-Smith 2005^24^ (for multivariable analysis), Lloyd-Smith 2012^25^ (for multivariable analysis among men, but did include stats among women [because was “significant” in unadjusted analysis within stepwise regression], Milloy 2010^30^ (for multivariable analysis, dropped in stepwise regression), Pollini 2010^39^ (for multivariable analysis, dropped in stepwise regression).

Heroin type/formulation

- One ecological study [Ciccarone 2016]^9^ found that rates of SSTI was higher in cities with predominantly Mexican-sourced (tar) heroin compared to cities with predominantly Colombian-sourced (powder) heroin; this was true after covariate adjustment (aOR 2.05, p<0.001). The confidence interval values were not reported, so we extracted them from the figure.

###### Cocaine

Multiple related outcomes:

- Hope 2014^17^ and Hope 2015^18^ had separate estimates for abscess and cellulitis (“redness, swelling, and tenderness”), so we kept only abscess
- Lloyd-Smith 2010^28^ and Milloy 2010^30^ have baseline/cross-sectional OR and longitudinal/incidence HR (or RR). So we kept only HR (or RR) and not OR.

Other:

- Roux 2020^42^ did not specify cocaine the text (only saying “stimulant”), but next variable described is “speedball” (which is a combination of cocaine and heroin), so we interpret “stimulant” here to mean specifically cocaine.
- Hope 2010^19^ found that people who reported injecting only “stimulants” (not otherwise specified) had a lower risk of prevalent injecting-site infection than people who inject only opiates, for abscess (aOR 0.49, 95%CI 0.34-0.71) and cellulitis (aOR 0.47, 0.30-0.75). However, a small minority of participants used only stimulants (206 out of 4,484; 4.6%). As not specified (and would be outlier for cocaine anyway), left out of synthesis.

###### Methamphetamine and amphetamines

Reported no difference, without statistics:

- Hope 2014^17^ and Hope 2015^18^ had separate estimates for abscess and cellulitis, but all were reported as “not associated” with no statistics, so were excluded.
- Hope 2008^20^ reported only “not associated” so was excluded

###### Speedball (cocaine and heroin together) and goofball (methamphetamine and heroin/fentanyl together)

Multiple related outcomes

- Lloyd-Smith 2012, Lloyd-Smith-2009, and Milloy 2010^25,27,30^ all had cross-sectional/baseline odds ratios and longitudinal/incidental HRs or RRs, so only kept longitudinal measurements

###### Alcohol use

Multiple related exposures:

- Phillips 2008^37^ had two related exposure measurements of hazardous alcohol use, AUDIT >= 8 and alcohol intoxication days in past month. We kept alcohol intoxication days in past month.

Collapsing multiple levels of exposure

- Wilson 2002^56^ had three levels: no alcohol; 1-21 drinks per week, and >21 drinks. We kept only >21 drinks per week and categorized as “Hazardous”

###### Drug policy factors

###### Police contacts and arrests

Multiple related outcomes:

- One study [Cooper 2005]^10^ was an ecological study assessing the impact of policing crackdowns on local rates of hospitalization for injection drug use-associated SSTI and endocarditis. While multiple effect estimates are presented, we included the IRR for the “first crackdown quarter” based on the authors’ stated hypothesis.
- Two studies [Hope 2014; Hope 2015]^17,18^ provided two related effect estimates for associations between arrest in the past year with abscess and with cellulitis (in the past month for Hope 2014, and in the past year for Hope 2015). Hope 2014 reported only “no associations found” between arrest and the abscess outcomes, so we included only the cellulitis outcome. Hope 2015 reported only “not associated” for the cellulitis outcome, so we included only the abscess outcome.

Multiple related exposures:

- One study [Pollini 2010]^39^ assessed several different exposures related to police contacts amongst the same sample of PWID in Tijuana, Mexico. This included unadjusted effect estimates for being arrested for sterile syringes, arrested for used syringes, arrested for track marks, and police asking you for money, and unadjusted and adjusted estimates for “Police affected where you used drugs”. We included all these separate estimates.

###### Assisted injecting, or requiring help to inject

Other:

- One study [Lloyd-Smith 2012]^25^ provided one unadjusted OR for the whole sample, but for adjusted OR only provided sex-stratified estimates.

###### Injecting with others

Multiple related exposures:

- Pollini 2010^39^ assessed three different exposures in the same sample (alone/never injected alone, with friends, with family/spouse). Included all.
- Roux 2020^42^ had sample broken up into three levels, (a) injected alone or did not inject (as reference), (b) with someone else, (c) in group. Included all.
- Smith 2015^49^ had exposure in three levels: (a) injected alone (as reference), (b) with friends, (c) with family/spouse. Included all.

###### Harm reduction and drug treatment

###### Needle and syringe distribution programs

Multiple related exposures:

- Dunleavy 2017^15^ reported two separate effect estimates for uptake of sterile needles and syringes and for uptake of sterile “paraphernalia” (i.e., filters and cookers/spoons), both of which we included. They reported a third effect estimate which was a composite of these two variables; we excluded this to avoid double-counting.

Multiple estimates from subsamples:

- Hope 2010^19^ reported one effect estimate for the whole sample and a second effect estimate for the subsample of participants who reported injecting in the past four weeks; since the exposure and outcome definitions related to the past year (rather than only the past four weeks) we included the effect estimate for the full sample and excluded the one for the subsample to avoid double-counting.

###### Opioid agonist treatment

Continuity correction:

- In Sierra 2006^47^ there were zero injecting-related infections in the methadone group, resulting in an infinity confidence interval that could not be included in meta-analysis. When we perform a continuity correction by adding 0.5 to all cells in the 2 by 2 table, the study OR changes from 0.03 (0.00 – 0.19) to 0.08 (0.00 – 1.50) and the summary uOR changes from uOR 0.76 (95%CI 0.65-0.89) to 0.75 (0.63 – 0.89).

Inverted effect esimates:

- Two studies [Betts 2016; Hope 2008]^4,20^ provided an effect estimate for NOT being on OAT, so we took inverse.

Handling more than two exposure levels

- Several studies [Dunleavy 2017; Hope 2010; Hope 2008]^15,19,20^ assessed currently being on OAT vs. previously being on OAT, treating never on OAT as a distinct category. We included this current OAT vs. previous OAT effect estimate. In these studies, being “previously” on OAT was associated with higher risk of infections compared to never being on OAT. This may include people without opioid use disorder or people without substance dependence.

Other:

- Several studies compared multiple types of OAT, rather than OAT vs. no OAT, and were excluded from the meta-analysis of OAT vs. no OAT.
- One study [Bertin 2020]^3^ compared incidence of injecting-related bacterial infections amongst patients starting buprenorphine, methadone, and second-line/alternative OAT with morphine sulfate:
  - Crude incidence per 100,000 PY, by cohort -- MS: 7.0 (4.7 – 10.6); Bupe: 2.2 (1.8 – 2.5); Methadone: 1.6 (1.2 – 2.0)
  - aHR 2.8 (1.8-4.4) for MS vs. patients starting buprenorphine
  - aHR 3.6 (2.2-5.9) for MS vs. patients starting methadone
  - aHR Adjusted for age, sex, socioeconomic status, chronic alcohol consumption, concurrent BZD use, major chronic somatic or psychiatric comorbid disease
- One study [Oviedo-Joekes 2017]^35^ reported rates of cellulitis or abscess as potential adverse effects within a randomized trial of injectable hydromorphone (7 episodes among 100 patients) vs. injectable diacetylmorphine (17 episodes among 100 patients).

###### Supervised consumption sites

Multiple related outcomes:

- One study [Scherbaum 2010]^45^ reported rates of past-month skin abscesses among PWID at their first time attending a supervised consumption site and compared this to past-month rates at one, two, and three months follow-up. Since these three effect estimates are among the same sample, we included only the data at the first month follow-up because most the sample was lost to follow-up after that.

##### Outcomes during treatment for injecting-related infections

###### Healthcare-seeking for injecting-related infections

Multiple related outcomes:

- Hope 2015^18^ had highly related outcomes of abscess and cellulitis (“redness, swelling, tenderness”). Kept abscess because it may be more persistent and recognized as a medical issue.

###### Against medical advice discharge

###### Gender/sex

Inverted effect estimates:

- Jo 2021^63^ had male as exposure. Inverted from aOR 0.83 (0.62 – 1.11) to aOR 1.20 (0.90 – 1.61)
- Mertz 2008^70^ had male gender as exposure. Inverted from aOR 1.2 (0.6 – 2.2) to aOR 0.83 (0.45 – 1.67)
- Serota 2021^76^ had male gender as exposure. Inverted from aOR 0.86 (.80, .92) to aOR 1.16 (1.09-1.25)

###### Age

Inverted effect estimates:

- Jo 2021^63^ inverted to aOR 1.04 (95%CI 1.03-1.06)
- Mertz 2008^70^ inverted to aOR 1.25 (95%CI 0.83-2)

###### Health insurance

Multiple levels of exposures:

- Jo 2021^63^: uninsured is reference (vs. levels government-funded insurance or private insurance). Inverted government funded insurance from aOR 0.45 (0.33, 0.61) to aOR 2.22 (1.64 – 3.03)
- Kimmel 2020^64^: Medicaid is reference. Kept “self” as source of payment; aOR 1.37 (1.13 – 1.66)

###### All-cause rehospitalization

###### Gender/sex

Inverted effect estimates

- Jo 2021^63^ had females as reference. Inverted to aOR 1.12 (95%CI 0.9-1.41)

###### Opioid agonist treatment

Multiple outcomes:

- Barocas 2020^82^ had separate effect estimates for hazard of all-cause rehospitalization within 30 days and within 1 year. Since exposure was OAT within 30 days, that outcome likely affected by immortal time. Kept 1 year outcome.
- Wang 2020^81^ had separate effect estimates for binary yes/no rehospitalization within 30 days and within 90 days. Since exposure was OAT provided on hospital discharge, kept 30 days outcome

##### Colonization outcomes

Multiple related exposures:

- Leibler 2019^104^ had multiple related exposures, kept “sleeping in homeless shelter” and excluded sleeping on the street (“no associations were observed”). Moved :”sleeping >1 place during the last week” and “use of public shower facilities” and “sharing bedding” to it’s own “other” category.
- Miller 2007^3^ had two related exposures: “Homeless during past six months” and “spent time in a homeless shelter during the past six months”. Kept only “Homeless” in meta-analysis, as assumed this was inclusive of both.

### Appendix 17. Synthesis and meta-analyses for studies where outcome is incident or prevalent injecting-related infections

Sixty studies assessed factors associated with incident or prevalent injecting-related infections. Demographic characteristics included gender/sex; age; race/ethnicity; education; income/employment; relationship status; and migration status. Social and housing support characteristics included incarceration history; sex work; food insecurity; unstable housing/homelessness. Substance use factors included overdose history; heroin formulation; heroin use; prescription-type opioids; cocaine use; amphetamines; prescription-type stimulants; other/combined stimulant use; speedball use; other/polysubstance use; alcohol use; and smoking. Drug policy and injecting context factors included drug policy changes; drug purchasing network; injecting in public; shooting gallery; police contacts and arrests; assisted injecting/requiring help to inject; and injecting with others. Health and social services factors included access to needle and syringe programs; opioid agonist treatment; supervised consumption sites.

#### Sociodemographic characteristics

###### Gender/sex

35 studies investigated associations between gender/sex and incident or prevalent injecting-related infections.^1,4,6,11,12,14–21,23–28,30–32,36,37,39,42,44,46,47,49,50,56–59^ We combined gender and sex into one category because studies tended to use these terms interchangeably and did not typically define how they were ascertained. Doran 2020^14^ reported effect estimates from two different study samples (the “Care & Prevent Study” [C&P] and the Unlinked Anonymous Monitoring survey [UAM]) and we included both of these.

Thirty-one unadjusted effect estimates were included in meta-analysis, resulting in a summary unadjusted odds ratio (uOR) for woman/female (vs. man/male) of 1.56 (95% confidence interval [CI] 1.34-1.82; Figure 1). There were four outliers [Hope 2010; Morin 2020; Dunleavy 2017; Lloyd-Smith 2005].^15,19,24,31^ Removing these changed the summary to uOR 1.65 (95CI% 1.47-1.86; I^2^ 56.8%, p=0.0002).

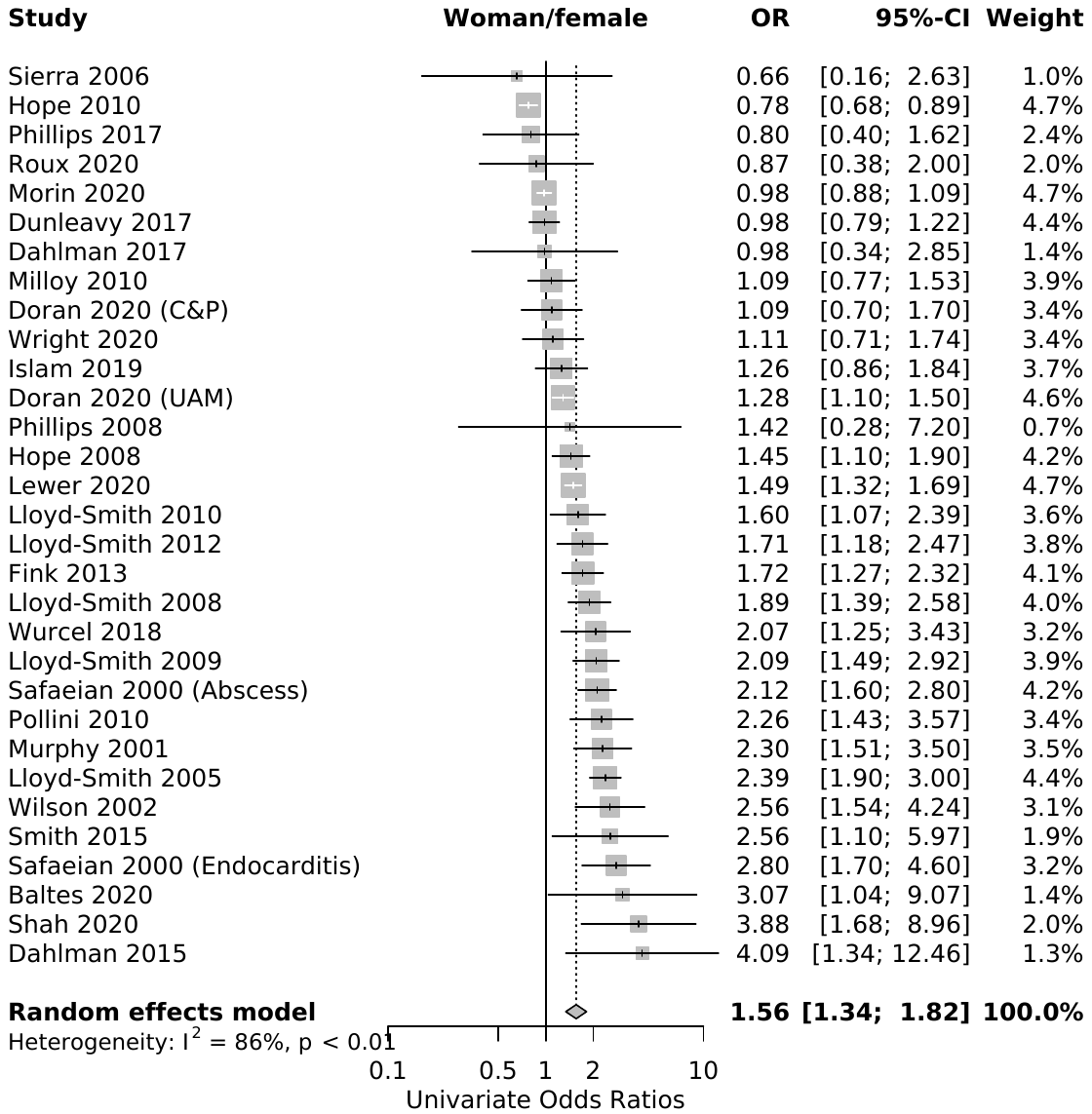
Figure 1. Meta-analysis of unadjusted effect estimates of relationship between woman/female gender/sex and incident or prevalent injection drug use-associated bacterial infections.

Twenty-two fully-adjusted effect estimates were included in meta-analysis, resulting in an adjusted odds ratio (aOR) of 1.59 (95%CI 1.33-1.89) for woman/female gender (Figure 2). There were two outliers [Morin 2020; Safaeian 2000 (Endocarditis)],^31,44^ and removing these changed aOR to 1.57 (95%CI 1.36-1.83; I^2^ 46.8%, p=0.01).

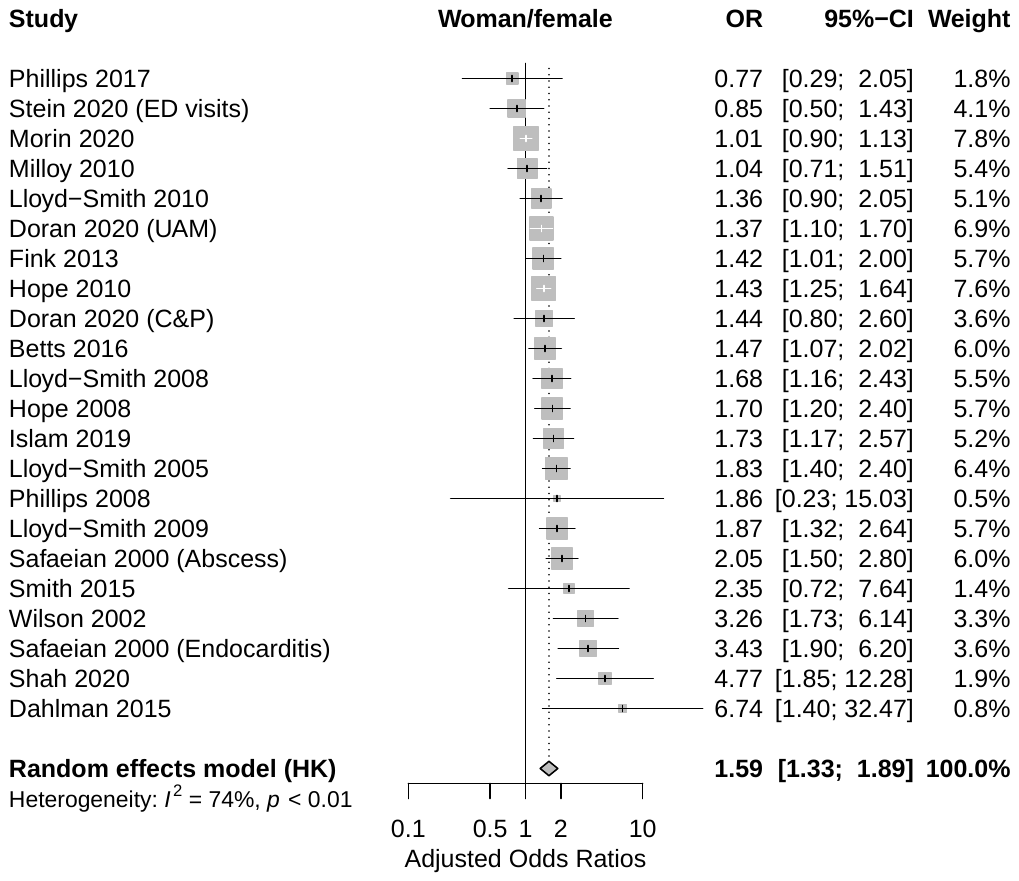
Figure 2. Meta-analysis of fully-adjusted effect estimates of relationship between woman/female gender/sex and incident or prevalent injection drug use-associated bacterial infections.

###### Age

Thirty-nine studies reported analyses of age and incident or prevalent infections.^1,4,6,8,10–12,14–21,25–28,30–32,34,36–39,42–44,46,47,50–52,56–59^ In most studies, participants were grouped by categories of older vs. younger ages, but definitions varied (e.g., age 30 years or older vs. younger; age 35 years or older vs. younger than 35; increasing age in years, treated continuously). Two studies [Lloyd-Smith 2012; Wurcel 2018] provided only sex-stratified effect estimates, and we included all of these.^25,59^

Summary results for 32 unadjusted effect estimates for older age (vs. younger) was uOR 1.00 (95%CI 0.96-1.03; Figure 3). Removing 10 outlier effect estimates changed the unadjusted meta-analysis summary estimate to uOR 1.00 (95%CI 0.99-1.01; I^2^ 25.3%, p=0.14).

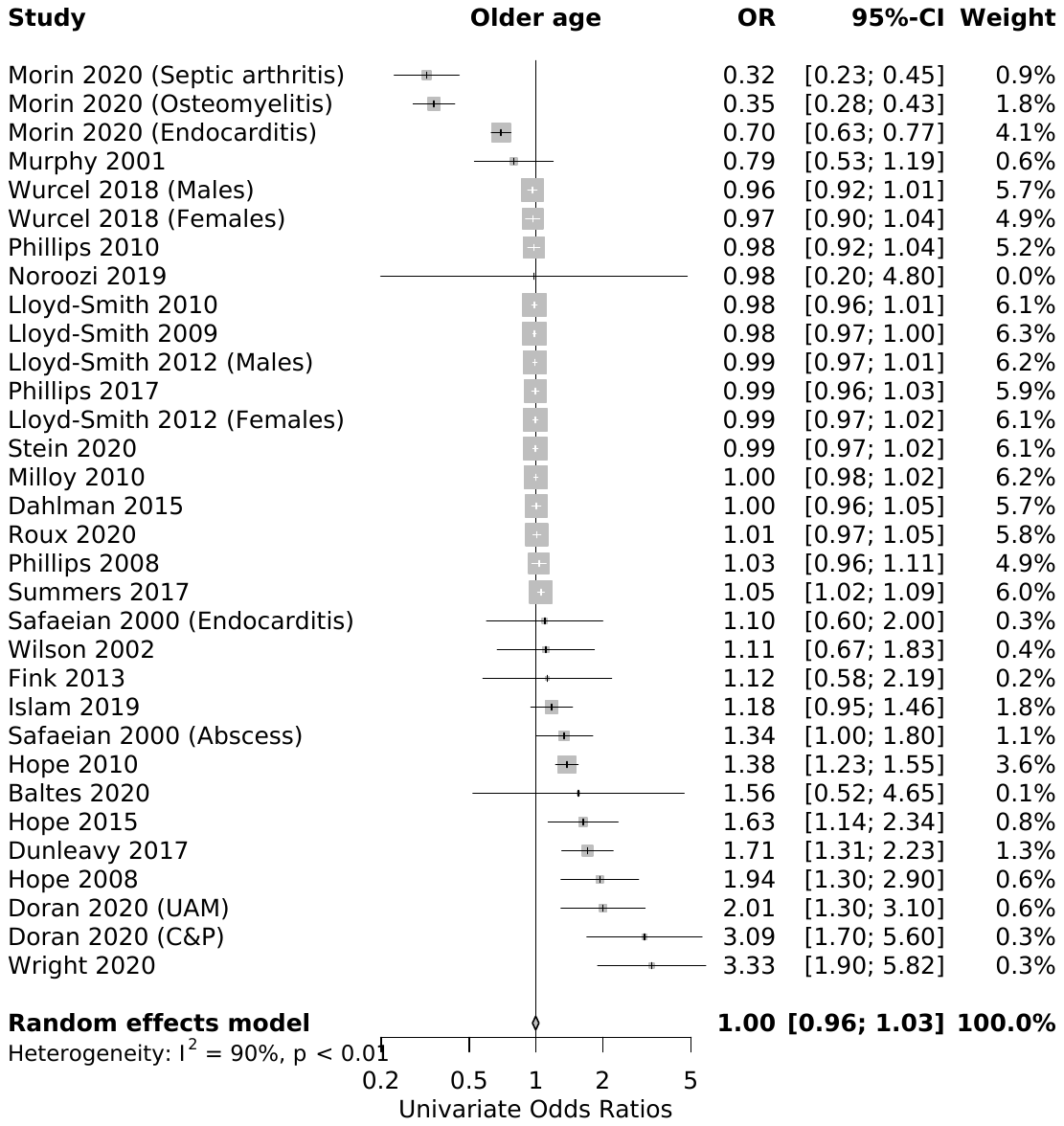
Figure 3. Meta-analysis of unadjusted effect estimates of relationship between older age and incident or prevalent injection drug use-associated bacterial infections.

Meta-analytic summary for 20 adjusted effect estimates was aOR 0.97 (95%CI 0.76-1.24; Figure 4). Removing seven outlier adjusted effect estimates changed the summary estimate to aOR 1.00 (95%CI 0.89-1.10; I^2^ 84.1%, p<0.0001).

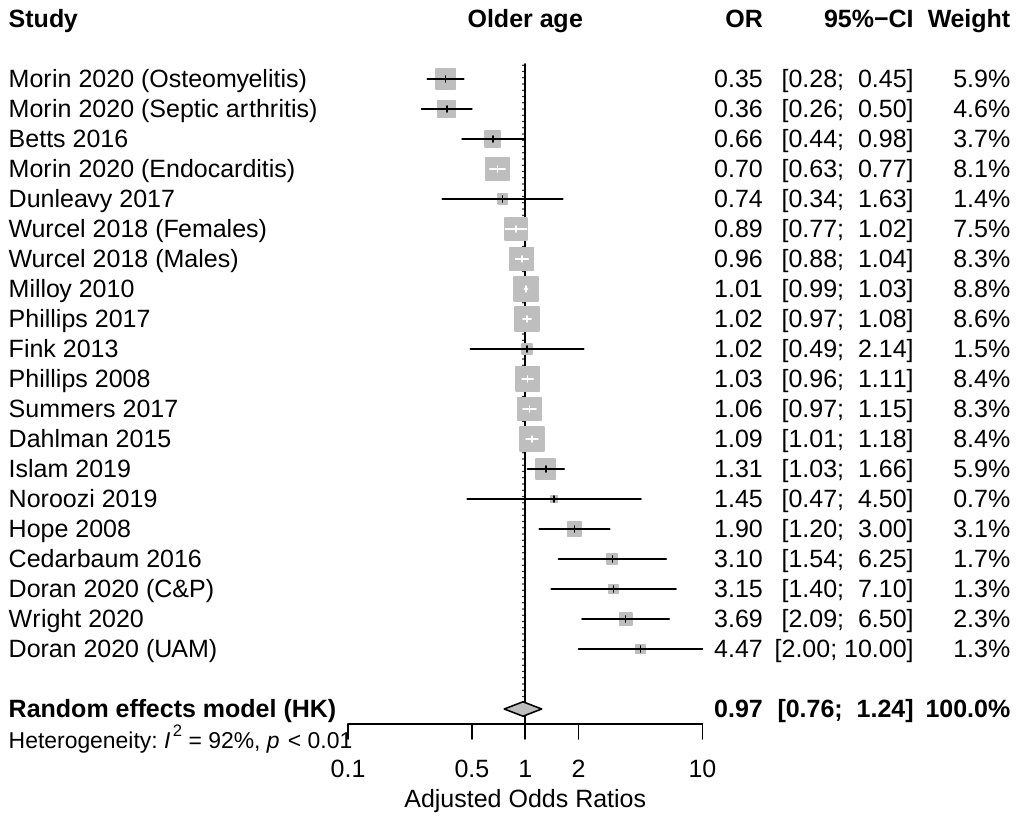
Figure 4. Meta-analysis of fully-adjusted effect estimates of relationship between older age and incident or prevalent injection drug use-associated bacterial infections.

Several of the studies reported age distributions among participants with and without infections, and so were not included in the above meta-analyses of binary effect estimates. In three studies [Pollini 2010^39^, Saeland 2014^43^; Sierra 2006^47^] the age distributions did not significantly differ between groups. In one study [Shah 2020]^46^ participants with endocarditis (mean age 35.5, SD 8.4 years) were younger than participants without (mean age 40.0, SD 11.0 years, p=0.03).

###### Race/ethnicity

Seventeen studies included analyses of race/ethnicity and risk of incident or prevalent injecting-related bacterial infections.^10,12,14,16,30,32,36–38,44,46,49–51,58,59^ Most samples were majority white participants (sometimes defined as “non-Hispanic white”), and as a result most investigators compared risk among white participants to non-white participants (which among various studies included Black, Indigenous, and Latino participants). Few studies described explicitly how participants’ race was identified (i.e., if it was self-reported or presumed by the researcher).

Among 18 unadjusted effect estimates, meta-analysis summary for white race vs. non-white was uOR 0.99 (95%CI 0.97-1.01). Since Cooper 2005 was an ecological study (i.e., the exposure was proportion of neighbourhood residents that were non-Hispanic white, rather than an individual-characteristic), we repeated the meta-analysis without Cooper 2005. Cooper 2005 had also contributed >98% of the weighting in meta-analysis. The updated summary uOR was 1.14 (95%CI 0.90-1.44; Figure 5). Among 12 fully-adjusted effect estimates the summary aOR was 0.99 (95%CI 0.98-1.00). Without Cooper 2005, this changed to aOR 1.15 (95%CI 0.92-1.44; Figure 6).

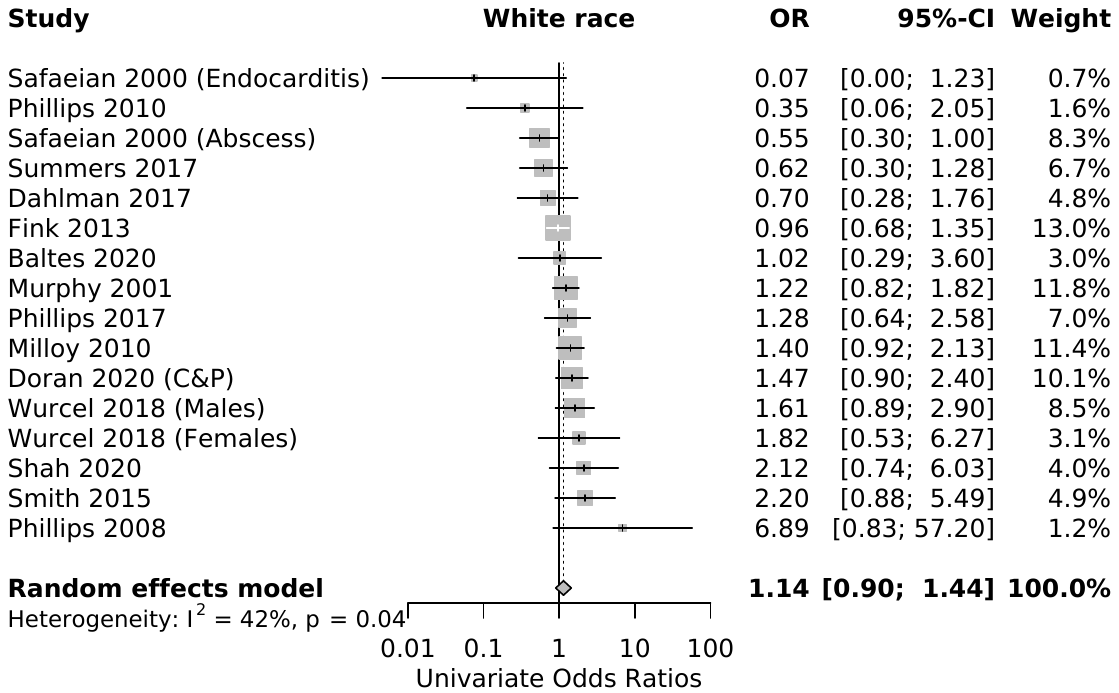
Figure 5. Meta-analysis of unadjusted effect estimates of relationship between white race (vs. other races) and incident or prevalent injection drug use-associated bacterial infections. (Without Cooper 2005.)

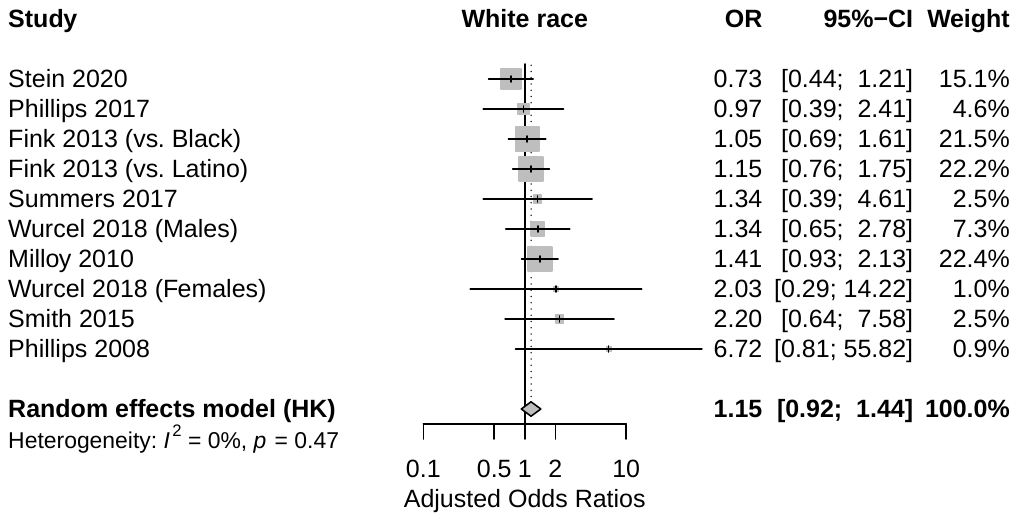
Figure 6. Meta-analysis of fully-adjusted effect estimates of relationship between white race (vs. other races) and incident or prevalent injection drug use-associated bacterial infections. (Without Cooper 2005.)

###### Education

In eleven studies, investigators assessed relationships between level of education and incident or prevalent injecting-related bacterial infections.^4,16,32,34,36,42–44,46,56,59^ Most studies compared a binary measure of more vs. less years of education (typically, completing secondary school vs. did not complete secondary school). Eleven unadjusted effect estimates were available for inclusion in meta-analyses, and the summary was uOR 0.98, 95%CI 0.80-1.21; Figure 7). Two studies [Shah 2020^46^; Phillips 2017^36^] were outliers. After removing these, summary odds ratio was uOR 0.87 (95%CI 0.76-0.99; I^2^ 0.0%, p=0.6). There were four fully-adjusted effect estimates, and the meta-analytic summary was aOR 0.86 (95%CI 0.13-5.55; Figure 8).

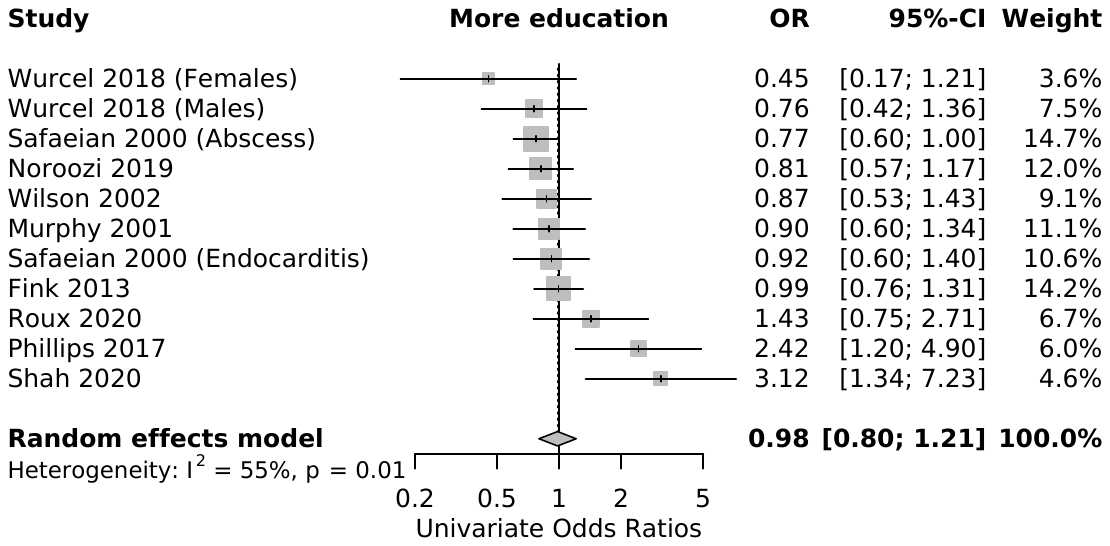
Figure 7. Meta-analysis of unadjusted effect estimates of relationship between greater educational attainment and incident or prevalent injection drug use-associated bacterial infections.

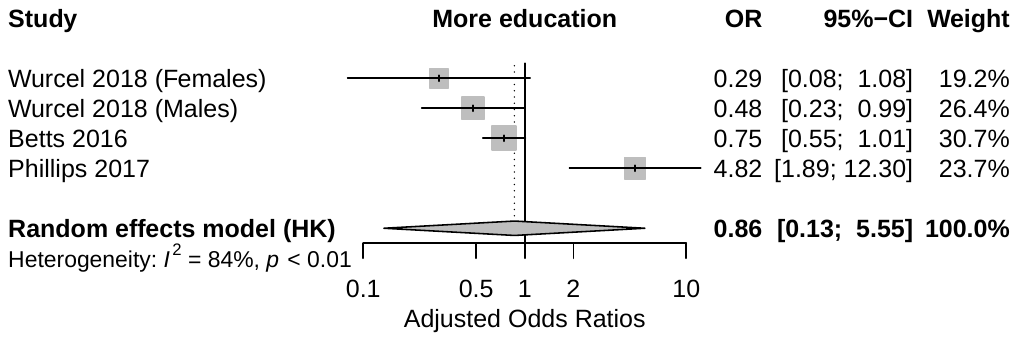
Figure 8. Meta-analysis of fully-adjusted effect estimates of relationship between greater educational attainment and incident or prevalent injection drug use-associated bacterial infections.

One study [Saeland 2014]^43^ reported distributions of years in school among people with abscess (median 11; IQR 9-13 years) and people without (median 11; IQR 9-12 years; p=0.6).

###### Income/employment

Thirteen studies reported assessments between income or employment and injecting-related bacterial infections.^9,10,14,17,18,31,32,34,39,42,44,46,51^ In most studies, investigators treated lower income, poverty, unemployment, or illicit/illegal income generation as exposed and higher income/legal employment as the referent group. Meta-analysis of 16 unadjusted effect estimate resulted in summary uOR 1.44 (95% 1.22-1.71; Figure 9) for lower income and unemployment or illicit/illegal work vs. higher income and employment or legal work. Meta-analysis of 7 fully-adjusted effect estimates resulted in summary aOR 1.16 (95%CI 0.81-1.65; Figure 10).

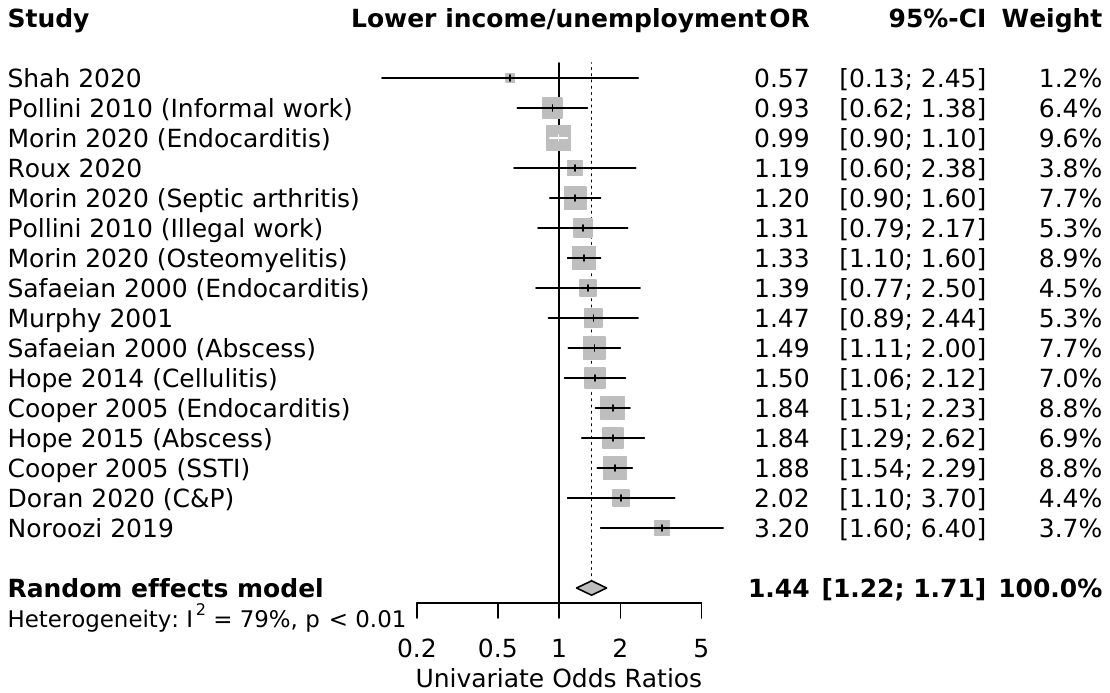
Figure 9. Meta-analysis of unadjusted effect estimates of relationship between lower income/unemployment and incident or prevalent injection drug use-associated bacterial infections.

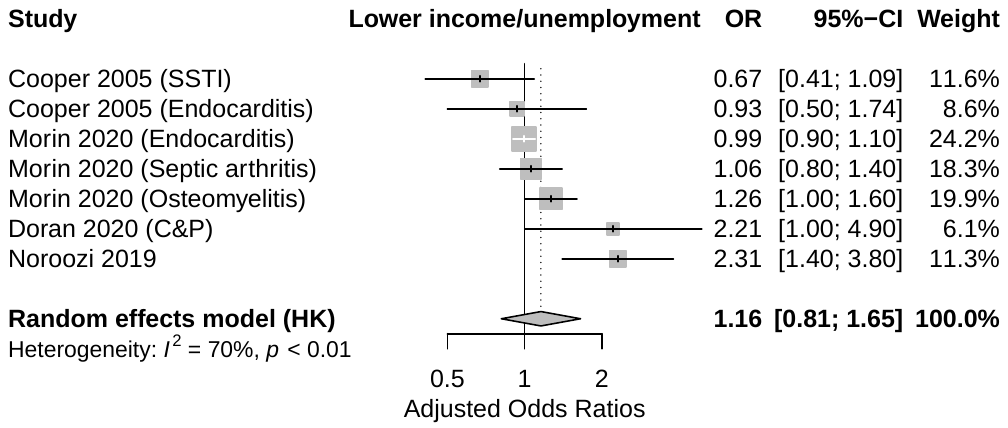
Figure 10. Meta-analysis of fully-adjusted effect estimates of relationship between lower income/unemployment and incident or prevalent injection drug use-associated bacterial infections.

###### Relationship status

Two studies assessed associations between relationship status and incident or prevalent injecting-related infections.^34,42^ Summary meta-analysis for two unadjusted effect estimates (for married or living with spouse, vs. not) was uOR 0.61 (95%CI 0.001-377.0; I^2^ 83.9%, p=0.01) and one adjusted odds ratio was aOR 0.38 (95%CI 0.17-0.85).

###### Migration

Two studies analysed injecting-related infection risk and migration. Doran 2020^14^ did not find that people with SSTI in the past year were more likely to be born in the UK vs. outside the UK (uOR 1.2, 95%CI 0.8–1.6). In Hope 2015^18^, the number of years lived in the current area was associated with having had abscess in the past year in unadjusted analyses (e.g., 10.9% among people who had lived in the current area up to 1 year, and 18.7% among people who had lived in the current area more than 20 years, p=0.02). This relationship was reported as “not associated” in fully adjusted analyses (following stepwise regression) in the manuscript. While we were unable to tell from the data presented, it is possible that people who lived the current area for more than 20 years tended to be older than people who lived in the area up to 1 year, and older age was associated with increased risk of abscess in this study. Migration was also “not associated” with cellulitis (no data reported), in a separate analysis in the same study.

#### Social and housing support characteristics

###### Incarceration history

I identified 11 studies assessing associations between incarceration history with incident or prevalent injecting-related bacterial infections.^14,15,17–20,24,30,39,43^ Meta-analytic summary for six unadjusted effect estimates was uOR 1.27 (95%CI 1.06-1.53; Figure 11) and for two fully-adjusted effect estimates was aOR 1.60 (95%CI 0.99-2.59; Figure 12).

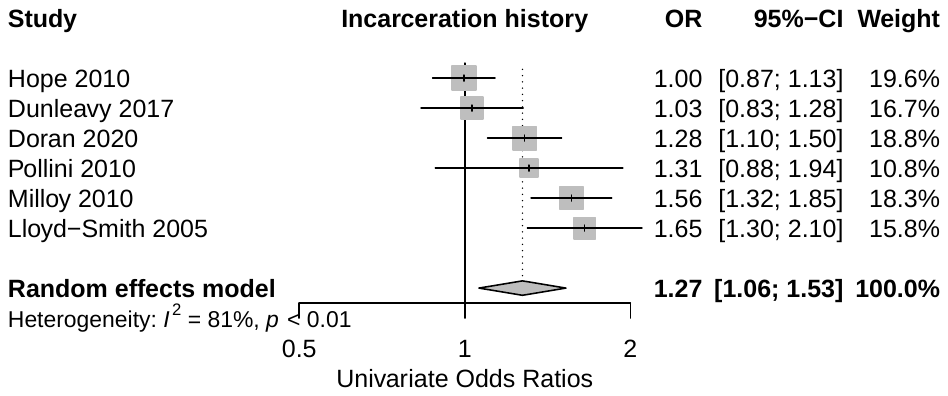
Figure 11. Meta-analysis of unadjusted effect estimates of relationship between incarceration history and incident or prevalent injection drug use-associated bacterial infections.

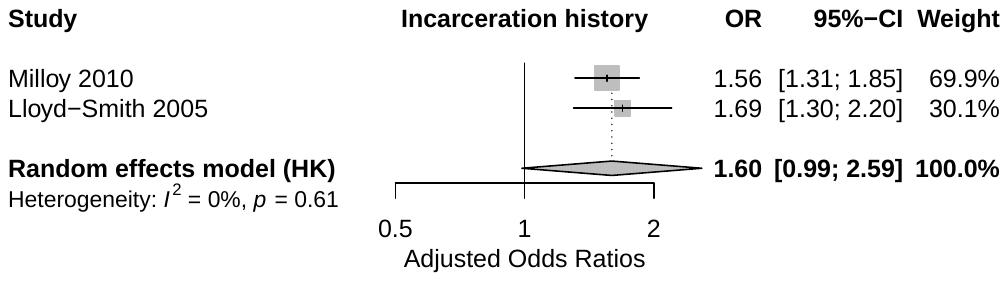
Figure 12. Meta-analysis of fully-adjusted effect estimates of relationship between incarceration history and incident or prevalent injection drug use-associated bacterial infections.

###### Sex work

Seven studies assessed relationships between sex work and incident or prevalent injecting-related bacterial infections.^14,18,24,26,30,39,59^ The summary odds ratio for nine unadjusted effect estimates was uOR 1.66 (95%CI 1.09–2.53; Figure 13). Removing one outlier [Pollini 2010] changed the summary to uOR 1.49, 95%CI 1.06-2.09; I^2^ 88.6%, p<0.0001). Meta-analysis for six fully adjusted effect estimates was aOR 1.58 (95%CI 0.72–3.50; Figure 14).

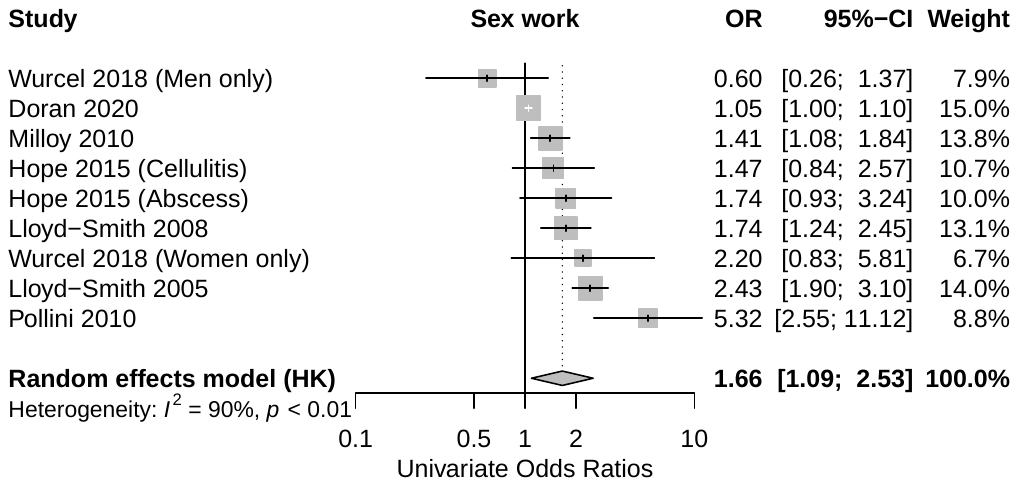
Figure 13. Meta-analysis of unadjusted effect estimates of relationship between sex work and incident or prevalent injection drug use-associated bacterial infections.

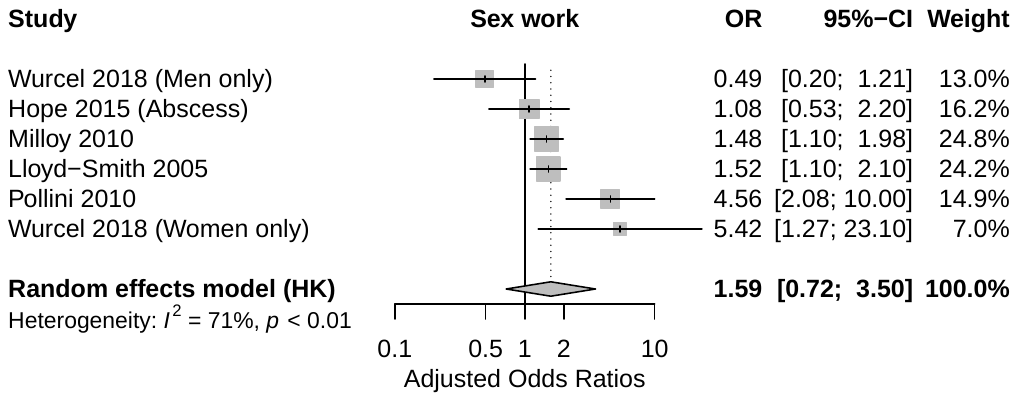
Figure 14. Meta-analysis of fully-adjusted effect estimates of relationship between sex work and incident or prevalent injection drug use-associated bacterial infections.

###### Unstable housing and homelessness

There were 31 studies assessing relationships between unstable housing/homelessness and incident or prevalent injecting-related bacterial infections.^1,4,6,12,14–20,24–28,30,34,36–39,42,46,47,49,51,52,56,57,59^ The summary odds ratio for 32 unadjusted effect estimates was uOR 1.34 (95%CI 1.16–1.55; Figure 15). There were three outliers [Fink 2013; Dunleavy 2017; Noroozi 2019]^15,16,34^. Removing these changed the summary to uOR 1.34 (95%CI 1.20-1.49; I^2^ 43.3%, p=0.008).

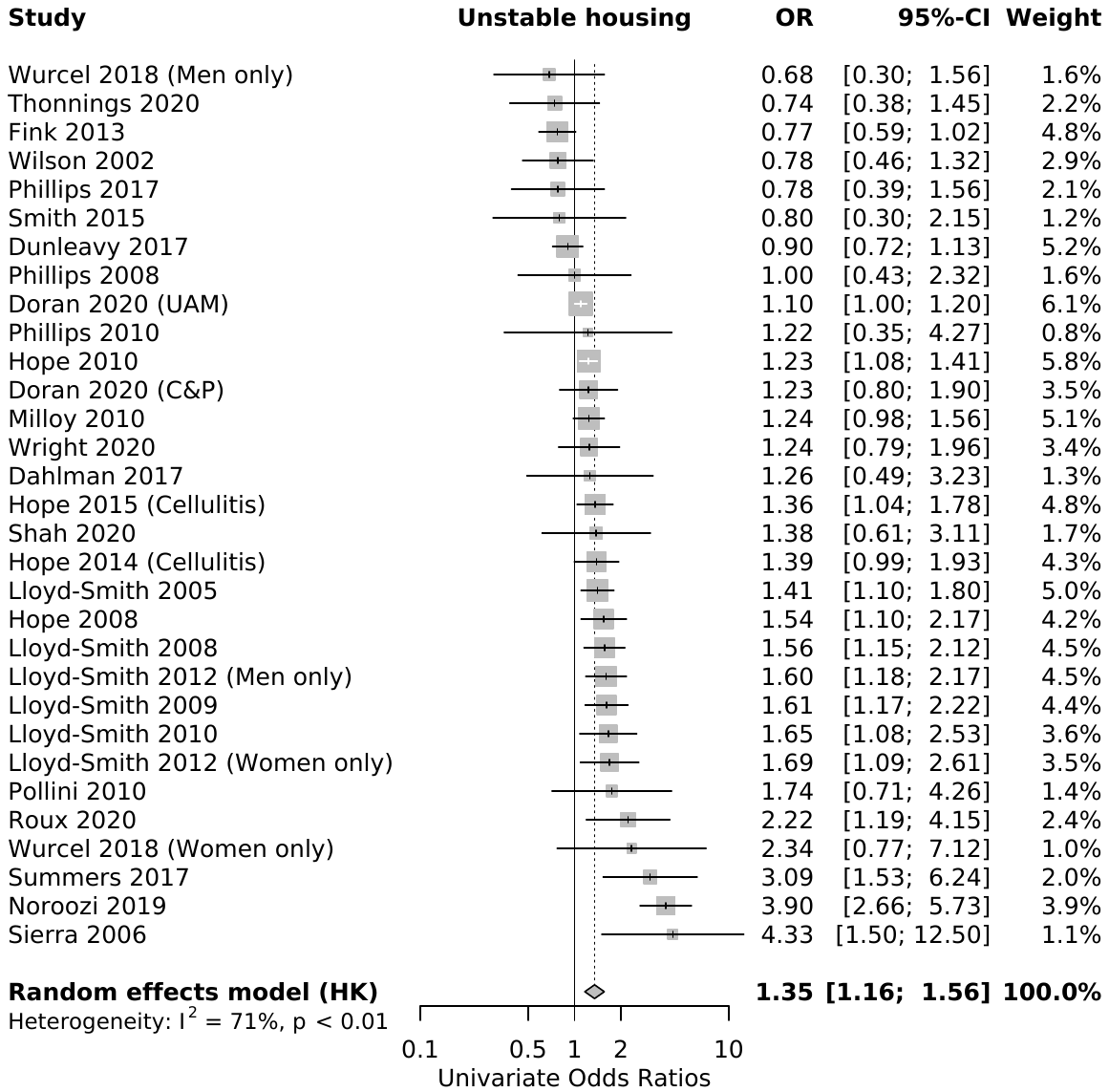
Figure 15. Meta-analysis of unadjusted effect estimates of relationship between unstable housing and incident or prevalent injection drug use-associated bacterial infections.

Meta-analytic summary for 14 fully adjusted effect estimates was aOR 1.29 (95%CI 1.10–1.50; Figure 16). Removing the single outlier [Wurcel 2018 (Men only)]^59^, changed the summary to aOR 1.29 (95%CI 1.16-1.44; I^2^ 8.7%, p=0.4).

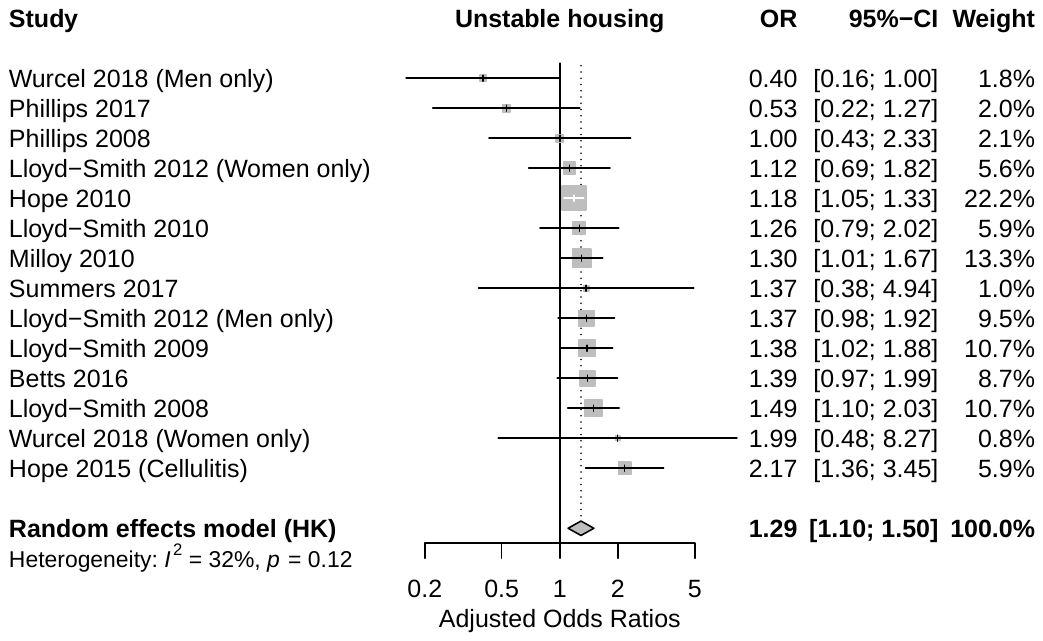
Figure 16. Meta-analysis of fully-adjusted effect estimates of relationship between unstable housing and incident or prevalent injection drug use-associated bacterial infections.

For this analysis, five studies^25–28,30^ used data from the Scientific Evaluation of Supervised Injecting (SEOSI) cohort study in Vancouver, British Columbia, Canada. They reported separate but related outcomes (e.g., self-reported abscess, emergency department visit for injecting-related infection, hospital admission, etc.) for very similar or identical samples of participants. We performed two sensitivity analyses to test the impact of having multiple related estimates from the same sample. Keeping only the earliest study [Lloyd-Smith 2008]^26^ and removing the rest changed the summary effect estimates to uOR 1.29 (95%CI 1.13-1.47; I^2^ 72.6%, p<0.0001) and aOR 1.34 (95%1.02-1.75; I^2^ 42.9%, p=0.09). Keeping only Milloy 2010^30^ (which was the only one of these studies where incarceration history was specified in the research question or treated as the main exposure of interest) changed the summary effect estimates to uOR 1.26 (95%CI 1.12-1.43; I2 71.7%, p<0.0001) and aOR 1.29 (95%CI 1.02-1.64; I2 36.3%, p=0.1). In this case, the meta-analyses appeared robust (i.e., remained significant) after removing these additional effect estimates from the SEOSI cohort.

###### Food insecurity

One study [Saeland 2014]^43^ reported two analyses of associations between food insecurity and risk for injecting-related infections. Participants who had a current injecting-related abscess reported fewer meals in the past 24 hours (median 2, IQR 1-3) compared to participants who did not have a current abscess (median 3, IQR 2-4). “Limited access to food” (not otherwise defined) was associated with an imprecise effect estimate (uOR 1.76; 95%CI 0.84 – 3.68) that could include meaningful differences in risk.

###### Health insurance

One study [Baltes 2002]^1^ assessed health insurance status among people with and without injecting-related SSTI, and estimated an imprecise effect size with a wide confidence interval that could include meaningful differences (uOR 0.79, 95%CI 0.18-3.56).

#### Substance use-related factors

###### Overdose history

Three studies^17,18,43^ report analyses of associations between overdose history and risk of injecting-related infections. Meta-analysis summary effect estimates for two unadjusted odds ratios was uOR 2.28 (95%CI 0.42-12.26; Figure 17). For two fully adjusted effect estimates, the summary was aOR 1.87 (95%CI 0.25-14.06; Figure 18).

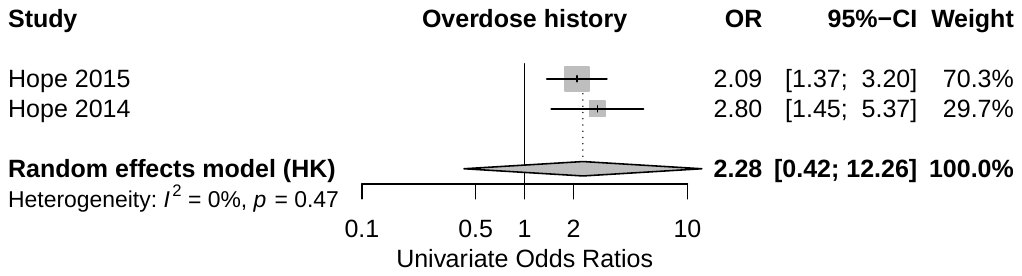
Figure 17. Meta-analysis of unadjusted effect estimates of relationship between history of overdose and incident or prevalent injection drug use-associated bacterial infections.

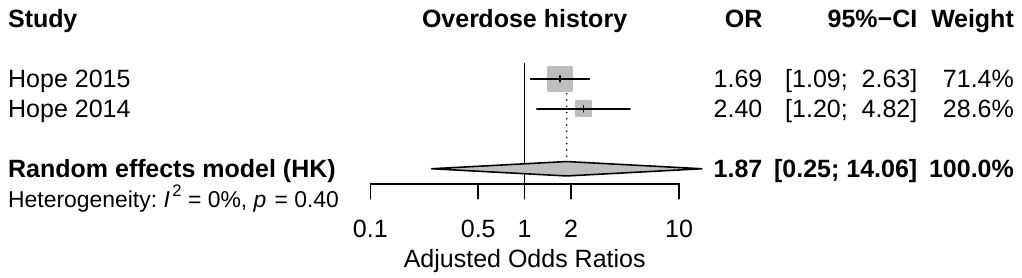
Figure 18. Meta-analysis of fully-adjusted effect estimates of relationship between history of overdose and incident or prevalent injection drug use-associated bacterial infections.

###### Heroin type/formulation

Three studies assessed associations between heroin formulation and risk of infection.^9,39,51^ One ecological study [Ciccarone 2016]^9^ across U.S. cities found that the proportion of opiate-related hospital admissions comprising skin and soft-tissue infections was 10.7% in Mexican “tar” heroin-dominant cities vs. 5.2% in Colombian “powder” heroin-dominant cities (p<0.001). Two studies [Pollini 2010; Summers 2017]^39,51^ assessed individual use of tar vs. powder heroin. Meta-analysis of two unadjusted effect estimates was uOR 7.44 (95%CI 0.31-176.41; Figure 19) and two fully adjusted effect estimates was aOR 3.65 (95%CI 0.0009-15058; Figure 20).

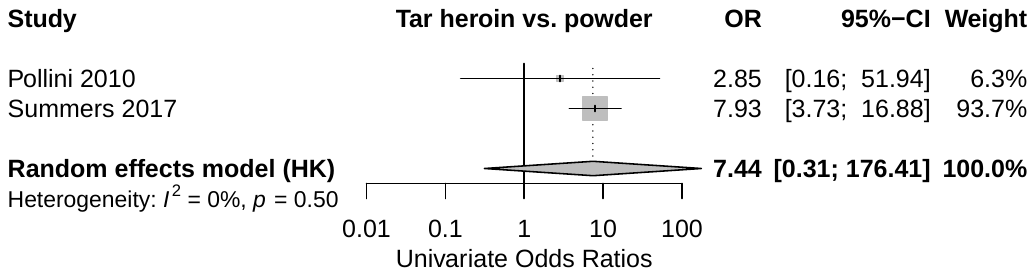
Figure 19. Meta-analysis of unadjusted effect estimates of relationship between heroin type (tar vs. powder) and incident or prevalent injection drug use-associated bacterial infections.

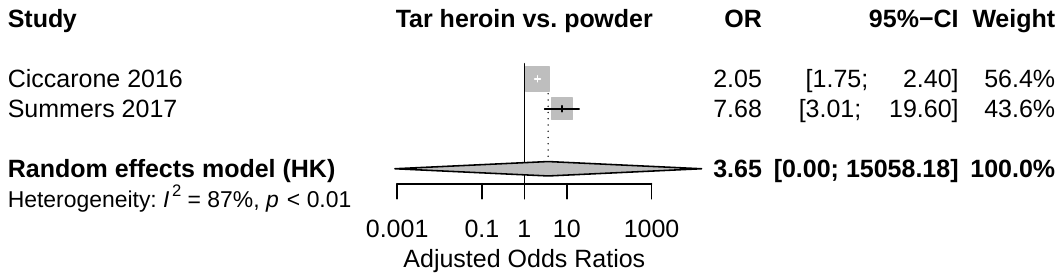
Figure 20. Meta-analysis of fully-adjusted effect estimates of relationship between heroin type (tar vs. powder) and incident or prevalent injection drug use-associated bacterial infections.

###### Heroin

Twenty-two studies assessed relationships between heroin use and risk of infections.^1,11,12,16–19,24–27,30,32,37–39,42,43,46,47,49,59^ Twenty-three unadjusted effect estimates were eligible for inclusion in meta-analysis, with summary effect estimate of uOR 1.34 (95%CI 1.13-1.61; Figure 21). There were three outliers [Sierra 2006; Milloy 2010; Phillips 2008].^30,37,47^ Removing these changed the summary to uOR 1.53 (1.29-1.82; I^2^ 66.2%, p<0.0001). Heroin use exposures were categorized differently across studies as “heroin is main drug”, “any heroin use” (vs. no heroin use), and “frequent heroin use” (e.g., at least daily; vs. less frequent use). Among these exposure subgroupings, only “any heroin use” (vs. no heroin use) had a statistically significant association with incident or prevalent injecting-related infections. “Heroin is main drug” and “heroin only” had imprecise effect estimates that could include meaningful differences.

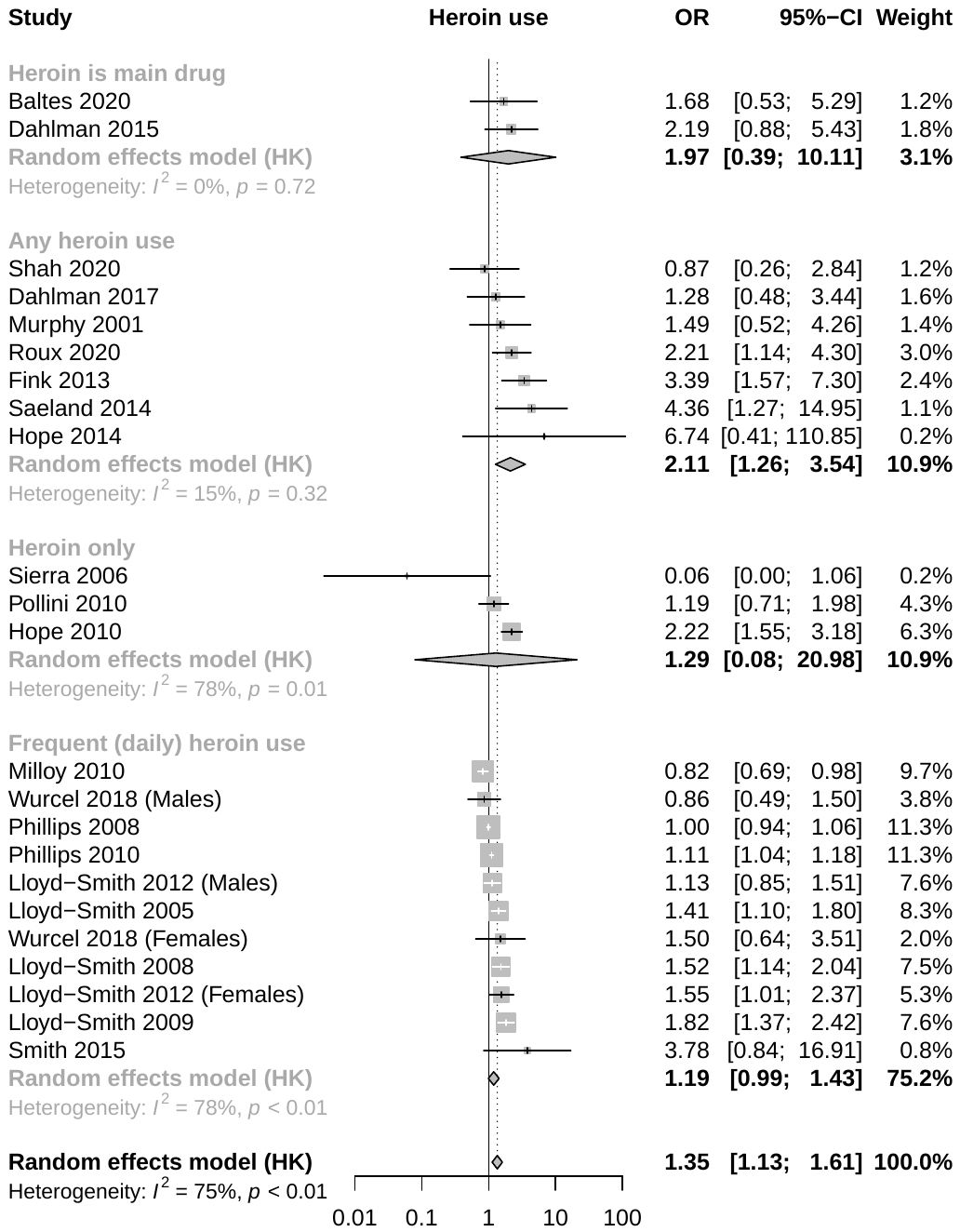
Figure 21. Meta-analysis of unadjusted effect estimates of relationship between heroin use and incident or prevalent injection drug use-associated bacterial infections.

Among seven adjusted effect estimates, summary was aOR 1.28 (0.95-1.74; Figure 22).

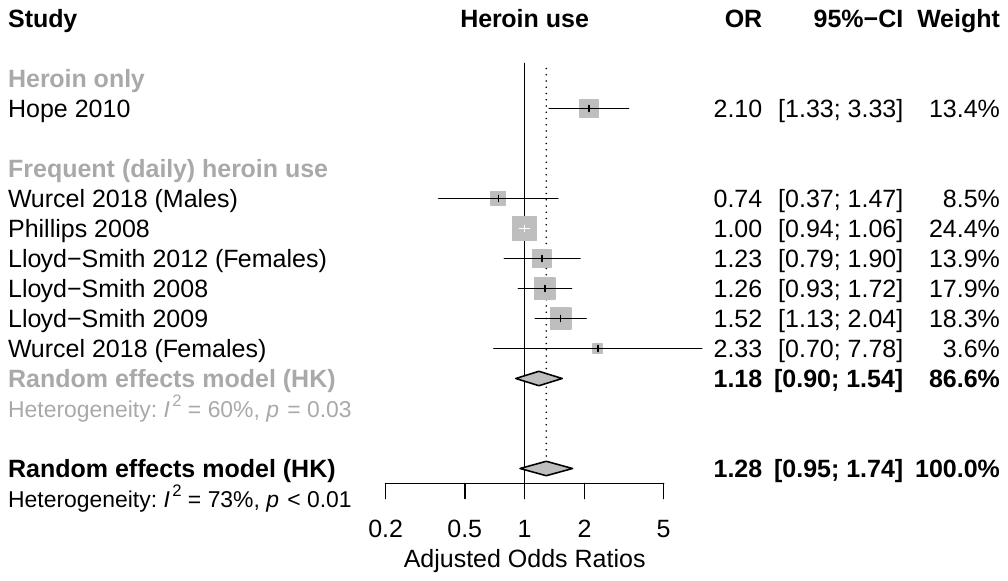
Figure 22. Meta-analysis of fully-adjusted effect estimates of relationship between heroin use and incident or prevalent injection drug use-associated bacterial infections.

###### Prescription opioids

Six studies^1,11,46,55,108^ assessed relationships between injecting prescription-type opioids (prescription fentanyl, oxycodone, hydromorphone, methadone, or buprenorphine) and development of injecting-related infections.

One study [Shah 2020]^46^ assessed injecting prescription fentanyl formulations and risk of endocarditis, in a case-control study. Fentanyl patch injection use (uOR 10.1, 95%CI 1.01–100.70) and fentanyl tablet injection use (uOR 0.76; 95%CI 0.08-7.10) were both associated with imprecise effect estimates, but fentanyl patch injection demonstrated statistically significantly elevated risk. We did not identify any studies assessing exposure to unregulated or illicit sources of fentanyl use and risk of infections.

The same study [Shah 2020]^46^ found no evidence of an association (with imprecise effect estimates) between prevalent injecting-related infections and injection use of oxycodone (uOR 0.83, 95%CI 0.36-1.90), controlled-release hydromorphone capsules (uOR 2.29, 95%CI 0.63-8.29), or immediate-release hydromorphone tablets (uOR 0.59, 95%CI 0.26-1.38). A second study [Silverman 2020]^48^ (among people with health care administrative codes consistent with injection drug use) found that compared with people who had recently filled prescriptions for opioids other than hydromorphone, people who filled a prescription for controlled-release hydromorphone had higher rates of injecting-related endocarditis (aOR 3.3, 95%CI 2.1-5.6) while people who filled a prescription for immediate-release hydromorphone did not (aOR 1.7, 95%CI 0.9-3.6).

There were no significant associations between injecting-related infections and injecting methadone identified in three studies [Baltes 2020; Dahlman 2015; Roux 2020]^1,11,42^, and injecting buprenorphine in one study [Dahlman 2015^11^].

###### Cocaine

Twenty-one studies assessed relationships between cocaine injecting and infections.^1,7,12,16–18,20,24–28,30,32,36,37,42,46,47,49,59^ Thirty unadjusted effect estimates were eligible for inclusion in meta-analysis, and the summary unadjusted effect estimate was uOR 1.29 (95%CI 1.10-1.51; Figure 23). When categorized by whether the study assessed “crack” cocaine (base) use or powder cocaine (hydrochloride) use, crack use was associated with excess risk (uOR 1.37, 95%CI 1.04-1.82) but powder cocaine was not (uOR 1.19; 95%CI 0.78-1.81). Note that these studies did not compare risks of crack vs. powder use, but instead asked participants whether they used crack or powder vs. did not use crack or powder cocaine. When sub-grouped by frequent (e.g. daily) use vs. less frequent use, summary was uOR 1.29 (95%CI 1.07-1.55); and when grouped by any use vs. no use, summary was uOR 1.26 (95%CI 0.92-1.71).

There were three outliers [Hope 2014 (Crack); Hope 2014 (Powder); Phillips 2008]^17,37^, and removing these changed the summary uOR to 1.42 (95%CI 1.25-1.61; I^2^ 49.3%, p=0.003). The updated summary for the crack cocaine subgroup was uOR 1.55 (95%CI 1.33-1.80; I^2^ 0%, p=0.56) and powder cocaine subgroup was uOR 1.35 (95%CI 1.00-1.83; I^2^ 46%, p=0.09).

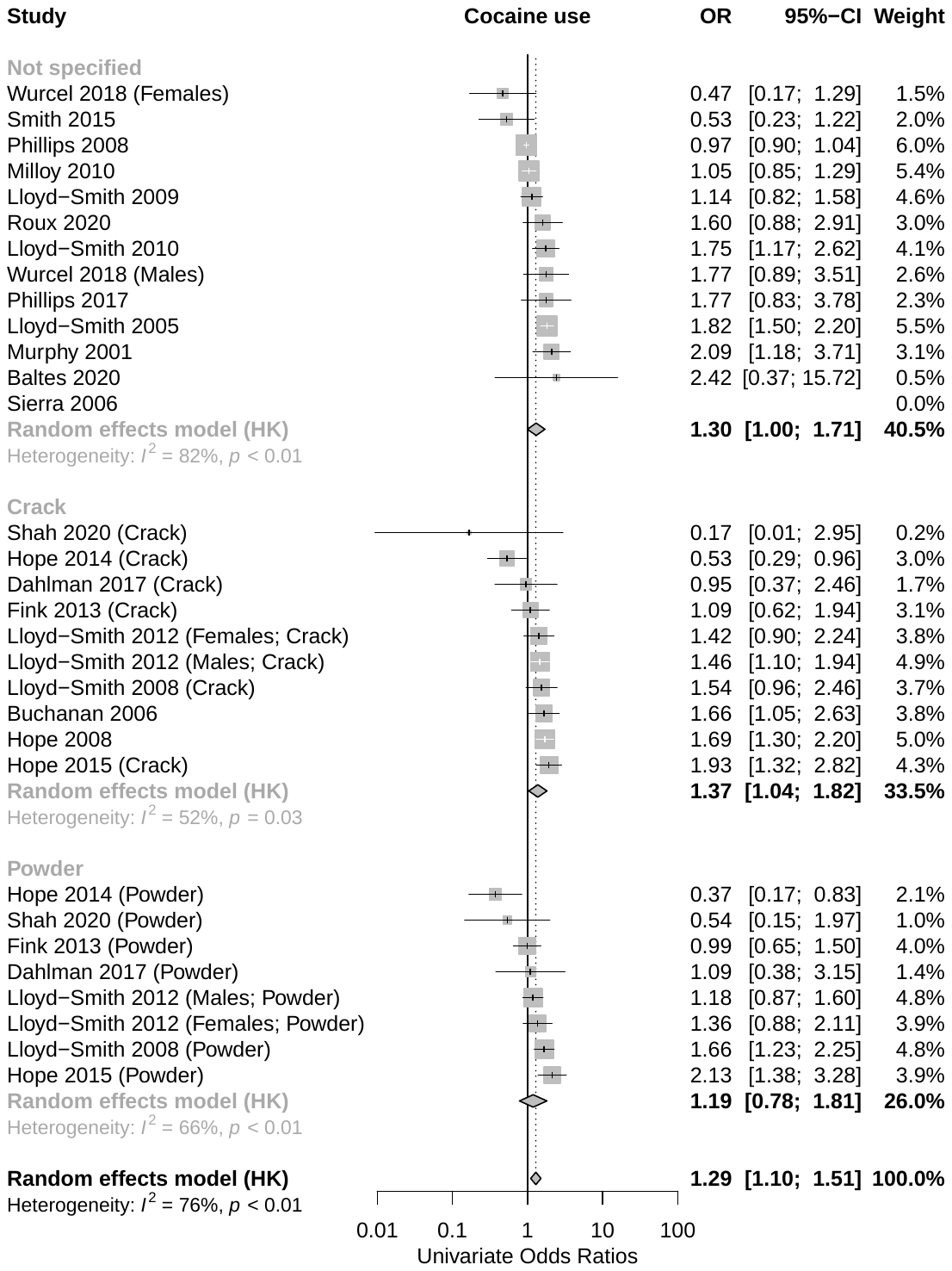
Figure 23. Meta-analysis of unadjusted effect estimates of relationship between cocaine and incident or prevalent injection drug use-associated bacterial infections. Subgroups by whether study assessed "crack" or "powder" forumluations of cocaine, or did not specify.

Meta-analytic summary for 10 adjusted effect estimates was aOR 1.31 (95%CI 1.02–1.69; Figure 24). When separated by frequency of use, frequent (daily) use vs. less-often was aOR 1.27 (95%CI 0.82-1.99; I2 80%, p<0.01) and any use vs. no use was aOR 1.41 (95%CI 0.92-2.15; I2 17.1%, p=0.3).

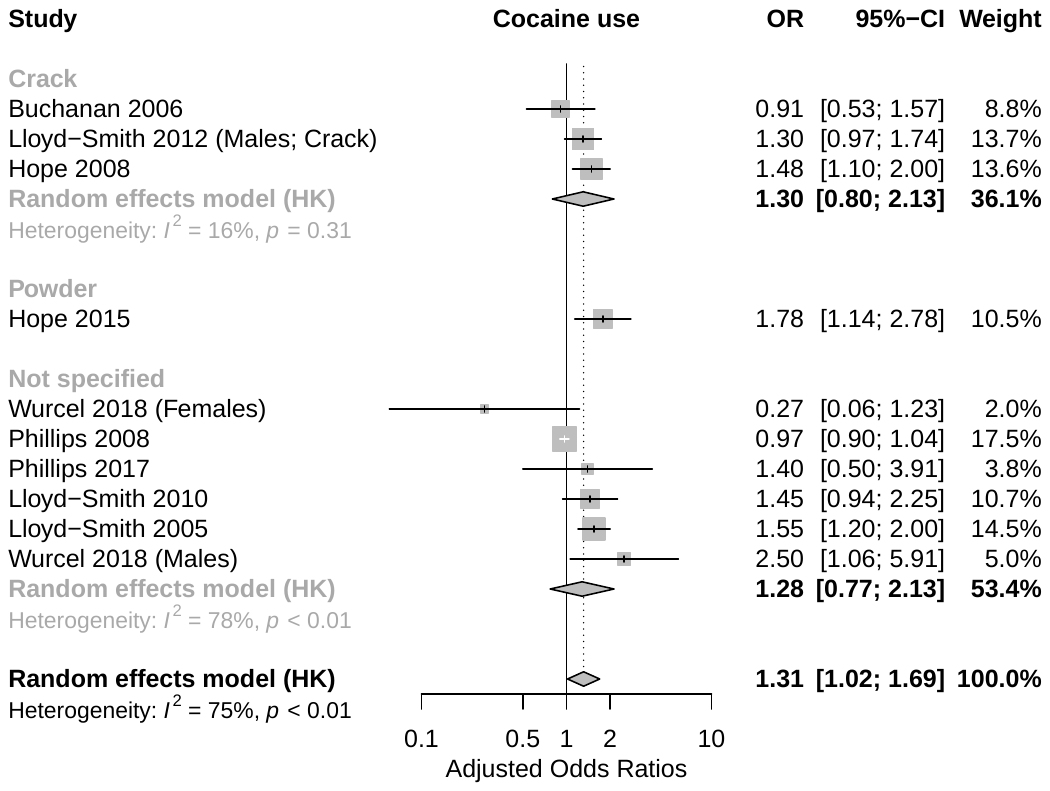
Figure 24. Meta-analysis of fully-adjusted effect estimates of relationship between cocaine and incident or prevalent injection drug use-associated bacterial infections. Subgroups by whether study assessed "crack" or "powder" formulations of cocaine, or did not specify.

###### Amphetamines (including methamphetamine)

Fourteen studies assessed unregulated/illicit amphetamines (including methamphetamine) and incident or prevalent injecting-related infections.^1,12,14,16–18,20,26,29,32,34,39,43,46^ Twelve unadjusted effect estimates were eligible for inclusion in meta-analysis, with summary uOR 0.81 (95%CI 0.49-1.33; Figure 25). One study [Murphy 2001]^32^ was an outlier; removing this changed summary to uOR 0.90 (95%CI 0.54-1.48; I^2^ 82.4%, p<0.0001).

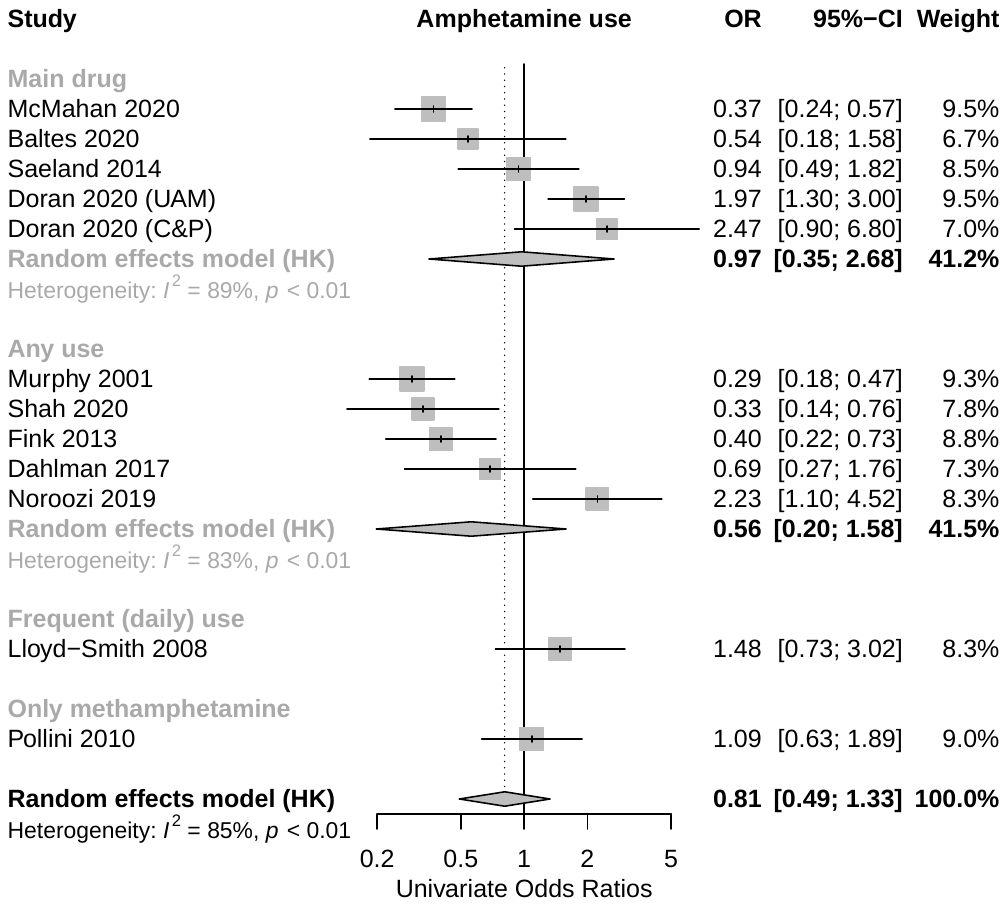
Figure 25. Meta-analysis of unadjusted effect estimates of relationship between amphetamines (including methamphetamine) and incident or prevalent injection drug use-associated bacterial infections.

Two fully-adjusted effect estimates were eligible for inclusion in meta-analysis, with aOR 1.74 (1.39-2.23; Figure 26).

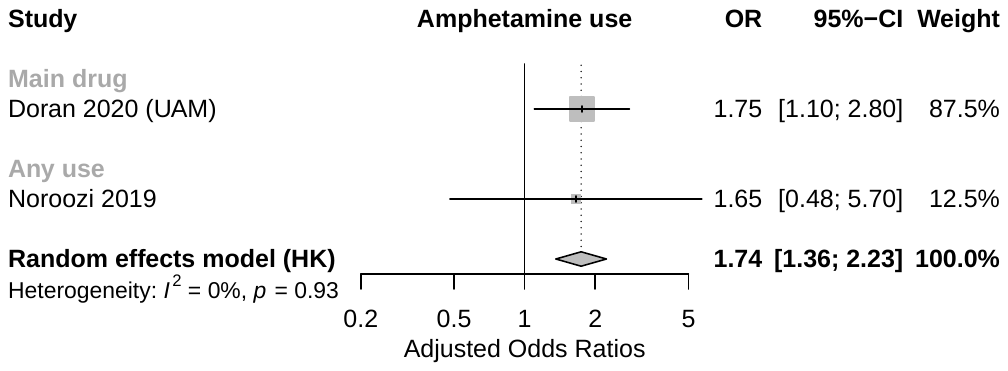
Figure 26. Meta-analysis of fully-adjusted effect estimates of relationship between amphetamines (including methamphetamine) and incident or prevalent injection drug use-associated bacterial infections.

One study [Pollini 2010]^39^ assessed colour of methamphetamine and past 6-months abscess. For people who used “clear” methamphetamine vs. “other”, the effect estimate was uOR 1.37 (95%CI 0.86-2.19).

###### Prescription stimulants

Two studies [Dahlman 2015; Shah 2020]^11,46^ included assessments of relationship between injecting prescription stimulants and injecting-related infections, and did not find evidence of an association with injecting methylphenidate (summary uOR 0.73, 95%CI 0.19-2.82, Figure 27) or bupropion (n=1 study; uOR was infinity, as only one participant reported bupropion injecting and they developed an infection).

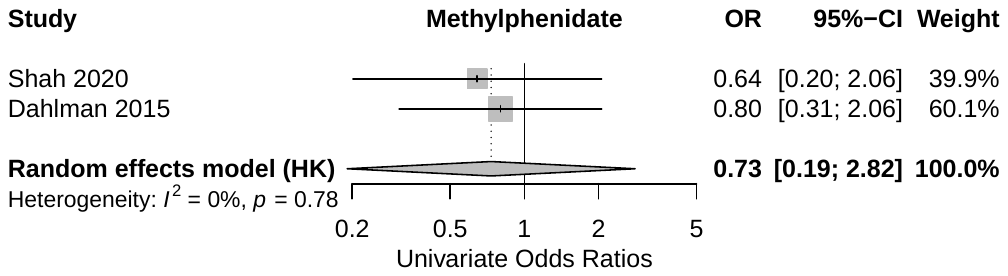
Figure 27. Meta-analysis of unadjusted effect estimates of relationship between methylphenidate injecting and incident or prevalent injection drug use-associated bacterial infections.

###### Novel psychoactive stimulants

Yeung 2017^60^ found that people self-reporting use of ethylphenidate (a novel psychoactive stimulant associated with high frequency of injecting) had higher weekly rates of S. pyogenes or S. aureus infections compared to people who did not report use of ethylphenidate (aRR 1.81, 95%CI 1.12-2.93).

###### Speedball (cocaine and heroin together) and goofball (methamphetamine and heroin together)

Nine studies assessed use of speedball or goofballs and injecting-related infections.^12,16,25,27,28,30,32,38,49^ Twelve eligible unadjusted effect estimates had summary uOR 1.34 (95%CI 1.15-1.57; Figure 28). When separated, goofball (n=2; uOR 1.22, 95%CI 0.98-1.52) and speedball (n=10; uOR 1.38, 95%CI 1.14-1.67) summary estimates had similar point estimates. Two fully adjusted effect estimates for speedball use had summary aOR 1.35 (95%CI 0.38-4.77; I^2^ 0.0%, p=0.6).

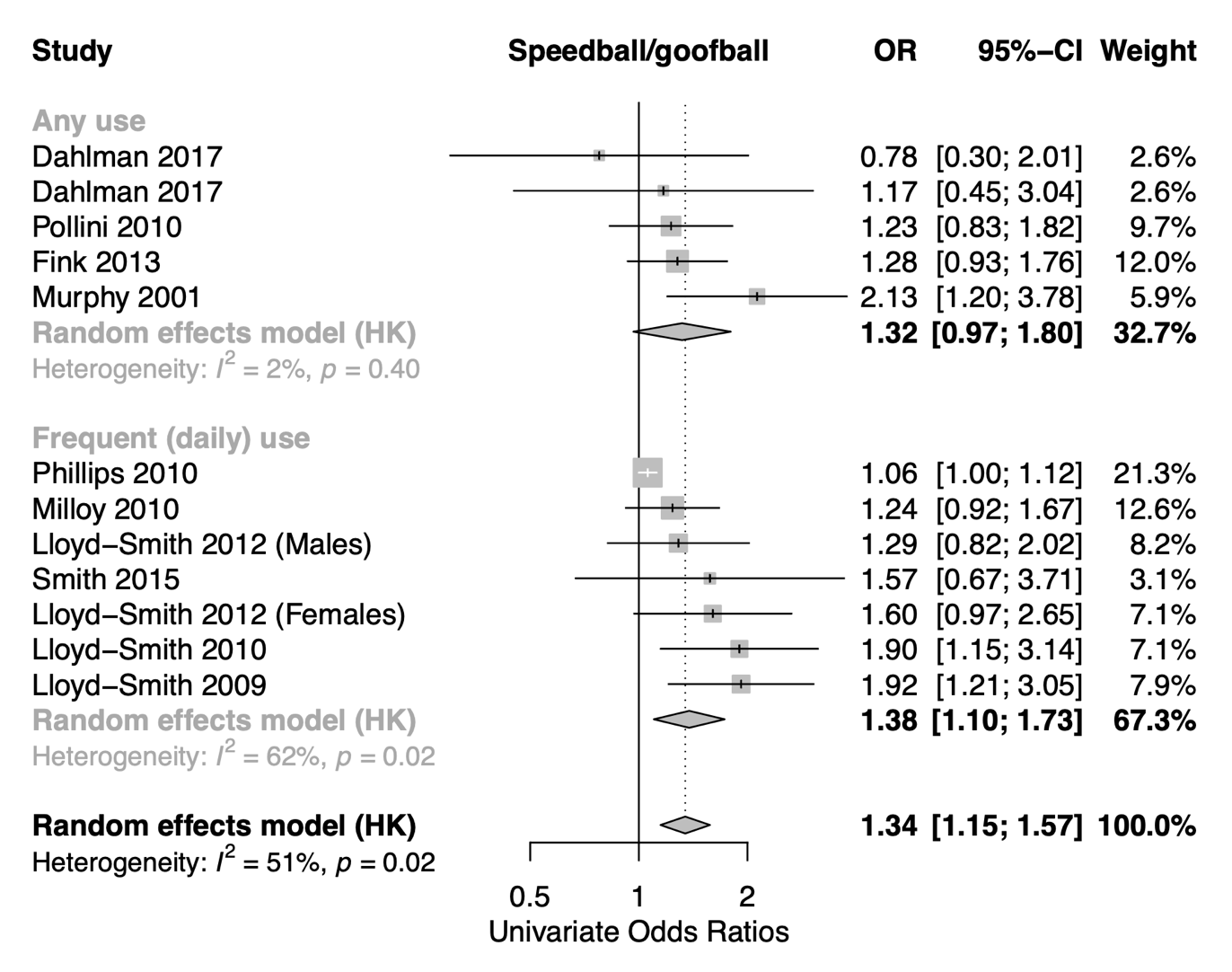
Figure 28. Meta-analysis of unadjusted effect estimates of relationship between speedball/goofball injection and incident or prevalent injection drug use-associated bacterial infections.

###### Alcohol use

Seven studies assessed relationships between alcohol use and incident or prevalent infections.^15,32,36,37,42,44,56^ When seven eligible unadjusted effect estimates were combined in meta-analysis, summary uOR was 0.94 (95%CI 0.77-1.14; Figure 29). Alcohol use exposures included “any” use or measures of hazardous alcohol use (e.g., Alcohol Use Disorders Identification Test [AUDIT]). Three eligible fully-adjusted effect estimates were combined in meta-analysis, with summary aOR 0.59 (95%CI 0.13-2.67; Figure 30). All of these studies assessed measures of hazardous alcohol use.

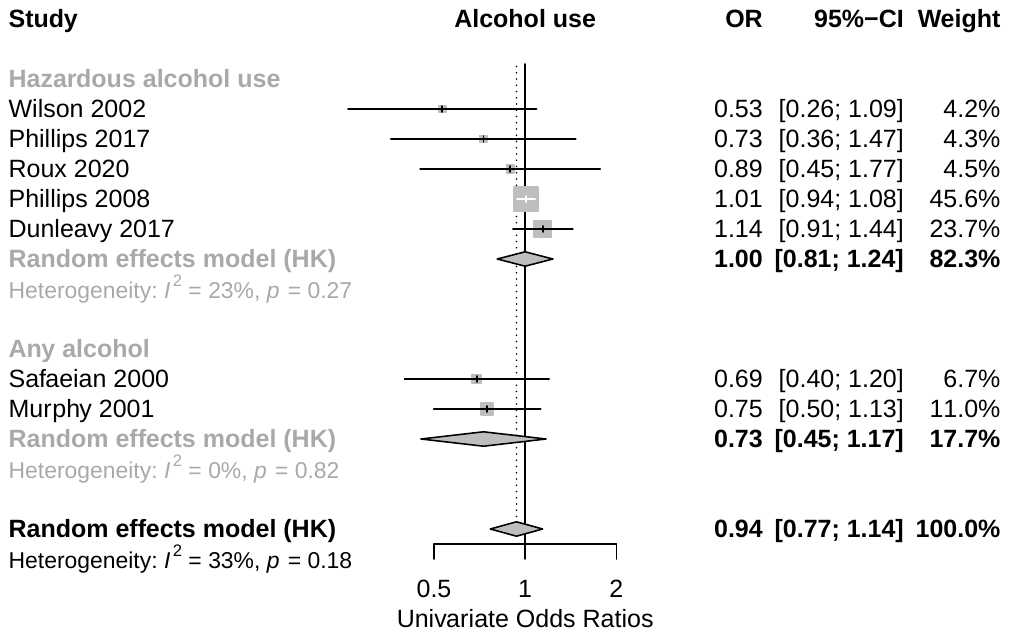
Figure 29. Meta-analysis of unadjusted effect estimates of relationship between alcohol use and incident or prevalent injection drug use-associated bacterial infections.

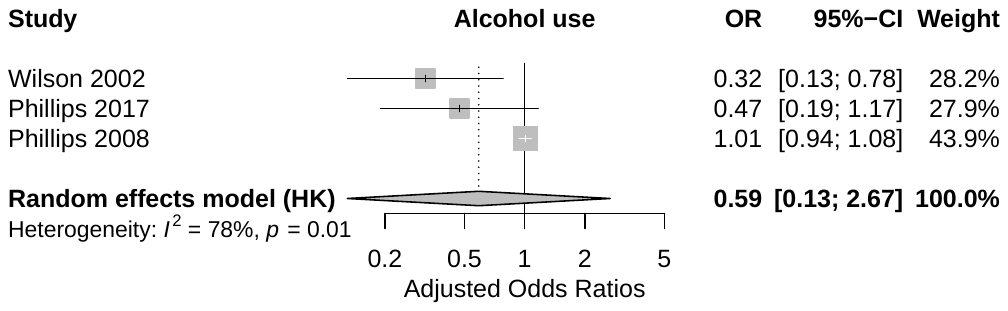
Figure 30. Meta-analysis of fully-adjusted effect estimates of relationship between alcohol use and incident or prevalent injection drug use-associated bacterial infections.

###### Smoking

Four studies assessed relationships between smoking substances and incident or prevalent injecting-related infections. Three studies included unadjusted effect estimates for cigarette smoking and injecting-related infections. Murphy 2001 (uOR 1.73; 95%CI 0.79-3.79) and Safaeian 2000 (uOR 0.6; 95%CI 0.3-1.2) did not find evidence of an association between smoking with abscess and endocarditis, respectively. In the same sample, Safaeian 2000 found an association between cigarette smoking and abscess (aOR 1.8; 95%CI 1.1-3.2).

Pollini 2010 found people who smoked methamphetamine were more likely to report having had an abscess (aOR 1.65; 95%CI 1.05-2.62), and Saeland 2014 did not find evidence of a relationship between smoking cannabis and current abscess (uOR 1.16; 95%CI 0.60-2.25).

#### Drug policy and injecting context

###### Drug policy change

Four studies assessed the impact of drug policy changes on risk of injecting-related infections.^13,33,55,60^ DiGiorgio 2019^13^ and Nagar 2015^33^ assessed the impact of state-wide opioid prescribing restrictions on rates of hospital admissions for injection drug use-associated spinal epidural abscess in Louisiana and Kentucky, respectively. In both states, increases were seen after state-wide restrictions on opioid prescribing – both thought to be due to people switching from prescription oral pills to injecting heroin when they could no longer access prescription tablets. Weir 2019^55^ found no relationship between delisting of extended-release oxycodone in Ontario and the proportion of hospital admissions for endocarditis attributable to injection drug use. Yeung 2017^60^ found no evidence of an immediate step-change (relative risk [RR] 1.11, 95%CI 0.46-2.70) but a gradual trend reduction (RR 0.88, 95%CI 0.82–0.94) in the weekly rate of *S. pyogenes* and *S. aureus* infections associated with injection drug use in Lothian, Scotland, after a temporary class order on ethylphenidate (a novel psychoactive stimulant associated with high frequency of injecting).

###### Drug purchasing network

One case-control study [Sierra 2006]^47^ assessed drug purchasing networks as part of an outbreak investigation into invasive *S. pyogenes* infections in Barcelona. Exposures including purchasing from one drug seller who was likely colonized (uOR 73, 95%CI 8-3090) and purchasing drugs from one physical location where this drug seller worked (uOR 34, 95%CI 7-175).

###### Injecting in public

I identified four studies reporting analyses of associations between public injecting and risk of injecting-related bacterial infections.^12,30,42,54^ The summary odds ratio for four unadjusted effect estimates was uOR 1.54 (95%CI 1.27-1.86; Figure 31) and for two fully adjusted effect estimates was aOR 1.40 (95% CI 1.05–1.88; Figure 32).

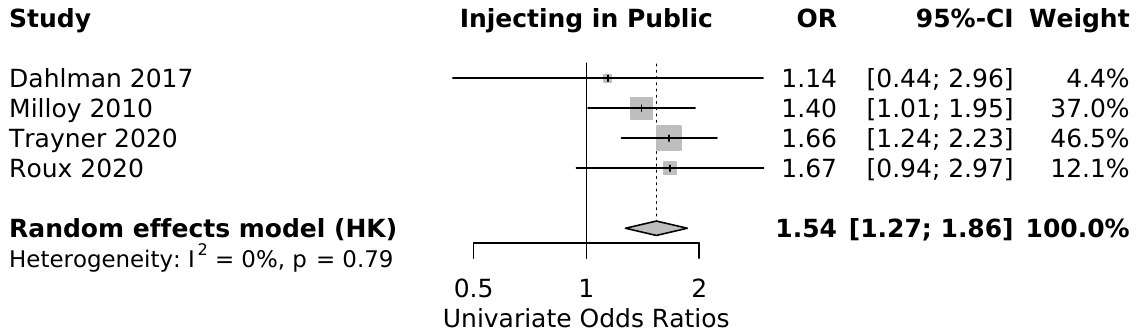
Figure 31. Meta-analysis of unadjusted effect estimates of relationship between public injecting and incident or prevalent injection drug use-associated bacterial infections.

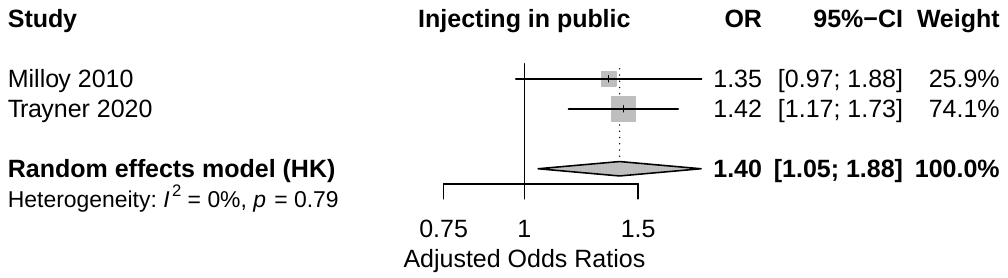
Figure 32. Meta-analysis of fully-adjusted effect estimates of relationship between public injecting and incident or prevalent injection drug use-associated bacterial infections.

###### Shooting galleries

Two studies assessed associations between use of “shooting galleries” (central locations where people can rent or borrow needles and syringes, and inject) and risk for injecting-related infections.^37,39^ Meta-analysis summary for two unadjusted effect estimates was uOR 0.84 (95%CI 0.17–4.02; Figure 33). One fully adjusted effect estimate from Phillips 2008^37^ was aOR 1.33 (95%CI 0.31–5.73).

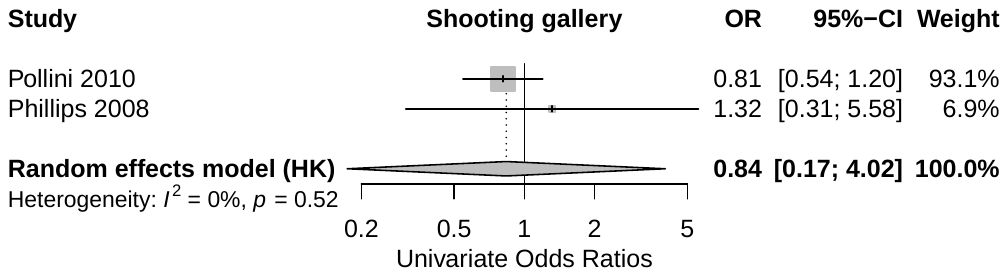
Figure 33. Meta-analysis of unadjusted effect estimates of relationship between "shooting gallery" use and incident or prevalent injection drug use-associated bacterial infections.

###### Police contacts and arrests

I identified four studies reporting analyses of associations between policing contacts (including arrests) and incident injecting-related bacterial infections.^10,17,18,39^ The meta-analytic summary for nine unadjusted effect estimates was uOR 1.16 (95%CI 0.85-1.59; Figure 34) and for four fully adjusted effect estimates was aOR 1.19 (95%CI 0.61-2.31; Figure 35).

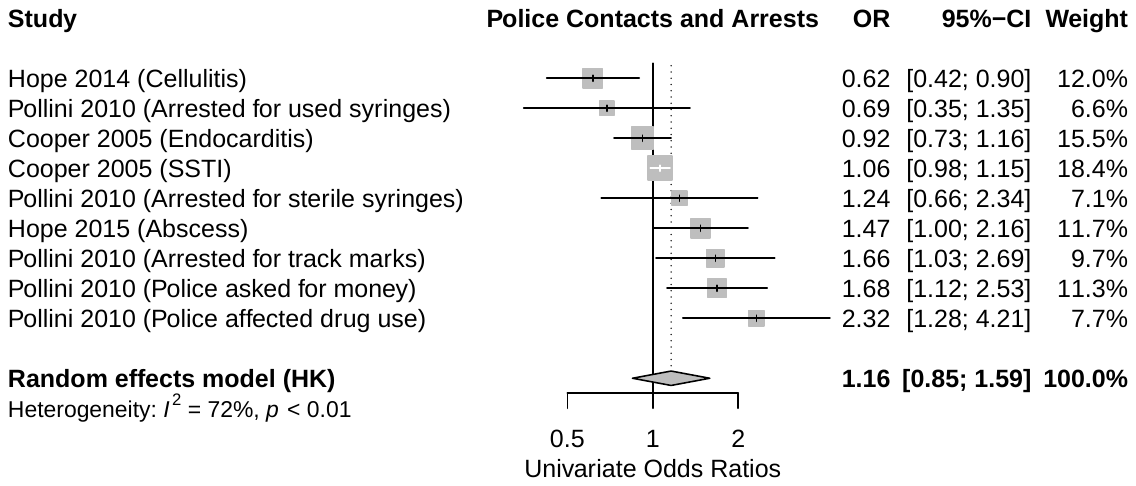
Figure 34. Meta-analysis of unadjusted effect estimates of relationship between police contacts and incident or prevalent injection drug use-associated bacterial infections.

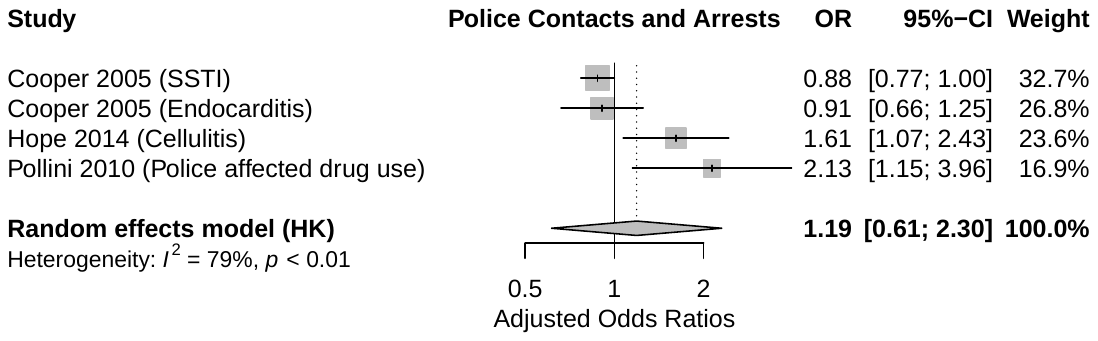
Figure 35. Meta-analysis of fully-adjusted effect estimates of relationship between police contacts and incident or prevalent injection drug use-associated bacterial infections.

Since Cooper 2005^10^ is an ecological study design and does not include exposure information for individual participants, we conducted a sensitivity analysis excluding Cooper 2005. It made no meaningful difference to the summary effect estimate for unadjusted (uOR 1.26, 95%CI 0.80–1.98; I2 75.8%, p=0.0004) or adjusted analyses (aOR 1.76, 95%CI 0.34-9.06; I2 0%, p=0.5).

###### Assisted injecting, or requiring help to inject

I identified eight studies^12,22,25–27,39,41,42^ reporting analyses of associations between requiring (or receiving) injecting assistance and risk for injecting-related infections. There were eight unadjusted effect estimates and eight fully adjusted effect estimates for meta-analysis. Meta-analysis of eight unadjusted analyses results in uOR 2.09 (95%CI 1.61–2.71; Figure 36) and of eight covariate-adjusted analyses results in aOR 1.78 (95%CI 1.40–2.27; Figure 37).

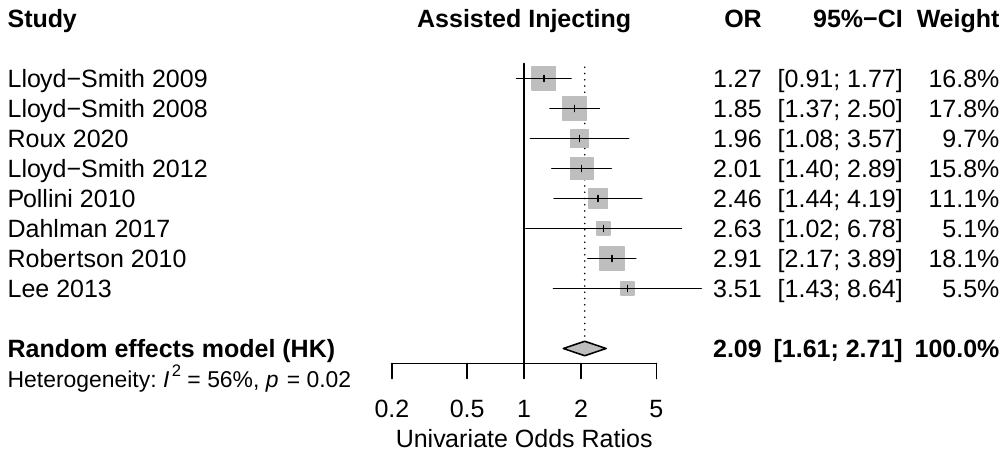
Figure 36. Meta-analysis of unadjusted effect estimates of relationship between requiring/receiving injecting assistance and incident or prevalent injection drug use-associated bacterial infections.

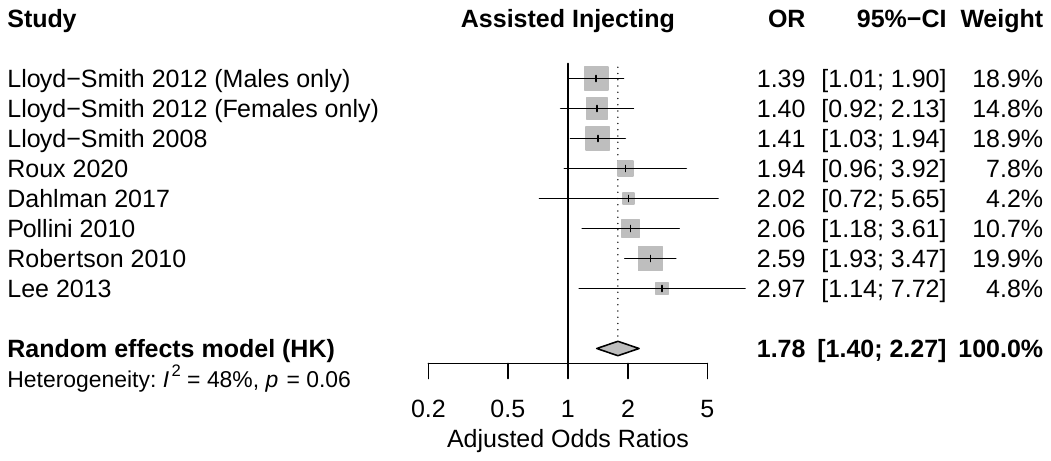
Figure 37. Meta-analysis of fully-adjusted effect estimates of relationship between requiring/receiving injecting assistance and incident or prevalent injection drug use-associated bacterial infections.

###### Injecting with others

Three studies assessed the impact of injecting with others or in groups, on infection risk.^39,42,49^ Seven eligible unadjusted effect estimates resulted in summary uOR 1.44 (95%CI 0.88-2.34; Figure 38). Two fully adjusted effect estimates, both from Smith 2015^49^, assessed whether a participant injects with friends (aOR 1.65, 95%CI 0.42-6.47) or with family/spouse (aOR 4.05, 95%CI 0.99-16.58). Summary aOR was 2.55 (95%CI 0.008-769.91; I^2^ 0.0%, p=1.00).

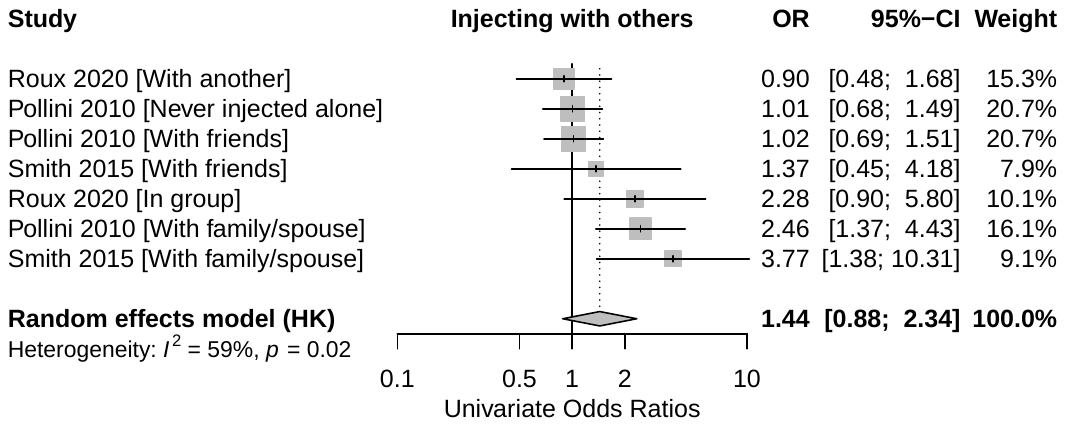
Figure 38. Meta-analysis of unadjusted effect estimates of relationship between injecting in groups and incident or prevalent injection drug use-associated bacterial infections.

#### Harm reduction and drug treatment

###### Needle and syringe distribution programs

I identified eight studies reporting analyses of associations between use of needle and syringe programs and risk of injecting-related bacterial infections.^5,6,15,16,19,34,53,59^ There were eight eligible unadjusted effect estimates, with a summary uOR 0.85 (95%CI 0.58-1.25; Figure 39). There were seven fully adjusted effect estimates, with summary aOR 0.84 (95%CI 0.53-1.34; Figure 40). Hope 2010 was an outlier for both analyses. Removing Hope 2010 changed the summary estimates to uOR 0.75 (95% 0.54-1.05; I^2^ 63.9%, p=0.01)) and aOR 0.75 (95% CI 0.54-1.03; I^2^ 47.1%, p=0.09).

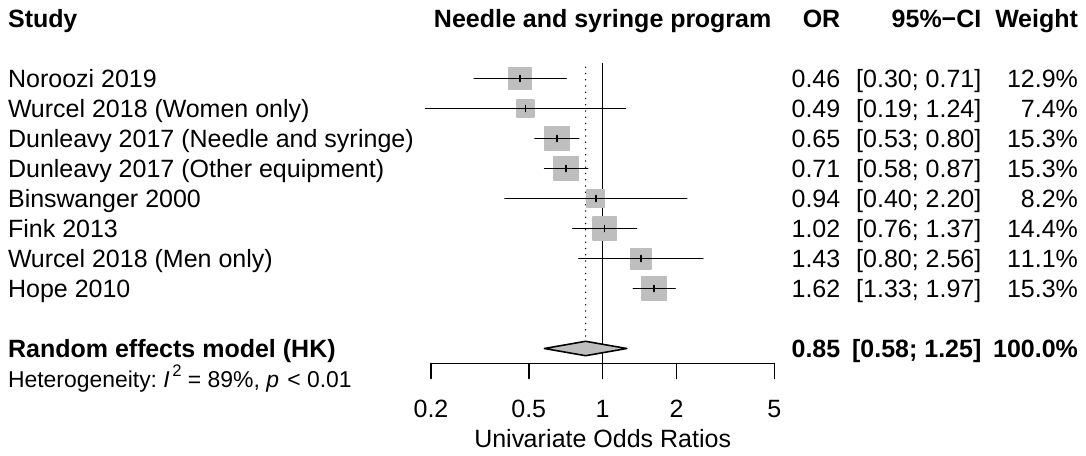
Figure 39. Meta-analysis of unadjusted effect estimates of relationship between needle/syringe program use and incident or prevalent injection drug use-associated bacterial infections.

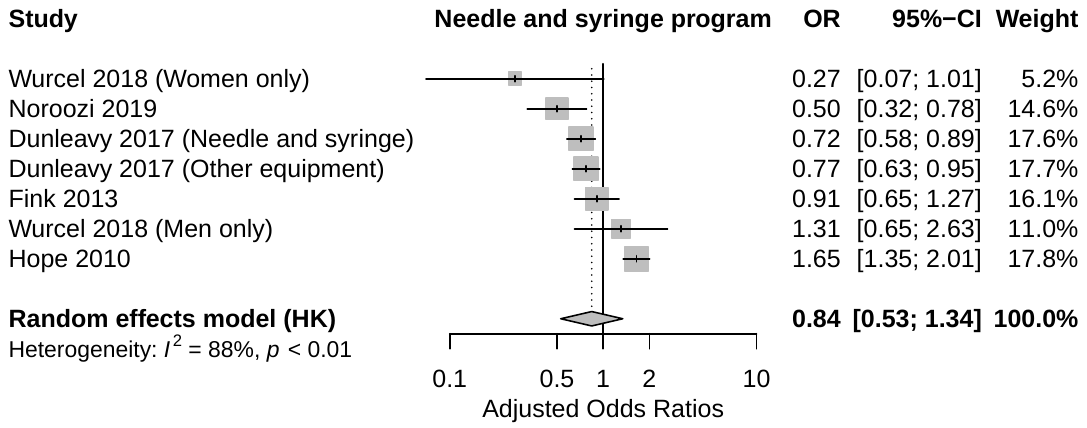
Figure 40. Meta-analysis of fully-adjusted effect estimates of relationship between needle/syringe program use and incident or prevalent injection drug use-associated bacterial infections.

One study [Bhattacharya 2006]^5^ was an ecological study assessing the proportion of local people who inject drugs with injection-site abscesses before and after the implementation of a needle and syringe program. Abscess prevalence was 23% in the month before implementation and declined to very low levels (at times 0%; e.g. one year after implementation). There was no statistical analysis or reported frequencies, so we could not include this in meta-analysis.

Another study [Tomolillo 2007]^53^ included an ecological analysis, correlating the weekly number of abscesses treated at a community clinic with the activity of the associated and co-located needle exchange program. A time series analysis identified “significant negative relationships” between the number of abscesses treated and both the number of needles exchanged (b = -0.001, p=0.002) and the number of needle exchange program visits (b = -0.12m p<0.001) per week. The authors report unstandardized coefficients from their regression model but do not provide any other detail on the model methods. We could not include these in meta-analysis. It also reports a second individual-level study correlating self-reported abscesses with number of needles exchanged (R2 = 0.10, p=0.01) and “more use of sterile equipment” (R2 = 0.10, p=0.10), with no timelines or frequencies reported. We also could not include this in meta-analysis. Finally, Tomolillo and co-authors describe the average weekly rate of abscesses treated at the community clinic before and after the implementation of a policy restricting the number of needles and syringes to be distributed to each client and requiring pre-arranged appointments for access. The average weekly number of needles distributed decreased from mean (SD) 3268 (965) needles to 471 (321) needles, and the average weekly number of abscesses treated at the community clinic increased from 8.5 (3.2) to 14.3 (6.0). No statistics or frequencies are reported for us to calculate.

One additional study [Pollini 2010b]^40^ assessed relationships between being refused/overcharged syringes when trying to purchase at a pharmacy in Tijuana, Mexico. There were imprecise effect estimates for having an abscess in the past 6 months (uOR 0.97, 95%CI 0.57–1.65), nor with ever having had an abscess (uOR 1.10, 95%CI 0.71–1.69). There was a small positive associated with average number of life abscesses (aOR 1.02, 95%CI 1.00–1.03).

###### Opioid agonist treatment

I identified 13 studies reporting analyses of associations between opioid agonist treatment use and risk for injecting-related infections.^2–4,15,19,20,30,35,42,47,50,52^ Eleven unadjusted effect estimates and ten multivariable effect estimates for meta-analysis. Only four studies provided both, and otherwise unadjusted and adjusted effect estimates were from separate studies. Only two of the studies [Bassetti 2002; Milloy 2010] assessed exposure to opioid agonist treatment and development of injecting related infections over the same time period; otherwise, the timelines did not align, which may represent misclassification bias and undermines our ability to infer any causal relationship. For example, Betts 2016 assessed currently taking opioid agonist treatment in relation to risk of SSTI in the past month; Dunleavy 2017 assessed current opioid agonist treatment use in relation to risk of SSTI in the past year.

Summary effect estimates for 11 univariable effect estimates was uOR 0.75 (95%CI 0.63-0.89; Figure 41). One study [Hope 2010]^19^ was an outlier. Removing this changed the summary effect estimate to uOR 0.71 (95%CI 0.62-0.81; I^2^ 18.1%, p=0.28).

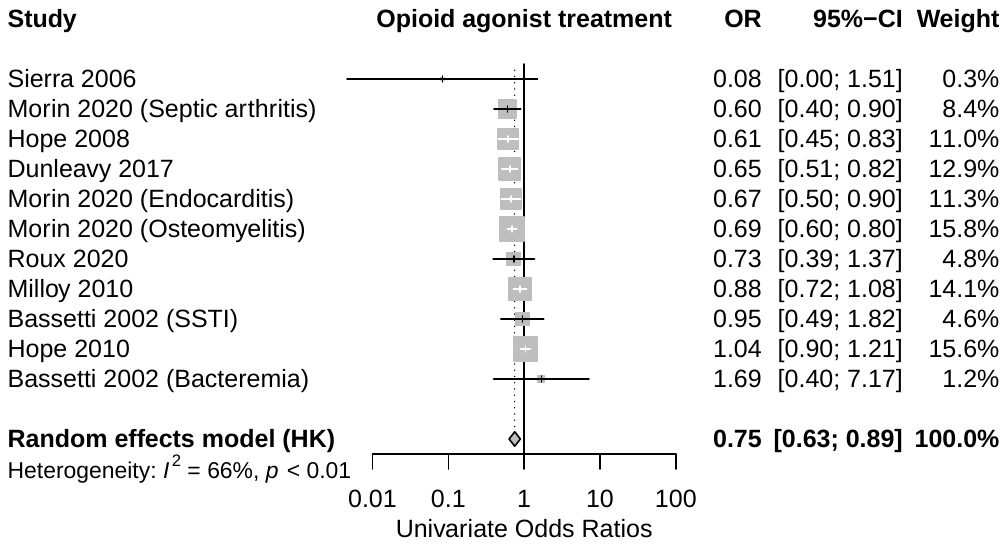
Figure 41. Meta-analysis of unadjusted effect estimates of relationship between opioid agonist treatment (OAT) use and incident or prevalent injection drug use-associated bacterial infections.

Summary effect estimate for 10 multivariable adjusted analyses was aOR 0.92 (95%CI 0.86–0.97; Figure 42). One study [Hope 2008]^20^ was an outlier; removing this changed summary aOR to 0.92 (95%CI 0.89-0.95; I^2^ 50.2%, p=0.04)

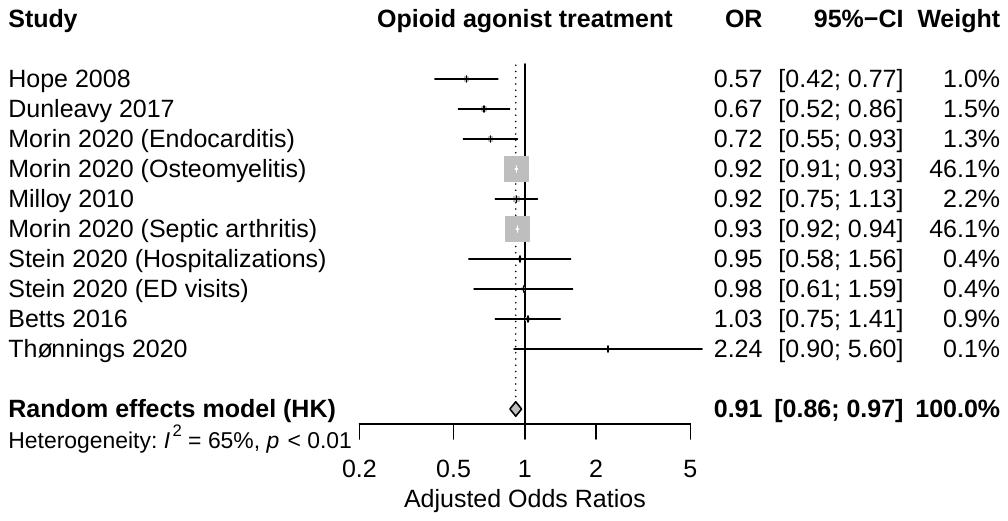
Figure 42. Meta-analysis of fully-adjusted effect estimates of relationship between opioid agonist treatment (OAT) use and incident or prevalent injection drug use-associated bacterial infections.

Two studies [Bertin 2020; Oviedo-Joekes 2017] compared multiple types of opioid agonist treatment, rather than any vs. no opioid agonist treatment. Bertin 2020 compared incidence of hospitalization with injecting-related bacterial infections amongst patients prescribed first-line treatment with buprenorphine or methadone, and second-line/alternative treatment with morphine sulfate. The crude incidence per 100,000 person-years in the buprenorphine group was 2.2 (95%CI 1.8-2.5), in the methadone group was 1.6 (95%CI 1.2-2.0), and in the morphine group was 7.0 (4.7-10.6). For patients receiving morphine, aHR was 2.8 (1.8-4.4) compared to buprenorphine and aHR 3.6 (2.2-5.9) compared to methadone. One study [Oviedo-Joekes 2017]^35^ reported rates of cellulitis or abscess as potential adverse effects within a randomized trial of injectable hydromorphone (7 episodes among 100 patients) vs. injectable diacetylmorphine (17 episodes among 102 patients; no statistical test reported).

###### Combined harm reduction interventions

I identified one study [Dunleavy 2017]^15^ assessing exposure of combined needle and syringe program and opioid agonist treatment use. “High” use (defined as currently prescribed opioid agonist treatment and >200% uptake of needles and syringes) was associated with reduced risk of past year SSTI (uOR 0.55, 95%CI 0.41–0.73; aOR 0.62, 95%CI 0.46–0.83).

###### Supervised consumption sites

I identified three studies reporting analyses of relationships between supervised consumption site use and risk for injecting-related infections.^26,30,45^ Meta-analysis of three unadjusted effect estimates resulted in summary uOR 0.74 (95%CI 0.17-3.26; Figure 43). Only one study [Lloyd-Smith 2008]^26^ reported an adjusted effect estimate (aOR 0.59, 95% 0.29-1.19). Overall, these summary effect estimates were imprecise and confidence intervals were wide enough to include potentially meaningful differences in risk.

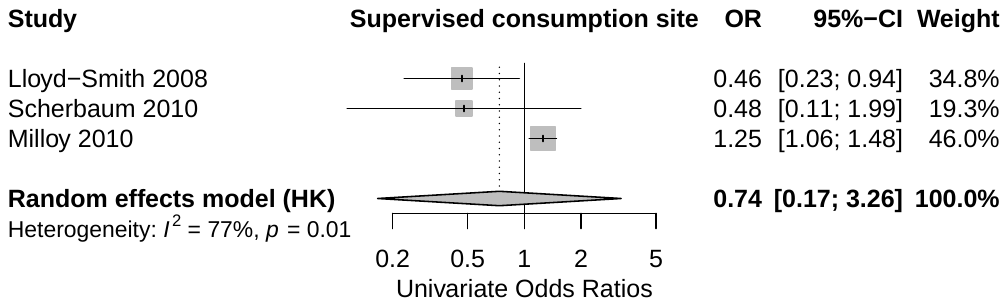
Figure 43. Meta-analysis of unadjusted effect estimates of relationship between supervised consumption site use and incident or prevalent injection drug use-associated bacterial infections.

#### Intersectionality

Few studies assessed the intersections of multiple identities or social positions in relation to risk of injecting-related bacterial infections. For example, no studies examined differential health risks across multiple intersections of race and gender categories. Several studies presented analyses stratified by gender-sex. In Wurcel 2018^59^, sex work was associated with ever having had an abscess among females but not males. In Smith 2015^49^, women with abscesses more often reported unstable housing (n=6, 40.0% versus n=6, 14.6%; P=0.119). In Wurcel 2016^58^, the investigators found that among U.S.-wide hospital admissions for endocarditis the proportion attributed to injection drug use increased especially quickly among younger white people (from 57.0% in 2000 to 80.3% in 2013, p<0.001). In Lloyd-Smith 2012, requiring injecting assistance was associated with increased risks of injecting-related infections among men and women in unadjusted analyses, but in fully-adjusted analyses this was only significant among men.

### Appendix 18. Synthesis and meta-analyses of studies with outcomes occurring during treatment for injecting-related infections

Included studies assessed several different outcomes that occurred during treatment for injecting-related bacterial infections:

(a) healthcare-seeking for injecting-related infection;
(b) self-treatment of abscess;
(c) hospital admissions among people with an injecting-related SSTI;
(d) premature hospital discharges against medical advice, among people hospitalized with injecting-related infections;
(e) new/secondary bloodstream infections among people receiving antibiotic treatment;
(f) in-hospital death; and
(g) other outcomes (development of endogenous endophthalmitis, and respiratory failure among people with botulism).

Table 2. Summary of outcomes and associated exposures assessed among studies where outcome occurs during treatment for injecting-related infections.

| **Outcomes** | **Exposures assessed** | **Number of studies** |
| --- | --- | --- |
| Health care-seeking for injecting-related infections | gender/sex; age; income/employment; sex work; unstable housing; incarceration; overdose history; migration status; heroin use; cocaine use; amphetamine use; opioid agonist treatment; supervised consumption site use | 4^18,20,25,28^ |
| Self-treatment of abscess | gender/sex; age; race/ethnicity; unstable housing; heroin use; cocaine use; needle and syringe programs; several measures of access to health care (e.g., having a primary care provider or having health insurance) | 2^18,78^ |
| Hospital admissions among people with an injecting-related SSTI | gender/sex; age; race/ethnicity; education; income/employment; sex work; migration status; unstable housing/homelessness; incarceration history; overdose history; heroin; cocaine; amphetamines; alcohol use; needle and syringe program use; access to health care (e.g., insurance, having a primary care provider); self-treatment of infections; hospital admission history | 2^18,78^ |
| Premature hospital discharges against medical advice, among people hospitalized with injecting-related infections | gender/sex; age; race/ethnicity; income/employment; unstable housing; overdose history; opioid use; cocaine; alcohol; other substance use; health care access; opioid agonist treatment; in-hospital addiction treatment; hospital characteristics; hospital policy; surgery during hospitalization | 10^61–64,66,70,72,73,76,77,81^ |
| New/secondary bloodstream infections among people receiving antibiotic treatment | gender/sex; age; unstable housing and homelessness; substance use (heroin, stimulants, polysubstance use, other); substance use treatment; insertion of peripherally-inserted intravenous central catheters (PICC lines) for parenteral antimicrobial treatment | 1^79^ |
| In-hospital death | gender/sex; age; race/ethnicity; overdose history; substance use (opioids, stimulants); health care access (insurance); hospital policies; surgery during hospital admission | 5^68–70,75,76^ |
| Development of endogenous endophthalmitis | gender/sex; race/ethnicity; alcohol use; infection of central venous catheter | 1^80^ |
| Respiratory failure among people with botulism | gender/sex; age | 1^74^ |

Subsections below are organized by outcome and then by each exposure assessed in association with that outcome. See Appendix 13 for a list of all extracted effect estimates in this section.

##### Healthcare-seeking for injecting-related infections

Four studies [Hope 2008; Hope 2015; Lloyd-Smith 2010; Lloyd-Smith 2012]^18,20,25,28^ assessed associations between seeking treatment for injecting-related infections (once they had developed) and the following exposures: gender/sex; age; income/employment; sex work; unstable housing; incarceration; overdose history; migration status; heroin use; crack and powder cocaine use; amphetamine use; opioid agonist treatment; and supervised consumption site use.

Results were mixed and none of these social-structural, substance use, or health services correlates were significantly associated with seeking treatment for injecting-related infections in more than one study (and for most “non-significant” associations, study authors did not report frequencies or statistics). Seeking medical advice about injection-site infections was associated with female sex in Hope 2015^18^ (aOR 3.04, 95%CI 1.14-8.13) but not in Hope 2008^20^ (no statistics provided). Recent incarceration was associated with decreased healthcare seeking in Hope 2015^18^ for cellulitis (uOR 0.55; 95%CI 0.34- 0.88), but there was no evidence an association for abscess in Hope 2015^18^ or for any SSTI in Hope 2008^20^. Seeking medical advice was not associated with age [Hope 2008^20^; Hope 2015^18^]; illicit/illegal work as main source of employment (uOR 1.50; 95%CI 0.99 – 2.78) [Hope 2015^18^]; sex work in the preceding year (uOR 0.45; 0.19-1.07) [Hope 2015^18^]; or unstable housing/homelessness [Hope 2008^20^; Hope 2015^18^].

Seeking medical advice for cellulitis was more likely in people who injected powder cocaine (aOR 2.37; 1.36-4.14, for cellulitis only) and crack cocaine (uOR 1.71; 1.12-2.63) in Hope 2015^18^ (vs. people who did not inject cocaine), but not for abscesses in Hope 2015 and not for SSTI in Hope 2008^20^. Advice-seeking was not associated with overdose history [Hope 2008^20^; Hope 2015^18^]; heroin use [Hope 2015^18^]; or amphetamine use [Hope 2008^20^; Hope 2015^18^].

In one study [Hope 2008^20^], people who never received opioid agonist treatment (aOR 0.3; 95%CI 0.1-0.7) and people who previously received opioid agonist treatment (aOR 0.5; 95%CI 0.3-0.9) were less likely to seek health care for an injecting-site infection than people currently receiving opioid agonist treatment. In Lloyd-Smith 2012^25^ and Lloyd-Smith 2010^28^, people who received a referral from a nurse at a supervised consumption site were more likely to have ED visit or hospital admission (respectively) for an injecting-related infection – however, these analyses were confounded as people with infections would be referred more often than people without infections.

##### Self-treating abscess

Two studies [Fink 2013; Monteiro 2020]^16,71^ assessed factors associated with self-treatment of an abscess: gender/sex; age; race/ethnicity; unstable housing; heroin use; cocaine use; needle and syringe programs; and several measures of access to health care.

Self-treating abscesses was not associated with most of these, but effect estimates tended to be imprecise with wide confidence intervals. The only statistically significant findings were that self-treating abscess was more common among Latino vs. Black participants in Fink 2013^16^ (aOR 2.83; 95%CI 1.65-5.10). Self-treating abscess was also less likely among people who reported having a “usual place” to access health care (aOR 0.61; 95%CI 0.40-0.92) [Fink 2013^16^], but self-treatment was not significantly associated with other measures of health care access (e.g., having a primary care provider or having health insurance). These analyses also had wide confidence intervals that could potentially include meaningful effects.

##### Hospital admission, among people presenting for healthcare with SSTI

Two studies [Hope 2015; Takahashi 2007]^18,78^ assessed associations between hospital admission (among people with injecting-related infections) and the following exposures: gender/sex; age; race/ethnicity; education; income/employment; sex work; migration status; unstable housing/homelessness; incarceration history; overdose history; heroin; cocaine; amphetamines; alcohol use; needle and syringe program use; access to health care (e.g., insurance, having a primary care provider); self-treatment of infections; and hospital admission history.

Almost all exposures (e.g., education, income/employment, sex work, incarceration history, health insurance, self-treatment of infections, and others) were not significantly associated with risks of hospital admission, often in the context of small sample sizes and imprecise effect estimates. Reporting two or more hospitalizations in the past year was significantly associated with hospital admission for SSTI in the one study in which it was assessed (aOR 4.4; 95%CI 1.6-11.8) [Takahashi 2007^78^].

The following exposures were inconsistently associated with hospital admission, between multiple studies. Older age was associated with hospital admission in one study [Hope 2015^18^] (for age 35 years or older, uOR 3.71, 95%CI 1.77-7.81) for only those with abscesses (not those with cellulitis), and was not significantly associated with hospital admission in another study [Takahashi 2007^78^]. Similarly, female sex was associated with decreased likelihood (uOR 0.40, 95%CI 0.17-0.96) only for abscess, but not cellulitis, in one study [Hope 2015^18^] and not in a second [Takahashi 2007^78^]. In one study [Takahashi 2007^7^], “living in a shelter” was associated with increased risk of hospital admission (aOR 4.2, 95%CI 1.2–15.1), but “living on the street” was not (aOR 1.4, 95%CI 0.5–4.1). Housing status was not significantly associated with the outcome in a second study [Hope 2015^2^]. In one study [Hope 2015^18^], injecting crack was associated with a large positive effect (aOR 7.49, 2.50-22.50) only for people with injection-site abscesses, and was not significantly associated for people reporting injection-site cellulitis.

##### Against medical advice discharge

Ten studies assessed relationships between social, substance use, and health services exposures and risks of premature hospital discharges against medical advice (among people admitted to hospital with an injecting-related infection).^61–64,66,70,72,73,76,77,81^ The exposures included gender/sex; age; race/ethnicity; income/employment; unstable housing; overdose history; opioid use; cocaine; alcohol; other substance use; health care access; opioid agonist treatment; in-hospital addiction treatment; hospital characteristics; hospital policy; surgery during hospitalization.

###### Gender/sex

Six studies assessed relationships between gender/sex and risk of premature hospital discharge against medical advice, among patients hospitalized with injecting-related infections.^62–64,70,72,76^ Meta-analysis of two unadjusted effect estimates resulted in summary uOR 2.34 (95%CI 0.90-6.10; Figure 44). Meta-analysis of six fully-adjusted effect estimate resulted in aOR 1.22 (95%CI 0.99-1.50; Figure 45).

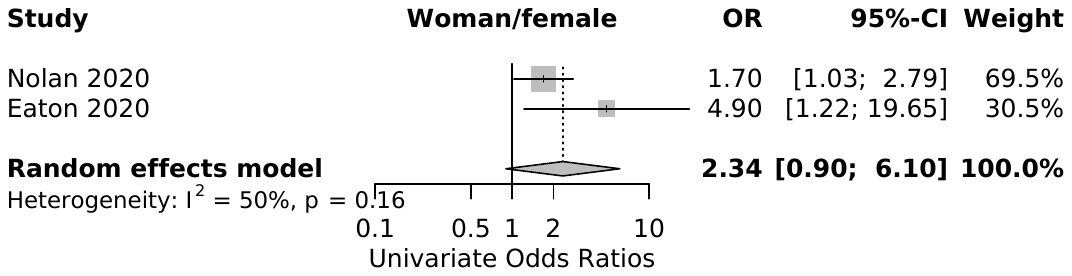

Figure 44. Meta-analysis of unadjusted effect estimates of relationship between woman/female gender/sex and premature hospital discharge against medical advice, among people hospitalized with injection drug use-associated bacterial infections.

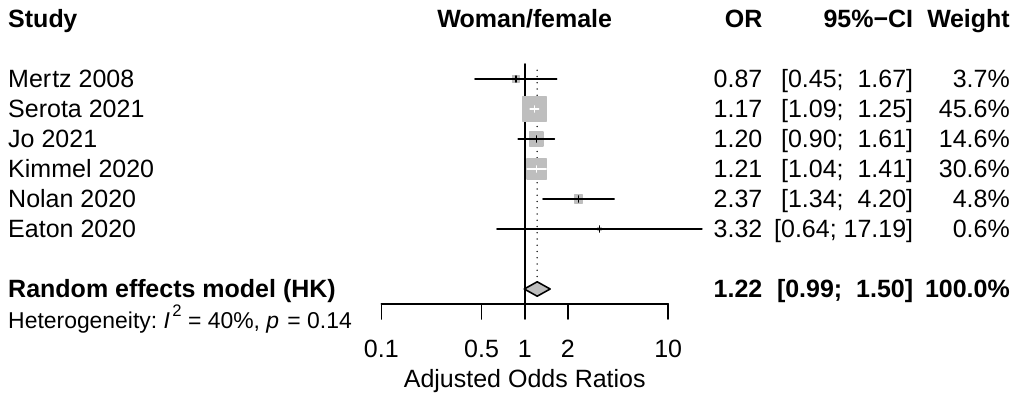

Figure 45. Meta-analysis of fully-adjusted effect estimates of relationship between woman/female gender/sex and premature hospital discharge against medical advice, among people hospitalized with injection drug use-associated bacterial infections.

###### Age

Four studies assessed risk of premature hospital discharge against medical advice in relation to age.^63,64,70,76^ Younger age was associated with increased likelihood of premature discharge when measured categorically in two studies^64,76^ (e.g., aOR 3.02; 95%CI 2.10-4.34 for age 18-24 vs. 56-65 years)^64^. Younger age was associated with premature hospital discharge when assessed linearly in one study [Jo 2021^63^] (but intervals not defined; aOR 1.04; 95%CI 1.03-1.06) but not in another [Mertz 2008^70^] (per 10-year intervals; aOR 1.25; 0.83-2.00).

###### Race/ethnicity

Three studies assessed relationships between race/ethnicity and risk of premature hospital discharge.^63,64,76^ Associations were nonsignificant in two studies. In the third [Kimmel 2020]^64^, compared to white patients, Hispanic patients had a higher risk (aOR 1.31; 95%CI 1.03-1.69). Differences were nonsignificant for Black, Asian, Native American, and “Other” patients.

###### Income/employment

In the one study that assessed it [Kimmel 2020^64^], lower neighbourhood income quartile was associated with higher risk of premature hospital discharge (e.g., aOR 1.56; 95% 1.21, 1.99, for lowest income quartile vs. highest income quartile).

###### Unstable housing and homelessness

People with unstable housing or homelessness were more likely to have premature discharge against medical advice in one study [Cooksey 2020^61^] (uOR 4.6; 95%CI 1.4-15.0) but not in a second [Nolan 2020^72^] (aOR 1.39; 95%CI 0.62–3.12).

###### Overdose history

In one study [Serota 2021^76^], people with diagnostic codes for overdose were not more likely to have premature hospital discharge against medical advice (aOR 0.87; 95%CI 0.74-1.01). It was unclear if these overdose-related codes were during the same hospital admission, or a prior one.

###### Substance use (opioids, stimulants, alcohol, other substance use)

Five studies analysed substance use and risk of premature hospital discharge against medical advice, among people hospitalized with injecting-related infections.^63,64,70,72,76^ Compared to people who use only opioids, premature hospital discharge was more common among people who use only stimulants (aOR 1.09; 1.00-1.19) in one study [Serota 2021^76^]; differences were nonsignificant compared to people who use both stimulants and opioids (aOR 1.23, 0.85-1.77 in Jo 2021^63^; aOR 1.19, 95%CI 0.24-5.88 in Nolan 2020^72^). Risk of premature discharge was not associate with sedative or cannabis use in Kimmel 2020^64^, and was not associated with alcohol use in Kimmel 2020^64^ nor Mertz 2008^70^.

###### Opioid agonist treatment

Six studies assessed relationships between receiving opioid agonist treatment in hospital and risk of premature discharge against medical advice.^62,63,66,72,77,81^ Among four unadjusted effect estimates, meta-analysis summary was uOR 0.65 (95%CI 0.42-1.01; Figure 46). Among three fully-adjusted effect estimates, summary was aOR 0.69 (95%CI 0.31-1.56; Figure 47).

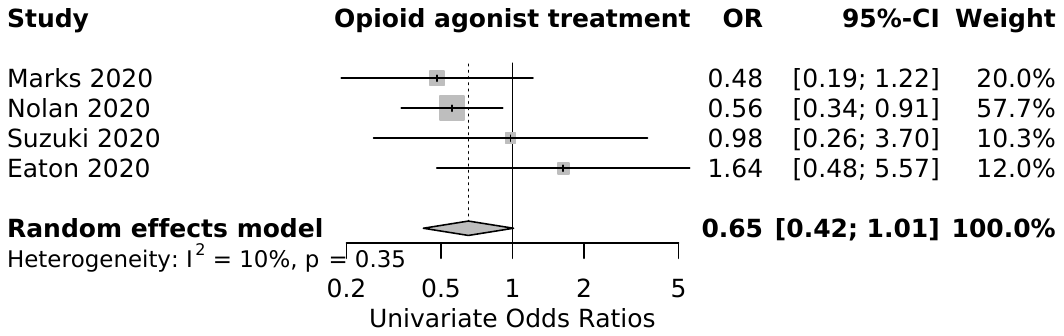
Figure 46. Meta-analysis of unadjusted effect estimates of relationship between opioid agonist treatment receipt and premature hospital discharge against medical advice, among people hospitalized with injection drug use-associated bacterial infections.

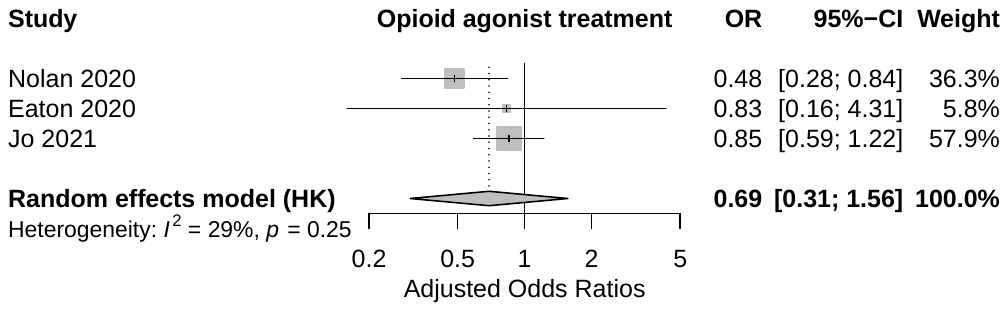
Figure 47. Meta-analysis of fully-adjusted effect estimates of relationship between opioid agonist treatment receipt and premature hospital discharge against medical advice, among people hospitalized with injection drug use-associated bacterial infections.

###### In-hospital addiction treatment

Hospital inpatient addiction medicine consultation was associated with lower risk of premature hospital discharge against medical advice (uOR 0.19, 95%CI 0.08–0.48), in one study [Marks 2020^66^].

###### Health care access (health insurance)

Increased risk of premature discharge was experienced among people without health insurance in all four studies that assessed it.^63,64,72,76^ One unadjusted effect estimate was uOR 3.93 (95%CI 2.17-7.13). Summary effect estimate for four adjusted effect estimates was aOR 2.06 (95%CI 1.09-3.91; Figure 48).

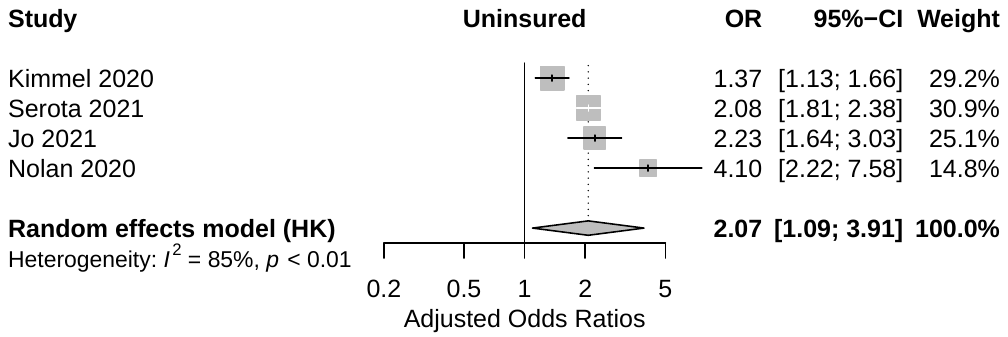
Figure 48. Meta-analysis of fully-adjusted effect estimates of relationship between lack of health insurance and premature hospital discharge against medical advice, among people hospitalized with injection drug use-associated bacterial infections.

###### Hospital characteristics

Risk of premature hospital discharge was not associated with hospital location/teaching status, nor hospital bed size, in Kimmel 2020.^64^

###### Hospital policies

In one study [Cooksey 2020^61^], following implementation of a new hospital-wide policy (to search patient’s belongings, supervise and limit all visitation, restrict cell phone access, provide analgesics and sedatives only in liquid formulation, make patients who inject drugs wear self-identifying gowns, and flag their medical chart), premature hospital discharges increased from 6% to 35% (p<0.001).

In Wang 2020^81^, before and after implementation of a hospital protocol to identify opioid use disorder and facilitate opioid agonist treatment, premature discharges were similar (from 42.2% to 40.8%, p=0.85).

###### Heart valve surgery during hospitalization

Among patients admitted to hospital with injecting-related endocarditis, having heart valve surgery during the hospital stay was associated with decreased risk of premature hospital discharges in two studies [Kimmel 2020^64^; Rudasill 2019^73^]. One unadjusted effect estimate was uOR 0.22 (95%CI 0.19-0.27; Rudasill 2019^73^) and one adjusted effect estimate was aOR 0.23 (95%CI 0.16-0.33; Kimmel 2020^64^).

##### New/secondary bloodstream infection during treatment

One study [Tan 2020^79^] assessed the following exposures in relation to developing a new (secondary) bloodstream infection during treatment for injection drug use-associated infective endocarditis: gender/sex; age; unstable housing and homelessness; substance use (heroin, stimulants, polysubstance use, other); substance use treatment; and insertion of peripherally-inserted intravenous central catheters (PICC lines) for parenteral antimicrobial treatment.

Increased risk of new bloodstream infections was seen among people who inject opiates (uOR 7.44; 95%CI 1.77-31.19, and 87% of participants injected opiates) and people who inject more than one substance (uOR 2.57; 1.27-5.21, and 76% injected more than one substance). Reduced risk weas seen among patients receiving an inpatient addiction medicine consultation (aHR 0.53l 95%CI 0.32-0.88).

Differences were nonsignificant for all other exposures, including experiencing homelessness (uOR 1.77; 95%CI 0.99-3.18), referral to outpatient addiction treatment (uOR 1.43; 95%CI 0.87-2.33), and PICC line insertion (aHR 0.60; 95%CI 0.14-2.56).

##### In-hospital death

Five studies assessed exposures associated with in-hospital mortality, during a hospital admission for injecting-related infection: gender/sex; age; race/ethnicity; overdose history; substance use (opioids, stimulants); health care access (insurance); hospital policies; and surgery during hospital admission.^68–70,75,76^ Associations were nonsignificant for most exposures. Significant associations are highlighted below.

###### Race/ethnicity

Hispanic people had higher risk of in-hospital mortality than non-Hispanic white people in one study [Serota 2021^76^] (aOR 1.27; 95%CI 1.01-1.61), while Black people’s risk of in-hospital death did not differ from white people (aOR 0.85; 95%CI 0.68-1.07). In another study [Saydain 2010^75^], the effect estimate for race and in-hospital mortality was imprecise and could include meaningful differences (uOR 1.33; 95%CI 0.28-6.30).

###### Substance use

In a U.S. nationwide study [Serota 2021^76^], higher risk of in-hospital death was associated with only-stimulant use (vs., only-opioid use; aOR 1.26, 95%CI 1.03-1.46), a history of overdose (aOR 1.26, 95%CI 1.01-1.59), and Medicaid insurance (publicly financed insurance, primarily for people with low income; aOR 1.41, 95%CI 1.09-1.82 vs. private insurance). People who were uninsured had lower risk of in-hospital death (aOR 0.74, 95%CI 0.55-0.98).

###### Hospital policy

Two studies assessed policy change as exposures. Following implementation of a policy of searching, surveillance, and restricting movement of people admitted to hospital with injection drug use-associated endocarditis, in-hospital mortality rates decreased from 11% to 0% (p=0.003) [Cooksey 2020^61^]. Following implementation of a new U.S. nationwide policy of reporting outcomes of aortic valve replacement surgery, the in-hospital mortality rate for patients with injection drug use-associated endocarditis changed from 3.7% (95% CI 2.2%–6.2%) to 3.2% (95% CI 1.8%–5.4%) [Kimmel 2020^65^].

###### Heart valve surgery

In both the studies that assessed it [Martín-Dávila 2005^68^; Rudasill 2019^73^], receiving valve surgery during hospitalization with injection drug use-associated endocarditis was associated with decreased in-hospital mortality.

##### Other outcomes during treatment

One study [Uppuluri 2021^80^] on risk factors for endogenous endophthalmitis among patients with injecting-related infections found these associated with female sex, non-white race/ethnicity, and infection of a central intravenous catheter. Endophthalmitis was less common among people with diagnostic codes for alcohol use disorder. One study [Sandrock 2001^74^] on risk factors for respiratory failure among people who inject drugs with botulism, identified no significant differences by gender or age.

### Appendix 19. Synthesis and meta-analyses of studies with outcomes occurring after treatment for injecting-related infections

Included studies assessed several different outcomes that occurred after initial treatment for injecting-related infections:

(a) infection-related rehospitalization;
(b) all-cause rehospitalization;
(c) overdose-related rehospitalization;
(d) all-cause mortality;
(e) other outcomes (failure of outpatient parental antimicrobial therapy [OPAT] and change in visual acuity following treatment for endogenous endophthalmitis).

Table 3. Summary of outcomes and associated exposures assessed among studies where outcome occurs after initial treatment for injecting-related infections.

| **Outcomes** | **Exposures assessed** | **Number of studies** |
| --- | --- | --- |
| Infection-related rehospitalization (after discharge from an initial hospital admission with injecting-related infections) | gender/sex; age; race/ethnicity; rural residency; substance use (injecting prescription opioids); opioid agonist treatment; other substance use treatment; hospital policy; premature hospital discharge against medical advice; cardiac surgery during admission | 8^52,82,89,90,94,95,97,101^ |
| All-cause rehospitalization | gender/sex; age; race/ethnicity; unstable housing; access to healthcare (health insurance); substance use (heroin, cocaine, methamphetamine, other); opioid agonist treatment; other addiction treatment; hospital policies; antibiotic treatment models; surgery during hospital admission | 9^61,63,73,81–83,92,95,99^ |
| Overdose-related rehospitalization | gender/sex; age; substance use; opioid agonist treatment; hospital policy | 2^83,81^ |
| All-cause mortality | gender/sex; age; unstable housing; substance use (opioid, stimulant, polysubstance use); premature hospital discharge against medical advice; opioid agonist treatment; other addiction medicine treatment; hospital policy; surgery during hospital admission | 14^61,67,79,81,85,91,93,94,96,98–102^ |
| Failure of outpatient parental antimicrobial therapy [OPAT] | age; discharge setting | 3^84,87,88^ |
| Change in visual acuity following treatment for endogenous endophthalmitis | gender/sex; age | 1^86^ |

See Appendix 14 for a list of all extracted effect estimates in this section.

##### Infection-related rehospitalization

Eight studies assessed relationships between the following exposures and infection-related rehospitalization (after people were discharged from an initial hospital admission with injecting-related infections): gender/sex; age; race/ethnicity; rural residency; substance use (injecting prescription opioids); opioid agonist treatment; other substance use treatment; hospital policy; premature hospital discharge against medical advice; cardiac surgery during admission.^52,82,89,90,94,95,97,101^ Most exposures (including gender/sex, race/ethnicity, rural residency, premature hospital discharge against medical advice, cardiac surgery during admission), were not significantly associated with risks of infection-related rehospitalization, though effect estimates often had wide confidence intervals that could include meaningful differences.

###### Age

Increasing age (measured continuously, in years) was associated with increased risk of infection related rehospitalization in one study [Barocas 2020^82^] (aOR 1.01; 95%CI 1.01–1.01), but not in two others [Huang 2018^90^; Thønnings 2020^52^].

###### Substance use (prescription opioids)

In one study [Huang 2018^90^], people who had multiple hospital admissions for endocarditis were more likely to inject prescription opioids compared to people who had only one single hospital admission for endocarditis.

###### Opioid agonist treatment

Four studies assessed relationships between opioid agonist treatment and infection-related rehospitalization.^52,82,89,101^ In unadjusted analyses, people prescribed opioid agonist treatment had lower rates of rehospitalization (10.3 [95%CI 9.87-10.64] per 100 person-years vs. 18.7 [95%CI 18.53-18.78] per 100 person-years) in one study [Barocas 2020^82^], and rates did not differ between groups in three other studies [Hilbig 2020^89^; Suzuki 2020^101^; Thønnings *2020*^52^]. Two fully-adjusted effect estimates for opioid agonist treatment from the same study [Barocas 2020^82^], were aHR 0.49 (95%CI 0.18-1.23) for infection-related rehospitalization by 30 days and aHR 0.41 (95%CI 0.42–0.91) for infection-related rehospitalization by 1 year. A new hospital policy to identify opioid use disorder and facilitate opioid agonist treatment did not change 90-day rates of infection-related rehospitalization in one study [Ray 2020^95^].

###### Other addiction treatment

In two studies [Thønnings 2020^52^*;* Rodger 2019^97^], referrals to outpatient addiction treatment were not associated with risk of infection-related rehospitalization.

##### All-cause rehospitalization

Nine studies assessed factors associated with all-cause rehospitalization (following an initial hospital admission with injecting-related infections): gender/sex; age; race/ethnicity; unstable housing; access to healthcare (health insurance); substance use (heroin, cocaine, methamphetamine, other); opioid agonist treatment; other addiction treatment; hospital policies; antibiotic treatment models; surgery during hospital admission.^61,63,73,81–83,92,95,99^ Several of the exposures (including age; unstable housing; health insurance; heroin cocaine; methamphetamine; benzodiazepine use) were not significantly associated with the outcome.

###### Gender/sex

Woman/female gender/sex was associated with increased risk of all-cause rehospitalization. Summary meta-analysis of three fully-adjusted effect estimates was aOR 1.22 (95%CI 1.08-1.38; Figure 49). One unadjusted effect estimate was nonsignificant at uOR 1.23 (95%CI 0.77-1.96).

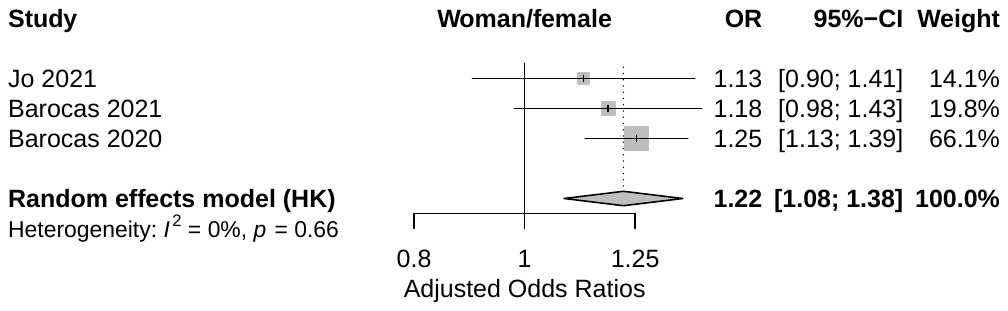

Figure 49. Meta-analysis of fully-adjusted effect estimates of relationship between woman/female gender/sex and all-cause rehospitalization, following discharge from an initial hospital admission with injection drug use-associated bacterial infections.

###### Substance use

People who had diagnostic codes for multiple substances (including alcohol, cannabis, hallucinogens, or sedatives), were at higher risk of all-cause rehospitalization in one study (aHR 1.29; 95%CI 1.11–1.50) [Barocas 2020^82^].

###### Opioid agonist treatment

Four studies assessed receipt of opioid agonist treatment and risk of all-cause rehospitalization; the studies measured opioid agonist treatment receipt in hospital or at discharge, or received within 30 days of hospital discharge.^63,81–83^ Summary of two unadjusted effect estimates was uOR 0.82 (95%CI 0.42-1.60; Figure 50). Meta-analytic summary of three fully-adjusted effect estimates was aOR 0.98 (95%CI 0.75-1.29; Figure 51).

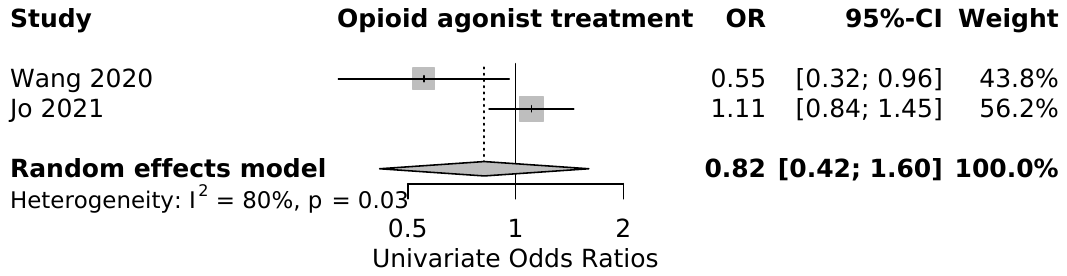
Figure 50. Meta-analysis of unadjusted effect estimates of relationship between receiving opioid agonist treatment (during hospitalization or at discharge) and all-cause rehospitalization, following discharge from an initial hospital admission with injection drug use-associated bacterial infections.

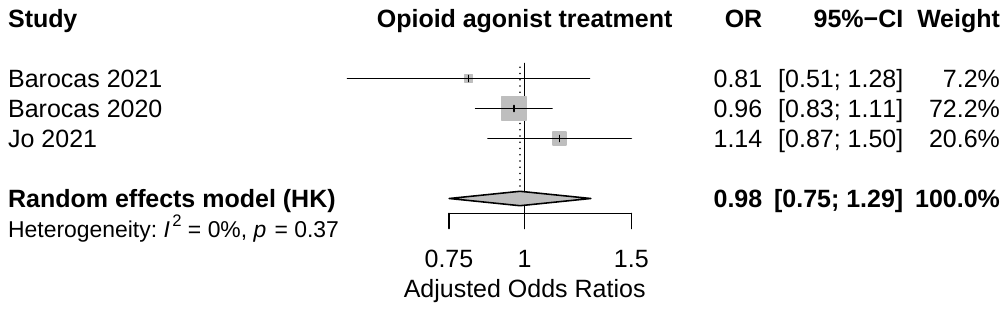
Figure 51. Meta-analysis of fully-adjusted effect estimates of relationship between receiving opioid agonist treatment (during hospitalization or within 30 days following discharge) and all-cause rehospitalization, following discharge from an initial hospital admission with injection drug use-associated bacterial infections.

###### In-hospital addiction medicine treatment

In two studies, receiving an inpatient addiction medicine consultation was associated with reduced risk of all-cause rehospitalization; summary of two unadjusted effect estimates was uOR 0.46 (95%CI 0.33-0.63; Figure 52) and one fully-adjusted effect estimate was aOR 0.57 (95%CI 0.38–0.86; Marks 2019^66^).

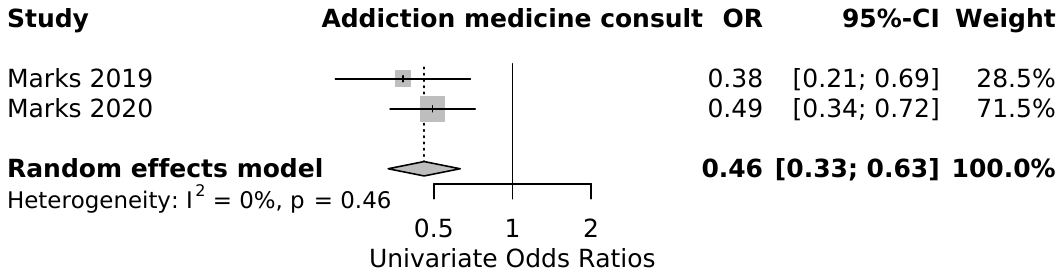

Figure 52. Meta-analysis of unadjusted effect estimates of relationship between receiving an addiction medicine consultation (during hospitalization) and all-cause rehospitalization, following discharge from an initial hospital admission with injection drug use-associated bacterial infections.

###### Hospital policy

Three studies assessed the impact of hospital policy change on all-cause readmission, following an initial hospital admission with an injecting-related infection.^61,81,95^ In Cooksey 2020^61^, implementation of a new hospital policy of searching, surveillance, and restricting movement of people who use drugs was associated with a decrease in 90-day all-cause readmissions (aOR 0.2, 95%CI 0.08-0.6; from 48% pre-implementation to 34% post-implementation). In two other studies [Ray 2020^95^; Wang 2020^81^], implementation of hospital policies to facilitate access to opioid agonist treatment were not associated with changes in readmission rates.

###### Antimicrobial treatment mode

One study [Marks 2020^92^] assessed whether patients with injecting-related bacterial infections who left hospital prematurely received oral antibiotics to finish out their course. Compared to completing a full inpatient course of intravenous antibiotics, people receiving partial intravenous/partial oral treatment courses had similar risk of all-cause readmission (aHR 0.99, 95%CI 0.62-1.62). People receiving partial intravenous/no oral treatment had higher risk of all-cause readmission (aHR 2.32, 95%CI 1.41 – 3.82).

###### Surgery during hospitalization

Three studies assessed the impact of surgical intervention during a hospital admission for injecting-related infections, on risk of all-cause rehospitalization.^73,92,99^ Surgery was associated with decreased risks of readmission in the two studies that assessed it [Marks 2020^92^; Rudasill 2019^73^]. One study [Slaughter 2019^99^] compared different types of cardiac surgery in people with tricuspid valve endocarditis; valvectomy was associated with higher rates of all-cause readmission than repair or replacement (aOR 5.42, 95%CI 2.33–12.57, vs. repair).

##### Overdose-related rehospitalization

Two studies [Barocas 2021^83^; Wang 2020^81^] assessed factors associated with opioid overdose-related rehospitalization, following an initial hospital admission with injecting-related infections: gender/sex; age; substance use; opioid agonist treatment; hospital policy.^81,83^ Risks of rehospitalization with overdose was associated with age (aHR 0.97; 95%CI 0.95–0.99) but not female sex (aHR 0.89; 95%CI 0.47–1.63) nor substance use (diagnostic codes related to alcohol, cannabis, hallucinogens, or sedatives; aHR 0.86; 95%CI 0.26–2.84) in one study.

Both studies assessed the impact of opioid agonist treatment. In one study [Barocas 2021^83^], receiving a prescription within 30 days of hospital discharge was not associated with rehospitalization for overdose (aHR 0.86; 95%CI 0.26–2.91), but the effect estimate was nonspecific and could include meaningful differences. In the second [Wang 2020^81^], having opioid agonist treatment continued at hospital discharge was associated with decreased risk of 30-day (uRR 0.34; 95% CI 0.16-0.74) and 90-day opioid-related readmission (uRR 0.46; 95%CI 0.24-0.88). Implementation of a hospital protocol to facilitate access to opioid agonist treatment did not lead to significant changes in opioid-related readmission (e.g., uRR 0.83, 95% CI 0.44-1.57 at 30 days) [Wang 2020^81^].

##### All-cause mortality

Fourteen studies assessed the following exposures in relation to all-cause mortality after surviving an initial hospital admission with an injecting-related infection: gender/sex; age; unstable housing; substance use (opioid, stimulant, polysubstance use); premature hospital discharge against medical advice; opioid agonist treatment; other addiction medicine treatment; hospital policy; surgery during hospital admission.^61,67,79,81,85,91,93,94,96,98–102^ All-cause mortality after hospital discharge was not associated with several outcomes (gender/sex, unstable housing, stimulant-only use, polysubstance use, and premature hospital discharge against medical advice). Risk of all-cause mortality was increased among people who use only opioids (vs. use other substances or multiple substances) in one study (uRR 1.72; 95%CI 1.06–2.80) [Rodger 2018^96^].

###### Opioid agonist treatment

Four studies assessed relationships between opioid agonist treatment and all-cause mortality after hospital discharge.^67,91,96,101^ Three studies provided unadjusted effect estimates of the relationship between opioid agonist treatment prescriptions provided at hospital discharge and risk of all-cause mortality. Meta-analysis summary was uOR 0.58 (95%CI 0.24-1.45; Figure 53). In the only fully-adjusted effect estimate [Kimmel 2020^91^], opioid agonist treatment was associated with reduced risks of all-cause death in the month within which it was received (when treated as a time-varying exposure; aHR 0.30; 95% CI 0.10-0.89).

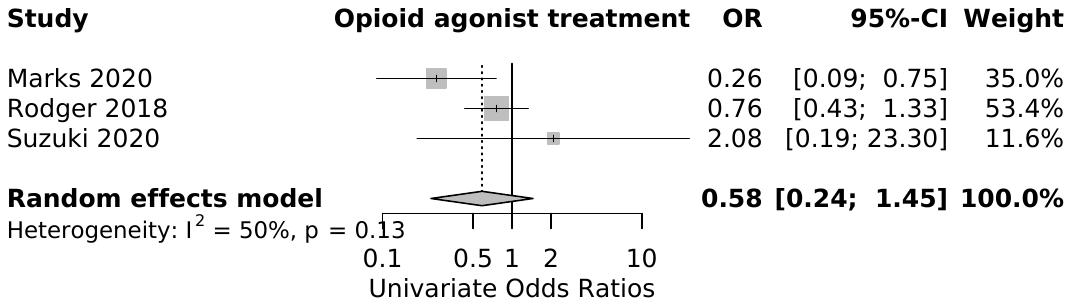

Figure 53. Meta-analysis of unadjusted effect estimates of relationship between receiving opioid agonist treatment at hospital discharge and all-cause mortality, following discharge from an initial hospital admission with injection drug use-associated bacterial infections.

###### In-hospital addiction medicine consultation

Three studies assessed risk of all-cause mortality after hospital discharge in relation to receiving other addiction treatment in hospital. In one study [Nguemeni Tiako 2020^93^], 0 out of 20 people with endocarditis who received “comprehensive addiction treatment” (defined as psychiatry and social work consultation and/or opioid agonist treatment during hospitalization) died at 24 months after hospital discharge, compared to 7 out of 22 who did not. In Tan 2020^79^, receiving an inpatient addiction medicine consultation was not statistically significantly associated with all-cause mortality after discharge (aHR 0.64, 95%CI 0.32–1.29), though the confidence interval was wide and could include meaningful differences.

###### Other substance use treatment

In Rodger 2018^96^, referral to outpatient addiction treatment at hospital discharge was associated with reduced all-cause mortality (aHR 0.29; 95%CI 0.12–0.73). However, it seems as if the all-cause mortality outcome here included some patients who died in hospital, so this may have simply indicated those who survived to hospital discharge and reflect “immortal time”.

###### Hospital policies

In Cooksey 2020^61^, following implementation of a hospital policy of searching, surveillance, and restricting movement of people with injecting-related infections, all-cause mortality at 12 months decreased from 7% to 4% (aOR 0.25; 95%CI 0.07–0.89). In Wang 2020^81^, following implementation of a hospital policy to facilitate access to opioid agonist treatment, all-cause mortality by 3 months changed from 2.8% (2/71) to 3.9% (3/76), and they concluded, “These numbers were too small to compare”.

###### Surgery during hospitalization

Five studies assessed the impact of heart valve surgery during hospitalization with injecting-related endocarditis, and all-cause mortality following discharge. Meta-analysis summary of three unadjusted effect estimates was uOR 0.95 (95%CI 0.49-1.82; Figure 54), and for four fully-adjusted effect estimates was aOR 0.61 (95%CI 0.19-1.93; Figure 55).

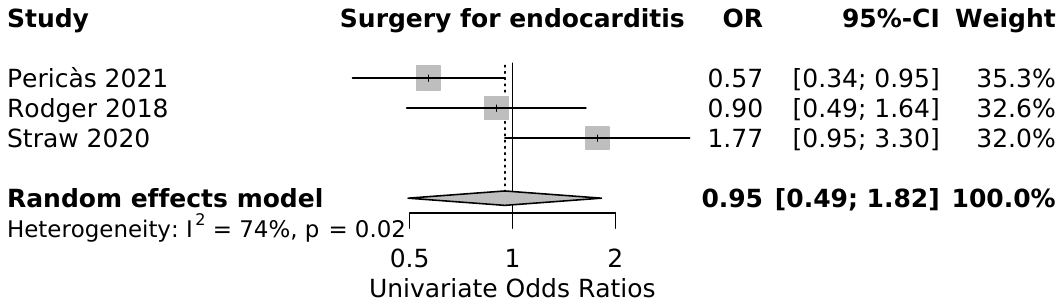
Figure 54. Meta-analysis of unadjusted effect estimates of relationship between receiving surgery and all-cause mortality following discharge from an initial hospital admission with injection drug use-associated endocarditis.

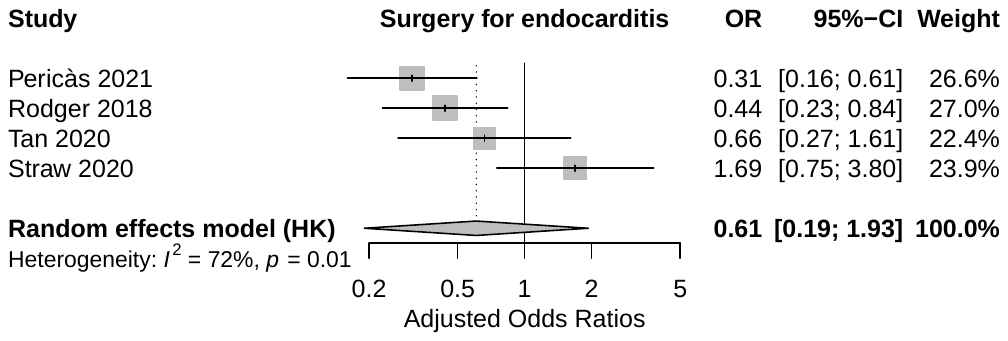
Figure 55. Meta-analysis of fully-adjusted effect estimates of relationship between receiving surgery and all-cause mortality following discharge from an initial hospital admission with injection drug use-associated endocarditis.

One of these studies compared types of heart valve surgery among people with injecting-related tricuspid valve endocarditis. Compared to valve repair, valvectomy (removal of the valve without replacement) was associated with higher risk of all-cause mortality (aOR 5.42; 95%CI 2.33–12.57), and valve replacement did not have significantly different risk (aOR 1.04; 95%CI 0.47–2.27).

##### Other outcomes

Studies assessed other outcomes after treatment for injecting-related bacterial infections, including outpatient parenteral antibiotic treatment (“OPAT”) failure^84,87,88^ and change in visual acuity after treatment for endogenous endophthalmitis.^86^

OPAT failure (after initial hospitalization with an injecting-related infection) was not associated with age^84^ and did not differ according to whether patients were discharged home or to a post-acute care nursing facility.^87^ In a pilot trial [Fanucchi 2020]^88^ that randomized 20 patients to either complete antibiotic treatment as a hospital inpatient vs. OPAT integrated with opioid agonist treatment, all participants completed the recommended antibiotic treatment course. In one study [Connell 2010]^86^, improvement in visual acuity after treatment for endogenous endophthalmitis was seen more commonly among men than among women, but did not differ by age group.

### Appendix 20. Synthesis and meta-analyses of studies where outcome is colonization with pathogenic bacteria.

Five studies assessed factors associated with colonization with specific pathogenic bacteria among people who inject drugs, including *Staphylococcus aureus* and methicillin-resistant *S. aureus*: gender/sex; age; race/ethnicity; education; employment; relationship status; unstable housing and homelessness; incarceration; substance use (heroin, cocaine, crack, speedball, methamphetamines, prescription opioids; cannabis); public injecting; injecting in groups; opioid agonist treatment; other addiction treatment; recent hospital admission; and other (e.g. using public shower facilities).^103–107^ See for all effect estimates extracted for this section.

Most exposures were not significantly associated with colonization. Several exposures had significant associations in some studies but not others. Meta-analysis for four unadjusted effect estimates on the relationship between homelessness and colonization with pathogenic bacteria was uOR 1.84 (95%CI 0.81-4.18; Figure 56), and we identified no fully-adjusted effect estimates. Recent hospital admission was significantly associated with colonization in one study (uOR 4.3; 95%CI 1.34-13.80) [Packer 2019^107^], but not in two others [Leung 2015^105^; Miller 2007^106^].

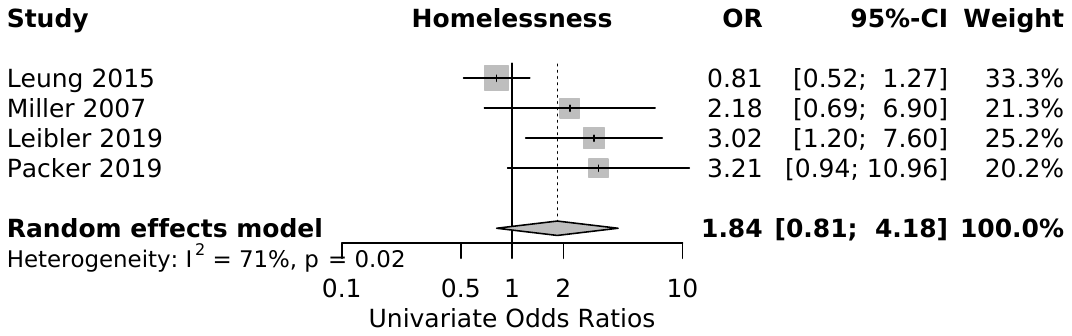

Figure 56. Meta-analysis of unadjusted effect estimates of relationship between homelessness and colonization with pathogenic bacteria among people who inject drugs.

Several other exposures has significant associations in single studies. In one study [Packer 2019^107^], public injecting (uOR 5.5; 95%CI 1.34-22.73) and frequently injecting in groups of three or more people (uOR 15.8; 95%CI 2.51-99.28) were both associated with colonization. In a second study [Leibler 2019^4^], positive associations were seen with sleeping at more than one place during the last week (uOR OR 3.1; 1.3-7.6), use of public shower facilities in the last week (uOR 13.7, 95%CI 1.4-132.8), and sharing bedding with other people (uOR 2.2; 95%CI 1.0 –4.7).
